# Estimates of protection against SARS-CoV-2 infection and severe COVID-19 in Germany before the 2022/2023 winter season - the IMMUNEBRIDGE project

**DOI:** 10.1101/2023.02.16.23285816

**Authors:** Berit Lange, Veronika K Jaeger, Manuela Harries, Viktoria Rücker, Hendrik Streeck, Sabine Blaschke, Astrid Petersmann, Nicole Toepfner, Matthias Nauck, Max J Hassenstein, Maren Dreier, Isabell Von Holt, Axel Budde, Antonia Bartz, Julia Ortmann, Marc-André Kurosinski, Reinhard Berner, Max Borsche, Gunnar Brandhorst, Melanie Brinkmann, Kathrin Budde, Marek Deckena, Geraldine Engels, Marc Fenzlaff, Christoph Härtel, Olga Hovardovska, Alexander Katalinic, Katja Kehl, Mirjam Kohls, Stefan Krüger, Wolfgang Lieb, Kristin M Meyer-Schlinkmann, Tobias Pischon, Daniel Rosenkranz, Nicole Rübsamen, Jan Rupp, Christian Schäfer, Mario Schattschneider, Anne Schlegtendal, Simon Schlinkert, Lena Schmidbauer, Kai Schulze-Wundling, Stefan Störk, Carsten Tiemann, Henry Völzke, Theresa Winter, Christine Klein, Johannes Liese, Folke Brinkmann, Patrick F Ottensmeyer, Jens-Peter Reese, Peter Heuschmann, André Karch

**Author notes:** **Correspondence:** Berit Lange & André Karch. Members of the IMMUNEBRIDGE STUDY GROUP, listed in alphabetical order. indicates shared first authorship (Berit Lange, Veronika K Jaeger). indicates last authorship (André Karch).

## Abstract

Despite the need to generate valid and reliable estimates of protection against SARS-CoV-2 infection and severe course of COVID-19 for the German population in summer 2022, there was a lack of systematically collected population-based data allowing for the assessment of the protection level in real-time.

In the IMMUNEBRIDGE project, we harmonised data and biosamples for nine population-/hospital-based studies (total number of participants n=33,637) to provide estimates for protection levels against SARS-CoV-2 infection and severe COVID-19 between June and November 2022. Based on evidence synthesis, we formed a combined endpoint of protection levels based on the number of self-reported infections/vaccinations in combination with nucleocapsid/spike antibody responses (“confirmed exposures”). Four confirmed exposures represented the highest protection level, and no exposure represented the lowest.

Most participants were seropositive against the spike antigen; 37% of the participants ≥79 years had less than four confirmed exposures (highest level of protection) and 5% less than three. In the subgroup of participants with comorbidities, 46-56% had less than four confirmed exposures. We found major heterogeneity across federal states, with 4%-28% of participants having less than three confirmed exposures.

Using serological analyses, literature synthesis and infection dynamics during the survey period, we observed moderate to high levels of protection against severe COVID-19, whereas the protection against SARS-CoV-2 infection was low across all age groups. We found relevant protection gaps in the oldest age group and amongst individuals with comorbidities, indicating a need for additional protective measures in these groups.

## Introduction

During the SARS-CoV-2 pandemic, there was a lack of population-based panels in Germany capable of providing rapid and adaptable estimates of population immunity, vaccination coverage, infection dynamics, and underdetection of notified infections over time (1). As a partial substitute, several individual studies tried to bridge this gap, e.g. by providing estimates of population immunity at defined time points early in the pandemic (2-8). However, these studies were often cross-sectional and could not inform real-time infectious disease modelling rapidly and continuously enough to serve as a basic and pivotal information layer for what needed to be communicated to political decision-makers (9).

The absence of centrally organised population panels resulted in a lack of data suited to provide evidence on population immunity in Germany after the summer of 2022. Additionally, there was relevant uncertainty regarding the choice of adequate endpoints for correlates of protection against infection or severe COVID-19 that would allow their use in real-time decision-making (10).

To derive estimates of protection against SARS CoV-2 infection and severe COVID-19 in the German general population, the IMMUNEBRIDGE project was established within the Network of University Medicine (NUM). It aimed to bring together established large population-based cohorts with newly designed cross-sectional studies by using central data linkage structures, targeted literature synthesis, and central laboratory infrastructure, to allow the preparation of action plans for winter 2022. Rapid and continuous communication to a newly established modelling network for severe infectious diseases (MONID) was essential to allow usage of results in the communication of harmonised scenario modelling from this network to decision-makers and the public.

Here, we report estimates for the level of protection against SARS-CoV-2 infection and severe COVID-19 from nine epidemiological studies surveying 33,637 participants between June and November 2022 based on a combined endpoint developed for this study.

## Methods

The IMMUNEBRIDGE project aimed to provide a comprehensive picture of the protection level within the German population against SARS-CoV-2 infection and severe COVID-19 from June to November 2022 using existing population-based cohort studies and newly set-up cross-sectional studies. Antibodies against the spike (S-) and nucleocapsid (N-) antigen of SARS-CoV-2 were measured within the participating studies, and seropositivity proportions for predefined subgroups (age, sex, and comorbidities) were derived. In addition, information on vaccination and infection history of the study participants were obtained. These data were harmonised across participating studies using a jointly developed minimal data set (MDS). The project was structured to enable early ad-hoc feedback to consortia of a newly established modelling network for severe infectious diseases (MONID)(11) from early August onwards. For this, aggregated data were presented in a model-usable format in two interim reports in August (12) and October 2022 (13) to support the MONID modelling consortia. Within the framework of IMMUNEBRIDGE, a targeted literature synthesis was carried out to derive a categorisation of protection levels against infection and severe COVID-19 into a “combined endpoint” based on self-reported infections/vaccinations and immune correlates (Table 1 and Supplement 2).

**Table 1:** Definition of the combined endpoint for estimating protection against SARS-CoV-2 infections and overview of protective effects against SARS-CoV-2 derived from the literature. The range of vaccine effectiveness (VE) point estimates is provided of n included studies on adults aged above 18 years (reference group: no vaccination, infection, or antibodies; further and more detailed results on children and adolescents, adults > 60 years, and different reference groups are provided in the Supplement 2, S3-S5). A confirmed exposure is either a vaccination or an infection with humoral immune correlates^1^

| Combined endpoint definition (a confirmed exposure is either a vaccination or an infection with humoral immune correlates<sup>1</sup>) |  | Literature based protective effects of each combined endpoint category |  |  |  |  |  | Level of protection to be used in model parameterisation depending on variants<sup>2</sup> |
| --- | --- | --- | --- | --- | --- | --- | --- | --- |
| Abbreviated definition (used in text/tables/figures) | Definition | Vaccine effectiveness against: | Vaccine effectiveness for studies without follow-up time | Vaccine effectiveness 0-13 days after vaccination | Vaccine effectiveness 14-90 days after vaccination | Vaccine effectiveness 91-180 days after vaccination | Vaccine effectiveness >180 days after vaccination |  |
| 3 + 1 confirmed exposures (referred to as 4 exposures in the text) | Three exposures (vaccination or infection) with humoral immune correlates<sup>1</sup>+ one infection or vaccination in 2022 | Infection<sup>3</sup> | 34-83% (n=5) | 36% (n=1) | 20-92% (n=4) | 5-82% (n=4) |  | Probably high protection against severe progression; Low protection against infection |
|  |  | Severe course<sup>4</sup> | 68-100% (n=5) |  | 44-98% (n=5) | 37-86% (n=2) |  |  |
|  |  | Death | 81% (n=1) |  |  |  |  |  |
| 3 confirmed exposures | Three exposures (vaccination or infection) with humoral immune correlates<sup>1</sup> | Infection<sup>3</sup> | -7-78% (n=16) | 18-69% (n=8) | 1-91% (n=17) | 5-67% (n=8) | -25-55% (n=3) | Moderate protection against severe progression; Very low protection against infection |
|  |  | Severe course<sup>4</sup> | 31-99% (n=9) | 65-96% (n=4) | 31-97% (n=10) | 29-95% (n=14) | 31-88% (n=3) |  |
|  |  | Death | 87-98% (n=3) | 71-86% (n=1) | 78-88% (n=3) | 74-87% (n=2) | 77% (n=1) |  |
| 1-<3 confirmed exposures | 3 or more exposures without immune correlate, 1-2 exposures with or without immune correlate or 0 exposure with at least 1 proven humoral immune correlate | Infection<sup>3</sup> | -46-65% (n=12) | 10-23% (n=2) | 8-55% (n=13) | 1-54% (n=15) | -14-52% (n=11) | Low protection against severe progression; Very low protection against infection |
|  |  | Severe course<sup>4</sup> | 33-86% (n=11) |  | -9-96% (n=15) | 24-87% (n=14) | 12-92% (n=9) |  |
|  |  | Death | 66-90% (n=4) | -9-29% (n=2) | 60-62% (n=2) | 57-70% (n=1) | 49-57% (n=2) |  |
| 0 Exposure | No humoral immune correlates and no exposures |  |  |  |  |  |  | No protection against severe course or infection |

<sup>1</sup>A confirmed exposure requires one humoral immune correlate: antibody to S-antigen for vaccination or antibody to N-antigen for infection.
<sup>2</sup>The categorisation given is only applicable for adults aged 18 and above and cannot be directly applied to children and adults above 60 years.
<sup>3</sup>Different definitions were used for infection: any, symptomatic, documented infection
<sup>4</sup>Different definitions were used for severe course of disease: Hospitalisation, treatment in intensive care, and/or death.

### Participating studies

From June to the end of November 2022 new survey rounds on vaccination status and previous infections were performed, blood samples were collected, and antibodies against SARS-Cov-2 were measured in four existing population-based cohort studies: the ELISA-Study (7, 14), MuSPAD (4), the German National Cohort (NAKO) (8), and STAAB (5). Four new cross-sectional studies were set up, one to provide regional depth (GUIDE Study; DRKS00029693) and three to provide relevant data on children (studies of the University Hospitals in Dresden (adolescents; DRKS00022549), Bochum (school children) (2), and Würzburg (pre-school children; Wü-Kita-CoV) (3)). Studies were included if they were able to quickly survey and sample in the general population regionally or supra-regionally, and provide the MDS. For the GUIDE study, capillary blood was collected by the participants themselves and put on dried blood cards; for all other studies venous blood samples were collected on-site. The principle desirable characteristics of epidemic panels such as speed, adaptability, and linkage to other sources were realised differently in the participating studies. If some characteristics were not available in a particular study, the network format of the project ensured that these aims could be reached for the overall project. For example, not all studies were able to collect samples in June 2022, but this was compensated by the capacity of other studies or study sites to do this. Further, as not all studies were able to provide regional depth, other studies were built up for this purpose (Supplement 1 Table S1).

In addition to the population-based studies, a prospective hospital-based study called IMMUNEBRIDGE_ED was conducted by the Central Emergency Department of the University Medical Center Göttingen to provide information on defined risk groups not covered in sufficient depth by the available population-based studies. Inclusion criteria for IMMUNEBRIDGE_ED comprised geriatric patients as well as patients with pre-existing immunodeficiency or severe pre-existing diseases.

We followed STROBE as a reporting guideline for observational studies (15). Further information on the participating studies, such as eligibility criteria, can be found in cohort profile publications of these studies (2-7, 16) (Supplement 1 Table S1).

For all participating studies, local ethics committees’ votes were obtained prior to the start of the IMMUNEBRIDGE project. Participants and patients were included only after written informed consent in each clinical study.

### Laboratory analyses

The serum samples obtained in the STAAB, MuSPAD, ELISA, all children cohorts and IMMUNEBRIDGE_ED by on-site blood collection were analysed in the Institute of Clinical Chemistry and Laboratory Medicine in Oldenburg. Serum samples from NAKO were analysed in the Institute of Clinical Chemistry and Laboratory Medicine in Greifswald. The Elecsys® Anti-SARS-CoV-2 S and Elecsys® Anti-SARS-CoV-2 NC (both Roche Diagnostics, Mannheim, Germany) were used to determine the quantitative antibody response to the S- and qualitative antibody response to the N-antigen of SARS-CoV-2. Seropositivity to the S-antigen was defined as ≥0.80 BAU/ml (BAU: Binding Antibody Units), seropositivity to the N-antigen as being above the cut-off index of 1.0.

For GUIDE, capillary blood was collected by the participants themselves, put on dried blood cards (Ahlstrom-Munksjö TFN 460), and sent by mail to the laboratory MVZ Labor Krone GbR (Bad Salzuflen). To analyse the dried blood spot (DBS) samples, the Anti-SARS-CoV-2-QuantiVac ELISA (IgG) was used for the S-antigen and the Anti-SARS-CoV-2-NCP ELISA (IgG) for the N-antigen. Both assays have been validated for use with sample material from DBS cards by both the manufacturer and the performing laboratory. Seropositivity against the S-antigen was defined as ≥35.2 BAU/ml, against the N-antigen as a ratio of >1.0. In the main analysis, DBS samples with borderline findings (ratio ≥0.8 to ≤1.0) for the N-antigen were defined as seropositive. This represents a conservative estimate of the waning function with the aim not to underestimate true protection in the population (as a positive antibody response served only as a confirmation of a self-reported infection in the main analysis).

### Data collection and linkage

Data pooling took place using the Serohub (www.serohub.net) based on the MDS previously agreed on with the participating studies. The Serohub was designed during the SARS-CoV-2 pandemic as a digital tool able to link and collect individual participant data using common data sharing and MDS documents. In the MDS, variables regarding demographic, socioeconomic, and medical characteristics of participants were agreed on, and reporting categories of these variables were defined. Furthermore, variables regarding test characteristics, assay results, and reporting categories were fixed (Supplement 1 Table S2).

### Combined endpoint for protection levels against SARS-CoV-2 infection and severe course of disease and data analysis

As a proxy for protection levels against SARS-CoV-2 infection and severe course of disease we formed a “combined endpoint” based on the number of previous self-reported infections and vaccinations hereafter called “exposures” as well as current antibody status against the N- and S-antigen. Based on a targeted literature synthesis, four categories of protection levels were formed and associated with graded levels of protection against (re-)infection and severe COVID-19 (Table 1). The four combined endpoint categories are as follows: *3+1 confirmed exposures* (with at least one confirmed exposure in 2022), corresponding to the highest level of protection; *3 confirmed exposures* (regardless of the timing of the exposures); *1*-<*3 confirmed exposures* (regardless of the timing of the exposures); and *0 exposures* (highest to lowest protection level respectively), indicating no protection, Table 1). We defined a confirmed exposure as a self-reported infection or vaccination with a corresponding humoral immune correlate (antibody to N-antigen for infection, antibody to S-antigen for vaccination).

The combined endpoint was reported stratified by age groups, self-reported comorbidities (hypertension, diabetes, lung diseases, cardiovascular diseases, cancers, diseases leading to immunosuppression), and self-reported sex. All estimates represent results that were age-standardised according to the 2021 population status update of the 2011 German census of the Federal Statistical Office using the *survey* package in R (17). Results that were not age-standardised can be found in Supplementary 1. In addition, the combined endpoint was presented stratified according to Nomenclature des Unités territoriales statistiques (NUTS) 2 regions as well as federal states (NUTS 1), using the R package *eurostat* (18). NUTS divides areas of the European Union into three levels, allowing for cross-border comparisons (19).

In the analyses comparing vulnerable groups from the population-based studies with the IMMUNEBRIDGE_ED cohort, the results of the population-based cohorts were age-standardised according to the age distribution in the IMMUNEBRIDGE_ED comorbidity groups.

### Sensitivity analyses

In the sensitivity analyses, we assessed the combined endpoint if only self-reported information on infection and vaccination and no humoral immunity correlates for confirmation were included. Additionally, we evaluated the combined endpoint and seropositivity if DBS samples with borderline findings (ratio ≥0.8 to ≤1.0) were treated as seronegative to provide a lower bound of protection levels compatible with the antibody analysis.

All analyses were carried out in R version 4.1 and 4.2 (20).

## Results

### General

A total of 33,637 participants from nine studies surveyed between June and November 2022 were included. Supplement 1, Table S1 describes the different participating studies, and Table 2 shows the baseline characteristics of the participants in these studies.

**Table 2:** Characteristics of participants with data from the minimal data set stratified by study (n=33,637).

|  | Population-based cohorts |  |  |  |  | Children's cohorts |  |  | Risk group | Total |
| --- | --- | --- | --- | --- | --- | --- | --- | --- | --- | --- |
|  | GUIDE*<br>(N=15932) | NAKO<br>(N=10595) | STAAB<br>(N=1711) | MuSPAD<br>(N=3032) | ELISA<br>(N=1389) | Wü-KITa-CoV<br>(N=275) | paedSAXCOVID<br>(N=91) | CorKID<br>(N=189) | Immunebridge_ED<br>(N=423) | Overall<br>(N=33637) |
| **Age (years)** |  |  |  |  |  |  |  |  |  |  |
| <18 | - | - | - | - | - | 275 (100.0%) | 75 (87.2%) | 186 (98.9%) | - | 536 (1.6%) |
| 18-29 | 1382 (8.7%) | 296 (2.8%) | - | 128 (4.4%) | 96 (6.9%) | - | 11 (12.8%) | <6 | 13 (3.1%) | 1928 (5.8%) |
| 30-34 | 1206 (7.6%) | 566 (5.3%) | - | 148 (5.1%) | 79 (5.7%) | - | - | - | 11 (2.6%) | 2010 (6.0%) |
| 35-39 | 1217 (7.6%) | 522 (4.9%) | 34 (2.0%) | 135 (4.6%) | 95 (6.8%) | - | - | - | 8 (1.9%) | 2011 (6.0%) |
| 40-49 | 2412 (15.1%) | 1607 (15.2%) | 166 (9.8%) | 345 (11.8%) | 231 (16.6%) | - | - | - | 19 (4.5%) | 4780 (14.3%) |
| 50-59 | 3612 (22.7%) | 3213 (30.3%) | 476 (28.0%) | 754 (25.8%) | 362 (26.1%) | - | - | - | 59 (13.9%) | 8476 (25.3%) |
| 60-64 | 1540 (9.7%) | 1479 (14.0%) | 265 (15.6%) | 388 (13.3%) | 193 (13.9%) | - | - | - | 39 (9.2%) | 3904 (11.7%) |
| 65-79 | 3992 (25.1%) | 2912 (27.5%) | 683 (40.2%) | 883 (30.2%) | 321 (23.1%) | - | - | - | 125 (29.6%) | 8916 (26.6%) |
| 80+ | 568 (3.6%) | - | 74 (4.4%) | 141 (4.8%) | 12 (0.9%) | - | - | - | 149 (35.2%) | 944 (2.8%) |
| Missing | 3 | 0 | 13 | 110 | 0 | 0 | 5 | 1 | 0 | 132 |
| **Sex** |  |  |  |  |  |  |  |  |  |  |
| Female | 8177 (51.3%) | 5382 (51.3%) | 891 (52.5%) | 1759 (60.1%) | 778 (56.0%) | 134 (48.7%) | 62 (68.1%) | 89 (47.1%) | 237 (57.0%) | 17509 (52.4%) |
| Male | 7755 (48.7%) | 5115 (48.7%) | 807 (47.5%) | 1169 (39.9%) | 611 (44.0%) | 141 (51.3%) | 29 (31.9%) | 100 (52.9%) | 179 (43.0%) | 15906 (47.6%) |
| Diverse | - | - | - | - | - | - | - | - | - | <6 |
| Missing | 0 | 98 | 13 | 103 | 0 | 0 | 0 | - | 7 | 221 |
| **Education** |  |  |  |  |  |  |  |  |  |  |
| No school-leaving qualification | 48 (0.3%) | - | 17 (1.0%) | 7 (0.2%) | <6 | - | - | - | 8 (1.9%) | 84 (0.4%) |
| GCSE (certification after 9 years) | 2300 (14.4%) | - | 345 (20.5%) | 265 (9.1%) | 93 (6.7%) | - | - | - | 190 (45.3%) | 3193 (14.3%) |
| GCSE (certification after 10 years) | 6925 (43.5%) | - | 514 (30.5%) | 674 (23.2%) | 344 (24.9%) | - | - | - | 125 (29.8%) | 8582 (38.5%) |
| A-levels (higher education) | 6645 (41.7%) | - | 786 (46.7%) | 1717 (59.1%) | 943 (68.1%) | - | - | - | 93 (22.2%) | 10184 (45.6%) |
| Other certification | 0 (0.0%) | - | 22 (1.3%) | 241 (8.3%) | 0 (0.0%) | - | - | - | <6 | 266 (1.2%) |
| Missing | 14 | - | 27 | 128 | 5 | - | - | - | 4 | 178 |
| **Employment** |  |  |  |  |  |  |  |  |  |  |
| Unemployed/seeking | 568 (3.6%) | - | 51 (3.0%) | 42 (1.5%) | 0 (0.0%) | - | - | - | <6 | 663 (3.1%) |
| Pensioner/ retired | 4772 (30.0%) | - | 435 (25.9%) | 1092 (38.5%) | 207 (27.9%) | - | - | - | 328 (78.1%) | 6834 (31.6%) |
| Employed (medical profession) | 1109 (7.0%) | - | 138 (8.2%) | 189 (6.7%) | 230 (31.0%) | - | - | - | 16 (3.8%) | 1682 (7.8%) |
| Employed (teacher/ childminder) | 650 (4.1%) | - | - | 122 (4.3%) | 120 (16.2%) | - | - | - | <6 | 894 (4.1%) |
| Employed (other) | 8416 (52.8%) | - | 967 (57.6%) | 1307 (46.1%) | 129 (17.4%) | - | - | - | 71 (16.9%) | 10890 (50.4%) |
| None of the above options | 417 (2.6%) | - | 89 (5.3%) | 81 (2.9%) | 57 (7.7%) | - | - | - | <6 | 645 (3.0%) |
| Missing | 0 | - | 31 | 199 | 646 | - | - | - | 3 | 879 |
| **Comorbidities** |  |  |  |  |  |  |  |  |  |  |
| Hypertension | 5089 (31.9%) | - | 842 (49.6%) | 923 (31.5%) | 358 (26.5%) | - | - | - | 275 (65.2%) | 7487 (33.5%) |
| Missing | 0 | - | 13 | 103 | 39 | - | - | - | 1 | 156 |
| Diabetes | 1684 (10.6%) | - | 148 (8.7%) | 176 (6.0%) | 53 (3.9%) | - | - | - | 116 (27.4%) | 2177 (9.7%) |
| Missing | 0 | - | 13 | 103 | 39 | - | - | - | 0 | 155 |
| Lung disease | 1943 (12.2%) | - | 256 (15.1%) | 227 (7.8%) | 138 (10.2%) | - | - | - | 89 (21.1%) | 2653 (11.9%) |
| Missing | 0 | - | 13 | 103 | 39 | - | - | - | 1 | 156 |
| Cardiovascular disease | 1272 (78.0%) | - | 406 (23.9%) | 290 (9.9%) | 123 (9.1%) | - | - | - | 211 (49.9%) | 2302 (10.3%) |
| Missing | 0 | - | 13 | 103 | 39 | - | - | - | 0 | 155 |
| Cancer (current or treated in the last year) | 947 (5.9%) | - | 61 (3.6%) | 61 (2.1%) | 30 (2.2%) | - | - | - | 115 (27.3%) | 1214 (5.4%) |
| Missing | 0 | - | 13 | 103 | 39 | - | - | - | 1 | 156 |
| Immunosuppression | 256 (1.6%) | - | 62 (3.7%) | 65 (2.2%) | 52 (3.9%) | - | - | 7 (3.8%) | 154 (37.7%) | 596 (2.6%) |
| Missing | 0 | - | 13 | 103 | 39 | - | - | 5 | 14 | 174 |
| **ER visit** |  | - |  |  |  | - | - |  |  |  |
| Yes | 2069 (13.0%) | - | 152 (10.2%) | 273 (9.0%) | 116 (8.6%) | - | - | 37 (20.1%) | 168 (39.8%) | 2815 (12.6%) |
| Not known | 0 (0.0%) | - | 0 (0.0%) | 171 (5.6%) | 0 (0.0%) | - | - |  | <6 | 174 (0.8%) |
| Missing | 42 | - | 223 | 0 | 39 | - | - | 5 | 1 | 310 |

\* Composed of samples in the payback panel and CATI. GCSE: General Certificate of Secondary Education; ER: Emergency room; ED: Emergency Department; Description of the participating studies (GUIDE; NAKO; STAAB; MuSPAD; ELISA; Wü-KITA-CoV; paedSAXCOVID; CorKID; Immunebridge\_ED) are given in the Supplement 1 TableS1.

### Combined endpoint for protection levels against SARS-CoV-2 infection and severe COVID-19

In the adult age groups, between 5% (of those over 79 years old) and 16% (of those 30-39 years old) of individuals had less than three confirmed exposures (Table 3, Figure 1A). Among persons over 79 years of age, 37% had less than four antibody-confirmed exposures (Table 3, Figure 1A). Among children and adolescents, 80% had less than three confirmed exposures, and 13% had neither a reported exposure nor an immunocorrelate. We found no relevant differences in the results when treating borderline antibody findings of the GUIDE study as seronegative (Supplement 1 Figure S6).

**Table 3:** Combined endpoint of infection, vaccination, and humoral immunity as well as proportion of antibodies detected against the S-antigen (S-AK) and the N-antigen (N-AK) stratified by age, sex, and pre-existing conditions. The results are age-standardised. Participants from IMMUNEBRIDGE_ED were excluded from this analysis as were participants with missing age information (n=132). The combined endpoint could not be formed for 12,185 participants due to missing information from the participants from NAKO (n=10,595) and for additional 1,590 participants from the other cohorts. Information on the S-AK and N-AK were not available for 1,517 and 1,503, respectively. (LCI: lower 95% confidence interval limit; UCI: upper 95% confidence interval limit).

|  | Combined endpoint |  |  |  |  |  |  |  |  |  |  |  | Antibodies |  |  |  |  |  |  |  |  |
| --- | --- | --- | --- | --- | --- | --- | --- | --- | --- | --- | --- | --- | --- | --- | --- | --- | --- | --- | --- | --- | --- |
|  |  | Three exposures +<br>infection or vaccination<br>in 2022 |  |  | Three exposures with<br>immunocorrelates |  |  | Three exposures without<br>immunocorrelates or less<br>than three exposures<br>(min. 1 exposure/<br>immunocorrelate) |  |  | No immunocorrelates<br>and no infection or<br>vaccination |  |  | S-AK |  |  |  | N-AK |  |  |  |
|  | N | Share (%) | LCI | UCI | Share (%) | LCI | UCI | Share (%) | LCI | UCI | Share (%) | LCI | UCI | N | Share (%) | LCI | UCI | N | Share (%) | LCI | UCI |
| Total | 20912 | 33.7 | 33.1 | 34.3 | 44.3 | 43.5 | 45.1 | 18.9 | 18.1 | 19.6 | 3.1 | 2.6 | 3.6 | 31566 | 94.8 | 94.2 | 95.4 | 31580 | 51.8 | 51.0 | 52.7 |
| Age (years) |  |  |  |  |  |  |  |  |  |  |  |  |  |  |  |  |  |  |  |  |  |
| 1-17 | 511 | 2.0 | 1.0 | 3.7 | 18.2 | 15.0 | 21.9 | 66.5 | 62.2 | 70.6 | 13.3 | 10.5 | 16.6 | 506 | 80.4 | 76.7 | 83.7 | 516 | 68.0 | 63.8 | 72.0 |
| 18-29 | 1522 | 39.7 | 37.2 | 42.2 | 45.9 | 43.3 | 48.4 | 13.4 | 11.8 | 15.2 | 1.1 | 0.6 | 1.7 | 1807 | 97.7 | 96.8 | 98.3 | 1809 | 59.7 | 57.3 | 61.9 |
| 30-34 | 1361 | 33.7 | 31.2 | 36.2 | 50.8 | 48.2 | 53.5 | 14.3 | 12.5 | 16.3 | 1.2 | 0.7 | 1.9 | 1914 | 97.0 | 96.1 | 97.7 | 1914 | 56.8 | 54.6 | 59.1 |
| 35-39 | 1402 | 34.4 | 31.9 | 36.9 | 49.9 | 47.3 | 52.6 | 14.8 | 13.0 | 16.8 | 0.9 | 0.5 | 1.6 | 1910 | 96.4 | 95.5 | 97.2 | 1910 | 56.8 | 54.5 | 59.0 |
| 40-49 | 2994 | 34.8 | 33.1 | 36.6 | 51.7 | 49.9 | 53.5 | 12.0 | 10.9 | 13.3 | 1.4 | 1.1 | 1.9 | 4586 | 96.8 | 96.2 | 97.3 | 4586 | 56.0 | 54.5 | 57.4 |
| 50-59 | 4906 | 32.1 | 30.8 | 33.4 | 58.4 | 57.0 | 59.8 | 8.2 | 7.5 | 9.0 | 1.2 | 1.0 | 1.6 | 8112 | 97.7 | 97.3 | 98.0 | 8112 | 48.4 | 47.3 | 49.4 |
| 60-64 | 2246 | 34.8 | 32.8 | 36.8 | 57.9 | 55.8 | 59.9 | 6.1 | 5.2 | 7.2 | 1.2 | 0.8 | 1.8 | 3728 | 98.0 | 97.5 | 98.5 | 3729 | 43.9 | 42.3 | 45.5 |
| 65-79 | 5300 | 49.7 | 48.4 | 51.1 | 45.2 | 43.9 | 46.6 | 4.1 | 3.6 | 4.7 | 0.9 | 0.7 | 1.2 | 8310 | 98.5 | 98.2 | 98.7 | 8311 | 38.1 | 37.1 | 39.2 |
| 80 and older | 670 | 62.5 | 58.7 | 66.2 | 32.5 | 29.0 | 36.3 | 4.6 | 3.2 | 6.6 | 0.3 | 0.1 | 1.2 | 693 | 99.1 | 98.0 | 99.6 | 693 | 28.3 | 25.0 | 31.8 |
| Sex |  |  |  |  |  |  |  |  |  |  |  |  |  |  |  |  |  |  |  |  |  |
| Female | 11123 | 33.5 | 32.5 | 34.5 | 45.3 | 44.1 | 46.4 | 18.3 | 17.0 | 19.5 | 3.0 | 2.3 | 3.7 | 16541 | 95.0 | 94.2 | 95.8 | 16550 | 51.1 | 49.9 | 52.3 |
| Male | 9788 | 33.9 | 32.9 | 35.0 | 43.3 | 42.0 | 44.5 | 19.6 | 18.2 | 20.9 | 3.2 | 2.4 | 4.0 | 14929 | 94.6 | 93.7 | 95.5 | 14934 | 52.7 | 51.4 | 53.9 |
| Comorbidities* |  |  |  |  |  |  |  |  |  |  |  |  |  |  |  |  |  |  |  |  |  |
| Cancer (current or<br>treated in the last year) | 1004 | 50.9 | 47.3 | 54.5 | 43.0 | 39.5 | 46.5 | 5.3 | 3.8 | 6.9 | 0.8 | 0.3 | 1.3 | 1006 | 97.9 | 97.1 | 98.7 | 1006 | 36.5 | 33.1 | 39.9 |
| Cardiovascular disease | 1885 | 53.8 | 51.1 | 56.4 | 41.0 | 38.4 | 43.6 | 4.7 | 3.6 | 5.8 | 0.6 | 0.3 | 0.9 | 1949 | 98.5 | 98.0 | 99.1 | 1949 | 36.9 | 34.4 | 39.4 |
| Diabetes | 1879 | 47.6 | 45.1 | 50.2 | 45.0 | 42.5 | 47.5 | 6.1 | 4.8 | 7.3 | 1.3 | 0.7 | 1.9 | 1904 | 97.7 | 97.0 | 98.5 | 1905 | 34.3 | 31.9 | 36.7 |
| Hypertension | 6621 | 46.4 | 45.1 | 47.7 | 47.3 | 46.0 | 48.6 | 5.5 | 4.9 | 6.2 | 0.8 | 0.5 | 1.0 | 6729 | 98.1 | 97.8 | 98.5 | 6730 | 38.1 | 36.8 | 39.4 |
| Immunosuppression | 414 | 47.5 | 42.2 | 52.8 | 47.7 | 42.5 | 53 | 4.4 | 2.3 | 6.5 | 0.4 | 0 | 0.9 | 415 | 81.9 | 67.6 | 96.1 | 415 | 36.3 | 31.3 | 41.3 |
| Lung disease | 2364 | 44.0 | 41.7 | 46.2 | 45.9 | 43.7 | 48.1 | 9.3 | 8.0 | 10.7 | 0.8 | 0.4 | 1.1 | 2393 | 97.6 | 97.0 | 98.3 | 2393 | 42.2 | 39.9 | 44.4 |
| First infections |  |  |  |  |  |  |  |  |  |  |  |  |  |  |  |  |  |  |  |  |  |
| No infection | 12790 | 19.6 | 18.9 | 20.3 | 59.1 | 58.0 | 60.2 | 15.9 | 14.8 | 17.1 | 5.4 | 4.5 | 6.2 | 12751 | 93.0 | 92.1 | 93.9 | 12762 | 24.8 | 23.6 | 26.0 |
| First infection 2020 | 444 | 33.6 | 28.2 | 39.0 | 42.3 | 36.5 | 48.2 | 24.1 | 18.4 | 29.8 | - | - | - | 443 | 98.0 | 96.6 | 99.4 | 443 | 70.6 | 65.2 | 75.9 |
| First infection 2021 | 921 | 30.0 | 26.5 | 33.5 | 36.0 | 32.0 | 40.1 | 33.9 | 29.5 | 38.4 | - | - | - | 912 | 98.0 | 97.0 | 98.9 | 912 | 76.3 | 72.9 | 79.7 |
| First infection 2022 | 6637 | 58.4 | 56.8 | 60.0 | 20.6 | 19.2 | 22.0 | 21.0 | 19.3 | 22.7 | - | - | - | 6613 | 95.6 | 94.6 | 96.7 | 6616 | 84.4 | 83.1 | 85.7 |
| Vaccinations |  |  |  |  |  |  |  |  |  |  |  |  |  |  |  |  |  |  |  |  |  |
| No vaccination | 1203 | 0.0 | 0.0 | 0.1 | 0.1 | 0.0 | 0.2 | 77.6 | 74.2 | 81.0 | 22.3 | 18.8 | 25.7 | 1173 | 63.4 | 59.2 | 67.7 | 1181 | 68.3 | 64.2 | 72.4 |
| One vaccination | 208 | 0.0 | 0.0 | 0.0 | 15.4 | 7.6 | 23.3 | 84.6 | 76.7 | 92.4 | - | - | - | 197 | 94.1 | 88.5 | 99.8 | 197 | 70.1 | 59.4 | 80.9 |
| Two vaccinations | 1715 | 7.1 | 5.7 | 8.4 | 35.0 | 32.3 | 37.7 | 58.0 | 55.2 | 60.8 | - | - | - | 1664 | 98.5 | 97.5 | 99.5 | 1667 | 55.8 | 52.8 | 58.7 |
| Three vaccinations | 14231 | 30.0 | 27.9 | 32.1 | 69.8 | 67.7 | 71.9 | 0.2 | 0.1 | 0.3 | - | - | - | 14231 | 99.8 | 99.7 | 99.9 | 14234 | 47.7 | 45.1 | 50.3 |
| Four vaccinations | 3554 | 96.0 | 94.6 | 97.3 | 3.6 | 2.3 | 4.9 | 0.4 | 0.0 | 0.9 | - | - | - | 3554 | 99.6 | 99.1 | 100.0 | 3554 | 30.5 | 27.5 | 33.5 |

\*only in ages ≥ 18years; S-AK: antibodies against S-antigen; N-AK: antibodies against nucleocapsid antigen

**Figure 1:**
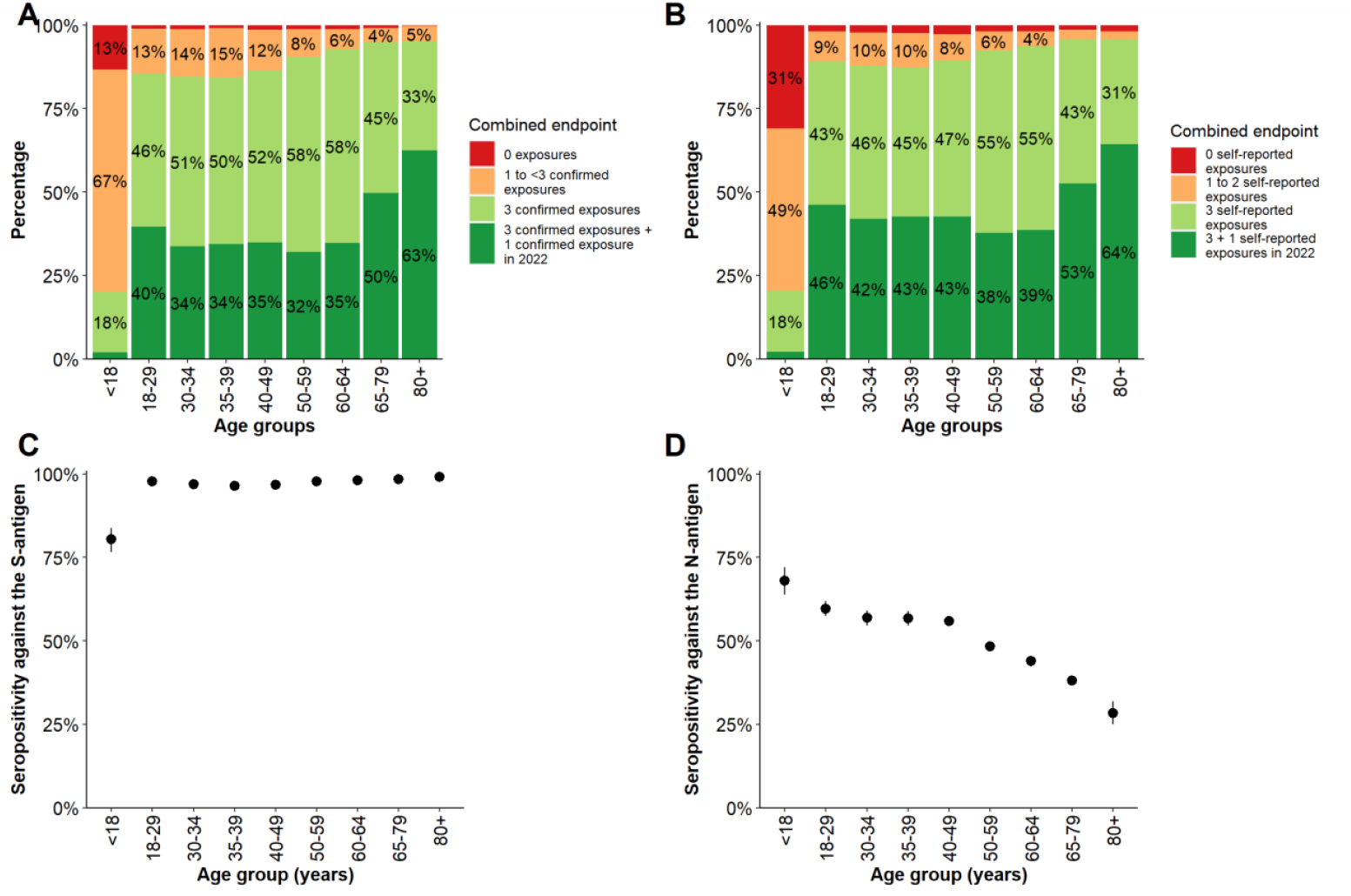
(A) The combined endpoint on the proportion of exposures by infection or vaccination with corresponding humoral immune response, stratified by age; for definition of combined endpoint see Table 1; (B) the combined endpoint of the proportion of exposures by infection or vaccination regardless of a humoral immune response, stratified by age; (C) seropositivity against the S-antigen and (D) against the N-antigen by age, with 95% confidence intervals. Participants from IMMUNEBRIDGE_ED were excluded from this analysis. The combined endpoint could not be formed for 12,185 participants due to missing information from the participants from NAKO (n=10,595) and for additional 1,590 participants from the other cohorts. Vaccination recommendations for children differed from those for adults, recommendations of > 1 dose only currently exist for those > 11 years old.

Among adults with self-reported comorbidities, 46%-56% (depending on the pre-existing comorbidity) had less than four confirmed exposures and 5%-10% had less than three confirmed exposures (Table 3).

When ignoring antibody results and building categories based on reported exposures only, the proportion of those with less than three exposures was lower in all age groups. However, the proportion of those with no reported exposure was higher, in particular in the age group under 18 years (Figure 1B). While seropositivity against the N-antigen decreased with older age (Figure 1D), this was not the case for seropositivity against the S-antigen, where a difference is only seen between children (80%) and adults (>95%; Figure 1C).

Figure 2A-F visualises the relative frequencies of the levels of the combined endpoint and seropositivity estimates on maps of Germany stratified by NUTS 2 units (usually German administrative districts). The combined endpoint is shown over time from June to September in Supplement 1 Figure S1, over NUTS-2 regions in Supplement 1 Figure S2, and over federal states in Supplement 1 Figure S3. The combined endpoint could not be formed for 12,185 participants due to missing information from the participants from NAKO (n=10,595) and for an additional 1,590 participants from the other cohorts. This was largely due to missing results from antibody tests due to difficulties in collecting blood for the DBS samples by the participants themselves and the resulting insufficient amount of sample material in the GUIDE study. Overall, between 0% and 12% of participants in all participating studies had missing laboratory results. In the GUIDE study, a higher proportion of missing lab results was seen among people in the older and very old age groups because of self-sampling. For between 0% and 13% of the study participants in the participating studies, information about vaccination and infection was missing.

**Figure 2.**
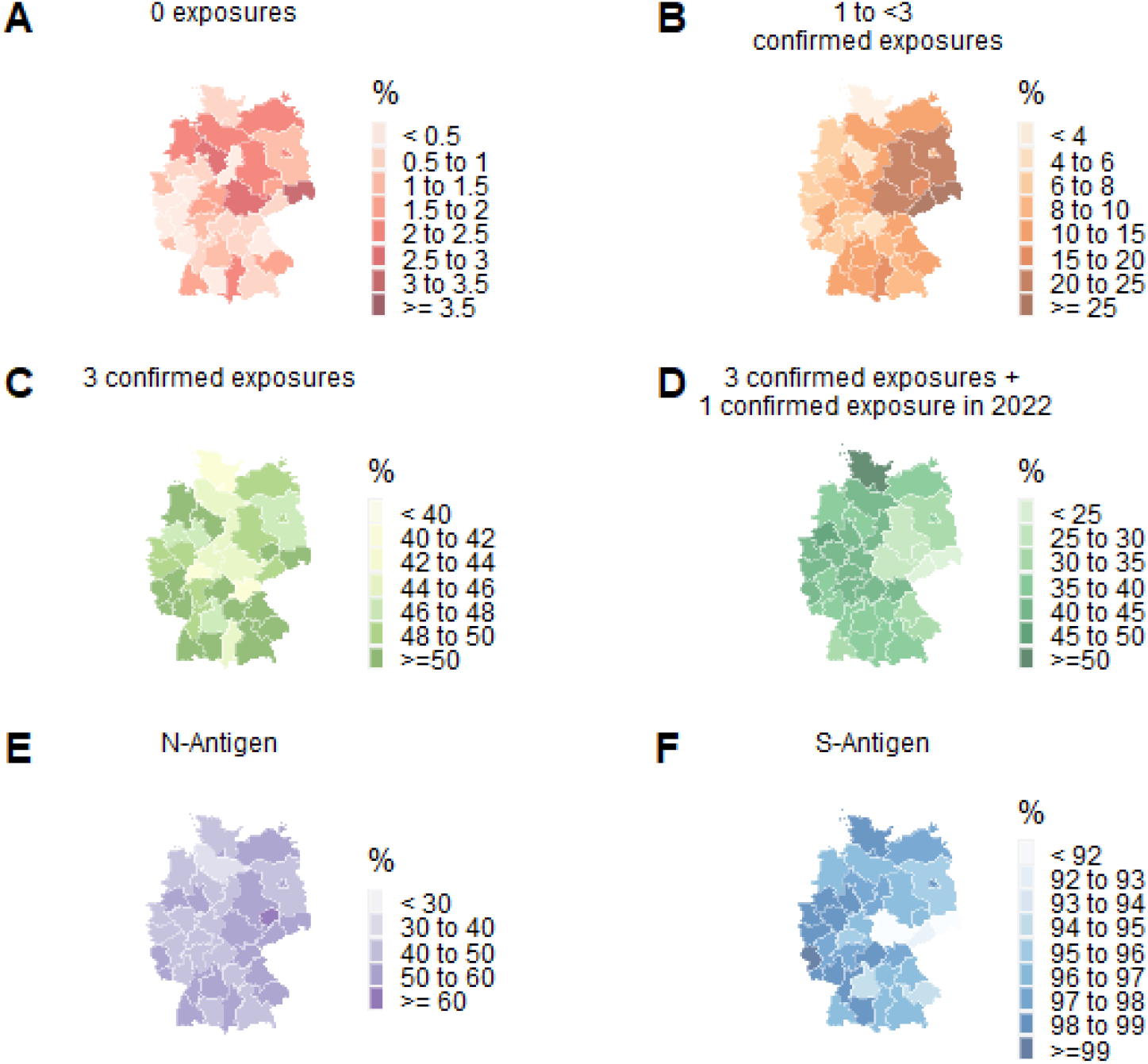
Age-standardised maps stratified by NUTS 2 region displaying (A-D) the combined endpoint of (A) no exposures, (B) 1 to <3 confirmed exposures, (C) 3 confirmed exposures, (D) 3 confirmed exposures and a confirmed exposure in 2022 and (E-F) seropositivity against (E) the S-antigen and (F) the N-antigen. Only data from the adult, population-based cross-sectional studies and cohorts (GUIDE, MuSPAD, NAKO, STAAB) are included in this analysis.

### Seroprevalence against the S- and N-antigen of SARS-CoV-2

Across all age groups, 95% of the study participants had antibodies against the S-antigen. In the age group 1-17, this proportion was lowest with 80% (Figure 1C; Table 3).

Across all age groups, 52% of the study participants had antibodies against the N-antigen. This proportion was highest in the age group 1-17 at 68% and lowest in people over 79 years of age at 28%; overall, there was a trend of decreasing proportions of antibodies against the N-antigen with increasing age (Figure 1D; Table 3). A lower proportion of older people (over 64 years of age) and people with comorbidities had antibodies against the N-antigen (Table 3). An antibody response against the N-antigen was present in 71% of participants who reported a first infection in 2020, in 76% of those who reported a first infection in 2021, and in 84% of those with a first reported infection in 2022 (Table 3). 25% of participants who reported no infection showed antibodies against the N-antigen (Table 3).

In participants reporting the first infection after being vaccinated at least two times, seropositivity for antibodies against the N-antigen was 89% up to 5 months after infection and dropped to 68% in those with reported infections longer than 5 months ago. This was similar albeit slightly less pronounced in persons without vaccination (91% up to 5 months and 76% after 5 months). This corresponded to markedly lower titre levels against the N-antigen in those participants of the GUIDE study with infections after at least two vaccinations compared to those with infections without vaccination and a higher decrease in antibody titres after 5 months amongst vaccinated persons.

### Regional heterogeneity

In relation to NUTS 2, the proportion of people with less than three confirmed exposures was highest in Dresden (28%) and lowest in Schleswig-Holstein (4%; Figure 2, Supplement 1 Figure S2). The proportion of people who had antibodies against the N-antigen ranged from 37% in Lüneburg to 60% in Leipzig (Figure 2).

### Comparison of data between population-based and hospital-based studies

In the hospital-based study IMMUNEBRIDGE_ED, the proportion of study participants who had less than three confirmed exposures (between 11% and 14%) was higher than that in the analogous strata in the population-based studies (between 4% and 7%; Figure 3).

**Figure 3:**
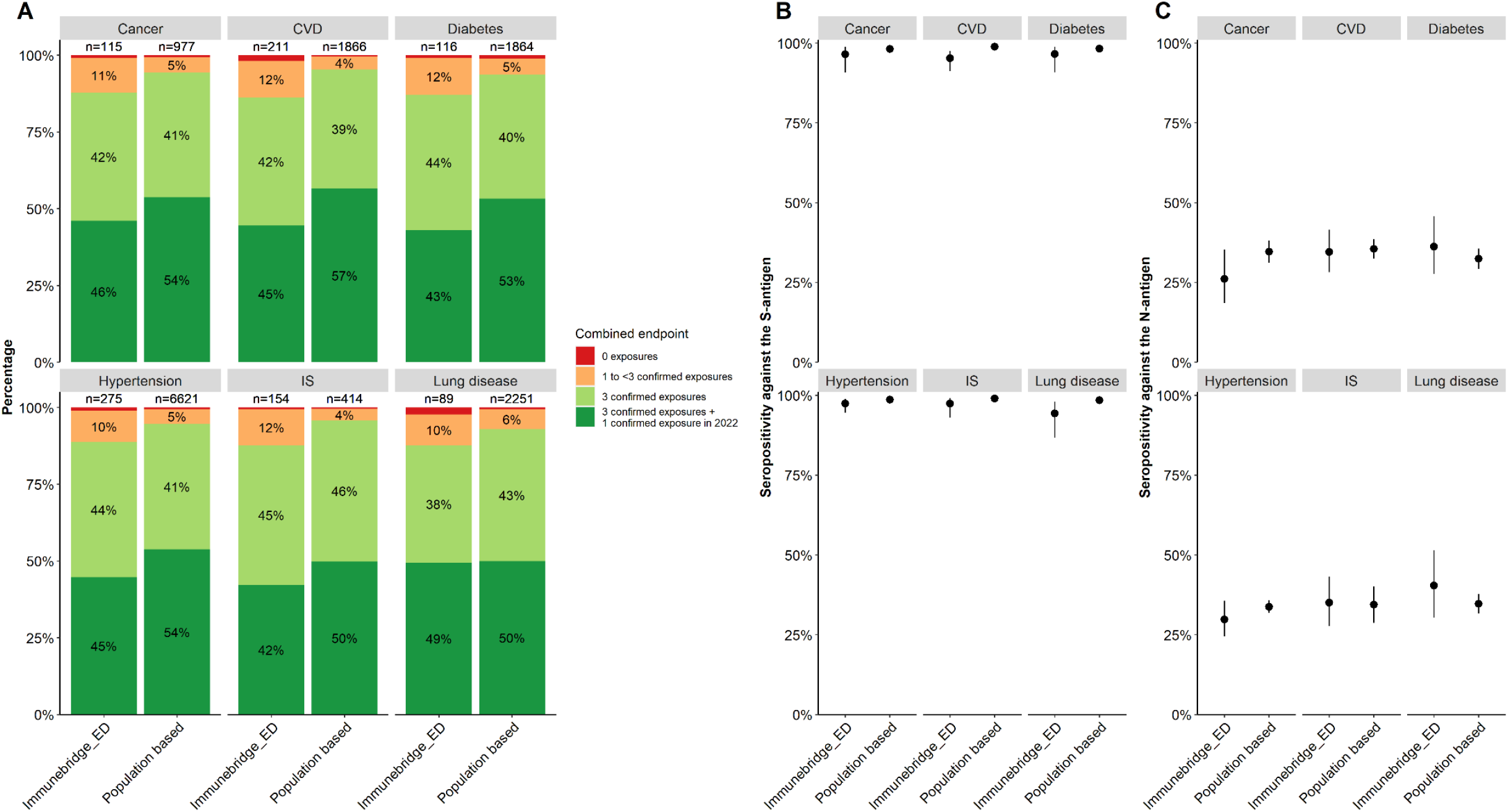
Comparison of (A) the combined endpoint and (B-C) seropositivity against the S- and N-antigen between the population-based studies and IMMUNEBRIDGE_ED in patients with cancer, cardiovascular disease (CVD), diabetes, hypertension, immunosuppression (IS), and lung disease. Post stratification weights based on the age-distribution in IMMUNEBRIDGE_ED comorbidity groups were applied to the population-based data.

Compared to the population-based studies, there were lower or similar proportions of study participants with antibodies against the S-antigen and against the N-antigen in IMMUNEBRIDGE_ED in the higher age groups and defined comorbidity groups (Figure 3).

## Discussion

We showed within the IMMUNEBRIDGE project that a network of existing cohort studies and newly established cross-sectional studies were able to successfully provide model-usable estimates of protection against infection and severe COVID-19 within two months of the start of sampling (first interim report in August 2022) and with high geographical resolution (up to NUTS-2). This was achieved by combining the advantages of established large population-based cohorts and newly designed cross-sectional studies, by using central data linkage structures and central laboratory infrastructures, and by collaboratively scaling up capacities for surveys and data analysis across various institutions in Germany. Continuous and rapid reporting of available results was achieved between August 2022 (12) and October 2022 (13) by direct communication to a newly established Modelling Network for Severe Infectious Diseases (MONID) (13). Different modelling groups simulated possible further pandemic courses for the 2022/23 winter based on the provided data (13, 21, 22).

By combining study results with the existing literature as well as the infection dynamics in the period of the survey, we noted that there was a moderate to high protection against severe COVID-19 (with the SARS-CoV-2 variant “Omicron BA.5” dominating in Germany at that time) in most age groups. Despite the high prevalence of antibodies against the S-antigen (95%) and N-antigen (52%) in the population, there was, however, low protection against infection, as confirmed by both the first BA.5 wave taking place when the surveys were performed as well as the second BA.5 wave in autumn 2022. Our results are in line with preliminary results from both the RKI-SOEP and the SeBluCo study based on data from February to March 2022 published in October and December 2022, and align with modelling results from the UK based on continuous population-based estimates available from large national population panels (10, 23, 24).

We identified gaps in protection against severe COVID-19 in people with pre-existing comorbidities as well as in certain population groups and in different regions of Germany. Among those with comorbidities, 49-56% had less than four confirmed exposures; 6-9% had less than three. The proportion of individuals with less than three confirmed exposures ranged from 4%-28% across the NUTS2 regions.

Analyses for vulnerable populations with severe comorbidities in IMMUNEBRDIGE_ED indicated that relevant gaps still exist in risk groups. The proportion of study participants who had less than three confirmed exposures was considerably higher in IMMUNEBRIDGE_ED than in the analogous strata in the population-based studies. This is most likely due to a combination of two phenomena: an accumulation of those persons with less protection in the emergency department and the fact that studies in the emergency department reach participants of those groups that population-based studies may find difficult to recruit. The extent to which each of these two phenomena explain the gap found here between population- and hospital-based results could be estimated in future modelling studies and would support efforts to understand generalisability of results from population-based studies to the German population (25).

In our results, there were also relevant differences between children or adolescents compared to adults. The clearly lower proportion of children (80%) with antibodies against the S-antigen results probably from a high proportion of unvaccinated children (62%). This reflects the different vaccination recommendation of the Standing Commission on Vaccination (STIKO) in this age group, recommending COVID-19 vaccinations initially only for adolescents and children with comorbidities (Supplement 1 Table S3). In contrast, the proportion of children with antibodies against the N-antigen following infection was 68%, i.e. the highest of all age groups (Table 3), although the proportion of children with self-reported infections (46%) was similar to the average of the adult age groups (Supplement 1 Table S4). The reasons for this discrepancy could be a higher proportion of oligo- or asymptomatic courses of infection in this age group, a longer lasting measurable immune response after infection, or an increased proportion of children compared to adults with an infection in more recent waves of the pandemic, especially during the circulation of the Omicron BA.5 variant.

Even if the proportion of persons with low protection initially seemed small compared to the majority of people with high protection, the infection of no more than 4% to 8% of the total population by the Wuhan and the delta variant of SARS-CoV-2 in the second and fourth wave of the pandemic at the end of 2020 and 2021 already led to a significant burden in the outpatient and inpatient care sector (10). This indicates that even small gaps of protection against severe COVID-19 may become relevant, in particular in light of waning immunity over the coming months and years (10, 21, 22).

Even though all included studies define a respective source population, they ultimately only provide a picture of the respective reference population with respect to predefined variables (e.g., age, sex, educational status). In addition, the under-recruitment of vaccination-averse groups across all studies may have led to an overestimation of protection. IMMUNEBRIDGE provides information on the presence of a humoral immune response against the S- and N-antigen of SARS-CoV-2, but not on the presence of neutralising activity or cellular immunity. However, such analyses were within the scope of some of the participating studies. There, an increasing activity of neutralising antibodies against the S-antigen for Wuhan and BA.5 variants could be detected with higher categories of the combined endpoint (26). The extent to which an antibody response is actually associated with a protective effect against infection or a severe course after infection is strongly dependent on the SARS-CoV-2 variant circulating. Our analysis also does not fully take into account that the probability of developing seropositivity to the N-antigen potentially depends on the vaccination history of the infected persons (27). Here, this may have led to an underestimation of the number of exposures in the group of vaccinated individuals.

In conclusion, we were able to quickly harmonise data collection, surveys, and data analysis across a network of nine existing and newly established population- and hospital-based studies to provide age-specific and regional estimates of protection against infection and severe COVID-19 in Germany. The combination of literature, data and infection dynamics during the survey period indicated moderate to high protection against severe COVID-19 with the BA.5 variant in most age groups, but low protection against infection in all age groups. The integration of this knowledge into current modelling studies that are required to reliably interpret the potential effects on infection dynamics was enabled by rapidly communicating preliminary results to a new modelling network for severe infectious diseases in Germany. Results of the IMMUNEBRIDGE project thus reflect the importance of networks covering a large number of scientific institutions in Germany when supported by relevant infrastructural and personnel resources.

## Data Availability

The aggregated data for this study will be made available to other academic researchers. The minimum dataset includes study site information, assay information, sample type, demographic information, self-administered diagnostic anamneses and lab results (NC, S Spike, IGRA and NAb). Institutions can apply for the data via. Data were provided to the modelling network for severe infectious diseases in Germany (MONID (13)).

## Statements

### Ethical statement

The responsible ethics committees of individual studies approved all study-related analyses: GUIDE: 202/22 approved by the Ethics Committee of the Medical Faculty of the Rheinische Friedrich-Wilhelm-University Bonn

ELISA: University of Lübeck (Az. 20-150)

NAKO: The study is continuously approved by the responsible local ethics committees of the German Federal States where all study centers are located in (original ethics approvals of the leading ethics committee of the Bayerische Landesärztekammer (protocol code 13023, Approval Date: 27 March 2013 and 14 February 2014 (rectification of documents, study protocol, consent form)). An external ethics advisory board has been established that accompanies NAKO over the full study period. A ‘Code of Ethics’ of NAKO (Ethikkodex) has been developed and the study is under steady surveillance by the ethics committees of the regional study centers (8).

STAAB: Ethics committee of the Medical Faculty of the University Würzburg (STAAB: #98/13) MuSPAD: Ethics committee of Hannover Medical School (9086_BO_S_2020 for MuSPAD), Dresden paedSAXCOVID: Ethics Committee of the Technische University (TU) Dresden (BO-EK-156042020).

Bochum CorKID: Ethics Committee of the Ruhr University Bochum (Nr. 20-6927_7) Würzburg Wü-KITa-CoV: Würzburg, Kennzeichen 105/21

IMMUNEBRIDGE_ED: Ethics Committee of University Medical Center Göttingen (21/6/22)

## Authors’ contributions

Conceptualisation: BL, AK, NT, AP, PH, HS, SB

Data quality: VKJ, MAK, MH, MJH, JO, VR

Formal analyses: VKJ, MJH, BL, AK, MH, AB, VR

Visualisation: VKJ, VR, AB

Project administration: BL, AK, CK, FB, NT, JL, SB, HS, PH

Laboratory analysis: AP, MN, GB, DR, KB, MF, TW, NT, FB, JL

Evidence synthesis: BL, MD, IvH, MB, OH

Writing-original draft: BL, VKJ, AK,

Funding acquisition: HS, SB, BL, AK

Writing-review & editing: All

All authors have read and agreed to the published version of the manuscript.

## Acknowledgements

The authors gratefully acknowledge all participants for their contribution, as well as the colleagues for data collection, in particular BOS112 and Heike Adam for the lab management, as well as Barbora Kessel for MuSPAD data management and Timo Ludwig for STAAB data management. We thank all participants who took part in the NAKO study and the staff of this research initiative. We thank the Federal State of Saxony for supporting the SchoolCoviDD19 (SchoolCoviDD19 (paedSAXCOVID) by a financial grant. We thank the Bavarian State Ministry of Health and Care/ State Office for Health and Food Safety for supporting Wü-KiTa-CoV by a financial grant. We thank Josephine Schneider for her excellent organisation of the SchoolCoviDD19 (paedSAXCOVID) study.

## Key public health message

In the IMMUNEBRIDGE project, a consortium of researchers and institutions harmonised data collection and data linkage of new surveys conducted between June and November 2022 across nine existing and newly established studies. The aim was to provide rapid estimates of the level of protection against COVID-19 in the German population that could be used in a newly established network of modelling groups for predictions of health care burden for the winter season 2022/23.

Researchers divided study participants into four categories based on whether persons reported vaccinations and/or infections of SARS-CoV-2 and whether these vaccinations or infections were confirmed with antibody assessments against N- and S-antigens in blood samples of participants. Different levels of protection against severe course of disease or infection were assumed according to these categories. The highest category comprised participants who had experienced four exposure events (i.e. vaccinations or infections), with at least one episode occurring during the year 2022. The lowest category comprised participants who had experienced no respective exposure event and exhibited no antibodies against the S- or N-antigen.

Results show that the majority of the population had moderate to high protection level against severe COVID-19 with age-related differences. Children had the lowest proportion of seropositivity against the S-protein, but the highest seropositivity against the N-protein, suggesting that protection in this age group has been established by infection during the Omicron BA.5 wave rather than by immunisation. There were relevant gaps in protection against severe COVID-19 in people with pre-existing comorbidities as well as in certain population groups and regions. Protection levels were highly variable across different regions in Germany.

Results were transferred rapidly to a new central modelling network and could be used in harmonised efforts from this network to communicate potential health care burden for the winter season 2022/23 to the public and decision makers. Overall, we showed how a network of existing epidemiological studies can serve as a crystallisation point for a rapid-response platform providing urgently needed public health estimates.

## References

1. Beermann S, Dörr M, Grill E, Karch A, Lange B, Zeeb H. Coronapandemie: Die Rolle epidemiologischer Forschung in Gesundheitskrisen. 2022;119(17):753–6.

2. Brinkmann F, Diebner HH, Matenar C, Schlegtendal A, Spiecker J, Eitner L, et al. Longitudinal Rise in Seroprevalence of SARS-CoV-2 Infections in Children in Western Germany—A Blind Spot in Epidemiology? Infectious Disease Reports. 2021;13(4):957–64.

3. Engels G, Hecker K, Forster J, Toepfner N, Hick E, Pietsch F, et al. High Seroprevalence of SARS-CoV-2 in Preschool Children in July 2022. Dtsch Arztebl Int. 2022;119(45):771–772.

4. Gornyk D, Harries M, Glöckner S, Strengert M, Kerrinnes T, Heise J-K, et al. SARS-CoV-2 Seroprevalence in Germany: A Population-Based Sequential Study in Seven Regions. Deutsches ärzteblatt international. 2021;118(48):824.

5. Eichner FA, Gelbrich G, Weißbrich B, Dölken L, Kurzai O, Deckert J, et al. Seroprävalenz von COVID-19 und psychosoziale Auswirkungen in der Allgemeinbevölkerung: Ergebnisse des STAAB-COVID-One Programms. 2021;83(12):965–75.

6. Streeck H. Grundimmunität gegen SARS-CoV-2 (COVID-19) in der Bevölkerung (GUIDE). 2022.

7. Klein C, Borsche M, Balck A, Föh B, Rahmöller J, Peters E, et al. One-year surveillance of SARS-CoV-2 transmission of the ELISA cohort: A model for population-based monitoring of infection risk. Science advances. 2022;8(15):eabm5016.

8. Peters A, Greiser KH, Goettlicher S, Ahrens W, Albrecht M, Bamberg F, et al. Framework and baseline examination of the German National Cohort (NAKO). European Journal of Epidemiology. 2022:1–18.

9. Neuhauser H, Buttmann-Schweiger N, Ellert U, Fiebig J, Hövener C, Offergeld R, et al. Seroepidemiologische Studien zu SARS-CoV-2 in Stichproben der Allgemeinbevölkerung und bei Blutspenderinnen und Blutspendern in Deutschland–Ergebnisse bis August 2021. 2021.

10. Antao E-M, Jung-Sedzik T, Buda S, Haas W, Diercke M, Schumacher J, et al. COVID-19-Pandemie: Surveillance und Studien des Robert Koch-Instituts zur Lage-und Maßnahmenbewertung. 2022.

11. Berndt JC, T. Hasenauer, J, Karch, A, Kheifetz, Y, Kirsten, H, et al. A. Szenarien für den Verlauf der SARS-CoV-2-Pandemie im Winter 2022/23 - Ergebnisse eines Workshops des Modellierungsnetzes für schwere Infektionskrankheiten (Modellierungsnetz). Zenodo 2022.

12. Lange B, Jäger, V, Rücker, V, Hassenstein, MJ., Harries, M, Berner, R, et al. A. Interimsanalyse des IMMUNEBRIDGE-Projektes zur Kommunikation von vorläufigen Ergebnissen an die Modellierungskonsortien der BMBF-geförderten Modellierungsplattform. preprint. 2022.

13. Lange B, Jäger, V, Rücker, V, Harries, M, Hassenstein, MJ., Dreier, M, et al. 2. Interimsanalyse des IMMUNEBRIDGE-Projektes zur Kommunikation von vorläufigen Ergebnissen an das Modellierungsnetz für schwere Infektionskrankheiten. 2022.

14. Balck A, Föh B, Borsche M, Rahmöller J, Vollstedt E-J, Waldeck F, et al. Protocol of the Luebeck longitudinal investigation of SARS-CoV-2 infection (ELISA) study–a prospective population-based cohort study. BMC Public Health. 2022;22(1):1–9.

15. Von Elm E, Altman DG, Egger M, Pocock SJ, Gøtzsche PC, Vandenbroucke JP, et al. The Strengthening the Reporting of Observational Studies in Epidemiology (STROBE) Statement: guidelines for reporting observational studies. 2014;12(12):1495–9.

16. Krause G. The German National Cohort: aims, study design and organisation. 2014.

17. Söllner R, Körner T. Der Registerzensus: Ziele, Anforderungen und Umsetzungsansätze. WISTA-Wirtschaft und Statistik. 2022;74(4):13–24.

18. Lahti L, Huovari J, Kainu M, Biecek P. Retrieval and Analysis of Eurostat Open Data with the eurostat Package. R J. 2017;9(1):385.

19. Commission E. Eurostat. TERCET Flat Files 2021 [Available from: https://gisco-services.ec.europa.eu/tercet/flat-files.

20. R Core Team R. R: A language and environment for statistical computing. R foundation for statistical computing Vienna, Austria; 2018.

21. Ärzteblatt D. Kleeblattprinzip: 93 Corona-intensivpatienten verlegt, das Konzept funktioniert: Deutsches Ärzteblatt; 2021 [Available from: https://www.aerzteblatt.de/nachrichten/129956/Kleeblattprinzip-93-Coronaintensivpatienten-verlegt-das-Konzept-funktioniert.

22. Faensen D, Claus H, Benzler J, Ammon A, Pfoch T, Breuer T, et al. SurvNet@ RKI–a multistate electronic reporting system for communicable diseases. Eurosurveillance. 2006;11(4):7–8.

23. Barnard RC, Davies NG, Jit M, Edmunds WJJNc. Modelling the medium-term dynamics of SARS-CoV-2 transmission in England in the Omicron era. 2022;13(1):1–15.

24. RKI. Corona-Monitoring bundesweit – Welle 2 (aktualisierte Version vom 14.12.2022). 2022.

25. Kuss O, Becher H, Wienke A, Ittermann T, Ostrzinski S, Schipf S, et al. Statistical Analysis in the German National Cohort (NAKO)–Specific Aspects and General Recommendations. European Journal of Epidemiology. 2022:1–8.

26. Harries M, Jäger, V, Rodiah, I, Hassenstein, MJ, Ortmann J, Dreier M, et al. Bridging the gap_Estimation of 2022/2023 SARS-CoV-2 healthcare burden in Germany based on multidimensional data from a rapid epidemic panel. medRxiv. 2022.

27. Follmann D, Janes HE, Buhule OD, Zhou H, Girard B, Marks K, et al. Antinucleocapsid antibodies after SARS-CoV-2 infection in the blinded phase of the randomised, placebo-controlled mRNA-1273 COVID-19 vaccine efficacy clinical trial. Annals of Internal Medicine. 2022;175(9):1258–65.

