## Supplement 1 for "Estimates of protection against SARS-CoV-2 infection and severe COVID-19 in Germany before the 2022/2023 winter season - the IMMUNEBRIDGE project"

### Supplementary material 1: Additional information on data collection and combined endpoint results

Figure S1: Combined endpoint on the proportion of exposures by survey month and age
Figure S2: Combined endpoint on the proportion of exposures by NUTS (age-standardised)
Figure S3: Combined endpoint on the proportion of exposures by federal states (age-standardised)
Figure S4: Combined endpoint on the proportion of exposures by NUTS (not age-standardised)
Figure S5: Combined endpoint on the proportion of exposures by federal states (not age-standardised)
Figure S6: Combined endpoint on the proportion of exposures: sensitivity analysis

Table S1: Description of the studies participating in IMMUNEBRIDGE
Table S2: Data collection instruments and infrastructures
Table S3: Vaccination status of participants stratified by age, sex, and pre-existing conditions and age-standardised
Table S4: Participants' self-reported first infections in 2020, 2021, and 2022 stratified by age, sex, and pre-existing conditions and age-standardised
Table S5: Combined endpoint of infection, vaccination, and humoral immunity as well as proportion of antibodies detected against the S-antigen (S-AK) and the N-antigen (N-AK) stratified by age, sex, and pre-existing conditions (not age-standardised)
Table S6: Vaccination status of participants stratified by age, sex, and pre-existing conditions (not age-standardised)
Table S7: Participants' self-reported first infections in 2020, 2021, and 2022 stratified by age, sex, and pre-existing conditions (not age-standardised)

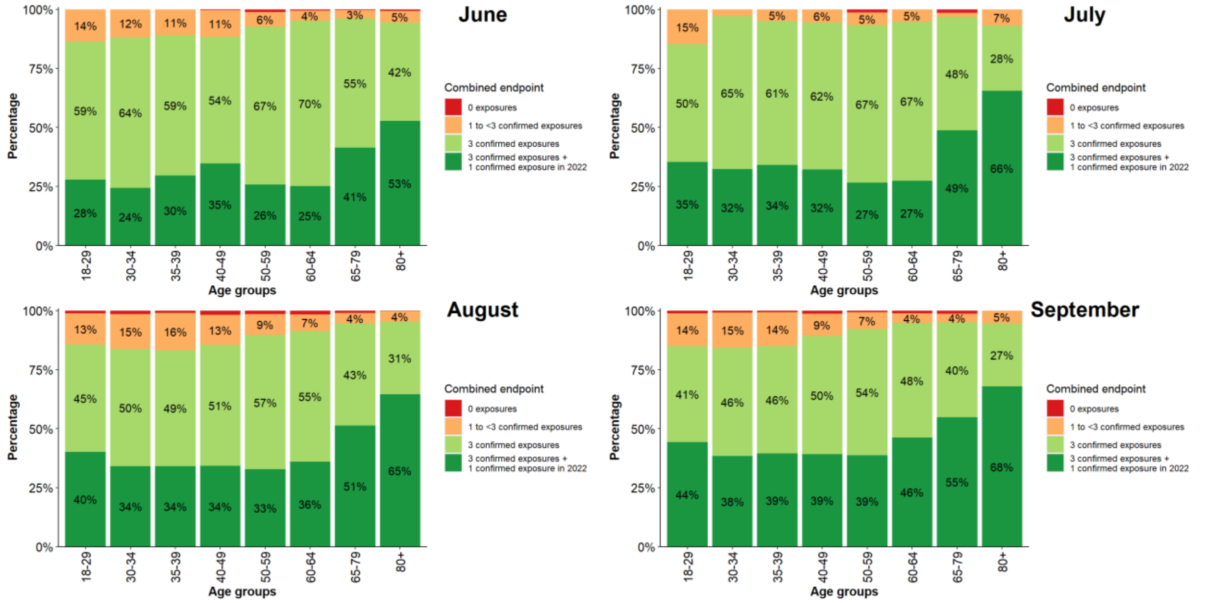


Supplement 1 Figure S1: Combined endpoint on the proportion of exposures through infection or vaccination with corresponding humoral immune response stratified by survey month and age (≥ 18 years); for the definition of endpoints see Table 1 in the main manuscript.

### Regional analysis of the combined endpoints

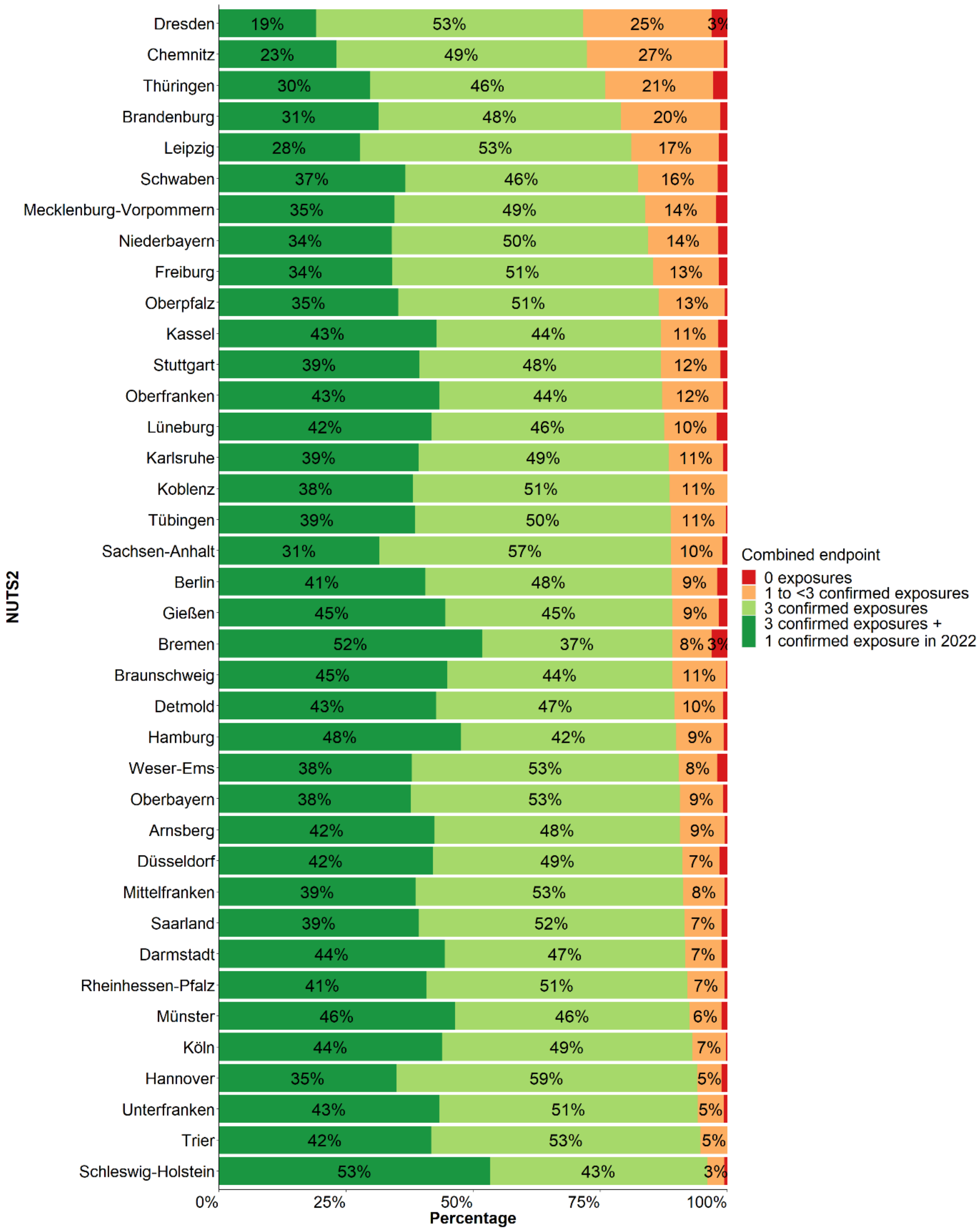


Supplement 1 Figure S2: Combined endpoint on the proportion of exposures through infection or vaccination with corresponding humoral immune response stratified by NUTS-2 and age-standardised (≥18 years).

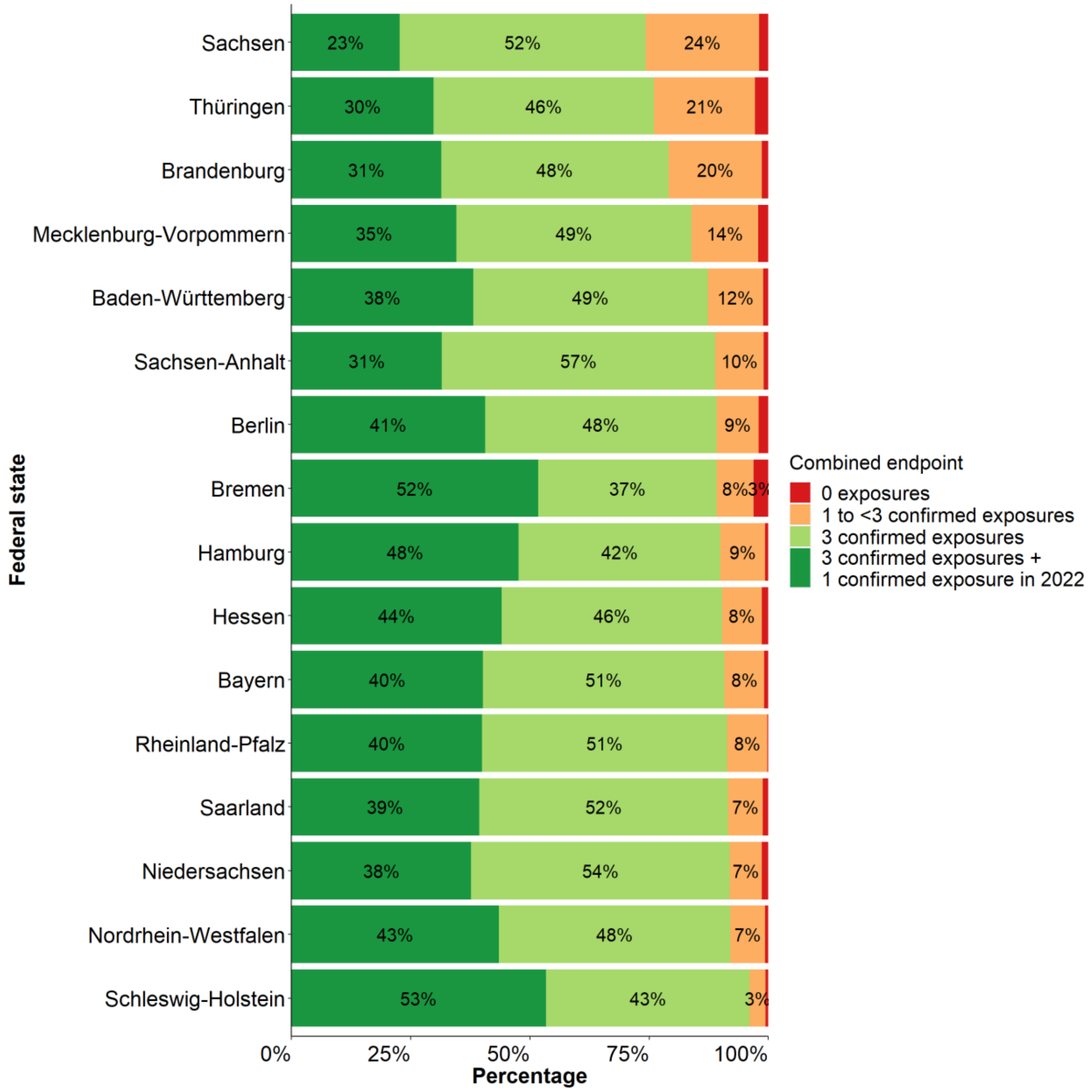


Supplement 1 Figure S3: Combined endpoint on the proportion of exposures through infection or vaccination with corresponding humoral immune response stratified by federal state and age-standardised (≥18 years).

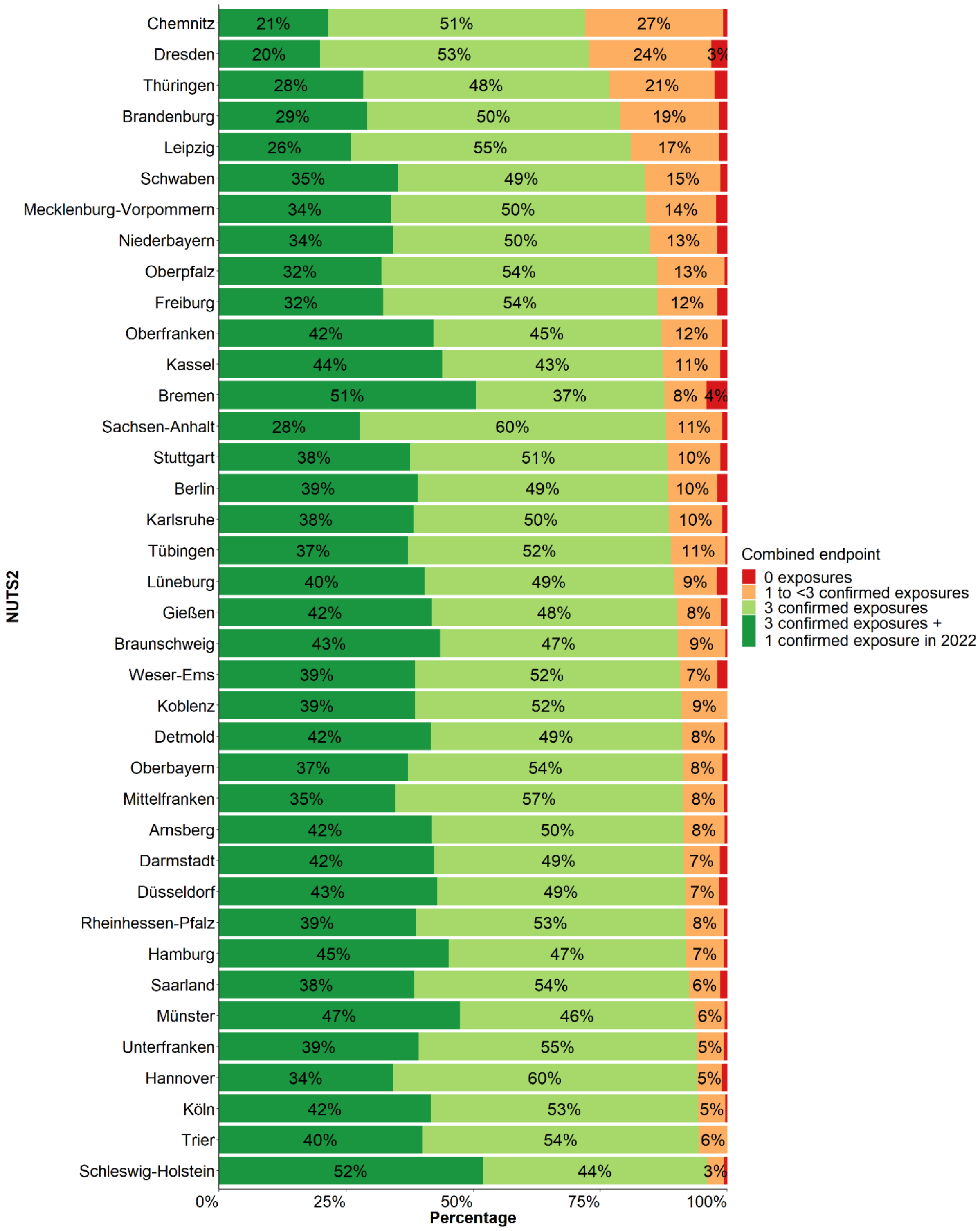


Supplement 1 Figure S4: Combined endpoint on the proportion of exposures through infection or vaccination with corresponding humoral immune response stratified by NUTS-2 (not age-standardised; ≥18 years).

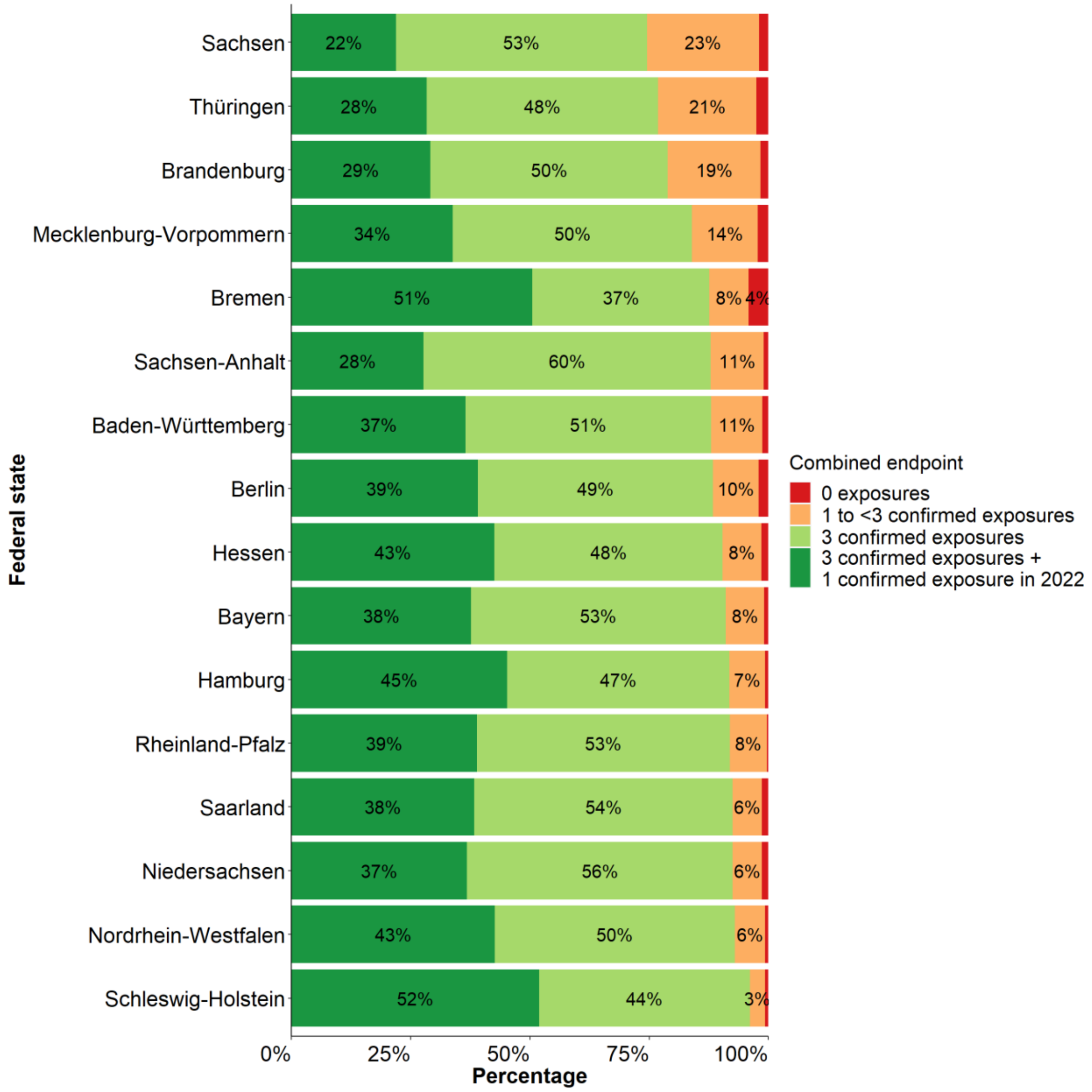


Supplement 1 Figure S5: Combined endpoint on the proportion of exposures through infection or vaccination with corresponding humoral immune response stratified by federal state (not age-standardised; ≥18 years).

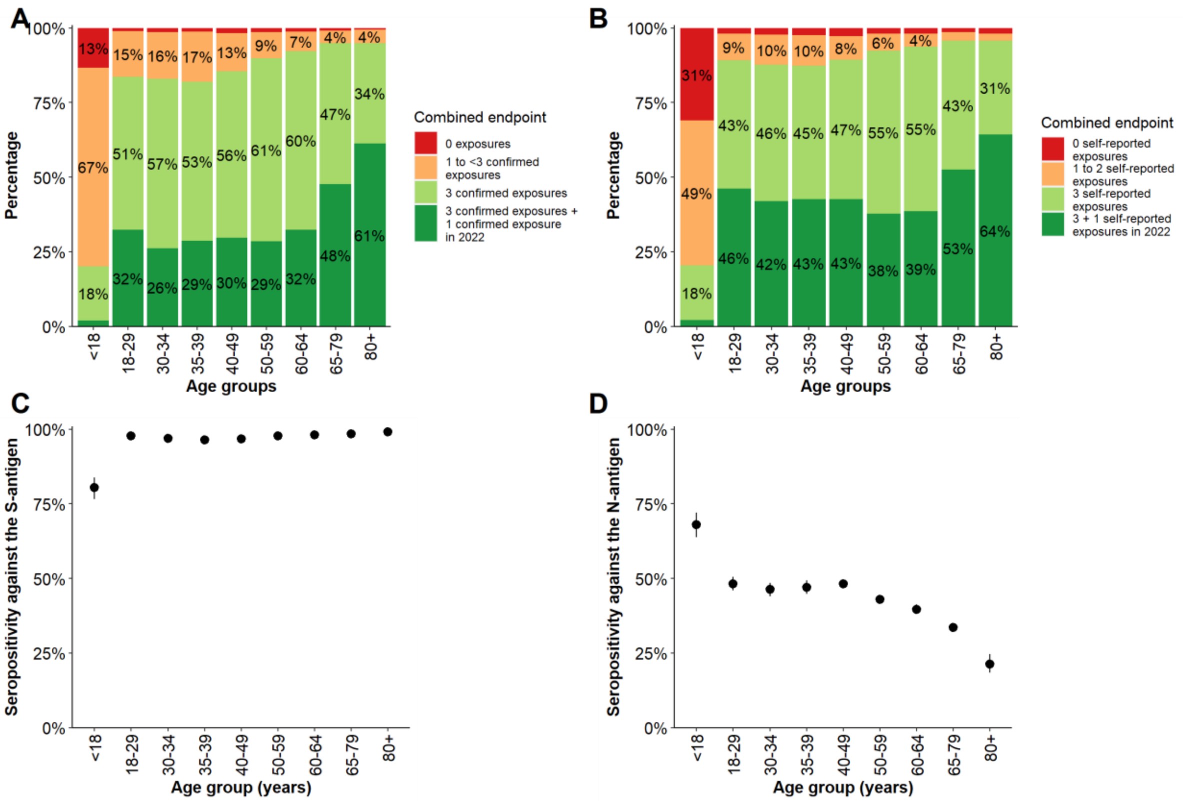


Supplement 1 Figure S6: Sensitivity analysis treating borderline N-antigen findings of the GUIDE study as seronegative (A) The combined endpoint on the proportion of exposures by infection or vaccination with corresponding humoral immune response, stratified by age (B) the combined endpoint of the proportion of exposures by infection or vaccination regardless of a humoral immune response, stratified by age; (C) seropositivity against the S-antigen and (D) against the N-antigen by age, with 95% confidence intervals.

Participants from IMMUNEBRIDGE_ED were excluded from this analysis. The combined endpoint could not be formed for 12,185 participants due to missing information from the participants from NAKO (n=10,595) and for additional 1,590 participants from the other cohorts. Vaccination recommendations for children differed from those for adults, recommendations of > 1 dose only currently exist for those > 11 years old.

Supplement 1 Table S1: Description of the studies participating in IMMUNEBRIDGE

| | Financing of data collection* | Description of the cohort | Cohort type | Region | N | Existing lab parameters | Existing data items | Restrictions |
|---|---|---|---|---|---|---|---|---|
| **Population-based cohorts (18 years and older)** | | | | | | | | |
| NAKO | Federal Ministry of Education and Research, federal statues, Helmholtz Association | 10.1007/s10654-014-9890-7 | Population-based; recruitment 2014-2019 via population registration offices | 18 locations nationwide | 10595 | N-AK, S-AK | Age, gender | Age at baseline 20-69 years |
| MuSPAD | Helmholtz Association, RESPINOW (Federal Ministry of Education and Research), own funds | 10.3238/arztebl.m2021.0364; https://hzi-c19-antikoerperstudie.de/teilnehmerunterlagen/ | Population-based; recruitment 2020 via population registration offices | Magdeburg | 1046 | N-AK, S-AK, S-IGRA | MDS** present, previous findings from 2 surveys | ≥18 |
| MuSPAD | Helmholtz Association, RESPINOW (Federal Ministry of Education and Research), own funds | 10.3238/arztebl.m2021.0364; https://hzi-c19-antikoerperstudie.de/teilnehmerunterlagen/ | Population-based; recruitment 2020 via population registration offices | Aachen | 870 | N-AK, S-AK, S-IGRA | MDS** present, previous findings from 2 surveys | ≥18 |
| MuSPAD | Helmholtz Association, IMMUNEBRIDGE (Federal Ministry of Education and Research), own funds | 10.3238/arztebl.m2021.0364; https://hzi-c19-antikoerperstudie.de/teilnehmerunterlagen/ | Population-based; recruitment 2020 via population registration offices | Hanover | 1116 | N-AK, S-AK | MDS** present, previous findings from 1 survey | ≥18 |
| STAAB | Federal Ministry of Education and Research, Bavarian State Ministry of Science and the Arts | 10.1177/2047487316680693 | Population-based; recruitment baseline 2013-2017 via population registration offices | Würzburg | 1711 | N-AK, S-AK | MDS** present, previous findings from 2 surveys | Age at baseline 30-79 years |
| ELISA | IMMUNEBRIDGE (Federal Ministry of Education and Research) | https://www.ncbi.nlm.nih.gov/pmc/articles/PMC9012459/pdf/sciadv.abm5016.pdf https://science.org/doi/10.1126/sciadv.abm5016 | Population-based; recruitment 2020 via population registration offices | Lübeck | 1389 | N-AK, S-AK | MDS** present | ≥18 |
| **Cross-sectional studies on children and populations with risk factors for a severe course of SARS-CoV-2 infection** | | | | | | | | |
| Dresden paedSAXCOVID | Funding by the Free State of Saxony "paedSaxCoviDD | 10.1136/bmjpo-2021-001036, 10.1007/s15010-022-01824-9 | School-based | Dresden | 91 | N-AK, S-AK | MDS** present | 12-17 |
| Würzburg Wü-KITa-CoV | Bavarian State Ministry of Health and Care/ State Office for Health and Food Safety | https://jamanetwork.com/journals/jamanetworkopen/fullarticle/2787578 https://jamanetwork.com/journals/jamanetworkopen/fullarticle/2796275 | Based on day care centres | Würzburg | 275 | N-AK, S-AK | MDS** present | 1-6 |
| Bochum CorKID | IMMUNEBRIDGE (Federal Ministry of Education and Research) | 10.3390/idr13040088 10.1001/jamanetworkopen.2021.42057 | Participating paediatric practices, recruitment July-August 2022 | Ruhr area | 189 | N-AK, S-AK | MDS** present | 1-17 |
| IMMUNEBRIDGE_ED | Own resources | - | Patients from the emergency room | Göttingen | 423 | N-AK, S-AK | MDS** present | ≥18 |
| **Cross-sectional study of the general population in Germany (UK Bonn)** | | | | | | | | |
| GUIDE (Payback + CATI) | IMMUNEBRIDGE (Federal Ministry of Education and Research) | https://www.drks.de/drks_web/navigate.do?navigationId=trial.HTML&TRIAL_ID=DRKS00029693 | Random draw from participants of the Payback Panel (online) + CATI survey (telephone) of persons older than 65 years of age | Germany-wide | 15932 | N-AK, S-AK (DBS) | MDS** present | ≥65 |
| **Total** | | | | | 33637 | | | |

*In addition, the central laboratory analysis in Oldenburg and Greifswald in the population cohort studies was financed via the IMMUNEBRIDGE project; **MDS = Minimal data set

Supplement 1 Table S2: Data collection instruments and infrastructures

| Characteristics needed | Definition/ Conditions | Survey infrastructures needed | N IMMUNEBRIDGE studies meeting this characteristic | (Network) infrastructures to ensure this | How Immunebridge met this characteristic |
|---|---|---|---|---|---|
| Population-based design | | Ability to work with population registers | 5 (ELISA,GUIDE, MuSPAD, NAKO, STAAB) | - | - |
| Longitudinal design | Scheduled follow-ups or ability to recontact and reinvite participants | | 4 (ELISA, MuSPAD, NAKO, STAAB) | - | - |
| Supra-regional design | Within surveys (not always needed) | Study centre ability in several locations | 4 (GUIDE, MuSPAD, NAKO, STAAB) | Geographic depths of the network either ensured or able to pull in additional sources on short notice | Set up additional cross sectional survey (GUIDE) to gain geographic depth |
| Speed | Survey preparation within 1-2 months | Logistics need to be set up to ensure short preparation time | 3 (MuSPAD, NAKO, STAAB) | Overview of studies and their potential preparation times needs to be available | Close consultation with the individual cohorts from the planning phase onwards |
| | Data linkage within 1-2 weeks | Ability to integrate minimal data sets quickly into current surveys | 8 (all) | Data linkage capacity and tool needs to be available | Relevant staff capacities of several research institutions were planned for this from the beginning |
| | Data analysis of aggregate estimates within 1-2 weeks | | 8 (all) | Data analysis capacity needs to be available | Relevant staff capacities of several research institutions were planned for this from the beginning |
| | Ensuring capacity to be speedy in set up of surveys and network proceedings by regular use (min 1/year) | Regular use of cohort for different purposes and pathogens | 4 (ELISA, MuSPAD, NAKO, STAAB) | Regular (min 1/year) use of network | Part of the surveys performed were already as part of the regular use of epidemic panels (e.g. MuSPAD within RESPINOW) |
| Adaptability | Of the survey to different pathogens (not needed in IB) | Different sampling methods need to be available (e.g., blood, swab, stool) | 3 (MuSPAD, NAKO, STAAB) | Network needs to be able to ensure that different sampling methods across studies are available | Did not need to be met, as only one pathogen was assessed |
| Linkage of survey and sampling across the network | Comparability of used biomarker assays | Potential to adapt to central laboratory infrastructure | 9 (all) | Central laboratory infrastructures, including IT linkage to ensure comparability | Central laboratory structure experience from NAKO and NUM-NAPKON was used here |
| | Quick set-up of minimal data sets | Generic minimal data sets implemented in individual studies | 9 (all) | Generic minimal data sets available within the network | A minimal data set was set up at the start of the project |
| Data linkage with other relevant sources | Surveillance data | | perspective | - | - |
| | Hospital data | | perspective | - | - |
| | Ability to link to clinical cohorts | | perspective | Either the network comprises clinical cohorts or has close collaboration with some | A clinical cohort as comparison (emergency department) was established |

Supplement 1 Table S3: Vaccination status of participants stratified by age, sex, and pre-existing conditions and age-standardised. Participants from IMMUNEBRIDGE_ED were excluded from this analysis. Information on the vaccination status of NAKO participants (n=10,595) and 262 participants from the other cohorts is unknown and are therefore not included in this analysis. (LCI: lower 95% confidence interval limit; UCI: upper 95% confidence interval limit)

| | | No vaccination | | | First dose | | | Second dose | | | Third dose | | | Fourth dose | | |
|---|---|---|---|---|---|---|---|---|---|---|---|---|---|---|---|---|
| | N | Share (%) | LCI | UCI | Share (%) | LCI | UCI | Share (%) | LCI | UCI | Share (%) | LCI | UCI | Share (%) | LCI | UCI |
| **Total** | 22225 | 14.0 | 13.4 | 14.7 | 1.4 | 1.1 | 1.7 | 11.5 | 10.8 | 12.2 | 58.8 | 58.1 | 59.5 | 14.3 | 13.9 | 14.7 |
| **Age (years)** | | | | | | | | | | | | | | | | |
| 1-17 | 534 | 61.8 | 57.5 | 65.9 | 3.4 | 2.1 | 5.4 | 26.6 | 22.9 | 30.6 | 8.2 | 6.1 | 11.0 | 0 | 0 | 8.8 |
| 18-29 | 1617 | 5.3 | 4.3 | 6.6 | 1.4 | 0.9 | 2.1 | 14.7 | 13.0 | 16.5 | 75.7 | 73.5 | 77.8 | 3.0 | 2.2 | 4.0 |
| 30-34 | 1430 | 6.8 | 5.6 | 8.2 | 1.3 | 0.8 | 2.0 | 13.0 | 11.3 | 14.9 | 75.9 | 73.6 | 78.1 | 3.0 | 2.2 | 4.1 |
| 35-39 | 1478 | 7.2 | 5.9 | 8.6 | 1.4 | 0.9 | 2.1 | 13.3 | 11.6 | 15.1 | 73.4 | 71.1 | 75.6 | 4.8 | 3.8 | 6.1 |
| 40-49 | 3138 | 6.7 | 5.9 | 7.7 | 1.4 | 1.0 | 1.9 | 9.6 | 8.6 | 10.7 | 76.1 | 74.6 | 77.6 | 6.2 | 5.4 | 7.1 |
| 50-59 | 5158 | 4.2 | 3.7 | 4.8 | 0.8 | 0.6 | 1.1 | 7.3 | 6.6 | 8.0 | 78.0 | 76.8 | 79.1 | 9.8 | 9.0 | 10.6 |
| 60-64 | 2364 | 3.1 | 2.5 | 3.9 | 0.7 | 0.4 | 1.1 | 5.6 | 4.7 | 6.6 | 74.3 | 72.5 | 76.0 | 16.3 | 14.9 | 17.9 |
| 65-79 | 5739 | 2.0 | 1.7 | 2.4 | 0.5 | 0.3 | 0.7 | 3.4 | 2.9 | 3.9 | 56.5 | 55.3 | 57.8 | 37.6 | 36.4 | 38.9 |
| 80 and older | 767 | 2.1 | 1.2 | 3.4 | 0.8 | 0.3 | 1.8 | 2.5 | 1.5 | 3.9 | 34.7 | 31.3 | 38.2 | 60.0 | 56.4 | 63.5 |
| **Sex** | | | | | | | | | | | | | | | | |
| Female | 11736 | 13.4 | 12.3 | 14.6 | 1.5 | 1.1 | 2.0 | 11.5 | 10.5 | 12.5 | 60.4 | 59.2 | 61.5 | 13.2 | 12.6 | 13.8 |
| Male | 10488 | 14.7 | 13.5 | 16.0 | 1.2 | 0.9 | 1.6 | 11.5 | 10.4 | 12.5 | 57.1 | 55.9 | 58.3 | 15.6 | 14.9 | 16.3 |
| **Comorbidities*** | | | | | | | | | | | | | | | | |
| Cancer (current or treated in the last year) | 1092 | 2.2 | 1.2 | 3.1 | 0.4 | 0.1 | 0.8 | 4.8 | 3.4 | 6.2 | 55.6 | 52.1 | 59.0 | 37.1 | 33.7 | 40.4 |
| Cardiovascular disease | 2014 | 1.8 | 1.3 | 2.4 | 0.5 | 0.1 | 0.8 | 3.8 | 2.9 | 4.8 | 52.0 | 49.4 | 54.5 | 41.9 | 39.4 | 44.5 |
| Diabetes | 2028 | 3.2 | 2.4 | 4.1 | 0.5 | 0.2 | 0.8 | 4.8 | 3.7 | 5.8 | 55.8 | 53.4 | 58.3 | 35.7 | 33.3 | 38.1 |
| Hypertension | 7082 | 2.6 | 2.2 | 3.1 | 0.7 | 0.5 | 0.9 | 5.1 | 4.5 | 5.6 | 61.6 | 60.4 | 62.9 | 30.0 | 28.8 | 31.2 |
| Immunosuppression | 434 | 1.4 | 0.3 | 2.5 | 0.6 | 0.1 | 1.3 | 4.9 | 2.7 | 7.1 | 57.9 | 52.8 | 63.1 | 35.2 | 30.2 | 40.2 |
| Lung disease | 2520 | 3.2 | 2.5 | 3.9 | 0.9 | 0.5 | 1.2 | 8.6 | 7.3 | 9.8 | 64.1 | 62.0 | 66.2 | 23.2 | 21.4 | 25.1 |

*only in the ≥ 18years

Supplement 1 Table S4: Proportion of participants' self-reported first infections in 2020, 2021, and 2022 stratified by age, sex, and pre-existing conditions and age-standardised. Participants from IMMUNEBRIDGE_ED were excluded from this analysis. Information on the date of self-reported infections of NAKO participants (n=10,595) and 365 participants from the other cohorts is unknown; these are therefore not included in this analysis. (LCI: lower 95% confidence interval limit; UCI: upper 95% confidence interval limit)

| | | No reported infection | | | First reported infection 2020 | | | First reported infection 2021 | | | First reported infection 2022 | | |
|---|---|---|---|---|---|---|---|---|---|---|---|---|---|
| | N | Share (%) | LCI | UCI | Share (%) | LCI | UCI | Share (%) | LCI | UCI | Share (%) | LCI | UCI |
| **Total** | 22122 | 58.6 | 57.7 | 59.5 | 2.1 | 1.9 | 2.3 | 4.5 | 4.1 | 4.9 | 34.8 | 33.9 | 35.7 |
| **Age (years)** | | | | | | | | | | | | | |
| 1-17 | 516 | 54.3 | 49.9 | 58.6 | 1.2 | 0.5 | 2.6 | 3.5 | 2.1 | 5.6 | 41.1 | 36.8 | 45.5 |
| 18-29 | 1616 | 41.8 | 39.4 | 44.3 | 3.3 | 2.5 | 4.4 | 6.4 | 5.3 | 7.7 | 48.5 | 46.0 | 50.9 |
| 30-34 | 1429 | 45.6 | 43.0 | 48.3 | 3.2 | 2.3 | 4.2 | 6.0 | 4.9 | 7.4 | 45.2 | 42.6 | 47.8 |
| 35-39 | 1476 | 46.5 | 43.9 | 49.1 | 2.3 | 1.6 | 3.2 | 6.4 | 5.3 | 7.8 | 44.8 | 42.2 | 47.4 |
| 40-49 | 3125 | 49.2 | 47.4 | 51.0 | 2.7 | 2.1 | 3.3 | 5.9 | 5.1 | 6.8 | 42.3 | 40.5 | 44.0 |
| 50-59 | 5118 | 62.1 | 60.8 | 63.4 | 2.2 | 1.8 | 2.6 | 4.5 | 4.0 | 5.1 | 31.2 | 30.0 | 32.5 |
| 60-64 | 2356 | 69.1 | 67.1 | 70.9 | 2.1 | 1.6 | 2.8 | 3.4 | 2.8 | 4.3 | 25.4 | 23.7 | 27.2 |
| 65-79 | 5717 | 76.1 | 74.9 | 77.2 | 1.4 | 1.2 | 1.8 | 2.6 | 2.2 | 3.1 | 19.9 | 18.9 | 21.0 |
| 80 and older | 769 | 83.1 | 80.2 | 85.6 | 1.2 | 0.6 | 2.3 | 3.0 | 2.0 | 4.5 | 12.7 | 10.5 | 15.4 |
| **Sex** | | | | | | | | | | | | | |
| female | 11670 | 58.5 | 57.2 | 59.8 | 1.9 | 1.6 | 2.2 | 4.2 | 3.7 | 4.7 | 35.5 | 34.2 | 36.7 |
| male | 10451 | 58.7 | 57.3 | 60.0 | 2.3 | 2.0 | 2.7 | 4.9 | 4.3 | 5.5 | 34.1 | 32.8 | 35.5 |
| **Antibodies** | | | | | | | | | | | | | |
| N-AB positive | 9084 | 30.0 | 28.6 | 31.5 | 3.0 | 2.6 | 3.5 | 7.0 | 6.3 | 7.7 | 60.0 | 58.5 | 61.5 |
| N-AB negative | 11646 | 85.6 | 84.8 | 86.4 | 1.3 | 1.1 | 1.5 | 2.2 | 1.9 | 2.5 | 11.0 | 10.2 | 11.7 |
| S-AB positive | 20038 | 57.7 | 56.8 | 58.7 | 2.2 | 1.9 | 2.5 | 4.7 | 4.3 | 5.1 | 35.4 | 34.5 | 36.3 |
| S-AB negative | 678 | 72.4 | 67.6 | 77.2 | 0.8 | 0.2 | 1.3 | 1.8 | 1.0 | 2.5 | 25.1 | 20.4 | 29.8 |
| **Comorbidities*** | | | | | | | | | | | | | |
| Cancer (current or treated in the last year) | 1091 | 70.9 | 67.9 | 73.9 | 1.8 | 1.1 | 2.5 | 3.5 | 2.3 | 4.7 | 23.8 | 21.0 | 26.7 |
| Cardiovascular disease | 2001 | 74.1 | 71.9 | 76.3 | 2.2 | 1.5 | 2.9 | 3.1 | 2.2 | 4.0 | 20.6 | 18.6 | 22.6 |
| Diabetes | 2028 | 72.4 | 70.3 | 74.5 | 1.6 | 1.0 | 2.3 | 5.1 | 4.0 | 6.3 | 20.9 | 19.0 | 22.7 |
| Hypertension | 7055 | 69.1 | 68.0 | 70.3 | 2.0 | 1.7 | 2.4 | 4.1 | 3.6 | 4.7 | 24.7 | 23.6 | 25.8 |
| Immunosuppression | 429 | 71.4 | 66.9 | 76.0 | 2.1 | 0.8 | 3.3 | 3.2 | 1.5 | 4.9 | 23.3 | 19.1 | 27.6 |
| Lung disease | 2507 | 61.5 | 59.4 | 63.6 | 2.9 | 2.1 | 3.7 | 4.2 | 3.4 | 5.1 | 31.4 | 29.4 | 33.4 |

*only in the ≥ 18years

Supplement 1 Table S4: Combined endpoint of infection, vaccination, and humoral immunity as well as proportion of antibodies detected against the S-antigen (S-AK) and the N-antigen (N-AK) stratified by age, sex, and pre-existing conditions. The results are not age-standardised. Participants from IMMUNEBRIDGE_ED were excluded from this analysis as were participants with missing age information. The combined endpoint could not be formed for 12,185 participants due to missing information from the participants from NAKO (n=10,595) and for additional 1,590 participants from the other cohorts. (LCI: lower 95% confidence interval limit; UCI: upper 95% confidence interval limit).

| | Combined endpoint | | | | | | | | | | | | | Antibodies | | | | | | | |
|---|---|---|---|---|---|---|---|---|---|---|---|---|---|---|---|---|---|---|---|---|---|
| | | Three exposures + infection or vaccination in 2022 | | | Three exposures with immunocorrelates | | | Three exposures without immunocorrelates or less than three exposures (min. 1 exposure/ immunocorrelate) | | | No immunocorrelates and no infection or vaccination | | | S-AK | | | | N-AK | | | |
| | N | Share (%) | LCI | UCI | Share (%) | LCI | UCI | Share (%) | LCI | UCI | Share (%) | LCI | UCI | N | Share (%) | LCI | UCI | N | Share (%) | LCI | UCI |
| **Total** | 20912 | 38.3 | 37.6 | 38.9 | 50.3 | 49.6 | 50.9 | 10.0 | 9.6 | 10.4 | 1.4 | 1.3 | 1.6 | 31566 | 97.4 | 97.3 | 97.6 | 31580 | 47.8 | 47.3 | 48.3 |
| **Age (years)** | | | | | | | | | | | | | | | | | | | | | |
| 1-17 | 511 | 2.0 | 1.0 | 3.7 | 18.2 | 15.0 | 21.9 | 66.5 | 62.2 | 70.6 | 13.3 | 10.5 | 16.6 | 506 | 80.4 | 76.7 | 83.7 | 516 | 68.0 | 63.8 | 72.0 |
| 18-29 | 1522 | 39.7 | 37.2 | 42.2 | 45.9 | 43.3 | 48.4 | 13.4 | 11.8 | 15.2 | 1.1 | 0.6 | 1.7 | 1807 | 97.7 | 96.8 | 98.3 | 1809 | 59.7 | 57.3 | 61.9 |
| 30-34 | 1361 | 33.7 | 31.2 | 36.2 | 50.8 | 48.2 | 53.5 | 14.3 | 12.5 | 16.3 | 1.2 | 0.7 | 1.9 | 1914 | 97.0 | 96.1 | 97.7 | 1914 | 56.8 | 54.6 | 59.1 |
| 35-39 | 1402 | 34.4 | 31.9 | 36.9 | 49.9 | 47.3 | 52.6 | 14.8 | 13.0 | 16.8 | 0.9 | 0.5 | 1.6 | 1910 | 96.4 | 95.5 | 97.2 | 1910 | 56.8 | 54.5 | 59.0 |
| 40-49 | 2994 | 34.8 | 33.1 | 36.6 | 51.7 | 49.9 | 53.5 | 12.0 | 10.9 | 13.3 | 1.4 | 1.1 | 1.9 | 4586 | 96.8 | 96.2 | 97.3 | 4586 | 56.0 | 54.5 | 57.4 |
| 50-59 | 4906 | 32.1 | 30.8 | 33.4 | 58.4 | 57.0 | 59.8 | 8.2 | 7.5 | 9.0 | 1.2 | 1.0 | 1.6 | 8112 | 97.7 | 97.3 | 98.0 | 8112 | 48.4 | 47.3 | 49.4 |
| 60-64 | 2246 | 34.8 | 32.8 | 36.8 | 57.9 | 55.8 | 59.9 | 6.1 | 5.2 | 7.2 | 1.2 | 0.8 | 1.8 | 3728 | 98.0 | 97.5 | 98.5 | 3729 | 43.9 | 42.3 | 45.5 |
| 65-79 | 5300 | 49.7 | 48.4 | 51.1 | 45.2 | 43.9 | 46.6 | 4.1 | 3.6 | 4.7 | 0.9 | 0.7 | 1.2 | 8310 | 98.5 | 98.2 | 98.7 | 8311 | 38.1 | 37.1 | 39.2 |
| 80 and older | 670 | 62.5 | 58.7 | 66.2 | 32.5 | 29.0 | 36.3 | 4.6 | 3.2 | 6.6 | 0.3 | 0.1 | 1.2 | 693 | 99.1 | 98.0 | 99.6 | 693 | 28.3 | 25.0 | 31.8 |
| **Sex** | | | | | | | | | | | | | | | | | | | | | |
| Female | 11123 | 37.9 | 37.0 | 38.8 | 50.7 | 49.8 | 51.6 | 9.9 | 9.3 | 10.4 | 1.5 | 1.3 | 1.7 | 16541 | 97.4 | 97.1 | 97.6 | 16550 | 46.7 | 45.9 | 47.4 |
| Male | 9788 | 38.7 | 37.7 | 39.6 | 49.8 | 48.8 | 50.8 | 10.2 | 9.6 | 10.8 | 1.3 | 1.1 | 1.5 | 14929 | 97.5 | 97.2 | 97.7 | 14934 | 49.0 | 48.2 | 49.8 |
| **Comorbidities*** | | | | | | | | | | | | | | | | | | | | | |
| Cancer (current or treated in the last year) | 1004 | 49.2 | 46.1 | 52.3 | 44.4 | 41.4 | 47.5 | 5.5 | 4.1 | 6.9 | 0.9 | 0.3 | 1.5 | 1006 | 97.5 | 96.6 | 98.5 | 1006 | 37.1 | 34.1 | 40.1 |
| Cardiovascular disease | 1885 | 50.8 | 48.5 | 53.0 | 43.6 | 41.4 | 45.8 | 4.9 | 4.0 | 5.9 | 0.7 | 0.3 | 1.1 | 1949 | 98.3 | 97.7 | 98.9 | 1949 | 37.5 | 35.4 | 39.7 |
| Diabetes | 1879 | 45.8 | 43.6 | 48.1 | 47.3 | 45.0 | 49.5 | 5.7 | 4.6 | 6.7 | 1.2 | 0.7 | 1.7 | 1904 | 97.7 | 97.1 | 98.4 | 1905 | 34.1 | 31.9 | 36.2 |
| Hypertension | 6621 | 44.6 | 43.4 | 45.8 | 49.4 | 48.2 | 50.6 | 5.3 | 4.7 | 5.8 | 0.8 | 0.6 | 1.0 | 6729 | 98.1 | 97.8 | 98.5 | 6730 | 38.9 | 37.8 | 40.1 |
| Immunosuppression | 414 | 48.8 | 44.0 | 53.6 | 46.4 | 41.6 | 51.2 | 4.3 | 2.4 | 6.3 | 0.5 | 0.0 | 1.2 | 415 | 98.8 | 97.7 | 99.8 | 415 | 36.9 | 32.2 | 41.5 |
| Lung disease | 2364 | 43.0 | 41.0 | 45.0 | 47.6 | 45.6 | 49.6 | 8.5 | 7.3 | 9.6 | 0.9 | 0.5 | 1.3 | 2393 | 97.5 | 96.9 | 98.2 | 2393 | 41.0 | 39.0 | 42.9 |
| **First infections** | | | | | | | | | | | | | | | | | | | | | |
| No infection | 12790 | 22.7 | 22.0 | 23.4 | 68.5 | 67.7 | 69.2 | 7.0 | 6.6 | 7.4 | 1.8 | 1.6 | 2.0 | 12751 | 96.6 | 96.3 | 96.9 | 12762 | 21.4 | 20.7 | 22.1 |
| First infection 2020 | 444 | 36.3 | 31.8 | 40.8 | 43.2 | 38.5 | 47.8 | 20.5 | 16.8 | 24.3 | - | - | - | 443 | 98.0 | 96.7 | 99.3 | 443 | 65.5 | 61.0 | 69.9 |
| First infection 2021 | 921 | 33.8 | 30.7 | 36.9 | 38.1 | 34.9 | 41.3 | 28.1 | 25.2 | 31.1 | - | - | - | 912 | 97.5 | 96.5 | 98.5 | 912 | 71.1 | 68.1 | 74.0 |
| First infection 2022 | 6637 | 72.5 | 71.4 | 73.6 | 19.3 | 18.3 | 20.3 | 8.2 | 7.5 | 8.9 | - | - | - | 6613 | 96.8 | 96.4 | 97.3 | 6616 | 81.8 | 80.9 | 82.7 |
| **Vaccinations** | | | | | | | | | | | | | | | | | | | | | |
| No vaccination | 1203 | 0.1 | 0.0 | 0.3 | 0.3 | 0.0 | 0.7 | 74.1 | 71.3 | 77.0 | 25.4 | 22.6 | 28.3 | 1173 | 49.0 | 46.2 | 51.8 | 1181 | 61.4 | 58.6 | 64.2 |
| One vaccination | 208 | 0.0 | 0.0 | 0.0 | 12.6 | 7.9 | 17.3 | 87.4 | 82.7 | 92.1 | - | - | - | 197 | 92.4 | 88.7 | 96.1 | 197 | 64.5 | 57.8 | 71.2 |
| Two vaccinations | 1715 | 9.0 | 7.6 | 10.4 | 35.1 | 32.8 | 37.4 | 55.9 | 53.4 | 58.3 | - | - | - | 1664 | 98.8 | 98.3 | 99.3 | 1667 | 55.4 | 53.0 | 57.7 |
| Three vaccinations | 14231 | 31.1 | 30.3 | 31.8 | 68.7 | 67.9 | 69.4 | 0.3 | 0.2 | 0.3 | - | - | - | 14231 | 99.7 | 99.7 | 99.8 | 14234 | 45.0 | 44.2 | 45.9 |
| Four vaccinations | 3554 | 97.0 | 96.4 | 97.5 | 2.7 | 2.2 | 3.2 | 0.3 | 0.1 | 0.5 | - | - | - | 3554 | 99.7 | 99.5 | 99.9 | 3554 | 28.0 | 26.6 | 29.5 |

*only in ages ≥ 18years; S-AK: antibodies against S-antigen; N-AK: antibodies against nucleocapsid antigen

Supplement 1 Table S5: Vaccination status of participants stratified by age, sex, and pre-existing conditions. The results are not age-standardised. Participants from IMMUNEBRIDGE_ED were excluded from this analysis as were participants with missing age information. Information on the vaccination status of NAKO participants (n=10,595) and 262 participants from the other cohorts is unknown and are therefore not included in this analysis. (LCI: lower 95% confidence interval limit; UCI: upper 95% confidence interval limit)

| | | No vaccination | | | First dose | | | Second dose | | | Third dose | | | Fourth dose | | |
|---|---|---|---|---|---|---|---|---|---|---|---|---|---|---|---|---|
| | N | Share (%) | LCI | UCI | Share (%) | LCI | UCI | Share (%) | LCI | UCI | Share (%) | LCI | UCI | Share (%) | LCI | UCI |
| **Total** | 22225 | 5.6 | 5.4 | 5.9 | 1.0 | 0.8 | 1.1 | 8.0 | 7.7 | 8.4 | 68.0 | 67.4 | 68.6 | 17.4 | 16.9 | 17.8 |
| **Age (years)** | | | | | | | | | | | | | | | | |
| 1-17 | 534 | 61.8 | 57.5 | 65.9 | 3.4 | 2.1 | 5.4 | 26.6 | 22.9 | 30.6 | 8.2 | 6.1 | 11.0 | 0 | 0 | 8.8 |
| 18-29 | 1617 | 5.3 | 4.3 | 6.6 | 1.4 | 0.9 | 2.1 | 14.7 | 13.0 | 16.5 | 75.7 | 73.5 | 77.8 | 3.0 | 2.2 | 4.0 |
| 30-34 | 1430 | 6.8 | 5.6 | 8.2 | 1.3 | 0.8 | 2.0 | 13.0 | 11.3 | 14.9 | 75.9 | 73.6 | 78.1 | 3.0 | 2.2 | 4.1 |
| 35-39 | 1478 | 7.2 | 5.9 | 8.6 | 1.4 | 0.9 | 2.1 | 13.3 | 11.6 | 15.1 | 73.4 | 71.1 | 75.6 | 4.8 | 3.8 | 6.1 |
| 40-49 | 3138 | 6.7 | 5.9 | 7.7 | 1.4 | 1.0 | 1.9 | 9.6 | 8.6 | 10.7 | 76.1 | 74.6 | 77.6 | 6.2 | 5.4 | 7.1 |
| 50-59 | 5158 | 4.2 | 3.7 | 4.8 | 0.8 | 0.6 | 1.1 | 7.3 | 6.6 | 8.0 | 78.0 | 76.8 | 79.1 | 9.8 | 9.0 | 10.6 |
| 60-64 | 2364 | 3.1 | 2.5 | 3.9 | 0.7 | 0.4 | 1.1 | 5.6 | 4.7 | 6.6 | 74.3 | 72.5 | 76.0 | 16.3 | 14.9 | 17.9 |
| 65-79 | 5739 | 2.0 | 1.7 | 2.4 | 0.5 | 0.3 | 0.7 | 3.4 | 2.9 | 3.9 | 56.5 | 55.3 | 57.8 | 37.6 | 36.4 | 38.9 |
| 80 and older | 767 | 2.1 | 1.2 | 3.4 | 0.8 | 0.3 | 1.8 | 2.5 | 1.5 | 3.9 | 34.7 | 31.3 | 38.2 | 60.0 | 56.4 | 63.5 |
| **Sex** | | | | | | | | | | | | | | | | |
| Female | 11736 | 5.8 | 5.4 | 6.2 | 1.0 | 0.8 | 1.2 | 7.9 | 7.4 | 8.4 | 69.0 | 68.2 | 69.9 | 16.3 | 15.7 | 17.0 |
| Male | 10488 | 5.5 | 5.1 | 5.9 | 0.9 | 0.7 | 1.1 | 8.2 | 7.6 | 8.7 | 66.9 | 66.0 | 67.8 | 18.6 | 17.9 | 19.3 |
| **Comorbidities*** | | | | | | | | | | | | | | | | |
| Cancer (current or treated in the last year) | 1092 | 2.2 | 1.3 | 3.1 | 0.6 | 0.1 | 1.0 | 5.0 | 3.7 | 6.2 | 57.8 | 54.9 | 60.7 | 34.5 | 31.7 | 37.3 |
| Cardiovascular disease | 2014 | 2.1 | 1.5 | 2.7 | 0.5 | 0.2 | 0.8 | 4.0 | 3.2 | 4.9 | 56.3 | 54.2 | 58.5 | 37.1 | 35.0 | 39.2 |
| Diabetes | 2028 | 3.0 | 2.3 | 3.8 | 0.6 | 0.3 | 0.9 | 4.4 | 3.5 | 5.3 | 58.2 | 56.1 | 60.3 | 33.8 | 31.8 | 35.9 |
| Hypertension | 7082 | 2.5 | 2.2 | 2.9 | 0.6 | 0.4 | 0.8 | 5.0 | 4.5 | 5.5 | 64.4 | 63.3 | 65.5 | 27.5 | 26.5 | 28.5 |
| Immunosuppression | 434 | 1.4 | 0.3 | 2.5 | 0.7 | -0.1 | 1.5 | 4.6 | 2.6 | 6.6 | 58.3 | 53.7 | 62.9 | 35.0 | 30.5 | 39.5 |
| Lung disease | 2520 | 3.2 | 2.5 | 3.9 | 1.0 | 0.6 | 1.3 | 7.8 | 6.7 | 8.8 | 64.0 | 62.1 | 65.9 | 24.1 | 22.4 | 25.7 |

*only in the ≥ 18years

Supplement 1 Table S6: Proportion of participants' self-reported first infections in 2020, 2021, and 2022 stratified by age, sex, and pre-existing conditions. The results are not age-standardised. Participants from IMMUNEBRIDGE_ED were excluded from this analysis as were participants with missing age information. Information on the date of self-reported infections of NAKO participants (n=10,595) and 365 participants from the other cohorts is unknown; these are therefore not included in this analysis. (LCI: lower 95% confidence interval limit; UCI: upper 95% confidence interval limit)

| | | No reported infection | | | First reported infection 2020 | | | First reported infection 2021 | | | First reported infection 2022 | | |
|---|---|---|---|---|---|---|---|---|---|---|---|---|---|
| | N | Share (%) | LCI | UCI | Share (%) | LCI | UCI | Share (%) | LCI | UCI | Share (%) | LCI | UCI |
| **Total** | 22122 | 61.6 | 61.0 | 62.2 | 2.1 | 1.9 | 2.3 | 4.4 | 4.1 | 4.7 | 31.9 | 31.3 | 32.5 |
| **Age (years)** | | | | | | | | | | | | | |
| 1-17 | 516 | 54.3 | 49.9 | 58.6 | 1.2 | 0.5 | 2.6 | 3.5 | 2.1 | 5.6 | 41.1 | 36.8 | 45.5 |
| 18-29 | 1616 | 41.8 | 39.4 | 44.3 | 3.3 | 2.5 | 4.4 | 6.4 | 5.3 | 7.7 | 48.5 | 46.0 | 50.9 |
| 30-34 | 1429 | 45.6 | 43.0 | 48.3 | 3.2 | 2.3 | 4.2 | 6.0 | 4.9 | 7.4 | 45.2 | 42.6 | 47.8 |
| 35-39 | 1476 | 46.5 | 43.9 | 49.1 | 2.3 | 1.6 | 3.2 | 6.4 | 5.3 | 7.8 | 44.8 | 42.2 | 47.4 |
| 40-49 | 3125 | 49.2 | 47.4 | 51.0 | 2.7 | 2.1 | 3.3 | 5.9 | 5.1 | 6.8 | 42.3 | 40.5 | 44.0 |
| 50-59 | 5118 | 62.1 | 60.8 | 63.4 | 2.2 | 1.8 | 2.6 | 4.5 | 4.0 | 5.1 | 31.2 | 30.0 | 32.5 |
| 60-64 | 2356 | 69.1 | 67.1 | 70.9 | 2.1 | 1.6 | 2.8 | 3.4 | 2.8 | 4.3 | 25.4 | 23.7 | 27.2 |
| 65-79 | 5717 | 76.1 | 74.9 | 77.2 | 1.4 | 1.2 | 1.8 | 2.6 | 2.2 | 3.1 | 19.9 | 18.9 | 21.0 |
| 80 and older | 769 | 83.1 | 80.2 | 85.6 | 1.2 | 0.6 | 2.3 | 3.0 | 2.0 | 4.5 | 12.7 | 10.5 | 15.4 |
| **Sex** | | | | | | | | | | | | | |
| female | 11670 | 61.8 | 60.9 | 62.6 | 1.9 | 1.7 | 2.1 | 4.1 | 3.7 | 4.4 | 32.3 | 31.4 | 33.1 |
| male | 10451 | 61.4 | 60.5 | 62.3 | 2.4 | 2.1 | 2.7 | 4.7 | 4.3 | 5.1 | 31.5 | 30.6 | 32.4 |
| **Antibodies** | | | | | | | | | | | | | |
| N-AB positive | 9084 | 30.1 | 29.2 | 31.0 | 3.2 | 2.8 | 3.6 | 7.1 | 6.6 | 7.7 | 59.6 | 58.6 | 60.6 |
| N-AB negative | 11646 | 86.1 | 85.5 | 86.7 | 1.3 | 1.1 | 1.5 | 2.3 | 2.0 | 2.5 | 10.3 | 9.8 | 10.9 |
| S-AB positive | 20038 | 61.5 | 60.8 | 62.1 | 2.2 | 2.0 | 2.4 | 4.4 | 4.2 | 4.7 | 31.9 | 31.3 | 32.6 |
| S-AB negative | 678 | 64.5 | 60.9 | 68.0 | 1.3 | 0.5 | 2.2 | 3.4 | 2.0 | 4.8 | 30.8 | 27.4 | 34.3 |
| **Comorbidities*** | | | | | | | | | | | | | |
| Cancer (current or treated in the last year) | 1091 | 69.8 | 67.0 | 72.5 | 2.2 | 1.3 | 3.1 | 3.5 | 2.4 | 4.6 | 24.6 | 22.0 | 27.1 |
| Cardiovascular disease | 2001 | 73.0 | 71.1 | 75.0 | 2.3 | 1.6 | 3.0 | 3.2 | 2.4 | 3.9 | 21.5 | 19.7 | 23.3 |
| Diabetes | 2028 | 72.4 | 70.4 | 74.3 | 1.5 | 1.0 | 2.0 | 4.4 | 3.5 | 5.3 | 21.7 | 19.9 | 23.5 |
| Hypertension | 7055 | 68.6 | 67.5 | 69.7 | 2.0 | 1.7 | 2.3 | 3.9 | 3.5 | 4.4 | 25.5 | 24.5 | 26.5 |
| Immunosuppression | 429 | 70.6 | 66.3 | 74.9 | 2.3 | 0.9 | 3.8 | 3.3 | 1.6 | 4.9 | 23.8 | 19.8 | 27.8 |
| Lung disease | 2507 | 63.7 | 61.8 | 65.5 | 2.6 | 1.9 | 3.2 | 4.4 | 3.6 | 5.2 | 29.4 | 27.7 | 31.2 |

*only in the ≥ 18years
