## Supplement 2 for "Estimates of protection against SARS-CoV-2 infection and severe COVID-19 in Germany before the 2022/2023 winter season - the IMMUNEBRIDGE project"

### Supplementary material 2: Ad hoc targeted literature synthesis

Figure S1: Flow chart of the literature search

Figure SF2-SF36: Forest plots

Table S1: Research questions of the systematic review (P Population, I (E) Intervention (Exposure), C Comparator, O Outcome, S Study design)
Table S2: Characteristics of the included studies from the review (n=90)
Table S3: Overview of protective effects (VE, vaccine effectiveness) against SARS-CoV-2 found in the literature: General population (adults > 18 years)
Table S4: Overview of protective effects (VE, vaccine effectiveness) against SARS-CoV-2 found in the literature: Older adults (>60 years)
Table S5: Overview of protective effects (VE, vaccine effectiveness) against SARS-CoV-2 found in the literature: Children and adolescents (5-17 years)

Table S6 Forest Plots: Stratification of study results
Table S7: List of figures of forest plots (SF2-SF36)
Table S8: Excluded primary studies with reason for exclusion (n=48)

Analogous to the current communication of the Robert Koch Institute as well as existing systematic reviews, meta-analyses, compilations of individual studies, and selected individual studies of the last few weeks, a translation of the antibody responses into protective profiles was also carried out in IMMUNEBRIDGE by means of a targeted literature synthesis on the protective effect of different exposure regimens and using epidemiological accompanying data as described above. This interpretation is exploratory and only usable under strong assumptions.

Targeted literature synthesis on the protective effect of different exposure combinations (summary)
Within the framework of IMMUNEBRIDGE, a targeted literature synthesis was carried out. The aim was to provide a systematic overview based on the currently available studies of the protection against infection and severe illness by the different omicron variants depending on various combinations of SARS-CoV-2 immunity by vaccination and/or previous infection. Table S1 shows the research question according to the PICOS scheme. The focus was on vaccines licensed in Germany (Comirnaty/ BNT162b2; Spikevax/mRNA-1273; Vaxzevria/ChAdOx1nCoV-19; JCOVDEN; Nuvaxovid). Risk groups such as chronically ill or immunosuppressed persons were explicitly included. Studies that exclusively examine healthcare workers or pregnant women were excluded.

Supplement 2 Table S1: Research questions of the systematic review (P Population, I (E) Intervention (Exposure), C Comparator, O Outcome, S Study design)

| | |
|---|---|
| P | General population of all ages, people at increased risk of severe COVID-19 disease |
| I (E) | Different combinations of vaccination against SARS-CoV-2, infection with SARS-CoV-2, and seroprevalence of antibodies against S and N antigen of SARS-CoV-2 |
| C | Individuals without vaccination/with lower numbers of vaccinations, without previous SARS-CoV-2 infection, without SARS-CoV-2 antibodies |
| O | Asymptomatic or symptomatic infection, severe COVID-19 disease (hospital admission, intensive care therapy), death from omicron variant |
| S | Randomised controlled trials, observational studies, population- or hospital-based studies |

Studies were identified via the VIEW-hub (last accessed on December 1, 2022), through which the International Vaccine Access Center, the Johns Hopkins Bloomberg School of Public Health, the World Health Organization, and the Coalition for Epidemic Preparedness Innovations provide a weekly online compilation of vaccine effectiveness studies based on a systematic search of preprint and published literature. A data extraction table was used to systematically collect information on study design, setting, population, and results on vaccine effectiveness (VE). The data was extracted by one person and checked by a second person. The data were synthesized qualitatively; a subsequent meta-analysis was not performed due to the great heterogeneity between the studies. The results are presented in tables (summary of the ranges of the point estimates) and in forest plots.

Data were stratified by several parameters:

- age group (children/adolescents, adults 18-60 years, adults 60+
- number of past vaccinations and/or infections
- reference group (not vaccinated, less vaccinated than comparison group)
- outcome (infection, severe disease, death)
- comorbidities (general population vs. diseased)
- days post exposure (=last vaccination or infection: < 14, 14-90, 90-180, > 180 Tage

### Results

In total, we included 90 studies, (see flow chart in Supplement 2 Figure S1), of those 24 studies examined children and adolescents, 53 adults above 18 years, and 28 adults above 60 years. Supplement 2 Table S2 provides an overview of the characteristics of the studies. The most frequently used study design was the test-negative case-control design, followed by retrospective cohort studies that mostly used registered or health claim based data. The older the study populations were, the more they examined higher exposure groups, and outcomes on severe disease or death. Almost half of the studies were published as preprints. Studies on children and adolescents mainly came from North

America (n=9), Asia and Europe (each n=5). Studies on adults predominantly were from the US (n=24), followed by Europe and Asia (each n=11), among the Asian studies five came from Quatar. Studies on older adults mostly took place in North Amerika (each n=5 in the US, and Canada) and Israel (n=6)

There is a high degree of heterogeneity due to differences in populations, vaccines and vaccine combinations, number of vaccinations, time period after last vaccination, survey of exposure (self-reported, documented, PCR test, antigen test, antibody test), definition of outcome (asymptomatic, positive test, severe disease, hospitalisation, intensive care treatment), study design (prospective or retrospective cohort study, case-control study, test-negative case-control study), analysis procedure, and consideration of covariates. Supplement 2 Tables S3, S4 and S5 qualitatively summarise the results on vaccine effectiveness from the primary studies identified so far for the different time periods after last exposure. Detailed results are presented in forest plots (Figures SF2-36) that are stratified by outcome, population group, exposure and reference group.

From the qualitative synthesis of results from studies on adult populations without special attention to risk groups, there is evidence of increasing protection analogous to the defined four categories based on vaccination, infection, and antibody status. With the same vaccination or immune status, there is a higher protection against severe COVID-19 diseases compared to a/symptomatic infections. Protection is highest 14 days to 3 months after the last exposure and then decreases again. This waning effect seems to be higher in older adults and higher for protection from infections rather than from severe course of disease. Compared to previously published systematic reviews, the added value of this review lies in the timeliness and number of identified studies, the differentiation of infection protection according to the time period after the last exposure, different reference groups, and, in particular, the consideration of different age groups.

Supplement 2 Figure S1: Flow chart of the literature search

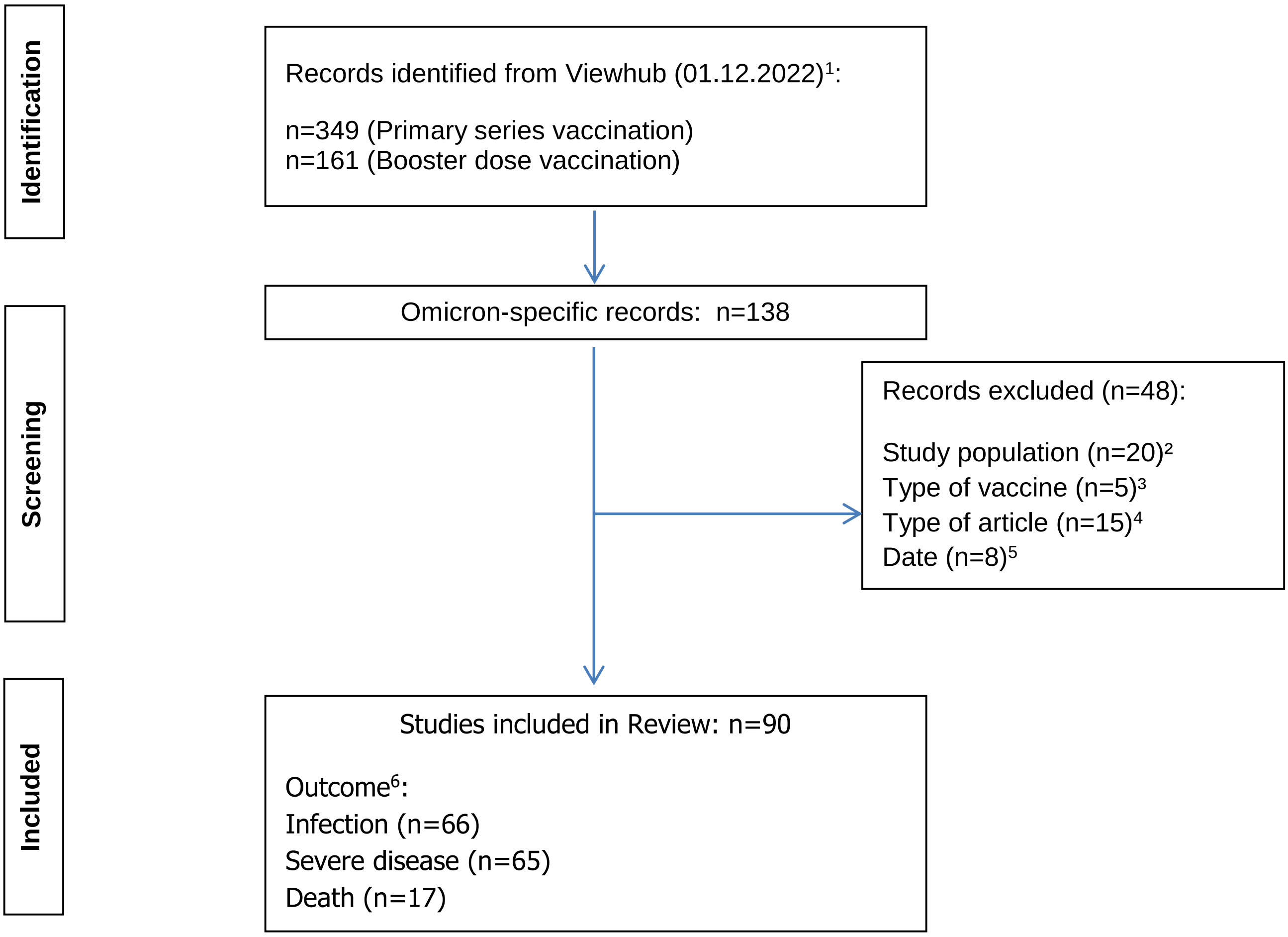

[1] https://view-hub.org/covid-19/effectiveness-studies
[2] Health care workers, prisoners, pregnant people, infants
[3] CoronaVac, Ad5-nCoV, BBIBP-CorV
[4] Letter, Editorial, Correspondence, Report, Comment
[5] before 12.02.2022 (RKI Review)
[6] Multiple outcomes possible

Supplement 2 Table S2: Characteristics of the included studies from the review (n=90)

| | | Children and adolescents | Adults 18+ | Adults 60+ |
|---|---|---|---|---|
| | | n | n | n |
| Total[1] | | 24 | 53 | 28 |
| Country | USA | 5 | 24 | 5 |
| | Canada | 4 | 3 | 5 |
| | Argentina | 2 | 1 | |
| | Brazil | 1 | 1 | |
| | Chile | | 1 | |
| | Hongkong | 1 | 2 | 3 |
| | Israel | 2 | 1 | 6 |
| | Quatar | 1 | 5 | |
| | Japan | 1 | 2 | 1 |
| | Singapore | 2 | 1 | |
| | UK, Scotland | 2 | 3 | 3 |
| | Sweden | 1 | | |
| | Finland | | | 1 |
| | Norway | 1 | | |
| | Denmark | | 4 | 1 |
| | Czech | | 1 | |
| | Italy | 1 | | 1 |
| | France | | 1 | |
| | Iceland | | 1 | |
| | Spain | | 1 | 1 |
| | Portugal | | | 1 |
| | Australia | | 1 | |
| Exposure[2] | 3+ | 2 | 16 | 16 |
| | 3 | 9 | 52 | 21 |
| | 1-3 | 24 | 35 | 15 |
| Outcome[2] | Infection | 18 | 33 | 19 |
| | Severe disease | 11 | 37 | 21 |
| | Death | 1 | 9 | 9 |
| Preprint | | 10 | 28 | 13 |
| Study design | Prospective cohort | 2 | 4 | 2 |
| | Retrospective cohort | 8 | 16 | 9 |
| | Case control study | | 2 | 3 |
| | Test negative design (CCS) | 13 | 30 | 11 |
| | Target trial emulation, cohort | 1 | 1 | 3 |

[1]Total sum of included studies is n=90, calculated sum exceeds this due to some studies examining more than one subpopulation, i.e. children/adolescents, adults 18+, or adults 60+.
[2] Multiple answers possible

Supplement 2 Table S3: Overview of protective effects (VE, vaccine effectiveness) against SARS-CoV-2 found in the literature: General population (adults > 18 years)

| **Exposure** | **VE [3] against** | **VE no information[4]** | **VE 0<14 days** | **VE 14-90 days** | **VE 91-180 days** | **VE >180 days** |
|---|---|---|---|---|---|---|
| **3+ Exposures** | Infection[2] | 34-83% (n=5)[5]<br>47-96% (n=2)[6] | 36% (n=1) | 20-92% (n=4)[5]<br>43-81% (n=1)[6] | 5-82% (n=4)[5]<br>-38-66% (n=2)[6] | |
| | Severe Course[1] | 68-100% (n=5)[5] | | 44-98% (n=5)[5] | 37-86% (n=2)[5] | |
| | Death | 81% (n=1)[5] | | | | |
| **3 Exposures** | Infection[2] | -7-78% (n=16)[5]<br>-4-96% (n=8)[6] | 18-69% (n=6)[5] | 1-91% (n=17)[5]<br>-8-91% (n=8)[6] | 5-67% (n=8)[5]<br>-23-62% (n=4)[6] | -25-55% (n=3)[5]<br>-20-50% (n=3)[6] |
| | Severe Course[1] | 31-99% (n=9)[5]<br>44-89% (n=5)[7] | 65-96% (n=4)[5] | 31-97% (n=10)[5]<br>44-98% (n=4)[7] | 29-95% (n=14)[5]<br>45-83% (n=2)[7] | 31-88% (n=3)[5]<br>84-89% (n=1)[7] |
| | Death | 87-98% (n=3)[5]<br>75-86% (n=1)[7] | 71-86% (n=1)[5] | 78-88% (n=3)[5]<br>75-99% (n=2)[7] | 74-87% (n=2)[5] | 77% (n=1)[5] |
| **1-3 Exposures** | Infection[2] | -46-65% (n=12)[5] | 10-23% (n=2)[5] | 8-55% (n=13)[5] | 1-54% (n=10)[5] | -14-52% (n=11)[5] |
| | Severe Course[1] | 33-86% (n=11)[5] | | -9-96% (n=15)[5] | 24-87% (n=14)[5] | 12-92% (n=9)[5] |
| | Death | 66-90% (n=4)[5] | -9-29% (n=1)[5] | 60-62% (n=2)[5] | 57-70% (n=1)[5] | 49-57% (n=2)[5] |

[1] Different definitions were used for severe course: Hospitalization, treatment in intensive care, and/or death.
[2] Different definitions were used for infection: any, symptomatic, documented
[3] VE, Vaccine effectiveness; Range of point estimates of n included studies is provided, stratified for days post exposure
[4] No information on time after last exposure
[5] Reference group: no exposure (vaccination and/or infection)
[6] Reference group: 3 exposures
[7] Reference group: 2 exposures

Supplement 2 Table S4: Overview of protective effects (VE, vaccine effectiveness) against SARS-CoV-2 found in the literature: Older adults (>60 years)

| **Exposure** | **VE [3] against** | **VE no information[4]** | **VE <14 days** | **VE 14-90 days** | **VE 91-180 days** | **VE >180 days** |
|---|---|---|---|---|---|---|
| **3+ Exposures** | Infection[2] | 49-81% (n=2)[5]<br>36% (n=1)[6] | 33-58% (n=2)[6] | 19-69% (n=2)[5]<br>8-81% (n=7)[6] | 36-60% (n=1)[5]<br>-5-18% (n=1)[6] | 18-44% (n=1)[5]<br>-24- -15% (n=1)[6] |
| | Severe Course[1] | 86% (n=1)[5]<br>64-72% (n=2)[6] | 86-98% (n=1)[5]<br>58% (n=1)[6] | 47-94% (n=4)[5]<br>8-89% (n=8)[6] | 78-89% (n=2)[5]<br>23-28% (n=1)[6] | 74% (n=1)[5]<br>6% (n=1)[6] |
| | Death | 49-82% (n=1)[5]<br>56-78% (n=3)[6] | | 76-90% (n=2)[6] | | |
| **3 Exposures** | Infection[2] | 48-67% (n=2)[5]<br>30-47% (n=3)[7] | 58-66% (n=1)[5] | -27-76% (n=6)[5]<br>54-58% (n=1)[7] | 23-56% (n=3)[5] | |
| | Severe Course[1] | 47-91% (n=5)[5]<br>16-64% (n=3)[7] | 66-95% (n=2)[5] | 51-98% (n=8)[5]<br>57-68% (n=1)[7] | 38-91% (n=6)[5]<br>65-66% (n=1)[7] | 75-85% (n=1)[5]<br>68% (n=1)[7] |
| | Death | 64-98% (n=3)[5]<br>13-86% (n=3)[7] | | 96-98% (n=1)[5] | | |
| **1-3 Exposures** | Infection[2] | 3-79% (n=3)[5] | | -70-56% (n=3)[5] | 24-46% (n=2)[5] | 4-20% (n=2)[5] |
| | Severe Course[1] | 39-82% (n=4)[5] | | 45-98% (n=5)[5] | 31-92% (n=3)[5] | 46-84% (n=3)[5] |
| | Death | 51-91% (n=3)[5] | | 68-93% (n=1)[5] | | |

[1] Different definitions were used for severe course: Hospitalization, treatment in intensive care, and/or death.
[2] Different definitions were used for infection: any, symptomatic, documented
[3] VE, Vaccine effectiveness; Range of point estimates of n included studies is provided, stratified for days post exposure
[4] No information on time after last exposure
[5] Reference group: no exposure (vaccination and/or infection)
[6] Reference group : 3 exposures
[7] Reference group : 2 exposures

Supplement 2 Table S5: Overview of protective effects (VE, vaccine effectiveness) against SARS-CoV-2 found in the literature: Children and adolescents (5-17 years)

| Exposure | VE [3] against | VE no information[4] | VE <14 days | VE 14-90 days | VE 91-180 days | VE >180 days |
|---|---|---|---|---|---|---|
| 3+ Exposures | Infection[2] | 96% (n=1)[5] | 78-90% (n=1)[5] | 80-90% (n=1)[5] | | |
| | Severe Course[1] | | | | | |
| | Death | | | | | |
| 3 Exposures | Infection[2] | 38-79% (n=5)[5] | 55-96% (n=5)[5] | 30-91% (n=4)[5] 76% (n=1)[6] | 34-96% (n=1)[5] | 29-76% (n=1)[5] |
| | Severe Course[1] | | | | | |
| | Death | | | | | |
| 1-3 Exposures | Infection[2] | -2-78% (n=9)[5] | -18-88% (n=6)[5] | -34-85% (n=15)[5] | -25-86% (n=13)[5] | -2-90% (n=4)[5] |
| | Severe Course[1] | 33-85% (n=10)[5] | 21-65% (n=1)[5] | 34-94% (n=8)[5] | -3-92% (n=5)[5] | 16-88% (n=3)[5] |
| | Death | 98% (n=1)[5] | | | | |

[1] Different definitions were used for severe course: Hospitalization, treatment in intensive care, and/or death.
[2] Different definitions were used for infection: any, symptomatic, documented
[3] VE, Vaccine effectiveness; Range of point estimates of n included studies is provided, stratified for days post exposure
[4] No information on time after last exposure
[5] Reference group: no exposure (vaccination and/or infection)
[6] Reference group: 3 exposures

### Forest Plots

Supplement 2 Table S6: Stratification of study results

| Feature | Category | Explanation |
|---|---|---|
| Outcome | Infection | Any, a-/symptomatic, documented, confirmed |
| | Severe disease | Hospitalisation, intensive care use , death |
| | Death | All cause moratlity, COVID-19 related mortality (within 30 days after infection) |
| Population | General population | Adults 18-59 years, all ages (studies without age specification) |
| | Older adults | 60+ years |
| | Children and adolescents | 5-17 years |
| Exposure | 3+ exposures | More than 3 exposures (vaccination or infection, thereof one in 2022) |
| | 3 exposures | 3 exposures (vaccination or infection) |
| | 1-3 exposures | 1-2 exposures (vaccination or infection) |
| Reference | Unvaccinated | No vaccination and/or infection |
| | Less exposures | Less vaccinations/infections |
| Time after last exposure | No information | No specification of time after last exposure |
| | <14 days post exposure | Within 2 weeks after exposure |
| | 14-90 days post exposure | Up to ~ 3 months after exposure |
| | 91-180 days post exposure | ~ 3 to 6 months after exposure |
| | >180 days post exposure | More than ~ 6 months after exposure |

**Multiple naming of a study within a forest plot is possible, due to further stratification within the study, e.g. for:** Vaccine type, Omicron subvariant, combination of exposures, more than one time periode within a category of time after exposure, reported outcome

Supplement 2 Table S7: List of figures of forest plots:

| | |
|---|---|
| SF2 | Infection: General population (adults 18+ years), Exposure 3+, Reference unvaccinated (n=9) |
| SF3 | Infection: General population (18+ years); Exposure 3+, Reference less exposures (3 exposures) (n=4) |
| SF4 | Infection: General population (adults 18+ years), Exposure 3, Reference unvaccinated (n=24) |
| SF5 | Infection: General population (adults 18+), Exposure 3, Reference less exposures (2 exposures) (n=14) |
| SF6 | Infection: General population (adults 18+), Exposure 1-3, Reference unvaccinated (n=21) |
| SF7 | Infection: Older adults (60+ years), Exposure 3+, Reference unvaccinated (n=4) |
| SF8 | Infection: Older adults (60+ years), Exposure 3+, Reference less exposures (3 exposures) (n=8) |
| SF9 | Infection: Older adults (60+years), Exposure 3, Reference unvaccinated (n=9) |
| SF10 | Infection: Older adults (60+ years), Exposure 3, Reference less exposures (2 exposures) (n=4) |
| SF11 | Infection: Older adults (60+ years), Exposure 1-3, Reference unvaccinated (n=8) |
| SF12 | Infection: Children and adolescents (5-17 years), Exposure 3+, Reference unvaccinated (n=2) |
| SF13 | Infection: Children and adolescents (5-17 years), Exposure 3, Reference unvaccinated (n=7) |
| SF14 | Infection: Children and adolescents (5-17 years), Exposure 3, Reference less exposures (2 exposures) (n=1) |
| SF15 | Infection: Children and adolescents (5-17 years), Exposure 1-3, Reference unvaccinated (n=17) |
| SF16 | Severe disease: General population (adults 18+ years), Exposure 3+, Reference unvaccinated (n=10) |
| SF17 | Severe disease: General population (adults 18+ years), Exposure 3, Reference unvaccinated (n=24) |

| | |
|---|---|
| SF18 | Severe disease: General population (adults 18+), Exposure 3, Reference less exposures (2 exposures) (n=9) |
| SF19 | Severe disease: General population (adults 18+ years), Exposure 1-3, Reference unvaccinated (n=23) |
| SF20 | Severe disease: Older adults (60+ years), Exposure 3+, Reference unvaccinated (n=4) |
| SF21 | Severe disease: Older adults (60+ years), Exposure 3+, Reference less exposures (3 exposures) (n=10) |
| SF22 | Severe disease: Older adults (60+ years), Exposure 3, Reference unvaccinated (n=12) |
| SF23 | Severe disease: Older adults (60+ years), Exposure 3, Reference less exposures (2 exposures) (n=4) |
| SF24 | Severe disease: Older adults (60+ years), Exposure 1-3, Reference unvaccinated (n=11) |
| SF25 | Severe disease: Children and adolescents (5-17 years), Exposure 3, Reference vaccinated (n=4) |
| SF26 | Severe disease: Children and adolescents (5-17 years), Exposure 1-3, Reference unvaccinated (n=11) |
| SF27 | Death: General population (adults 18+ years), Exposure 3+, Reference unvaccinated (n=1) |
| SF28 | Death: General population (adults 18+ years), Exposure 3, Reference unvaccinated (n=6) |
| SF29 | Death: General population (adults 18+ years), Exposure 3, Reference less exposures (2 exposures) (n=3) |
| SF30 | Death: General population (adults 18+ years), Exposure 1-3, Reference unvaccinated (n=4) |
| SF31 | Death: Older adults (60+ years), Exposure 3+, Reference unvaccinated (n=1) |
| SF32 | Death: Older adults (60+ years), Exposure 3+, Reference less exposures (3 exposures) (n=5) |
| SF33 | Death: Older adults (60+ years), Exposure 3, Reference unvaccinated (n=4) |
| SF34 | Death: Older adults (60+ years), Exposure 3, Reference less exposures (2 exposures) (n=3) |
| SF35 | Death: Older adults (60+ years), Exposure 1-3, Reference unvaccinated (n=4) |
| SF36 | Death: Children and adolescents (5-17 years), Exposure 1-3, Reference unvaccinated (n=1) |

**SF2:** Infection: General population (adults 18+ years), Exposure 3+, Reference unvaccinated (n=9)

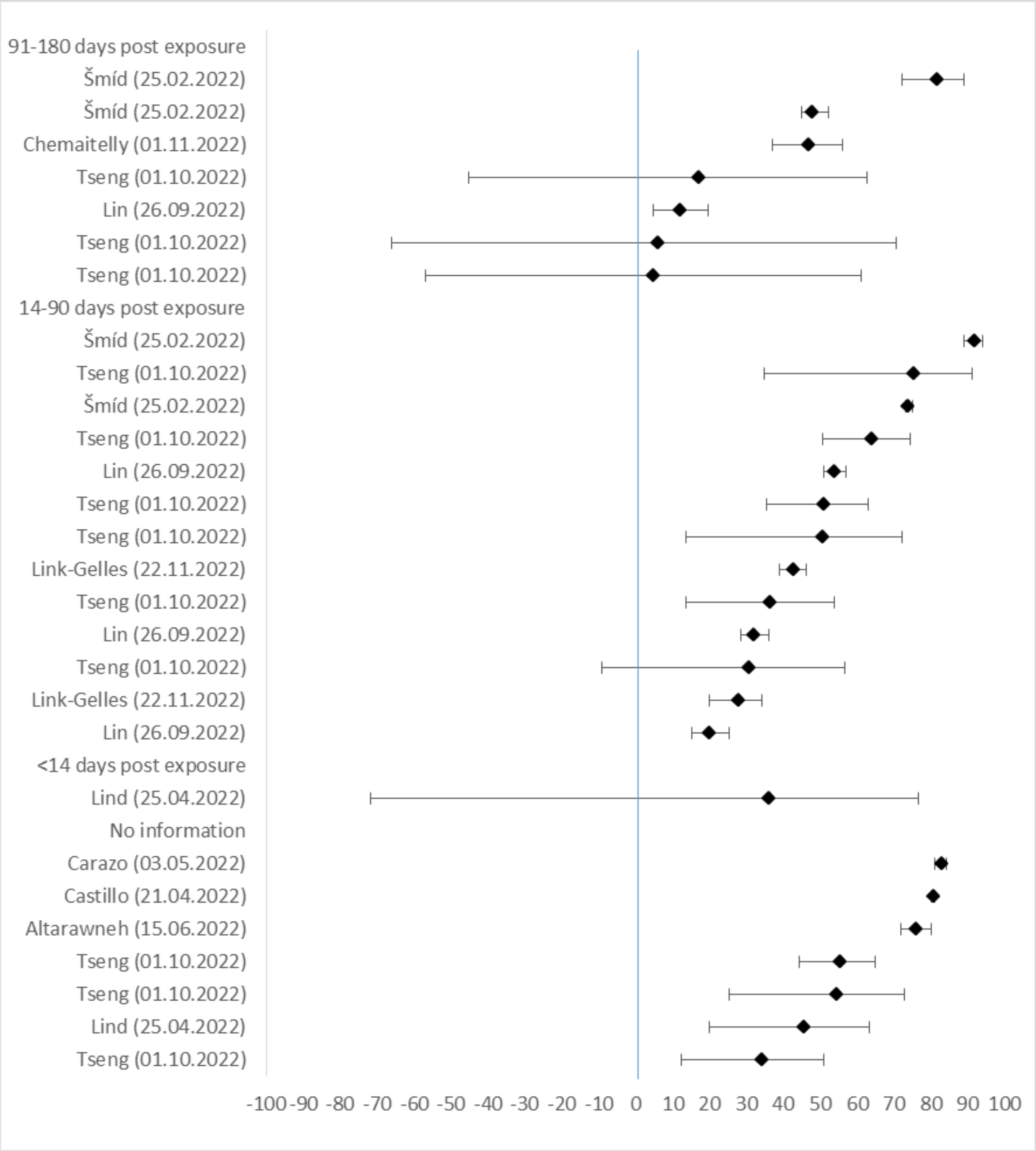

| Author (Date) | VE in % | lower CI 95% | upper CI 95% |
|---|---|---|---|
| **91-180 days post exposure** | | | |
| Šmíd (25.02.2022) | 82,0 | 72,0 | 89,0 |
| Šmíd (25.02.2022) | 48,0 | 45,0 | 52,0 |
| Chemaitelly (01.11.2022) | 47,0 | 37,0 | 56,0 |
| Tseng (01.10.2022) | 17,3 | -45,3 | 62,6 |
| Lin (26.09.2022) | 12,4 | 4,6 | 19,5 |
| Tseng (01.10.2022) | 6,3 | -66,3 | 70,4 |
| Tseng (01.10.2022) | 5,0 | -56,9 | 61,1 |
| **14-90 days post exposure** | | | |
| Šmíd (25.02.2022) | 92,0 | 89,0 | 94,0 |
| Tseng (01.10.2022) | 75,7 | 34,7 | 91,0 |
| Šmíd (25.02.2022) | 74,0 | 73,0 | 75,0 |
| Tseng (01.10.2022) | 64,3 | 50,7 | 74,2 |
| Lin (26.09.2022) | 53,9 | 50,8 | 56,8 |
| Tseng (01.10.2022) | 51,1 | 35,5 | 63,0 |
| Tseng (01.10.2022) | 50,9 | 13,4 | 72,1 |
| Link-Gelles (22.11.2022) | 43,0 | 39,0 | 46,0 |
| Tseng (01.10.2022) | 36,7 | 13,6 | 53,6 |
| Lin (26.09.2022) | 32,3 | 28,5 | 35,9 |
| Tseng (01.10.2022) | 30,8 | -9,2 | 56,5 |
| Link-Gelles (22.11.2022) | 28,0 | 20,0 | 34,0 |
| Lin (26.09.2022) | 20,3 | 15,2 | 25,2 |
| **<14 days post exposure** | | | |
| Lind (25.04.2022) | 36,3 | -71,8 | 76,4 |
| **No information** | | | |
| Carazo (03.05.2022) | 83,0 | 81,0 | 84,0 |
| Castillo (21.04.2022) | 81,0 | 80,0 | 81,0 |
| Altarawneh (15.06.2022) | 76,3 | 71,7 | 80,1 |
| Tseng (01.10.2022) | 55,7 | 44,2 | 64,9 |
| Tseng (01.10.2022) | 54,8 | 25,1 | 72,7 |
| Lind (25.04.2022) | 45,8 | 20,0 | 63,2 |
| Tseng (01.10.2022) | 34,3 | 12,2 | 50,8 |

**SF3:** Infection: General population (18+ years); Exposure 3+, Reference less exposures (3 exposures) (n=4)

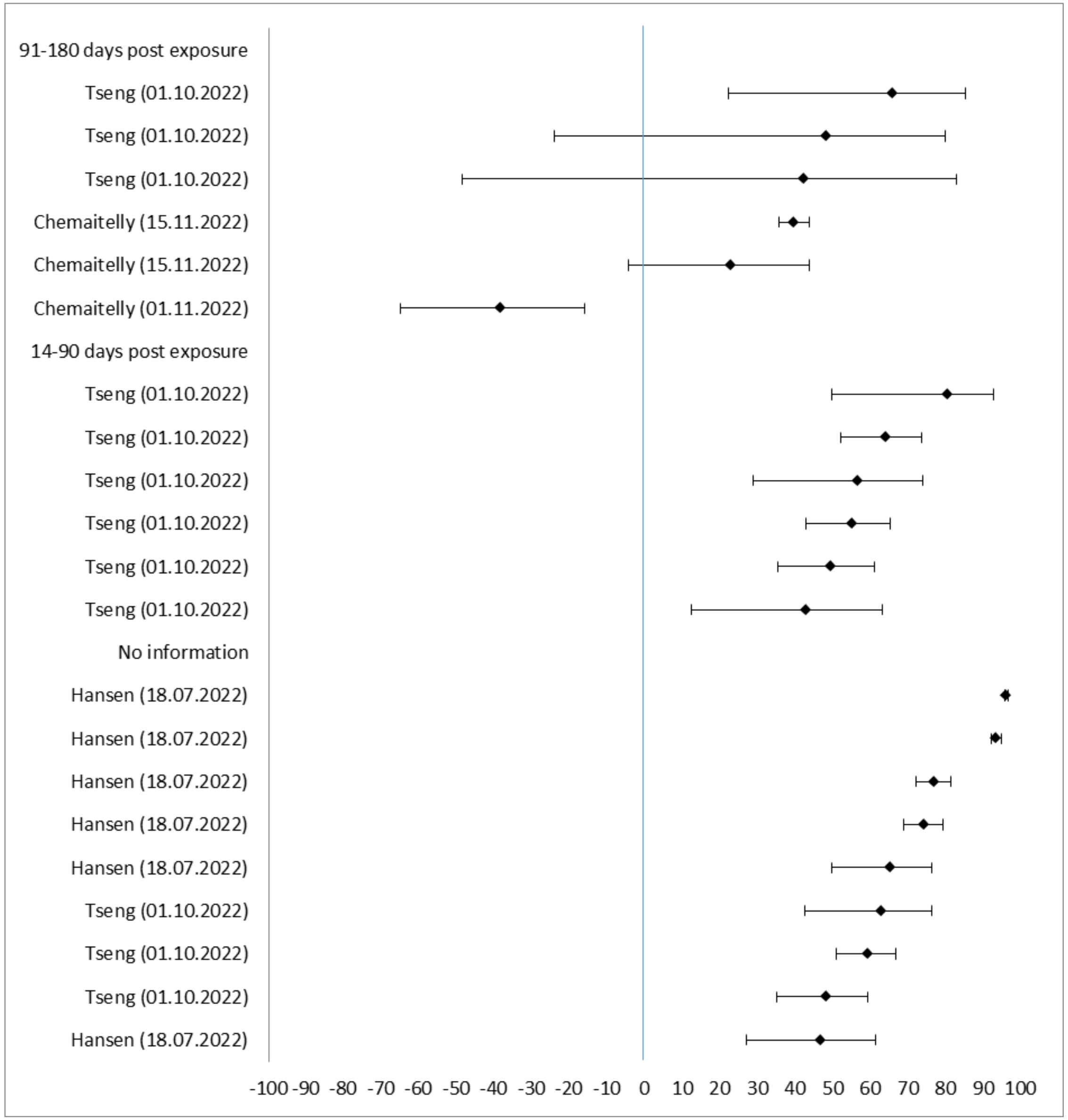

| Author (date) | VE in % | lower CI 95% | upper CI 95% |
|---|---|---|---|
| **91-180 days post exposure** | | | |
| Tseng (01.10.2022) | 66,1 | 22,1 | 85,3 |
| Tseng (01.10.2022) | 48,4 | -24,2 | 79,8 |
| Tseng (01.10.2022) | 42,4 | -48,5 | 82,9 |
| Chemaitelly (15.11.2022) | 39,8 | 35,7 | 43,6 |
| Chemaitelly (15.11.2022) | 23,2 | -4,5 | 43,7 |
| Chemaitelly (01.11.2022) | -38,0 | -65,0 | -16,0 |
| **14-90 days post exposure** | | | |
| Tseng (01.10.2022) | 80,8 | 49,6 | 92,7 |
| Tseng (01.10.2022) | 64,3 | 52,0 | 73,5 |
| Tseng (01.10.2022) | 56,9 | 28,8 | 73,9 |
| Tseng (01.10.2022) | 55,4 | 42,9 | 65,1 |
| Tseng (01.10.2022) | 49,8 | 35,3 | 61,1 |
| Tseng (01.10.2022) | 43,2 | 12,4 | 63,1 |
| **No information** | | | |
| Hansen (18.07.2022) | 96,3 | 95,8 | 96,7 |
| Hansen (18.07.2022) | 93,6 | 92,1 | 94,8 |
| Hansen (18.07.2022) | 77,2 | 72,2 | 81,3 |
| Hansen (18.07.2022) | 74,5 | 68,7 | 79,2 |
| Hansen (18.07.2022) | 65,4 | 49,8 | 76,2 |
| Tseng (01.10.2022) | 63,0 | 42,5 | 76,2 |
| Tseng (01.10.2022) | 59,6 | 50,9 | 66,8 |
| Tseng (01.10.2022) | 48,5 | 35,0 | 59,1 |
| Hansen (18.07.2022) | 46,9 | 27,0 | 61,3 |

**SF4:**Infection: General population (adults 18+ years), Exposure 3, Reference unvaccinated (n=24)

| Author (Date) | VE in % | lower CI 95% | upper CI 95 % |
|---|---|---|---|
| **>180 days post exposure** | | | |
| Tseng (01.10.2022) | 54,9 | 35,6 | 68,4 |
| Wang (25.03.2022) | 50,0 | 45,0 | 55,0 |
| Lind (25.04.2022) | 34,0 | 18,5 | 46,5 |
| Tseng (01.10.2022) | -16,4 | -35,8 | 8,2 |
| Tseng (01.10.2022) | -17,9 | -29,6 | -4,2 |
| Tseng (01.10.2022) | -24,9 | -32,3 | -16,7 |
| **91-180 days post exposure** | | | |
| Tseng (01.10.2022) | 67,3 | 62,0 | 71,9 |
| Chemaitelly (01.11.2022) | 57,0 | 52,0 | 62,0 |
| Gram (01.09.2022) | 52,3 | 48,0 | 56,2 |
| Gram (01.09.2022) | 51,3 | 49,8 | 52,7 |
| Castillo (21.04.2022) | 50,0 | 48,0 | 51,0 |
| Hansen (30.03.2022) | 40,5 | 38,9 | 42,2 |
| Hansen (30.03.2022) | 38,6 | 37,7 | 39,5 |
| Hansen (30.03.2022) | 37,9 | 33,4 | 42,0 |
| Kirsbom (01.05.2022) | 37,2 | -44,1 | 72,1 |
| Hansen (30.03.2022) | 36,9 | 34,8 | 38,9 |
| Kirsbom (01.05.2022) | 30,6 | 26,8 | 34,3 |
| Cerqueira-Silva (14.04.2022) | 29,2 | 25,0 | 33,1 |
| Tseng (01.10.2022) | 23,2 | -12,3 | 48,3 |
| Chemaitelly (02.06.2022) | 21,9 | 7,7 | 33,9 |
| Tseng (01.10.2022) | 20,7 | -1,6 | 38,2 |
| Lin (26.09.2022) | 11,7 | 9,9 | 13,5 |
| Tseng (01.10.2022) | 10,8 | 0,8 | 19,8 |
| Cerqueira-Silva (14.04.2022) | 5,4 | 3,2 | 7,5 |
| **14-90 days post exposure** | | | |
| Tseng (01.10.2022) | 90,6 | 30,6 | 98,7 |
| Šmíd (25.02.2022) | 86,0 | 85,0 | 88,0 |
| Tseng (01.10.2022) | 85,8 | 82,7 | 88,3 |
| Šmíd (25.02.2022) | 82,0 | 75,0 | 87,0 |
| Šmíd (25.02.2022) | 77,0 | 76,0 | 78,0 |
| Tseng (01.10.2022) | 76,3 | 73,9 | 78,6 |
| Tseng (01.10.2022) | 72,6 | -54,7 | 96,6 |
| Tseng (21.04.2022) | 71,6 | 69,7 | 73,4 |
| Castillo (21.04.2022) | 67,0 | 67,0 | 68,0 |
| Rennert (07.05.2022) | 66,4 | 46,1 | 79,0 |
| Wang (25.03.2022) | 65,0 | 63,0 | 66,0 |
| Kirsbom (01.05.2022) | 63,8 | 63,0 | 64,5 |
| Lin (26.09.2022) | 62,9 | 62,3 | 63,3 |
| Buchan (22.09.2022) | 61,0 | 56,0 | 65,0 |
| Tseng (01.10.2022) | 61,0 | 27,6 | 79,0 |
| Castillo (21.04.2022) | 59,0 | 59,0 | 60,0 |
| Castillo (21.04.2022) | 58,0 | 57,0 | 59,0 |
| Kirsbom (01.05.2022) | 57,3 | 56,4 | 58,2 |
| Tseng (01.10.2022) | 57,0 | 26,5 | 75,0 |
| Šmíd (25.02.2022) | 56,0 | 55,0 | 56,0 |
| Chemaitelly (02.06.2022) | 55,5 | 49,3 | 61,0 |
| Gram (01.09.2022) | 55,1 | 54,6 | 55,5 |
| Chemaitelly (02.06.2022) | 53,7 | 39,6 | 64,6 |
| Chemaitelly (02.06.2022) | 53,7 | 41,5 | 63,3 |
| Cerqueira-Silva (14.04.2022) | 53,4 | 51,4 | 55,3 |
| Kirsbom (01.05.2022) | 53,0 | 42,6 | 61,6 |
| Gram (01.09.2022) | 52,7 | 51,8 | 53,6 |
| Kirsbom (01.05.2022) | 51,7 | 38,9 | 61,8 |
| Chemaitelly (02.06.2022) | 51,5 | 45,0 | 57,2 |
| Gram (01.09.2022) | 51,1 | 50,4 | 51,7 |
| Tsang (25.08.2022) | 50,9 | 31,0 | 65,0 |
| Lin (26.09.2022) | 50,8 | 50,1 | 51,5 |
| Hansen (30.03.2022) | 47,9 | 47,4 | 48,3 |
| Hansen (30.03.2022) | 47,7 | 47,0 | 48,3 |
| Tseng (21.04.2022) | 47,4 | 40,5 | 53,5 |
| Kirsbom (01.05.2022) | 46,4 | 45,0 | 47,8 |
| Hansen (30.03.2022) | 45,5 | 44,9 | 46,2 |
| Rennert (07.05.2022) | 45,4 | 30,0 | 57,4 |
| Šmíd (25.02.2022) | 45,0 | 44,0 | 46,0 |
| Chemaitelly (02.06.2022) | 43,6 | 36,5 | 49,9 |
| Hansen (30.03.2022) | 43,5 | 42,2 | 44,7 |
| Cerqueira-Silva (14.04.2022) | 42,3 | 41,6 | 42,9 |
| Tsang (25.08.2022) | 41,4 | 23,2 | 55,2 |
| Tseng (01.10.2022) | 41,2 | 28,3 | 51,8 |
| Link-Gelles (22.11.2022) | 41,0 | 31,0 | 46,0 |
| Hansen (30.03.2022) | 41,0 | 40,3 | 41,7 |
| Hansen (30.03.2022) | 41,0 | 40,5 | 41,5 |
| Kirsbom (01.05.2022) | 40,8 | 18,6 | 56,9 |
| Lin (26.09.2022) | 40,6 | 39,7 | 41,6 |
| Chemaitelly (02.06.2022) | 37,3 | 25,4 | 47,3 |
| Chemaitelly (02.06.2022) | 32,6 | 17,8 | 44,8 |
| Chemaitelly (02.06.2022) | 31,5 | 20,3 | 41,1 |
| Šmíd (25.02.2022) | 21,0 | 19,0 | 23,0 |
| Tseng (01.10.2022) | 0,7 | -53,6 | 54,2 |
| **<14 days post exposure** | | | |
| Arashiro (03.08.2022) | 69,0 | 50,0 | 81,0 |
| Kirsbom (01.05.2022) | 61,2 | 40,9 | 74,9 |
| Kirsbom (01.05.2022) | 58,2 | 57,0 | 59,4 |
| Lind (25.04.2022) | 38,1 | 18,6 | 52,9 |
| Buchan (22.09.2022) | 36,0 | 29,0 | 43,0 |
| Castillo (21.04.2022) | 35,0 | 34,0 | 36,0 |
| Lind (25.04.2022) | 33,2 | 3,7 | 53,6 |
| Chemaitelly (02.06.2022) | 19,7 | -9,7 | 41,2 |
| Chemaitelly (02.06.2022) | 17,7 | 2,5 | 30,6 |
| **No information** | | | |
| Arashiro (03.08.2022) | 78,0 | 67,0 | 86,0 |
| Tseng (01.10.2022) | 76,6 | 74,4 | 78,6 |
| Carazo (03.05.2022) | 73,0 | 72,0 | 73,0 |
| Mimura (08.10.2022) | 71,8 | 60,1 | 80,1 |
| Carazo (03.05.2022) | 68,0 | 67,0 | 70,0 |
| Young-Xu (03.08.2022) | 64,0 | 63,0 | 65,0 |
| Kim (10.04.2022) | 62,0 | 48,0 | 72,0 |
| Risk (16.08.2022) | 57,0 | 51,0 | 62,0 |
| Lind (25.04.2022) | 56,9 | 52,1 | 61,2 |
| Mimura (08.10.2022) | 56,5 | 46 | 65 |
| Altarawneh (15.06.2022) | 55,5 | 51,8 | 59,0 |
| Altarawneh (15.06.2022) | 54,0 | 50,4 | 57,3 |
| Risk (16.08.2022) | 35,0 | 29,0 | 41,0 |
| Tseng (01.10.2022) | -2,2 | -10,5 | 6,4 |
| Tseng (01.10.2022) | -7,0 | -19,8 | 7,2 |
| Tseng (01.10.2022) | -7,2 | -27,9 | 16,4 |

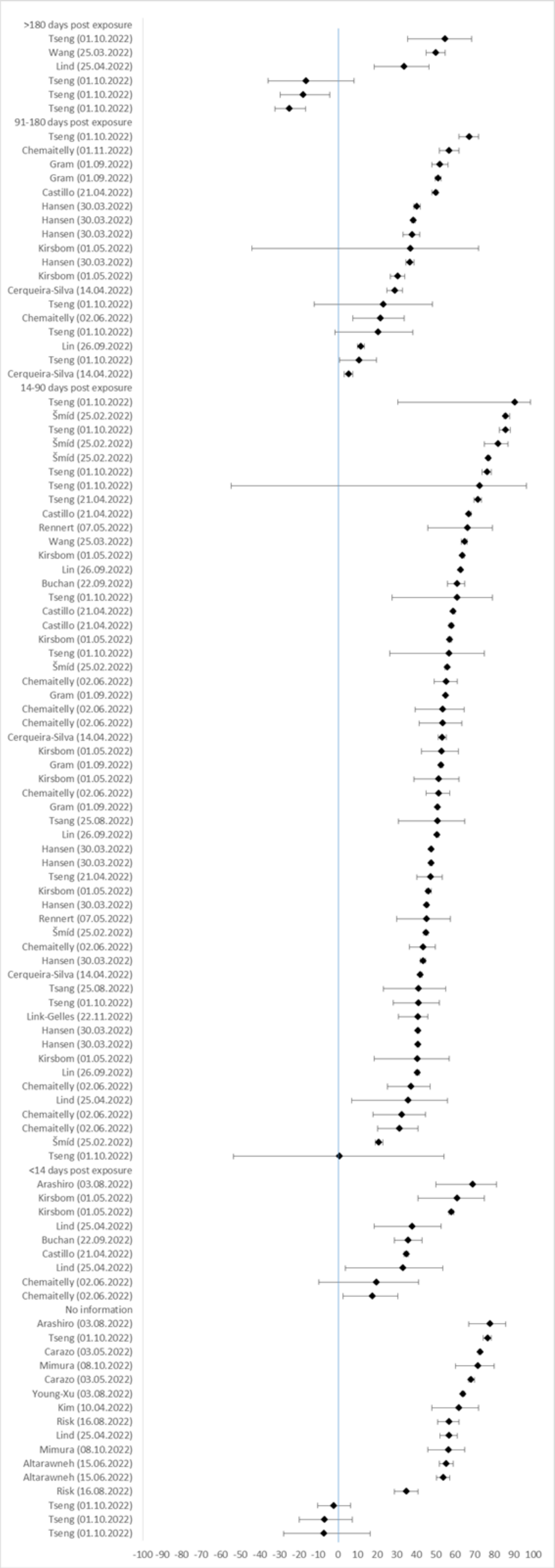
>180 days post exposure
Tseng (01.10.2022)
Wang (25.03.2022)
Lind (25.04.2022)
Tseng (01.10.2022)
Tseng (01.10.2022)
Tseng (01.10.2022)
91-180 days post exposure
Tseng (01.10.2022)
Chemaitelly (01.11.2022)
Gram (01.09.2022)
Gram (01.09.2022)
Castillo (21.04.2022)
Hansen (30.03.2022)
Hansen (30.03.2022)
Hansen (30.03.2022)
Kirsbom (01.05.2022)
Hansen (30.03.2022)
Kirsbom (01.05.2022)
Cerqueira-Silva (14.04.2022)
Tseng (01.10.2022)
Chemaitelly (02.06.2022)
Tseng (01.10.2022)
Lin (26.09.2022)
Tseng (01.10.2022)
Cerqueira-Silva (14.04.2022)
14-90 days post exposure
Tseng (01.10.2022)
Šmíd (25.02.2022)
Tseng (01.10.2022)
Šmíd (25.02.2022)
Šmíd (25.02.2022)
Tseng (01.10.2022)
Tseng (01.10.2022)
Tseng (21.04.2022)
Castillo (21.04.2022)
Rennert (07.05.2022)
Wang (25.03.2022)
Kirsbom (01.05.2022)
Lin (26.09.2022)
Buchan (22.09.2022)
Tseng (01.10.2022)
Castillo (21.04.2022)
Castillo (21.04.2022)
Kirsbom (01.05.2022)
Tseng (01.10.2022)
Šmíd (25.02.2022)
Chemaitelly (02.06.2022)
Gram (01.09.2022)
Chemaitelly (02.06.2022)
Chemaitelly (02.06.2022)
Cerqueira-Silva (14.04.2022)
Kirsbom (01.05.2022)
Gram (01.09.2022)
Kirsbom (01.05.2022)
Chemaitelly (02.06.2022)
Gram (01.09.2022)
Tsang (25.08.2022)
Lin (26.09.2022)
Hansen (30.03.2022)
Hansen (30.03.2022)
Tseng (21.04.2022)
Kirsbom (01.05.2022)
Hansen (30.03.2022)
Rennert (07.05.2022)
Šmíd (25.02.2022)
Chemaitelly (02.06.2022)
Hansen (30.03.2022)
Cerqueira-Silva (14.04.2022)
Tsang (25.08.2022)
Tseng (01.10.2022)
Link-Gelles (22.11.2022)
Hansen (30.03.2022)
Hansen (30.03.2022)
Kirsbom (01.05.2022)
Lin (26.09.2022)
Chemaitelly (02.06.2022)
Lind (25.04.2022)
Chemaitelly (02.06.2022)
Chemaitelly (02.06.2022)
Šmíd (25.02.2022)
Tseng (01.10.2022)
<14 days post exposure
Arashiro (03.08.2022)
Kirsbom (01.05.2022)
Kirsbom (01.05.2022)
Lind (25.04.2022)
Buchan (22.09.2022)
Castillo (21.04.2022)
Lind (25.04.2022)
Chemaitelly (02.06.2022)
Chemaitelly (02.06.2022)
No information
Arashiro (03.08.2022)
Tseng (01.10.2022)
Carazo (03.05.2022)
Mimura (08.10.2022)
Carazo (03.05.2022)
Young-Xu (03.08.2022)
Kim (10.04.2022)
Risk (16.08.2022)
Lind (25.04.2022)
Mimura (08.10.2022)
Altarawneh (15.06.2022)
Altarawneh (15.06.2022)
Risk (16.08.2022)
Tseng (01.10.2022)
Tseng (01.10.2022)
Tseng (01.10.2022)
-100 -90 -80 -70 -60 -50 -40 -30 -20 -10 0 10 20 30 40 50 60 70 80 90 100

**SF5**: Infection: General population (adults 18+), Exposure 3, Reference less exposures (2 exposures) (n=14)

| Author (date) | VE in % | lower CI 95% | upper CI 95% |
|---|---|---|---|
| **>180 days post exposure** | | | |
| Tseng (01.10.2022) | 50,0 | 28,4 | 65,1 |
| Ng (26.08.2022) | 14,6 | 11,0 | 18,0 |
| Chemaitelly (15.11.2022) | 14,4 | 7,3 | 20,9 |
| Ng (26.08.2022) | 9,7 | 2,7 | 16,3 |
| Ng (26.08.2022) | 3,7 | 0,0 | 7,2 |
| Ng (26.08.2022) | -2,8 | -5,4 | 0,3 |
| Tseng (01.10.2022) | -13,1 | -36,3 | 15,6 |
| Tseng (01.10.2022) | -13,6 | -23,4 | -2,5 |
| Tseng (01.10.2022) | -17,1 | -31,4 | 0,2 |
| Chemaitelly (15.11.2022) | -20,3 | -55,0 | 29,0 |
| **91-180 days post exposure** | | | |
| Tseng (01.10.2022) | 62,0 | 55,5 | 67,5 |
| Chemaitelly (15.11.2022) | 41,2 | 40,1 | 42,3 |
| Chemaitelly (15.11.2022) | 41,1 | 40,0 | 42,1 |
| Chemaitelly (15.11.2022) | 38,5 | 37,3 | 39,8 |
| Tseng (01.10.2022) | 29,1 | -6,9 | 53,2 |
| Tseng (01.10.2022) | 22,5 | 12,8 | 31,2 |
| Patalon (09.06.2022) | 18,3 | 15,2 | 21,2 |
| Tseng (01.10.2022) | 18,0 | -6,7 | 37,3 |
| Patalon (09.06.2022) | 16,0 | 12,3 | 19,5 |
| Butt (03.05.2022) | -9,0 | -22,0 | 2,0 |
| Butt (03.05.2022) | -23,0 | -65,0 | 9,0 |
| **14-90 days post exposure** | | | |
| Tseng (01.10.2022) | 91,0 | 33,6 | 98,8 |
| Tseng (01.10.2022) | 81,0 | 77,0 | 84,4 |
| Tseng (01.10.2022) | 71,8 | -55,7 | 96,5 |
| Andersson (29.11.2022) | 69,8 | 69,2 | 70,5 |
| Tseng (01.10.2022) | 68,8 | 65,5 | 71,9 |
| Andersson (29.11.2022) | 65,1 | 62,8 | 67,4 |
| Tseng (01.10.2022) | 63,6 | 32,3 | 80,4 |
| Patalon (09.06.2022) | 59,4 | 54,9 | 63,5 |
| Chemaitelly (15.11.2022) | 57,1 | 55,9 | 58,3 |
| Tseng (01.10.2022) | 53,3 | 18,9 | 73,1 |
| Monge (02.06.2022) | 51,3 | 50,2 | 52,4 |
| Monge (02.06.2022) | 49,9 | 48,6 | 51,3 |
| Tseng (01.10.2022) | 48,5 | 36,7 | 58,1 |
| Andersson (29.11.2022) | 47,9 | 41,5 | 54,3 |
| Patalon (09.06.2022) | 43,2 | 38,2 | 47,8 |
| Ng (26.08.2022) | 41,3 | 39,4 | 43,1 |
| Butt (03.05.2022) | 40,0 | 35,0 | 44,0 |
| Andersson (29.11.2022) | 39,5 | 37,2 | 41,8 |
| Gonzales (27.09.2022) | 36,0 | 33,0 | 39,0 |
| Ng (26.08.2022) | 35,6 | 32,8 | 38,3 |
| Ng (26.08.2022) | 34,9 | 33,0 | 36,8 |
| Ng (26.08.2022) | 31,7 | 30,0 | 33,4 |
| Andersson (29.11.2022) | 31 | 25,5 | 36,6 |
| Butt (03.05.2022) | 30,0 | 23,0 | 36,0 |
| Patalon (09.06.2022) | 29,1 | 26,1 | 32,0 |
| Ng (26.08.2022) | 26,3 | 24,2 | 28,3 |
| Butt (03.05.2022) | 24,0 | 17,0 | 31,0 |
| Ng (26.08.2022) | 23,4 | 20,7 | 26,1 |
| Andersson (29.11.2022) | 23 | 21,3 | 24,8 |
| Butt (03.05.2022) | 23,0 | 18,0 | 27,0 |
| Ng (26.08.2022) | 18,3 | 15,9 | 20,7 |
| Butt (03.05.2022) | 12,0 | 1,0 | 21,0 |
| Ng (26.08.2022) | 11,9 | 9,7 | 14,0 |
| Gonzales (27.09.2022) | 8,0 | -4,0 | 19,0 |
| Andersson (29.11.2022) | 4,2 | 0,2 | 8,2 |
| Andersson (29.11.2022) | 0,8 | -1,0 | 2,6 |
| Butt (03.05.2022) | 0,0 | -7,0 | 6,0 |
| Tseng (01.10.2022) | -8,2 | -57,8 | 50,0 |
| **No information** | | | |
| Norddahl (01.03.2022) | 96,0 | 25,0 | 64,0 |
| Liu (21.06.2022) | 93,8 | 92,9 | 94,5 |
| Tseng (01.10.2022) | 70,3 | 67,4 | 73,0 |
| Petrie (16.04.2022) | 70,0 | 51,0 | 81,0 |
| Petrie (16.04.2022) | 66,0 | 46,0 | 79,0 |
| Mimura (08.10.2022) | 64,2 | 49,9 | 74,4 |
| Norddahl (01.03.2022) | 50,0 | 34,0 | 62,0 |
| Abu-Raddad (12.05.2022) | 49,4 | 47,1 | 51,6 |
| Mimura (08.10.2022) | 48,3 | 36,4 | 57,9 |
| Abu-Raddad (12.05.2022) | 47,3 | 40,7 | 53,3 |
| Norddahl (01.03.2022) | 47,0 | 36,0 | 56,0 |
| Ioannou (16.06.2022) | 46,5 | 44,1 | 48,7 |
| Ioannou (16.06.2022) | 44,6 | 42,5 | 46,6 |
| Ioannou (16.06.2022) | 44,3 | 42,2 | 46,3 |
| Ioannou (16.06.2022) | 43,3 | 41,4 | 45,4 |
| Ioannou (16.06.2022) | 42,3 | 40,6 | 43,9 |
| Ioannou (16.06.2022) | 39,6 | 36,9 | 42,1 |
| Ioannou (16.06.2022) | 39,0 | 36,4 | 41,6 |
| Ioannou (16.06.2022) | 36,4 | 33,3 | 39,4 |
| Norddahl (01.03.2022) | 30,0 | 14,0 | 43,0 |
| Norddahl (01.03.2022) | 27,0 | 9,0 | 41,0 |
| Butt (03.05.2022) | 19,0 | 17,0 | 22,0 |
| Norddahl (01.03.2022) | 14,0 | -37,0 | 47,0 |
| Tseng (01.10.2022) | 13,6 | 4,1 | 22,1 |
| Norddahl (01.03.2022) | 9,0 | -21,0 | 32,0 |
| Norddahl (01.03.2022) | 7,0 | -16,0 | 25,0 |
| Liu (21.06.2022) | 6,9 | 4,8 | 9,0 |
| Tseng (01.10.2022) | -1,1 | -26,1 | 24,5 |
| Tseng (01.10.2022) | -3,9 | -19,7 | 13,0 |

**SF6**: Infection: General population (adults 18+), Exposure 1-3, Reference unvaccinated (n=21)

| Author (Date) | VE in % | upper CI 95% | lower CI 95% |
|---|---|---|---|
| **>180 days post exposure** | | | |
| Arashiro (03.08.2022) | 52,0 | 37,0 | 63,0 |
| Lin (26.09.2022) | 50,7 | 46,3 | 54,7 |
| Lin (26.09.2022) | 49,0 | 46,4 | 51,4 |
| Lin (26.09.2022) | 47,2 | 45,8 | 48,5 |
| Lin (26.09.2022) | 37,5 | 36,0 | 39,0 |
| Lin (26.09.2022) | 30,9 | 28,9 | 32,9 |
| Lin (26.09.2022) | 30,6 | 29,1 | 32,0 |
| Lin (26.09.2022) | 30,3 | 28,4 | 32,2 |
| Lin (26.09.2022) | 26,4 | 24,6 | 28,2 |
| Lind (25.04.2022) | 15,3 | 10,4 | 20,2 |
| Lin (26.09.2022) | 14,5 | 13,0 | 16,0 |
| Tseng (21.04.2022) | 13,8 | 11,0 | 10,2 |
| Šmíd (25.02.2022) | 13,0 | 11,0 | 14,0 |
| Kim (10.04.2022) | 11,0 | -21,0 | 35,0 |
| Castillo (21.04.2022) | 11,0 | 10,0 | 13,0 |
| Kirsbom (01.05.2022) | 8,0 | 6,0 | 9,9 |
| Wang (25.03.2022) | 7,0 | 4,0 | 10,0 |
| Tseng (21.04.2022) | 5,9 | 0,4 | 11,0 |
| Buchan (22.09.2022) | 2,0 | -17,0 | 17,0 |
| Buchan (22.09.2022) | 1,0 | -8,0 | 10,0 |
| Lin (26.09.2022) | 0,8 | -1,5 | 3,0 |
| Chemaitelly (02.06.2022) | -9,0 | -14,5 | -3,7 |
| Chemaitelly (02.06.2022) | -13,7 | -21,3 | -6,6 |
| **91-180 days post exposure** | | | |
| Arashiro (03.08.2022) | 54,0 | 42,0 | 63,0 |
| Carazo (03.05.2022) | 51,0 | 38,0 | 60,0 |
| Carazo (03.05.2022) | 44,0 | 29,0 | 48,0 |
| Carazo (03.05.2022) | 44,0 | 38,0 | 48,0 |
| Chemaitelly (07.07.2022) | 38,1 | 36,3 | 39,8 |
| Gram (01.09.2022) | 31,3 | 30,3 | 32,4 |
| Hansen (30.03.2022) | 27,4 | 26,2 | 28,5 |
| Castillo (21.04.2022) | 26,0 | 24,0 | 27,0 |
| Tseng (21.04.2022) | 23,5 | 14,4 | 30,0 |
| Hansen (30.03.2022) | 23,3 | 21,1 | 25,5 |
| Chemaitelly (02.06.2022) | 18,7 | 11,3 | 25,5 |
| Chemaitelly (02.06.2022) | 16,3 | 9,7 | 22,5 |
| Castillo (21.04.2022) | 16,0 | 15,0 | 17,0 |
| Buchan (22.09.2022) | 15,0 | 8,0 | 22,0 |
| Hansen (30.03.2022) | 13,2 | 12,3 | 14,2 |
| Gram (01.09.2022) | 12,6 | 12,0 | 13,3 |
| Hansen (30.03.2022) | 9,8 | 9,2 | 10,4 |
| Tsang (25.08.2022) | 4,7 | -23,5 | 26,6 |
| Castillo (21.04.2022) | 3,0 | 2,0 | 4,0 |
| Tsang (25.08.2022) | 1,1 | -22,4 | 20,1 |
| **14-90 days post exposure** | | | |
| Arashiro (03.08.2022) | 55,0 | 34,0 | 69,0 |
| Chemaitelly (02.06.2022) | 47,8 | 40,8 | 53,9 |
| Kim (10.04.2022) | 45,0 | 14,0 | 66,0 |
| Tseng (21.04.2022) | 44,0 | 35,1 | 51,6 |
| Chemaitelly (02.06.2022) | 43,2 | 15,0 | 62,1 |
| Castillo (21.04.2022) | 43,0 | 41,0 | 45,0 |
| Šmíd (25.02.2022) | 43,0 | 42,0 | 44,0 |
| Gram (01.09.2022) | 40,0 | 38,6 | 41,3 |
| Hansen (30.03.2022) | 37,9 | 34,4 | 41,2 |
| Hansen (30.03.2022) | 37,0 | 35,6 | 38,0 |
| Buchan (22.09.2022) | 36,0 | 24,0 | 45,0 |
| Tsang (25.08.2022) | 35,3 | -4,2 | 59,8 |
| Gram (01.09.2022) | 32,3 | 30,9 | 33,7 |
| Castillo (21.04.2022) | 32,0 | 30,0 | 34,0 |
| Gram (01.09.2022) | 31,9 | 30,7 | 33,0 |
| Lind (25.04.2022) | 28,5 | 20,0 | 36,2 |
| Hansen (30.03.2022) | 27,4 | 26,3 | 28,4 |
| Hansen (30.03.2022) | 27,1 | 24,5 | 29,6 |
| Castillo (21.04.2022) | 27,0 | 26,0 | 29,0 |
| Hansen (30.03.2022) | 26,8 | 23,8 | 26,9 |
| Hansen (30.03.2022) | 26,6 | 25,3 | 27,9 |
| Wang (25.03.2022) | 26,0 | 22,0 | 30,0 |
| Rennert (07.05.2022) | 25,6 | -17,6 | 52,9 |
| Tsang (25.08.2022) | 20,4 | -30,9 | 51,6 |
| Buchan (22.09.2022) | 12,0 | 3,0 | 21,0 |
| Castillo (21.04.2022) | 12,0 | 9,0 | 14,0 |
| Rennert (07.05.2022) | 7,7 | -11,5 | 23,5 |
| **<14 days post exposure** | | | |
| Lind (25.04.2022) | 23,1 | 15,2 | 30,2 |
| Chemaitelly (02.06.2022) | 9,8 | -94,1 | 58,1 |
| Chemaitelly (02.06.2022) | 9,5 | -39,8 | 41,3 |
| **No information** | | | |
| Carazo (03.05.2022) | 65,0 | 63,0 | 67,0 |
| Lin (26.09.2022) | 52,3 | 49,3 | 55,1 |
| Castillo (21.04.2022) | 51,0 | 50,0 | 52,0 |
| Altarawneh (15.06.2022) | 50,8 | 45,4 | 55,7 |
| Altarawneh (15.06.2022) | 50,2 | 38,1 | 59,9 |
| Lin (26.09.2022) | 46,4 | 45,3 | 47,5 |
| Altarawneh (15.06.2022) | 46,1 | 39,5 | 51,9 |
| Lin (26.09.2022) | 43,2 | 42,0 | 44,4 |
| Carazo (03.05.2022) | 42,0 | 41,0 | 44,0 |
| Wang (25.03.2022) | 39,0 | 36,0 | 42,0 |
| Chemaitelly (02.06.2022) | 31,4 | 12,5 | 46,3 |
| Arashiro (03.08.2022) | 29,0 | -32,0 | 62,0 |
| Mimura (08.10.2022) | 21,2 | 11,0 | 30,3 |
| Kim (10.04.2022) | 21,0 | -6,0 | 41,0 |
| Tseng (21.04.2022) | 20,4 | 9,5 | 30,0 |
| Carazo (03.05.2022) | 20,0 | 16,0 | 24,0 |
| Risk (16.08.2022) | 16,0 | 5,0 | 26,0 |
| Mimura (08.10.2022) | 15,8 | 7,9 | 23,1 |
| Young-Xu (03.08.2022) | 12,0 | 10,0 | 15,0 |
| Chemaitelly (02.06.2022) | 9,5 | -39,9 | 41,5 |
| Altarawneh (15.06.2022) | -0,2 | -5,5 | 4,9 |
| Altarawneh (15.06.2022) | -1,1 | -7,1 | 4,6 |
| Altarawneh (15.06.2022) | -4,9 | -16,4 | 5,4 |
| Risk (16.08.2022) | -6,0 | -16,0 | 4,0 |
| Mimura (08.10.2022) | -24,4 | -112,3 | 27,1 |
| Mimura (08.10.2022) | -45,6 | -111,4 | 0,3 |

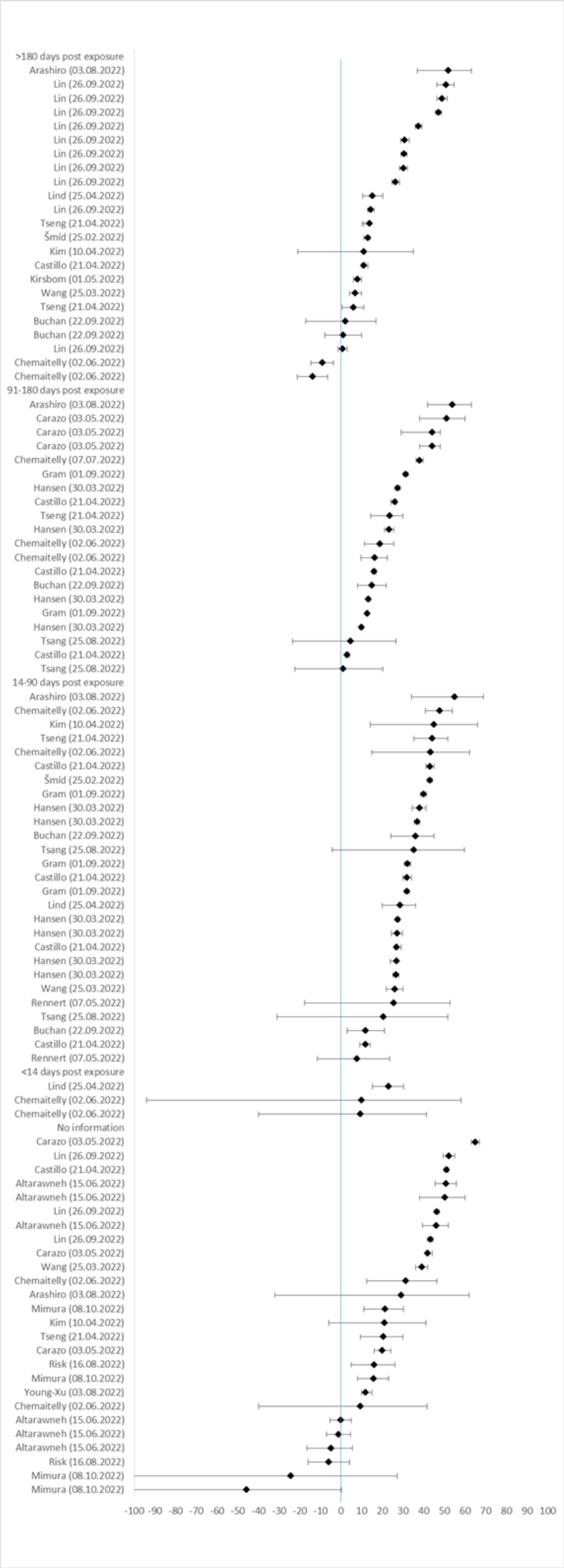
>180 days post exposure
Arashiro (03.08.2022)
Lin (26.09.2022)
Lin (26.09.2022)
Lin (26.09.2022)
Lin (26.09.2022)
Lin (26.09.2022)
Lin (26.09.2022)
Lin (26.09.2022)
Lin (26.09.2022)
Lind (25.04.2022)
Lin (26.09.2022)
Tseng (21.04.2022)
Šmíd (25.02.2022)
Kim (10.04.2022)
Castillo (21.04.2022)
Kirsbom (01.05.2022)
Wang (25.03.2022)
Tseng (21.04.2022)
Buchan (22.09.2022)
Buchan (22.09.2022)
Lin (26.09.2022)
Chemaitelly (02.06.2022)
Chemaitelly (02.06.2022)
91-180 days post exposure
Arashiro (03.08.2022)
Carazo (03.05.2022)
Carazo (03.05.2022)
Carazo (03.05.2022)
Chemaitelly (07.07.2022)
Gram (01.09.2022)
Hansen (30.03.2022)
Castillo (21.04.2022)
Tseng (21.04.2022)
Hansen (30.03.2022)
Chemaitelly (02.06.2022)
Chemaitelly (02.06.2022)
Castillo (21.04.2022)
Buchan (22.09.2022)
Hansen (30.03.2022)
Gram (01.09.2022)
Hansen (30.03.2022)
Tsang (25.08.2022)
Castillo (21.04.2022)
Tsang (25.08.2022)
14-90 days post exposure
Arashiro (03.08.2022)
Chemaitelly (02.06.2022)
Kim (10.04.2022)
Tseng (21.04.2022)
Chemaitelly (02.06.2022)
Castillo (21.04.2022)
Šmíd (25.02.2022)
Gram (01.09.2022)
Hansen (30.03.2022)
Hansen (30.03.2022)
Buchan (22.09.2022)
Tsang (25.08.2022)
Gram (01.09.2022)
Castillo (21.04.2022)
Gram (01.09.2022)
Lind (25.04.2022)
Hansen (30.03.2022)
Hansen (30.03.2022)
Castillo (21.04.2022)
Hansen (30.03.2022)
Hansen (30.03.2022)
Wang (25.03.2022)
Rennert (07.05.2022)
Tsang (25.08.2022)
Buchan (22.09.2022)
Castillo (21.04.2022)
Rennert (07.05.2022)
<14 days post exposure
Lind (25.04.2022)
Chemaitelly (02.06.2022)
Chemaitelly (02.06.2022)
No information
Carazo (03.05.2022)
Lin (26.09.2022)
Castillo (21.04.2022)
Altarawneh (15.06.2022)
Altarawneh (15.06.2022)
Lin (26.09.2022)
Altarawneh (15.06.2022)
Lin (26.09.2022)
Carazo (03.05.2022)
Wang (25.03.2022)
Chemaitelly (02.06.2022)
Arashiro (03.08.2022)
Mimura (08.10.2022)
Kim (10.04.2022)
Tseng (21.04.2022)
Carazo (03.05.2022)
Risk (16.08.2022)
Mimura (08.10.2022)
Young-Xu (03.08.2022)
Chemaitelly (02.06.2022)
Altarawneh (15.06.2022)
Altarawneh (15.06.2022)
Altarawneh (15.06.2022)
Risk (16.08.2022)
Mimura (08.10.2022)
Mimura (08.10.2022)
-100 -90 -80 -70 -60 -50 -40 -30 -20 -10 0 10 20 30 40 50 60 70 80 90 100

**SF7**: Infection: Older adults (60+ years), Exposure 3+, Reference unvaccinated (n=4)

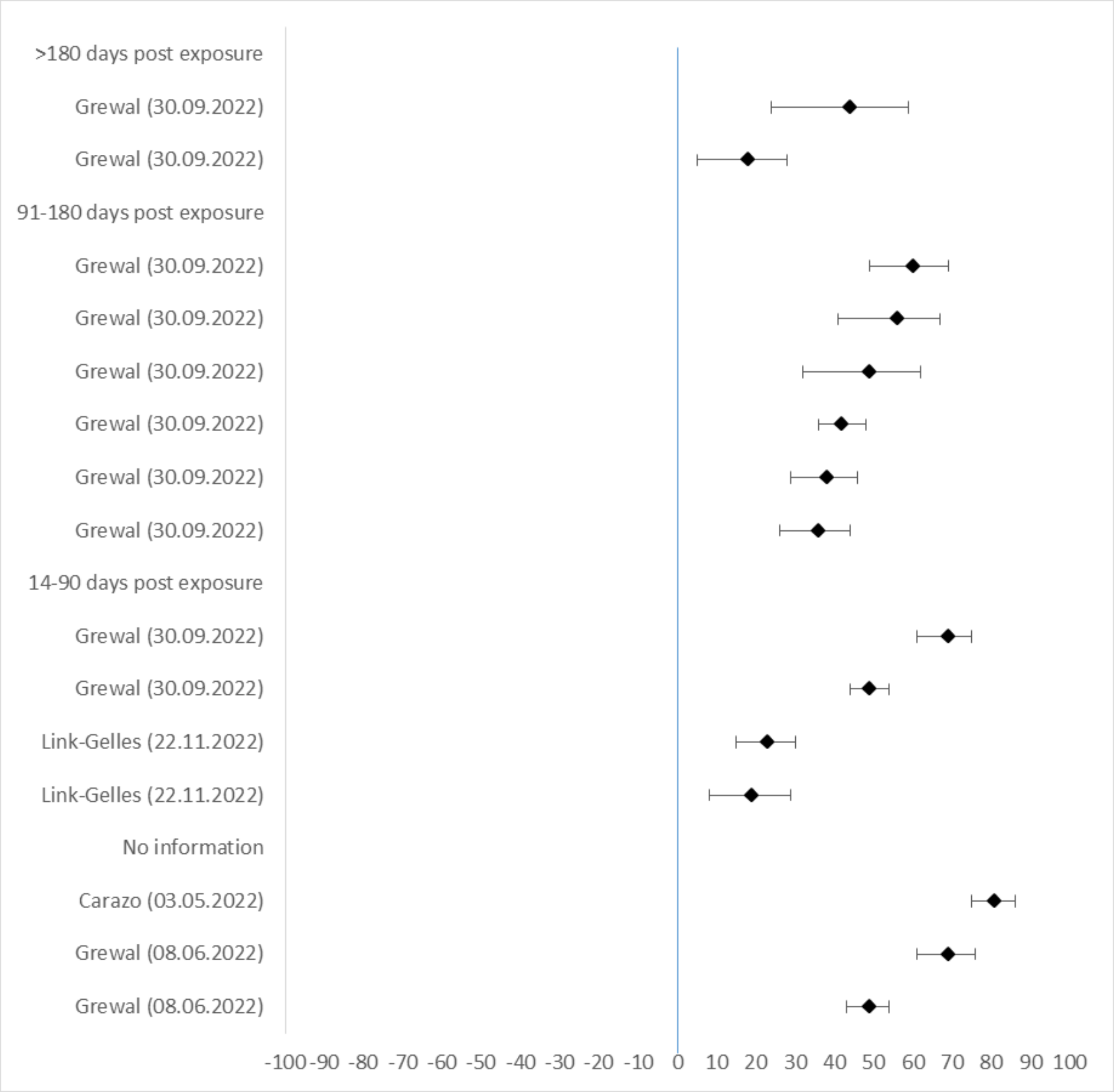

| Author (Date) | VE in % | upper CI 95% | lower CI 95% |
|---|---|---|---|
| **>180 days post exposure** | | | |
| Grewal (30.09.2022) | 44,0 | 24,0 | 59,0 |
| Grewal (30.09.2022) | 18,0 | 5,0 | 28,0 |
| **91-180 days post exposure** | | | |
| Grewal (30.09.2022) | 60,0 | 49,0 | 69,0 |
| Grewal (30.09.2022) | 56,0 | 41,0 | 67,0 |
| Grewal (30.09.2022) | 49,0 | 32,0 | 62,0 |
| Grewal (30.09.2022) | 42,0 | 36,0 | 48,0 |
| Grewal (30.09.2022) | 38,0 | 29,0 | 46,0 |
| Grewal (30.09.2022) | 36,0 | 26,0 | 44,0 |
| **14-90 days post exposure** | | | |
| Grewal (30.09.2022) | 69,0 | 61,0 | 75,0 |
| Grewal (30.09.2022) | 49,0 | 44,0 | 54,0 |
| Link-Gelles (22.11.2022) | 23,0 | 15,0 | 30,0 |
| Link-Gelles (22.11.2022) | 19,0 | 8,0 | 29,0 |
| **No information** | | | |
| Carazo (03.05.2022) | 81,0 | 75,0 | 86,0 |
| Grewal (08.06.2022) | 69,0 | 61,0 | 76,0 |
| Grewal (08.06.2022) | 49,0 | 43,0 | 54,0 |

**SF8**: Infection: Older adults (60+ years), Exposure 3+, Reference less exposures (3 exposures) (n=8)

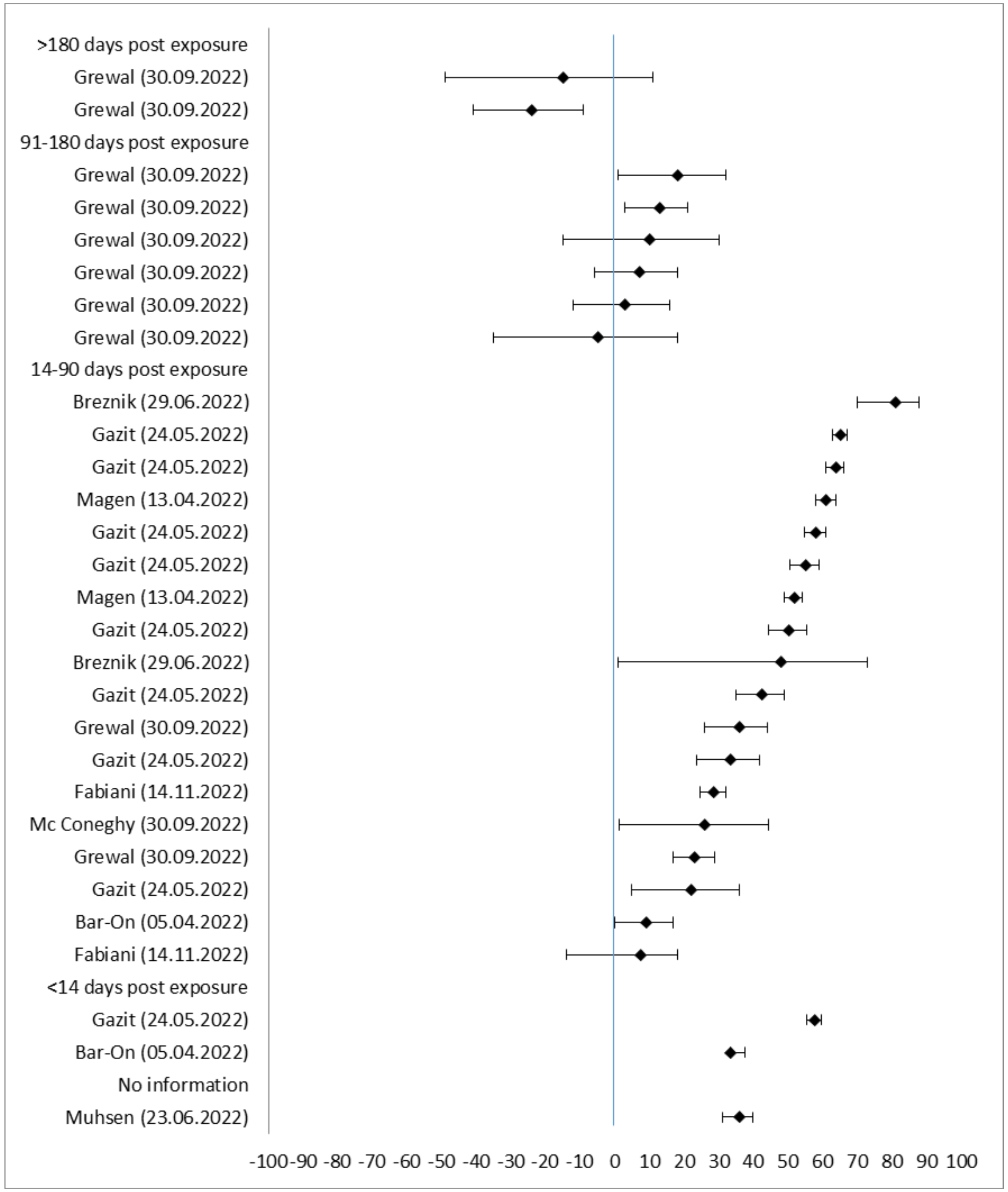

| Author (date) | VE in % | lower CI 95% | upper CI 95% |
|---|---|---|---|
| **>180 days post exposure** | | | |
| Grewal (30.09.2022) | -15,0 | -49,0 | 11,0 |
| Grewal (30.09.2022) | -24,0 | -41,0 | -9,0 |
| **91-180 days post exposure** | | | |
| Grewal (30.09.2022) | 18,0 | 1,0 | 32,0 |
| Grewal (30.09.2022) | 13,0 | 3,0 | 21,0 |
| Grewal (30.09.2022) | 10,0 | -15,0 | 30,0 |
| Grewal (30.09.2022) | 7,0 | -6,0 | 18,0 |
| Grewal (30.09.2022) | 3,0 | -12,0 | 16,0 |
| Grewal (30.09.2022) | -5,0 | -35,0 | 18,0 |
| **14-90 days post exposure** | | | |
| Breznik (29.06.2022) | 81,0 | 70,0 | 88,0 |
| Gazit (24.05.2022) | 65,1 | 63,0 | 67,1 |
| Gazit (24.05.2022) | 64,0 | 61,1 | 66,3 |
| Magen (13.04.2022) | 61,0 | 58,0 | 64,0 |
| Gazit (24.05.2022) | 58,1 | 54,8 | 61,1 |
| Gazit (24.05.2022) | 55,0 | 50,6 | 58,9 |
| Magen (13.04.2022) | 52,0 | 49,0 | 54,0 |
| Gazit (24.05.2022) | 50,2 | 44,5 | 55,3 |
| Breznik (29.06.2022) | 48,0 | 1,0 | 73,0 |
| Gazit (24.05.2022) | 42,5 | 35,1 | 49,1 |
| Grewal (30.09.2022) | 36,0 | 26,0 | 44,0 |
| Gazit (24.05.2022) | 33,4 | 23,8 | 41,8 |
| Fabiani (14.11.2022) | 28,5 | 24,7 | 32,1 |
| Mc Coneghy (30.09.2022) | 25,8 | 1,2 | 44,3 |
| Grewal (30.09.2022) | 23,0 | 17,0 | 29,0 |
| Gazit (24.05.2022) | 22,0 | 4,9 | 36,1 |
| Bar-On (05.04.2022) | 9,2 | 0,0 | 16,7 |
| Fabiani (14.11.2022) | 7,6 | -14,1 | 18,3 |
| **<14 days post exposure** | | | |
| Gazit (24.05.2022) | 57,7 | 55,6 | 59,7 |
| Bar-On (05.04.2022) | 33,3 | 33,3 | 37,5 |
| **No information** | | | |
| Muhsen (23.06.2022) | 36,0 | 31,0 | 40,0 |

**SF9**: Infection: Older adults (60+years), Exposure 3, Reference unvaccinated (n=9)

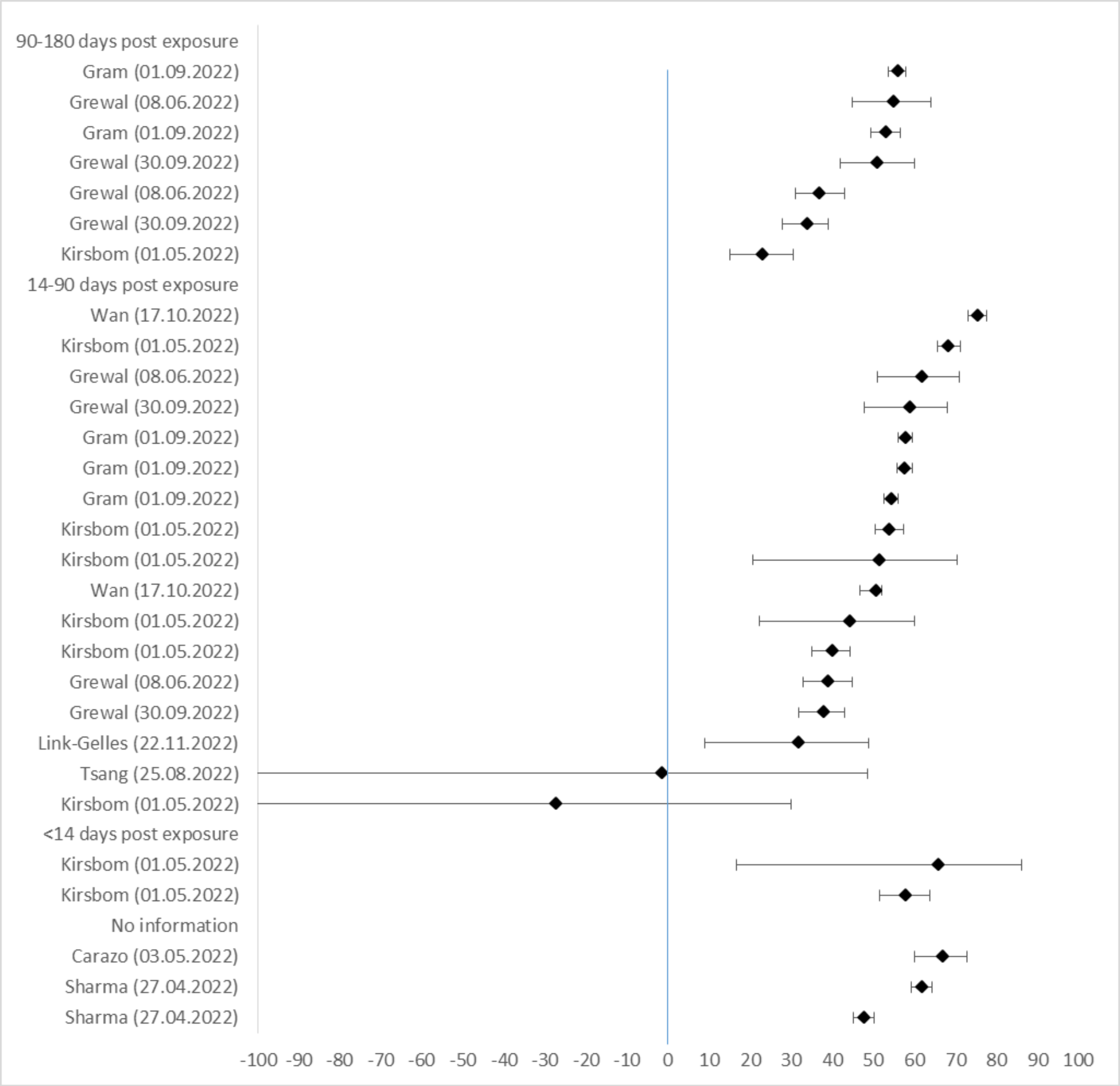

| Author (Date) | VE in % | lower CI 95% | upper CI 95% |
|---|---|---|---|
| **91-180 days post exposure** | | | |
| Gram (01.09.2022) | 56,0 | 53,7 | 58,1 |
| Grewal (08.06.2022) | 55,0 | 45,0 | 64,0 |
| Gram (01.09.2022) | 53,2 | 49,6 | 56,6 |
| Grewal (30.09.2022) | 51,0 | 42,0 | 60,0 |
| Grewal (08.06.2022) | 37,0 | 31,0 | 43,0 |
| Grewal (30.09.2022) | 34,0 | 28,0 | 39,0 |
| Kirsbom (01.05.2022) | 23,1 | 15,1 | 30,5 |
| **14-90 days post exposure** | | | |
| Wan (17.10.2022) | 75,6 | 73,2 | 77,8 |
| Kirsbom (01.05.2022) | 68,5 | 65,7 | 71,2 |
| Grewal (08.06.2022) | 62,0 | 51,0 | 71,0 |
| Grewal (30.09.2022) | 59,0 | 48,0 | 68,0 |
| Gram (01.09.2022) | 57,9 | 56,1 | 59,6 |
| Gram (01.09.2022) | 57,7 | 55,9 | 59,5 |
| Gram (01.09.2022) | 54,4 | 52,7 | 56,0 |
| Kirsbom (01.05.2022) | 54,1 | 50,5 | 57,5 |
| Kirsbom (01.05.2022) | 51,6 | 20,8 | 70,4 |
| Wan (17.10.2022) | 50,9 | 46,9 | 52,1 |
| Kirsbom (01.05.2022) | 44,5 | 22,4 | 60,2 |
| Kirsbom (01.05.2022) | 40,1 | 35,2 | 44,5 |
| Grewal (08.06.2022) | 39,0 | 33,0 | 45,0 |
| Grewal (30.09.2022) | 38,0 | 32,0 | 43,0 |
| Link-Gelles (22.11.2022) | 32,0 | 9,00 | 49,00 |
| Tsang (25.08.2022) | -1,5 | -101,0 | 48,7 |
| Kirsbom (01.05.2022) | -27,2 | -131,6 | 30,1 |
| **<14 days post exposure** | | | |
| Kirsbom (01.05.2022) | 66,1 | 16,6 | 86,3 |
| Kirsbom (01.05.2022) | 58,1 | 51,6 | 63,8 |
| **No information** | | | |
| Carazo (03.05.2022) | 67,0 | 60,0 | 73,0 |
| Sharma (27.04.2022) | 61,9 | 59,4 | 64,4 |
| Sharma (27.04.2022) | 47,8 | 45,2 | 50,3 |

**SF10**: Infection: Older adults (60+ years), Exposure 3, Reference less exposures (2 exposures) (n=4)

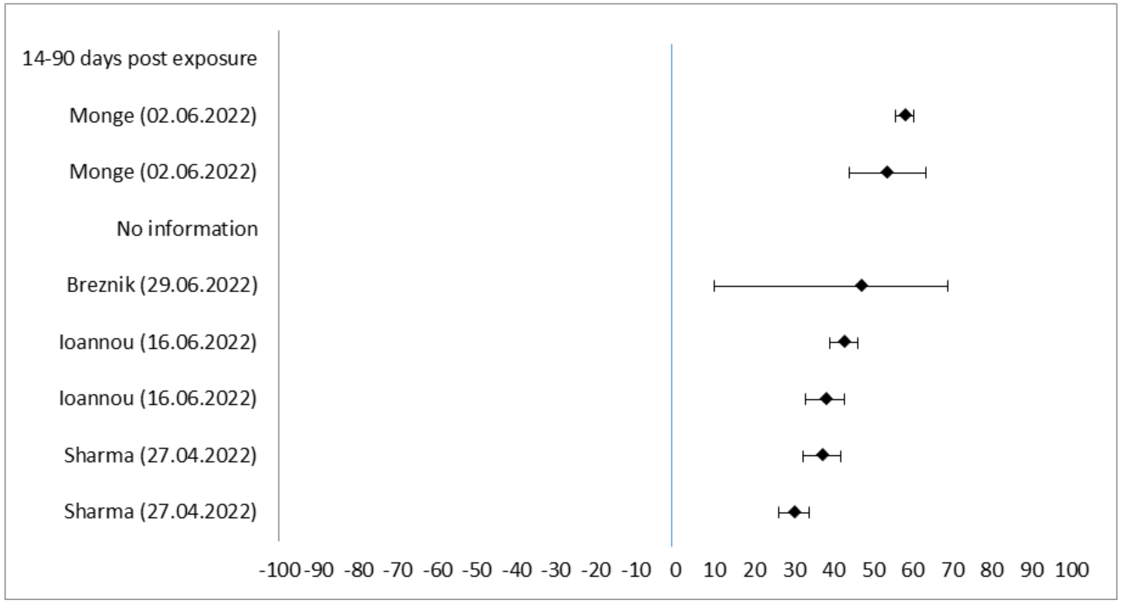

| Author (date) | VE in % | lower CI 95% | upper CI 95% |
|---|---|---|---|
| **14-90 days post exposure** | | | |
| Monge (02.06.2022) | 58,0 | 55,8 | 60,4 |
| Monge (02.06.2022) | 53,5 | 43,9 | 63,3 |
| **No information** | | | |
| Breznik (29.06.2022) | 47,0 | 10,0 | 69,0 |
| Ioannou (16.06.2022) | 42,8 | 39,1 | 46,2 |
| Ioannou (16.06.2022) | 38,1 | 33,0 | 42,8 |
| Sharma (27.04.2022) | 37,1 | 32,2 | 41,7 |
| Sharma (27.04.2022) | 30,1 | 26,2 | 33,7 |

**SF11**: Infection: Older adults (60+ years), Exposure 1-3, Reference unvaccinated (n=8)

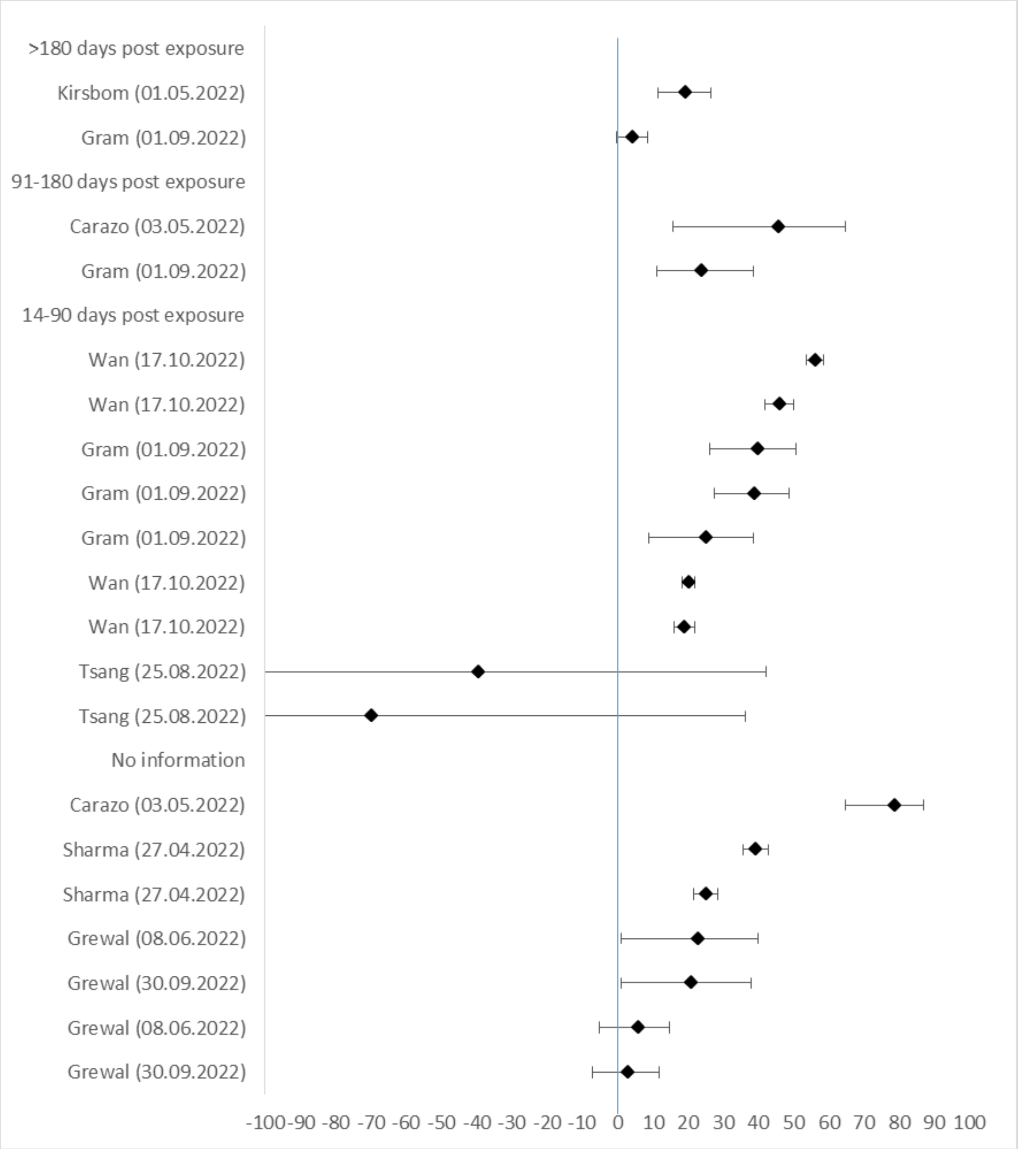

| Author (Date) | VE in % | lower CI 95% | upper CI 95% |
|---|---|---|---|
| **>180 days post exposure** | | | |
| Kirsbom (01.05.2022) | 19,5 | 11,7 | 26,6 |
| Gram (01.09.2022) | 4,4 | -0,1 | 8,7 |
| **91-180 days post exposure** | | | |
| Carazo (03.05.2022) | 46,0 | 16,0 | 65,0 |
| Gram (01.09.2022) | 24,0 | 11,4 | 38,8 |
| **14-90 days post exposure** | | | |
| Wan (17.10.2022) | 56,2 | 53,8 | 58,5 |
| Wan (17.10.2022) | 46,2 | 42,1 | 50 |
| Gram (01.09.2022) | 39,9 | 26,3 | 50,9 |
| Gram (01.09.2022) | 39,0 | 27,6 | 48,7 |
| Gram (01.09.2022) | 25,2 | 9,0 | 38,6 |
| Wan (17.10.2022) | 20,3 | 18,4 | 22,1 |
| Wan (17.10.2022) | 19,2 | 16,2 | 22 |
| Tsang (25.08.2022) | -39,3 | -236,4 | 42,4 |
| Tsang (25.08.2022) | -69,7 | -353,1 | 36,5 |
| **No information** | | | |
| Carazo (03.05.2022) | 79,0 | 65,0 | 87,0 |
| Sharma (27.04.2022) | 39,5 | 35,8 | 43,0 |
| Sharma (27.04.2022) | 25,3 | 21,8 | 28,7 |
| Grewal (08.06.2022) | 23,0 | 1,0 | 40,0 |
| Grewal (30.09.2022) | 21,0 | 1,0 | 38,0 |
| Grewal (08.06.2022) | 6,0 | -5,0 | 15,0 |
| Grewal (30.09.2022) | 3,0 | -7,0 | 12,0 |

**SF12:** Infection: Children and adolescents (5-17 years), Exposure 3+, Reference unvaccinated (n=2)

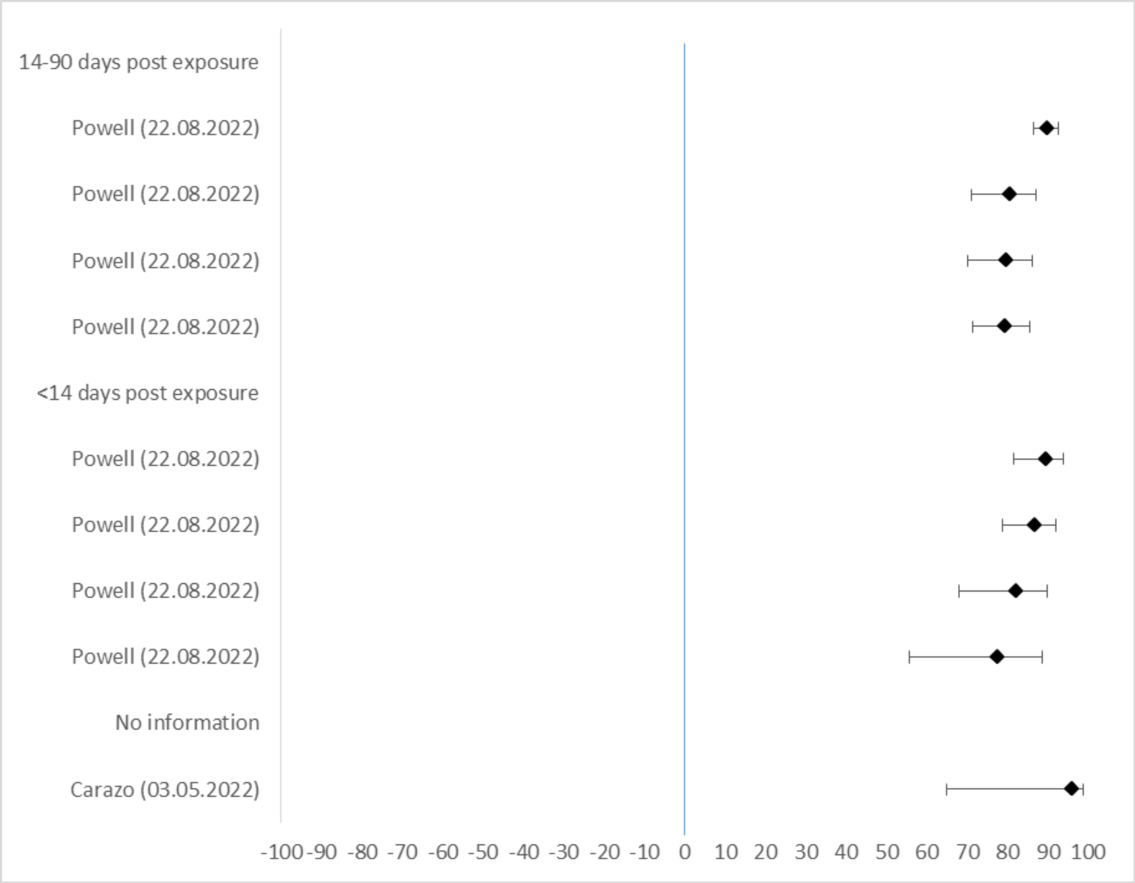

| Author (Date) | VE in % | lower CI 95% | upper CI 95% |
|---|---|---|---|
| **14-90 days post exposure** | | | |
| Powell (22.08.2022) | 90,1 | 86,6 | 92,7 |
| Powell (22.08.2022) | 80,7 | 71,1 | 87,1 |
| Powell (22.08.2022) | 79,8 | 70,4 | 86,3 |
| Powell (22.08.2022) | 79,6 | 71,4 | 85,5 |
| **<14 days post exposure** | | | |
| Powell (22.08.2022) | 89,5 | 81,7 | 94,0 |
| Powell (22.08.2022) | 87,0 | 78,8 | 92,0 |
| Powell (22.08.2022) | 82,2 | 68,1 | 90,1 |
| Powell (22.08.2022) | 77,7 | 55,7 | 88,8 |
| **No information** | | | |
| Carazo (03.05.2022) | 96,0 | 65,0 | 99,0 |

**SF13**: Infection: Children and adolescents (5-17 years), Exposure 3, Reference unvaccinated (n=7)

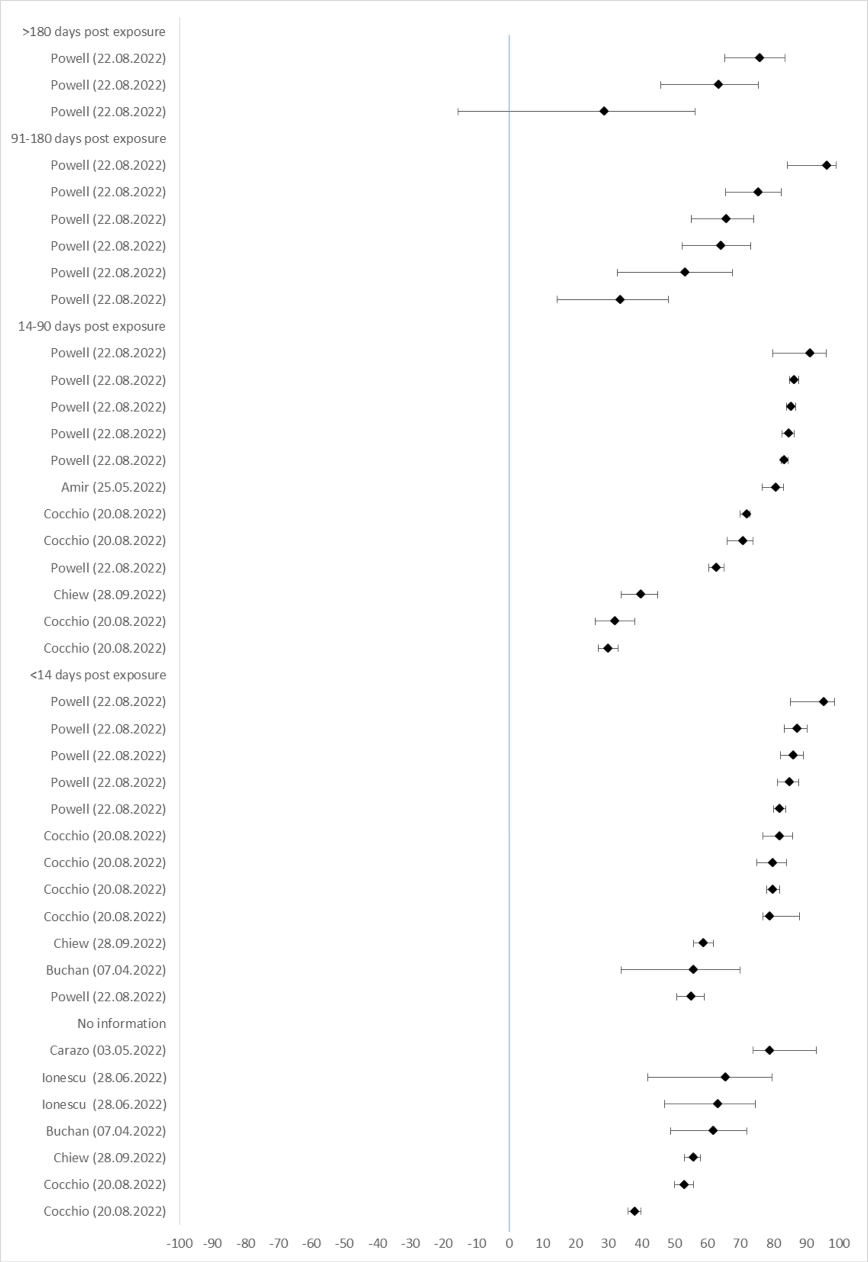

| Author (Date) | VE in % | lower CI 95% | upper CI 95% |
|---|---|---|---|
| **>180 days post exposure** | | | |
| Powell (22.08.2022) | 76,1 | 65,3 | 83,6 |
| Powell (22.08.2022) | 63,6 | 46,0 | 75,5 |
| Powell (22.08.2022) | 28,9 | -15,5 | 56,3 |
| **91-180 days post exposure** | | | |
| Powell (22.08.2022) | 96,4 | 84,4 | 99,1 |
| Powell (22.08.2022) | 75,5 | 65,6 | 82,5 |
| Powell (22.08.2022) | 65,9 | 55,2 | 74,1 |
| Powell (22.08.2022) | 64,3 | 52,4 | 73,3 |
| Powell (22.08.2022) | 53,4 | 32,7 | 67,7 |
| Powell (22.08.2022) | 33,6 | 14,6 | 48,3 |
| **14-90 days post exposure** | | | |
| Powell (22.08.2022) | 91,2 | 80,0 | 96,1 |
| Powell (22.08.2022) | 86,5 | 85,1 | 87,8 |
| Powell (22.08.2022) | 85,5 | 84,0 | 86,9 |
| Powell (22.08.2022) | 84,7 | 82,6 | 86,5 |
| Powell (22.08.2022) | 83,5 | 82,5 | 84,5 |
| Amir (25.05.2022) | 80,8 | 76,7 | 83,1 |
| Cocchio (20.08.2022) | 72,0 | 70,0 | 73,0 |
| Cocchio (20.08.2022) | 71,0 | 66,0 | 74,0 |
| Powell (22.08.2022) | 62,9 | 60,5 | 65,1 |
| Chiew (28.09.2022) | 40,0 | 34,0 | 45,0 |
| Cocchio (20.08.2022) | 32,0 | 26,0 | 38,0 |
| Cocchio (20.08.2022) | 30,0 | 27,0 | 33,0 |
| **<14 days post exposure** | | | |
| Powell (22.08.2022) | 95,5 | 85,3 | 98,6 |
| Powell (22.08.2022) | 87,4 | 83,5 | 90,4 |
| Powell (22.08.2022) | 86,1 | 82,3 | 89,1 |
| Powell (22.08.2022) | 84,9 | 81,3 | 87,8 |
| Powell (22.08.2022) | 82,1 | 80,1 | 83,9 |
| Cocchio (20.08.2022) | 82,0 | 77,0 | 86,0 |
| Cocchio (20.08.2022) | 80,0 | 75,0 | 84,0 |
| Cocchio (20.08.2022) | 80,0 | 78,0 | 82,0 |
| Cocchio (20.08.2022) | 79,0 | 77,0 | 88,0 |
| Chiew (28.09.2022) | 59,0 | 56,0 | 62,0 |
| Buchan (07.04.2022) | 56,0 | 34,0 | 70,0 |
| Powell (22.08.2022) | 55,1 | 50,7 | 59,1 |
| **No information** | | | |
| Carazo (03.05.2022) | 79,0 | 74,0 | 93,0 |
| Ionescu (28.06.2022) | 65,7 | 41,9 | 79,8 |
| Ionescu (28.06.2022) | 63,3 | 47,2 | 74,6 |
| Buchan (07.04.2022) | 62,0 | 49,0 | 72,0 |
| Chiew (28.09.2022) | 56,0 | 53,0 | 58,0 |
| Cocchio (20.08.2022) | 53,0 | 50,0 | 56,0 |
| Cocchio (20.08.2022) | 38,0 | 36,0 | 40,0 |

**SF14**: Infection: Children and adolescents (5-17 years), Exposure 3, Reference less exposures (2 exposures) (n=1)

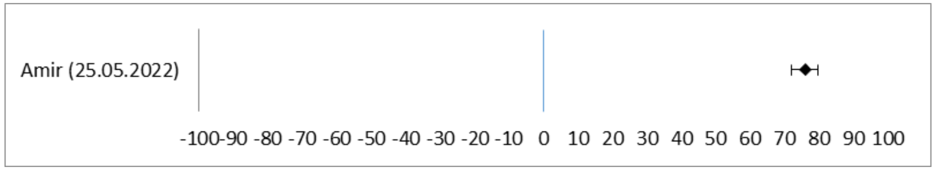

| Author (date) | VE in % | lower CI 95% | upper CI 95% |
|---|---|---|---|
| **14-90 days post exposure** | | | |
| Amir (25.05.2022) | 76,2 | 72,2 | 79,6 |

**SF15**: Infection: Children and adolescents (5-17 years), Exposure 1-3, Reference unvaccinated (n=17)

| **Author (Date)** | **VE in %** | **lower CI 95%** | **upper CI 95%** |
|---|---|---|---|
| **>180 days post exposure** | | | |
| Powell (22.08.2022) | 90,2 | 75,9 | 96,0 |
| Powell (22.08.2022) | 77,2 | 67,5 | 84,0 |
| Powell (22.08.2022) | 67,8 | 24,1 | 86,3 |
| Powell (22.08.2022) | 66,7 | 35,2 | 82,9 |
| Powell (22.08.2022) | 55,8 | 17,2 | 76,4 |
| Buchan (07.04.2022) | 29,0 | 17,0 | 38,0 |
| Powell (22.08.2022) | 25,7 | -4,2 | 47,0 |
| Powell (22.08.2022) | 19,4 | 11,7 | 26,4 |
| Powell (22.08.2022) | 14,7 | -63,3 | 55,4 |
| Powell (22.08.2022) | 12,8 | -1,6 | 25,1 |
| Nordstrom (24.10.2022) | 4,0 | 2,0 | 6,0 |
| Chemaitelly (26.07.2022) | -1,7 | -20,0 | 10,0 |
| **91-180 days post exposure** | | | |
| Powell (22.08.2022) | 85,5 | 77,5 | 90,6 |
| Powell (22.08.2022) | 76,2 | 63,7 | 70,3 |
| Powell (22.08.2022) | 74,4 | 67,2 | 78,4 |
| Powell (22.08.2022) | 69,5 | 64,5 | 73,8 |
| Powell (22.08.2022) | 65,9 | 60,5 | 70,6 |
| Fowlkes (11.03.2022) | 62,0 | -28,0 | 89,0 |
| Powell (22.08.2022) | 59,3 | 46,7 | 69,0 |
| Carazo (03.05.2022) | 57,0 | 36,0 | 71,0 |
| Powell (22.08.2022) | 52,4 | 50,9 | 53,8 |
| Ionescu (28.06.2022) | 50,9 | 44,9 | 56,3 |
| Florentino (08.08.2022) | 50,6 | 42,7 | 57,4 |
| Rudan (28.09.2022) | 48,7 | 22,0 | 66,3 |
| Ionescu (28.06.2022) | 47,5 | 37,6 | 55,8 |
| Ionescu (28.06.2022) | 46,0 | 40,9 | 50,7 |
| Ionescu (28.06.2022) | 44,6 | 40,0 | 49,0 |
| Ionescu (28.06.2022) | 37,7 | 22,7 | 49,7 |
| Powell (22.08.2022) | 36,6 | 32,9 | 40,1 |
| Nordstrom (24.10.2022) | 36,0 | 34,0 | 23,0 |
| Piche-Renaud (01.08.2022) | 35,0 | 21,0 | 46,0 |
| Ionescu (28.06.2022) | 33,9 | 24,1 | 42,4 |
| Ionescu (28.06.2022) | 33,9 | 27,4 | 39,9 |
| Powell (22.08.2022) | 32,7 | 27,7 | 37,4 |
| Powell (22.08.2022) | 29,8 | 24,9 | 34,2 |
| Buchan (07.04.2022) | 29,0 | 19,0 | 38,0 |
| Tan (20.07.2022) | 25,6 | 19,3 | 31,5 |
| Veneti (25.03.2022) | 23,3 | 2,7 | 39,5 |
| Cocchio (20.08.2022) | 23,0 | 20,0 | 26,0 |
| Ionescu (28.06.2022) | 22,2 | 8,4 | 33,9 |
| Nordstrom (24.10.2022) | 21,0 | 19,0 | 23,0 |
| Cocchio (20.08.2022) | 20,0 | 15,0 | 24,0 |
| Powell (22.08.2022) | 17,9 | 14,9 | 20,7 |
| Nordstrom (24.10.2022) | 10,0 | 8,0 | 12,0 |
| Rudan (28.09.2022) | 9,5 | -3,6 | 20,9 |
| Cocchio (20.08.2022) | 8,0 | 5,0 | 11,0 |
| Florentino (08.08.2022) | 5,9 | 2,2 | 9,4 |
| Rudan (28.09.2022) | 5,4 | -13,4 | 21,0 |
| Piche-Renaud (01.08.2022) | 4,0 | -12,0 | 18,0 |
| Rudan (28.09.2022) | 1,2 | -49,3 | 34,6 |
| Veneti (25.03.2022) | -5,3 | -32,9 | 16,6 |
| Chemaitelly (26.07.2022) | -9,5 | -30,0 | 35,0 |
| Veneti (25.03.2022) | -12,8 | -21,7 | -4,6 |
| Rudan (28.09.2022) | -24,2 | -46,5 | -5,3 |
| Rudan (28.09.2022) | -24,7 | -46,7 | -6,0 |
| **14-90 days post exposure** | | | |
| Powell (22.08.2022) | 85,3 | 83,7 | 86,8 |
| Florentino (08.08.2022) | 82,6 | 80,6 | 84,5 |
| Powell (22.08.2022) | 81,5 | 80,0 | 82,9 |
| Rudan (28.09.2022) | 81,2 | 77,7 | 84,2 |
| Powell (22.08.2022) | 79,6 | 44,9 | 92,4 |
| Powell (22.08.2022) | 78,8 | 77,9 | 79,5 |
| Florentino (08.08.2022) | 77,4 | 74,7 | 79,8 |
| Ionescu (28.06.2022) | 75,6 | 65,8 | 82,6 |
| Florentino (08.08.2022) | 69,6 | 66,3 | 72,6 |
| Rudan (28.09.2022) | 68,5 | 63,4 | 72,9 |
| Piche-Renaud (01.08.2022) | 67,0 | 60,0 | 72,0 |
| Rudan (28.09.2022) | 65,5 | 56,0 | 73,0 |
| Florentino (08.08.2022) | 65,4 | 61,9 | 68,7 |
| Florentino (08.08.2022) | 64,7 | 63,0 | 66,3 |
| Powell (22.08.2022) | 64,5 | 63,6 | 65,4 |
| Ionescu (28.06.2022) | 63,4 | 21,4 | 83,0 |
| Ionescu (28.06.2022) | 59,3 | 50,9 | 66,3 |
| Fowlkes (11.03.2022) | 59,0 | 22,0 | 79,0 |
| Cocchio (20.08.2022) | 59,0 | 55,0 | 62,0 |
| Amir (25.05.2022) | 58,3 | 54,6 | 61,5 |
| Florentino (08.08.2022) | 58,0 | 52,9 | 62,6 |
| Ionescu (28.06.2022) | 57,7 | 37,2 | 71,6 |
| Cocchio (20.08.2022) | 55,0 | 49,0 | 61,0 |
| Veneti (25.03.2022) | 53,1 | 42,6 | 61,7 |
| Florentino (08.08.2022) | 53,0 | 51,3 | 54,7 |
| Cocchio (20.08.2022) | 53,0 | 51,0 | 55,0 |
| Chemaitelly (26.07.2022) | 51,3 | 34,9 | 63,6 |
| Powell (22.08.2022) | 51,2 | 28,4 | 66,8 |
| Cohen-Stavi (29.06.2022) | 51,0 | 39,0 | 61,0 |
| Buchan (07.04.2022) | 51,0 | 38,0 | 61,0 |
| Chemaitelly (26.07.2022) | 49,6 | 28,5 | 64,5 |
| Ionescu (28.06.2022) | 48,1 | 39,9 | 55,1 |
| Cohen-Stavi (29.06.2022) | 48,0 | 29,0 | 63,0 |
| Nordstrom (24.10.2022) | 47,0 | 43,0 | 51,0 |
| Veneti (25.03.2022) | 45,7 | 34,8 | 54,7 |
| Florentino (08.08.2022) | 45,3 | 37,2 | 52,4 |
| Rudan (28.09.2022) | 43,4 | 26,9 | 56,2 |
| Rudan (28.09.2022) | 43,3 | 30,0 | 54,2 |
| Ionescu (28.06.2022) | 40,8 | 23,2 | 54,4 |
| Florentino (08.08.2022) | 40,6 | 38,8 | 42,4 |
| Tan (20.07.2022) | 37,6 | 35,7 | 39,3 |
| Tsang (25.08.2022) | 32,4 | -29,0 | 64,6 |
| Florentino (08.08.2022) | 32,0 | 30,0 | 33,9 |
| Fowlkes (11.03.2022) | 31,0 | 9,0 | 48,0 |
| Buchan (07.04.2022) | 31,0 | 20,0 | 41,0 |
| Rudan (28.09.2022) | 30,2 | 18,4 | 40,3 |
| Cocchio (20.08.2022) | 29,0 | 23,0 | 35,0 |
| Tan (20.07.2022) | 28,5 | 26,3 | 30,7 |
| Florentino (08.08.2022) | 25,3 | 22,9 | 27,6 |
| Piche-Renaud (01.08.2022) | 23,0 | 7,0 | 36,0 |
| Cocchio (20.08.2022) | 23,0 | 19,0 | 27,0 |
| Rudan (28.09.2022) | 22,8 | -6,4 | 44,0 |
| Cocchio (20.08.2022) | 22,0 | 19,0 | 24,0 |
| Rudan (28.09.2022) | 21,8 | 11,5 | 30,8 |
| Veneti (25.03.2022) | 21,2 | -5,1 | 53,8 |
| Powell (22.08.2022) | 18,8 | 17,2 | 20,3 |
| Cohen-Stavi (29.06.2022) | 18,0 | -2,0 | 34,0 |
| Cohen-Stavi (29.06.2022) | 17,0 | 7,0 | 25,0 |
| Florentino (08.08.2022) | 17,0 | 13,8 | 20,0 |
| Rudan (28.09.2022) | 16,9 | 8,7 | 24,4 |
| Veneti (25.03.2022) | 16,2 | -2,4 | 31,3 |
| Veneti (25.03.2022) | 15,1 | -56,0 | 53,8 |
| Rudan (28.09.2022) | 11,9 | -16,1 | 33,1 |
| Rudan (28.09.2022) | 19,1 | 8,9 | 30,3 |
| Tsang (25.08.2022) | 3,2 | -220,7 | 70,8 |
| Veneti (25.03.2022) | -1,3 | -22,4 | 16,2 |
| Veneti (25.03.2022) | -16,8 | -87,3 | 27,1 |
| Rudan (28.09.2022) | -22,4 | -52,3 | 1,6 |
| Veneti (25.03.2022) | -33,7 | -88,3 | 5,1 |
| **<14 days post exposure** | | | |
| Cocchio (20.08.2022) | 88,0 | 81,0 | 92,0 |
| Cocchio (20.08.2022) | 83,0 | 79,0 | 86,0 |
| Cocchio (20.08.2022) | 81,0 | 76,0 | 85,0 |
| Powell (22.08.2022) | 79,3 | 76,7 | 81,6 |
| Cocchio (20.08.2022) | 78,0 | 69,0 | 84,0 |
| Powell (22.08.2022) | 77,6 | 69,5 | 83,6 |
| Cocchio (20.08.2022) | 72,0 | 69,0 | 74,0 |
| Cocchio (20.08.2022) | 70,0 | 67,0 | 72,0 |
| Powell (22.08.2022) | 69,2 | 55,9 | 78,5 |
| Florentino (08.08.2022) | 61,0 | 56,9 | 64,8 |
| Florentino (08.08.2022) | 58,7 | 56,4 | 61,0 |
| Powell (22.08.2022) | 52,2 | 50,4 | 53,9 |
| Tan (20.07.2022) | 48,8 | 46,9 | 50,8 |
| Rudan (28.09.2022) | 46,9 | 37,0 | 55,3 |
| Florentino (08.08.2022) | 42,5 | 29,4 | 53,3 |
| Florentino (08.08.2022) | 37,5 | 28,5 | 45,3 |
| Rudan (28.09.2022) | 34,0 | 13,2 | 49,9 |
| Florentino (08.08.2022) | 19,7 | 10,6 | 27,9 |
| Powell (22.08.2022) | 15,2 | 9,9 | 20,1 |
| Rudan (28.09.2022) | 14,2 | -10,3 | 33,1 |
| Florentino (08.08.2022) | 10,1 | -14,2 | 29,1 |
| Veneti (25.03.2022) | 9,5 | -55,2 | 47,3 |
| Rudan (28.09.2022) | -18,4 | -89,3 | 26,0 |
| **No information** | | | |

| | | | |
|---|---|---|---|
| Carazo (03.05.2022) | 78,0 | 70,0 | 83,0 |
| Tan (20.07.2022) | 65,3 | 62,0 | 68,3 |
| Fowlkes (11.03.2022) | 59,0 | 24,0 | 78,0 |
| Ionescu (28.06.2022) | 55,2 | 49,5 | 60,3 |
| Piche-Renaud (01.08.2022) | 54,0 | 48,0 | 59,0 |
| Ionescu (28.06.2022) | 41,9 | 37,7 | 45,8 |
| Cocchio (20.08.2022) | 35,0 | 34,0 | 37,0 |
| Ionescu (28.06.2022) | 33,9 | 25,7 | 41,1 |
| Chemaitelly (26.07.2022) | 30,6 | 26,9 | 34,1 |
| Cocchio (20.08.2022) | 30,0 | 26,0 | 33,0 |
| Florentino (08.08.2022) | 28,0 | 26,3 | 29,7 |

| | | | |
|---|---|---|---|
| Chemaitelly (26.07.2022) | 25,7 | 10,0 | 38,6 |
| Florentino (08.08.2022) | 25,1 | 21,3 | 28,7 |
| Chiew (28.09.2022) | 25,0 | 21,0 | 29,0 |
| Tan (20.07.2022) | 24,3 | 19,5 | 28,9 |
| Chiew (28.09.2022) | 22,0 | 13,0 | 31,0 |
| Cocchio (20.08.2022) | 17,0 | 15,0 | 20,0 |
| Piche-Renaud (01.08.2022) | 13,0 | 3,0 | 21,0 |
| Nordstrom (24.10.2022) | -2,0 | -4,0 | 0,0 |

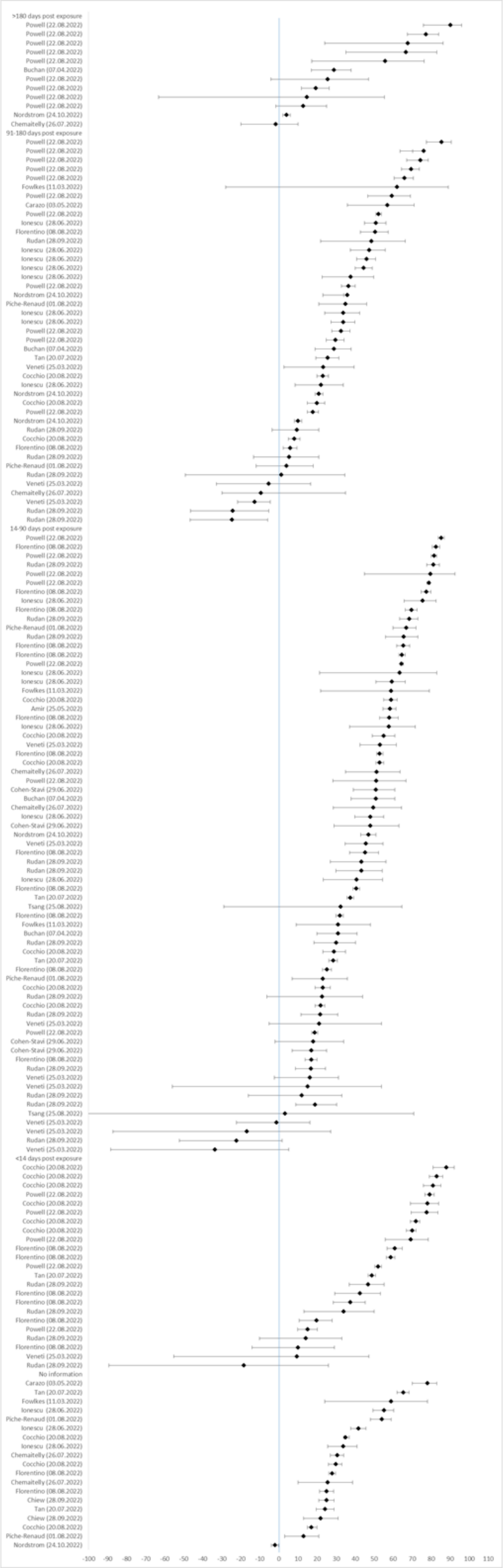
>180 days post exposure
91-180 days post exposure
14-90 days post exposure
<14 days post exposure
No information
-100 -90 -80 -70 -60 -50 -40 -30 -20 -10 0 10 20 30 40 50 60 70 80 90 100 110

**SF16:** Severe disease: General population (adults 18+ years), Exposure 3+, Reference unvaccinated (n=10)

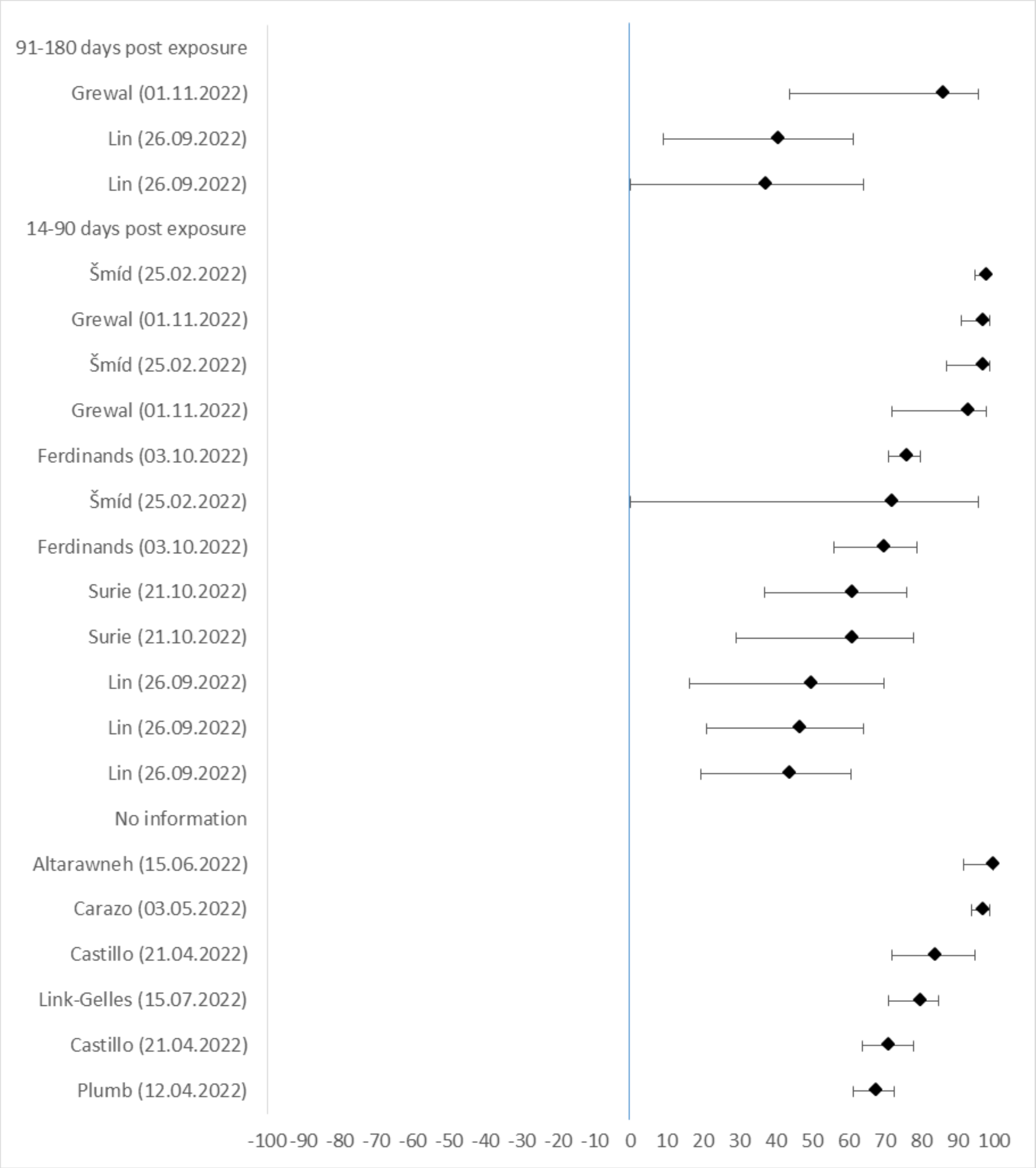

| Author (Date) | VE in % | lower CI 95% | upper CI 95% |
|---|---|---|---|
| **91-180 days post exposure** | | | |
| Grewal (01.11.2022) | 86,0 | 44,0 | 96,0 |
| Lin (26.09.2022) | 40,7 | 9,0 | 61,3 |
| Lin (26.09.2022) | 37,3 | 0,0 | 64,2 |
| **14-90 days post exposure** | | | |
| Šmíd (25.02.2022) | 98,0 | 95,0 | 98,0 |
| Grewal (01.11.2022) | 97,0 | 91,0 | 99,0 |
| Šmíd (25.02.2022) | 97,0 | 87,0 | 99,0 |
| Grewal (01.11.2022) | 93,0 | 72,0 | 98,0 |
| Ferdinands (03.10.2022) | 76,0 | 71,0 | 80,0 |
| Šmíd (25.02.2022) | 72,0 | 0,0 | 96,0 |
| Ferdinands (03.10.2022) | 70,0 | 56,0 | 79,0 |
| Surie (21.10.2022) | 61,0 | 37,0 | 76,0 |
| Surie (21.10.2022) | 61,0 | 29,0 | 78,0 |
| Lin (26.09.2022) | 49,7 | 16,2 | 69,8 |
| Lin (26.09.2022) | 46,8 | 21,1 | 64,2 |
| Lin (26.09.2022) | 43,8 | 19,4 | 60,9 |
| **No information** | | | |
| Altarawneh (15.06.2022) | 100,0 | 91,8 | 100,0 |
| Carazo (03.05.2022) | 97,0 | 94,0 | 99,0 |
| Castillo (21.04.2022) | 84,0 | 72,0 | 95,0 |
| Link-Gelles (15.07.2022) | 80,0 | 71,0 | 85,0 |
| Castillo (21.04.2022) | 71,0 | 64,0 | 78,0 |
| Plumb (12.04.2022) | 67,6 | 61,4 | 72,8 |

**SF17:** Severe disease: General population (adults 18+ years), Exposure 3, Reference unvaccinated (n=24)

| **Author (Date)** | **VE in %** | **lower CI 95%** | **upper CI 95%** |
|---|---|---|---|
| **>180 days post exposure** | | | |
| Grewal (01.11.2022) | 88,0 | 83,0 | 92,0 |
| Grewal (01.11.2022) | 87,0 | 81,0 | 92,0 |
| Tartof (30.06.2022) | 65,0 | 56,0 | 73,0 |
| Ferdinands (03.10.2022) | 41,0 | 35,0 | 47,0 |
| Ferdinands (03.10.2022) | 31,0 | 17,0 | 43,0 |
| **91-180 days post exposure** | | | |
| Castillo (21.04.2022) | 95,0 | 91,0 | 99,0 |
| Grewal (01.11.2022) | 94,0 | 91,0 | 96,0 |
| Link-Gelles (15.07.2022) | 94,0 | 62,0 | 99,0 |
| Castillo (21.04.2022) | 86,0 | 84,0 | 89,0 |
| Hansen (30.03.2022) | 83,6 | 77,7 | 88,0 |
| Cerqueira-Silva (14.04.2022) | 80,2 | 78,0 | 82,2 |
| Hansen (30.03.2022) | 79,0 | 76,5 | 81,3 |
| Hansen (30.03.2022) | 77,3 | 63,1 | 86,1 |
| Tartof (30.06.2022) | 76,0 | 69,0 | 82,0 |
| Stowe (01.04.2022) | 75,9 | 15,8 | 93,1 |
| Cerqueira-Silva (14.04.2022) | 75,8 | 55,0 | 87,0 |
| Tartof (30.06.2022) | 70,0 | 53,0 | 81,0 |
| Lin (26.09.2022) | 69,4 | 63,1 | 74,6 |
| Hansen (30.03.2022) | 66,2 | 61,1 | 70,7 |
| Ferdinands (03.10.2022) | 66,0 | 63,0 | 68,0 |
| Adams (14.06.2022) | 65,0 | 47,0 | 77,0 |
| Lin (26.09.2022) | 61,2 | 55,3 | 66,2 |
| Tartof (22.04.2022) | 53,0 | 36,0 | 66,0 |
| Link-Gelles (15.07.2022) | 52,0 | 44,0 | 59,0 |
| Surie (21.10.2022) | 41,0 | 23,0 | 55,0 |
| Link-Gelles (05.10.2022) | 41,0 | 23,0 | 55,0 |
| Link-Gelles (05.10.2022) | 36,0 | 29,0 | 42,0 |
| Chatzilena (12.09.2022) | 33,9 | 8,4 | 52,4 |
| Link-Gelles (05.10.2022) | 32,0 | 29,0 | 36,0 |
| Surie (21.10.2022) | 29,0 | 3,0 | 48,0 |
| **14-90 days post exposure** | | | |
| Stowe (01.04.2022) | 97,1 | 92,2 | 98,9 |
| Grewal (01.11.2022) | 97,0 | 96,0 | 98,0 |
| Šmíd (25.02.2022) | 97,0 | 80,0 | 100,0 |
| Grewal (01.11.2022) | 95,0 | 93,0 | 96,0 |
| Buchan (22.09.2022) | 95,0 | 87,0 | 98,0 |
| Stowe (01.04.2022) | 94,3 | 88,9 | 97,1 |
| Castillo (21.04.2022) | 94,0 | 92,0 | 97,0 |
| Šmíd (25.02.2022) | 93,0 | 43,0 | 99,0 |
| Castillo (21.04.2022) | 92,0 | 87,0 | 96,0 |
| Link-Gelles (15.07.2022) | 91,0 | 87,0 | 94,0 |
| Chemaitelly (02.06.2022) | 90,9 | 78,6 | 96,1 |
| Hansen (30.03.2022) | 90,2 | 87,3 | 92,5 |
| Chemaitelly (02.06.2022) | 90,1 | 80,6 | 95,0 |
| Šmíd (25.02.2022) | 90,0 | 87,0 | 92,0 |
| Šmíd (25.02.2022) | 90,0 | 84,0 | 94,0 |
| Stowe (01.04.2022) | 89,9 | 78,3 | 95,3 |
| Cerqueira-Silva (14.04.2022) | 89,8 | 88,9 | 90,6 |
| Ferdinands (03.10.2022) | 89,0 | 88,0 | 90,0 |
| Hansen (30.03.2022) | 88,8 | 87,3 | 90,1 |
| Hansen (30.03.2022) | 88,5 | 87,4 | 89,6 |
| Hansen (30.03.2022) | 87,8 | 84,5 | 90,4 |
| Hansen (30.03.2022) | 87,7 | 85,3 | 89,7 |
| Šmíd (25.02.2022) | 87,0 | 84,0 | 88,0 |
| Castillo (21.04.2022) | 87,0 | 80,0 | 93,0 |
| Ferdinands (03.10.2022) | 86,0 | 85,0 | 87,0 |
| Šmíd (25.02.2022) | 85,0 | 80,0 | 88,0 |
| Tartof (22.04.2022) | 85,0 | 80,0 | 89,0 |
| Hansen (30.03.2022) | 84,9 | 83,1 | 86,5 |
| Castillo (21.04.2022) | 84,0 | 82,0 | 87,0 |
| Šmíd (25.02.2022) | 83,0 | 75,0 | 89,0 |
| Lin (26.09.2022) | 82,7 | 77,6 | 86,7 |
| Castillo (21.04.2022) | 82,0 | 79,0 | 85,0 |
| Castillo (21.04.2022) | 82,0 | 78,0 | 86,0 |
| Chemaitelly (02.06.2022) | 81,8 | -49,5 | 97,8 |
| Cerqueira-Silva (14.04.2022) | 81,8 | 69,1 | 89,3 |
| Lin (26.09.2022) | 80,9 | 76,6 | 84,4 |
| Tartof (30.06.2022) | 80,0 | 74,0 | 84,0 |
| Adams (14.06.2022) | 80,0 | 74,0 | 84,0 |
| Surie (21.10.2022) | 79,0 | 74,0 | 84,0 |
| Šmíd (25.02.2022) | 79,0 | 75,0 | 83,0 |
| Tartof (22.04.2022) | 77,0 | 72,0 | 81,0 |
| Tartof (30.06.2022) | 74,0 | 47,0 | 87,0 |
| Tartof (30.06.2022) | 74,0 | 69,0 | 78,0 |
| Lin (26.09.2022) | 73,3 | 68,0 | 77,7 |
| Link-Gelles (15.07.2022) | 69,0 | 58,0 | 76,0 |
| Link-Gelles (05.10.2022) | 68,0 | 50,0 | 80,0 |
| Link-Gelles (05.10.2022) | 62,0 | 54,0 | 66,0 |
| Surie (21.10.2022) | 60,0 | 12,0 | 81,0 |
| Šmíd (25.02.2022) | 60,0 | 37,0 | 74,0 |
| Tartof (30.06.2022) | 59,0 | 40,0 | 72,0 |
| Chatzilena (12.09.2022) | 31,0 | -15,3 | 59,1 |
| **<14 days post exposure** | | | |
| Grewal (01.11.2022) | 96,0 | 91,0 | 98,0 |
| Buchan (22.09.2022) | 91,0 | 71,0 | 97,0 |
| Stowe (01.04.2022) | 90,7 | 56,0 | 98,1 |
| Castillo (21.04.2022) | 88,0 | 79,0 | 98,0 |
| Castillo (21.04.2022) | 88,0 | 78,0 | 98,0 |
| Castillo (21.04.2022) | 72,0 | 64,0 | 79,0 |
| Castillo (21.04.2022) | 65,0 | 57,0 | 73,0 |
| **No information** | | | |
| Tseng (21.04.2022) | 99,2 | 76,3 | 100,0 |
| Tseng (01.10.2022) | 97,5 | 96,3 | 98,3 |
| Altarawneh (15.06.2022) | 94,3 | 81,3 | 98,3 |
| Carazo (03.05.2022) | 94,0 | 91,0 | 96,0 |
| Altarawneh (15.06.2022) | 92,5 | 84,4 | 96,3 |
| Carazo (03.05.2022) | 91,0 | 91,0 | 92,0 |
| Young-Xu (03.08.2022) | 89,0 | 88,0 | 91,0 |
| Tseng (01.10.2022) | 82,0 | 64,5 | 90,8 |
| Adams (14.06.2022) | 77,0 | 71,0 | 82,0 |
| Yan (18.08.2022) | 75,5 | 38,9 | 90,2 |
| Tseng (01.10.2022) | 72,4 | 23,9 | 90,0 |
| Plumb (12.04.2022) | 34,6 | 25,5 | 42,5 |
| Chatzilena (12.09.2022) | 30,9 | 5,9 | 49,3 |

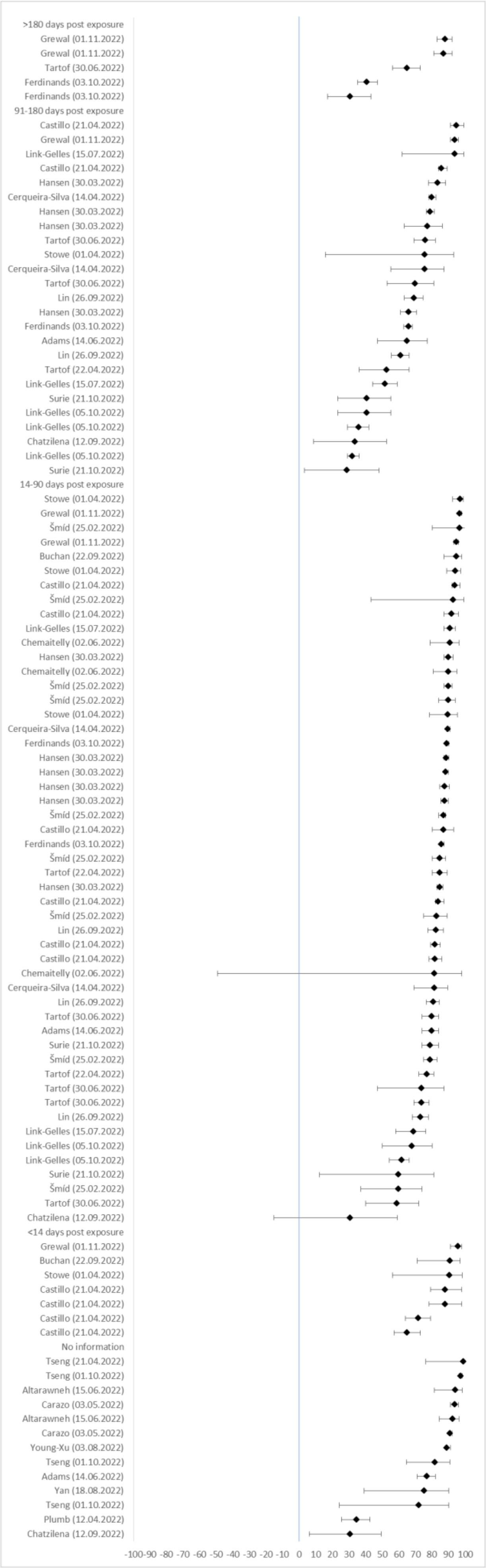

>180 days post exposure
Grewal (01.11.2022)
Grewal (01.11.2022)
Tartof (30.06.2022)
Ferdinands (03.10.2022)
Ferdinands (03.10.2022)
91-180 days post exposure
Castillo (21.04.2022)
Grewal (01.11.2022)
Link-Gelles (15.07.2022)
Castillo (21.04.2022)
Hansen (30.03.2022)
Cerqueira-Silva (14.04.2022)
Hansen (30.03.2022)
Hansen (30.03.2022)
Tartof (30.06.2022)
Stowe (01.04.2022)
Cerqueira-Silva (14.04.2022)
Tartof (30.06.2022)
Lin (26.09.2022)
Hansen (30.03.2022)
Ferdinands (03.10.2022)
Adams (14.06.2022)
Lin (26.09.2022)
Tartof (22.04.2022)
Link-Gelles (15.07.2022)
Surie (21.10.2022)
Link-Gelles (05.10.2022)
Link-Gelles (05.10.2022)
Chatzilena (12.09.2022)
Link-Gelles (05.10.2022)
Surie (21.10.2022)
14-90 days post exposure
Stowe (01.04.2022)
Grewal (01.11.2022)
Šmíd (25.02.2022)
Grewal (01.11.2022)
Buchan (22.09.2022)
Stowe (01.04.2022)
Castillo (21.04.2022)
Šmíd (25.02.2022)
Castillo (21.04.2022)
Link-Gelles (15.07.2022)
Chemaitelly (02.06.2022)
Hansen (30.03.2022)
Chemaitelly (02.06.2022)
Šmíd (25.02.2022)
Šmíd (25.02.2022)
Stowe (01.04.2022)
Cerqueira-Silva (14.04.2022)
Ferdinands (03.10.2022)
Hansen (30.03.2022)
Hansen (30.03.2022)
Hansen (30.03.2022)
Hansen (30.03.2022)
Šmíd (25.02.2022)
Castillo (21.04.2022)
Ferdinands (03.10.2022)
Šmíd (25.02.2022)
Tartof (22.04.2022)
Hansen (30.03.2022)
Castillo (21.04.2022)
Šmíd (25.02.2022)
Lin (26.09.2022)
Castillo (21.04.2022)
Castillo (21.04.2022)
Chemaitelly (02.06.2022)
Cerqueira-Silva (14.04.2022)
Lin (26.09.2022)
Tartof (30.06.2022)
Adams (14.06.2022)
Surie (21.10.2022)
Šmíd (25.02.2022)
Tartof (22.04.2022)
Tartof (30.06.2022)
Tartof (30.06.2022)
Lin (26.09.2022)
Link-Gelles (15.07.2022)
Link-Gelles (05.10.2022)
Link-Gelles (05.10.2022)
Surie (21.10.2022)
Šmíd (25.02.2022)
Tartof (30.06.2022)
Chatzilena (12.09.2022)
<14 days post exposure
Grewal (01.11.2022)
Buchan (22.09.2022)
Stowe (01.04.2022)
Castillo (21.04.2022)
Castillo (21.04.2022)
Castillo (21.04.2022)
Castillo (21.04.2022)
No information
Tseng (21.04.2022)
Tseng (01.10.2022)
Altarawneh (15.06.2022)
Carazo (03.05.2022)
Altarawneh (15.06.2022)
Carazo (03.05.2022)
Young-Xu (03.08.2022)
Tseng (01.10.2022)
Adams (14.06.2022)
Yan (18.08.2022)
Tseng (01.10.2022)
Plumb (12.04.2022)
Chatzilena (12.09.2022)
-100 -90 -80 -70 -60 -50 -40 -30 -20 -10 0 10 20 30 40 50 60 70 80 90 100

**SF18**: Severe disease: General population (adults 18+), Exposure 3, Reference less exposures (2 exposures) (n=9)

| Author (date) | VE in % | lower CI 95% | upper CI 95% |
|---|---|---|---|
| **>180 days post exposure** | | | |
| Ng (26.08.2022) | 89,0 | 78,9 | 94,2 |
| Ng (26.08.2022) | 87,3 | 84,2 | 89,8 |
| Ng (26.08.2022) | 86,3 | 44,7 | 96,6 |
| Ng (26.08.2022) | 84,4 | 73,4 | 90,9 |
| **91-180 days post exposure** | | | |
| Butt (03.05.2022) | 83,0 | 24,0 | 96,0 |
| Chemaitelly (15.11.2022) | 80,5 | 55,7 | 91,4 |
| Chemaitelly (15.11.2022) | 79,9 | 54,2 | 91,2 |
| Butt (03.05.2022) | 79,0 | -78,0 | 98,0 |
| Chemaitelly (15.11.2022) | 52,8 | -61,2 | 91,4 |
| Butt (03.05.2022) | 45,0 | 18,0 | 63,0 |
| **14-90 days post exposure** | | | |
| Ng (26.08.2022) | 97,5 | 89,7 | 99,4 |
| Andersson (29.11.2022) | 96,8 | 92,6 | 100 |
| Andersson (29.11.2022) | 94,4 | 85,5 | 100 |
| Ng (26.08.2022) | 93,8 | 88,2 | 96,8 |
| Butt (03.05.2022) | 90,0 | 22,0 | 99,0 |
| Andersson (29.11.2022) | 88,7 | 84,2 | 93,2 |
| Ng (26.08.2022) | 88,0 | 77,8 | 93,6 |
| Ng (26.08.2022) | 87,3 | 72,8 | 94,1 |
| Ng (26.08.2022) | 87,0 | 70,4 | 94,3 |
| Butt (03.05.2022) | 87,0 | 41,0 | 97,0 |
| Gonzales (27.09.2022) | 86,0 | 23,0 | 97,0 |
| Butt (03.05.2022) | 86,0 | -17,0 | 98,0 |
| Ng (26.08.2022) | 85,2 | 80,2 | 88,9 |
| Andersson (29.11.2022) | 83,7 | 79,0 | 88,4 |
| Ng (26.08.2022) | 81,0 | 76,3 | 84,7 |
| Andersson (29.11.2022) | 80,1 | 70,8 | 89,3 |
| Gonzales (27.09.2022) | 78,0 | 65,0 | 86,0 |
| Andersson (29.11.2022) | 77,9 | 65,2 | 90,6 |
| Andersson (29.11.2022) | 76,5 | 36,5 | 100 |
| Andersson (29.11.2022) | 76,3 | 59,9 | 93,1 |
| Butt (03.05.2022) | 63,0 | 45,0 | 74,0 |
| Butt (03.05.2022) | 62,0 | 43,0 | 75,0 |
| Butt (03.05.2022) | 58,0 | 46,0 | 67,0 |
| Butt (03.05.2022) | 55,0 | 38,0 | 67,0 |
| Butt (03.05.2022) | 50,0 | -68,0 | 85,0 |
| Butt (03.05.2022) | 47,0 | 20,0 | 65,0 |
| Butt (03.05.2022) | 44,0 | 31,0 | 55,0 |
| **No information** | | | |
| Tseng (01.10.2022) | 88,8 | 83,3 | 92,5 |
| Liu (21.06.2022) | 87,8 | 77,3 | 93,4 |
| Tseng (01.10.2022) | 87,5 | 51,8 | 96,8 |
| Butt (03.05.2022) | 83,0 | 65,0 | 92,0 |
| Abu-Raddad (12.05.2022) | 76,5 | 55,9 | 87,5 |
| Tseng (01.10.2022) | 75,0 | 47,6 | 88,1 |
| Liu (21.06.2022) | 65,2 | 60,8 | 69,1 |
| Ioannou (16.06.2022) | 63,2 | 56,4 | 69,0 |
| Ioannou (16.06.2022) | 54,0 | 46,1 | 60,8 |
| Ioannou (16.06.2022) | 53,7 | 45,8 | 60,4 |
| Ioannou (16.06.2022) | 53,3 | 43,2 | 61,6 |
| Ioannou (16.06.2022) | 53,3 | 48,1 | 58,0 |
| Ioannou (16.06.2022) | 53,1 | 45,7 | 59,5 |
| Liu (21.06.2022) | 52,9 | 36,9 | 64,8 |
| Ioannou (16.06.2022) | 52,9 | 45,6 | 59,2 |
| Butt (03.05.2022) | 52,0 | 46,0 | 57,0 |
| Liu (21.06.2022) | 49,4 | 30,8 | 63,0 |
| Ioannou (16.06.2022) | 43,8 | 35,2 | 51,3 |

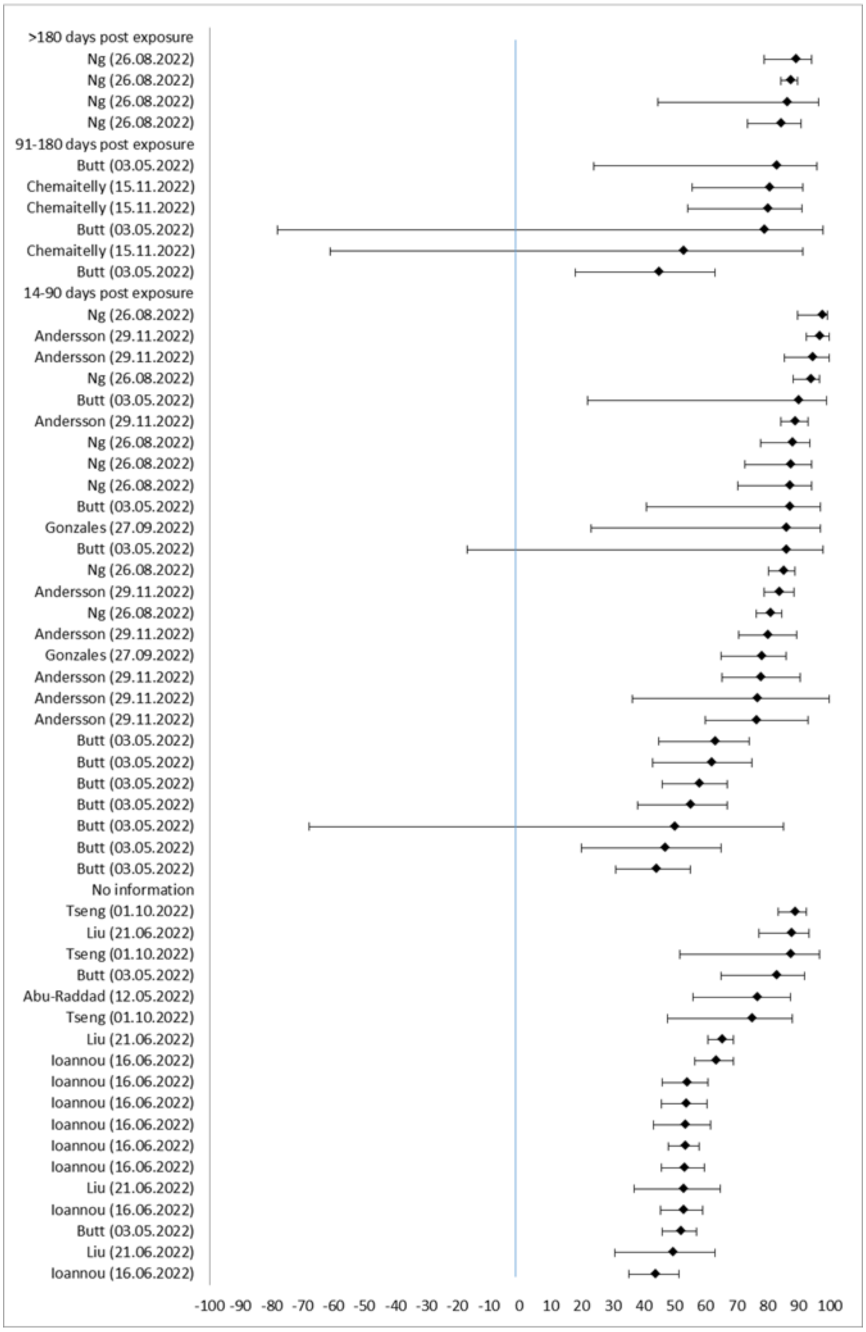
>180 days post exposure
Ng (26.08.2022)
Ng (26.08.2022)
Ng (26.08.2022)
Ng (26.08.2022)
91-180 days post exposure
Butt (03.05.2022)
Chemaitelly (15.11.2022)
Chemaitelly (15.11.2022)
Butt (03.05.2022)
Chemaitelly (15.11.2022)
Butt (03.05.2022)
14-90 days post exposure
Ng (26.08.2022)
Andersson (29.11.2022)
Andersson (29.11.2022)
Ng (26.08.2022)
Butt (03.05.2022)
Andersson (29.11.2022)
Ng (26.08.2022)
Ng (26.08.2022)
Ng (26.08.2022)
Butt (03.05.2022)
Gonzales (27.09.2022)
Butt (03.05.2022)
Ng (26.08.2022)
Andersson (29.11.2022)
Ng (26.08.2022)
Andersson (29.11.2022)
Gonzales (27.09.2022)
Andersson (29.11.2022)
Andersson (29.11.2022)
Andersson (29.11.2022)
Butt (03.05.2022)
Butt (03.05.2022)
Butt (03.05.2022)
Butt (03.05.2022)
Butt (03.05.2022)
Butt (03.05.2022)
Butt (03.05.2022)
No information
Tseng (01.10.2022)
Liu (21.06.2022)
Tseng (01.10.2022)
Butt (03.05.2022)
Abu-Raddad (12.05.2022)
Tseng (01.10.2022)
Liu (21.06.2022)
Ioannou (16.06.2022)
Ioannou (16.06.2022)
Ioannou (16.06.2022)
Ioannou (16.06.2022)
Ioannou (16.06.2022)
Ioannou (16.06.2022)
Liu (21.06.2022)
Ioannou (16.06.2022)
Butt (03.05.2022)
Liu (21.06.2022)
Ioannou (16.06.2022)
-100 -90 -80 -70 -60 -50 -40 -30 -20 -10 0 10 20 30 40 50 60 70 80 90 100

**SF19:** Severe disease: General population (adults 18+ years), Exposure 1-3, Reference unvaccinated (n=23)

| Author (Date) | VE in % | lower CI 95% | upper CI 95% |
|---|---|---|---|
| **>180 days post exposure** | | | |
| Šmíd (25.02.2022) | 92,0 | 86,0 | 96,0 |
| Šmíd (25.02.2022) | 88,0 | 72,0 | 94,0 |
| Grewal (01.11.2022) | 86,0 | 82,0 | 89,0 |
| Stowe (01.04.2022) | 82,3 | 67,7 | 90,3 |
| Grewal (01.11.2022) | 82,0 | 75,0 | 87,0 |
| Buchan (22.09.2022) | 82,0 | 62,0 | 91,0 |
| Grewal (01.11.2022) | 77,0 | 69,0 | 83,0 |
| Castillo (21.04.2022) | 68,0 | 58,0 | 79,0 |
| Šmíd (25.02.2022) | 66,0 | 15,0 | 86,0 |
| Tartof (30.06.2022) | 56,0 | 28,0 | 73,0 |
| Ferdinands (03.10.2022) | 55,0 | 51,0 | 58,0 |
| Ferdinands (03.10.2022) | 51,0 | 47,0 | 54,0 |
| Tartof (22.04.2022) | 51,0 | 43,0 | 59,0 |
| Castillo (21.04.2022) | 50,0 | 44,0 | 57,0 |
| Ferdinands (03.10.2022) | 48,0 | 42,0 | 53,0 |
| Tartof (22.04.2022) | 41,0 | 21,0 | 55,0 |
| Ferdinands (03.10.2022) | 40,0 | 32,0 | 47,0 |
| Surie (21.10.2022) | 37,0 | 12,0 | 55,0 |
| Surie (21.10.2022) | 34,0 | 20,0 | 46,0 |
| Tartof (30.06.2022) | 32,0 | 16,0 | 45,0 |
| Tartof (22.04.2022) | 31,0 | 16,0 | 43,0 |
| Ferdinands (03.10.2022) | 19,0 | 6,0 | 30,0 |
| Tartof (30.06.2022) | 19,0 | 6,0 | 31,0 |
| Tartof (30.06.2022) | 12,0 | -10,0 | 31,0 |
| **91-180 days post exposure** | | | |
| Chemaitelly (07.07.2022) | 88,6 | 70,9 | 95,5 |
| Šmíd (25.02.2022) | 87,0 | 73,0 | 94,0 |
| Gram (01.09.2022) | 85,8 | 73,8 | 92,3 |
| Grewal (01.11.2022) | 85,0 | 79,0 | 89,0 |
| Castillo (21.04.2022) | 85,0 | 76,0 | 93,0 |
| Šmíd (25.02.2022) | 83,0 | 0,0 | 98,0 |
| Šmíd (25.02.2022) | 81,0 | 40,0 | 94,0 |
| Castillo (21.04.2022) | 81,0 | 72,0 | 89,0 |
| Carazo (03.05.2022) | 81,0 | 66,0 | 89,0 |
| Gram (01.09.2022) | 77,6 | 72,6 | 81,6 |
| Chemaitelly (02.06.2022) | 77,5 | 67,8 | 84,3 |
| Buchan (22.09.2022) | 75,0 | 51,0 | 87,0 |
| Castillo (21.04.2022) | 75,0 | 58,0 | 91,0 |
| Altarawneh (15.06.2022) | 71,6 | 15,7 | 90,4 |
| Castillo (21.04.2022) | 70,0 | 65,0 | 76,0 |
| Chemaitelly (02.06.2022) | 68,4 | 46,1 | 81,5 |
| Castillo (21.04.2022) | 57,0 | 51,0 | 64,0 |
| Ferdinands (03.10.2022) | 57,0 | 51,0 | 62,0 |
| Link-Gelles (15.07.2022) | 52,0 | 43,0 | 59,0 |
| Hansen (30.03.2022) | 51,6 | 47,2 | 55,6 |
| Hansen (30.03.2022) | 47,2 | 33,7 | 57,9 |
| Castillo (21.04.2022) | 42,0 | 32,0 | 52,0 |
| Adams (14.06.2022) | 42,0 | 28,0 | 54,0 |
| Link-Gelles (05.10.2022) | 33,0 | 15,0 | 48,0 |
| Link-Gelles (05.10.2022) | 28,0 | 24,0 | 31,0 |
| Link-Gelles (05.10.2022) | 25,0 | 12,0 | 32,0 |
| Link-Gelles (15.07.2022) | 24,0 | 12,0 | 35,0 |
| **14-90 days post exposure** | | | |
| Gram (01.09.2022) | 96,2 | 72,9 | 99,5 |
| Chemaitelly (02.06.2022) | 87,1 | 40,2 | 97,2 |
| Stowe (01.04.2022) | 86,7 | 63,6 | 95,1 |
| Surie (21.10.2022) | 83,0 | 35,0 | 96,0 |
| Grewal (01.11.2022) | 79,0 | 60,0 | 89,0 |
| Gram (01.09.2022) | 79,0 | 57,5 | 89,6 |
| Castillo (21.04.2022) | 78,0 | 57,0 | 100,0 |
| Grewal (01.11.2022) | 77,0 | 65,0 | 84,0 |
| Stowe (01.04.2022) | 75,0 | 42,4 | 89,1 |
| Ferdinands (03.10.2022) | 73,0 | 63,0 | 80,0 |
| Link-Gelles (15.07.2022) | 71,0 | 65,0 | 75,0 |
| Gram (01.09.2022) | 70,5 | 40,3 | 85,5 |
| Chemaitelly (02.06.2022) | 70,4 | 45,0 | 84,0 |
| Ferdinands (03.10.2022) | 68,0 | 59,0 | 75,0 |
| Tartof (22.04.2022) | 68,0 | 48,0 | 80,0 |
| Castillo (21.04.2022) | 68,0 | 41,0 | 94,0 |
| Tartof (22.04.2022) | 64,0 | 51,0 | 73,0 |
| Link-Gelles (15.07.2022) | 64,0 | 52,0 | 73,0 |
| Surie (21.10.2022) | 63,0 | 46,0 | 75,0 |
| Ferdinands (03.10.2022) | 61,0 | 54,0 | 67,0 |
| Castillo (21.04.2022) | 60,0 | 47,0 | 73,0 |
| Šmíd (25.02.2022) | 59,0 | 3,0 | 82,0 |
| Šmíd (25.02.2022) | 57,0 | 32,0 | 72,0 |
| Link-Gelles (15.07.2022) | 57,0 | 19,0 | 77,0 |
| Tartof (30.06.2022) | 56,0 | -2,0 | 81,0 |
| Tartof (30.06.2022) | 54,0 | 38,0 | 65,0 |
| Adams (14.06.2022) | 54,0 | 32,0 | 69,0 |
| Hansen (30.03.2022) | 50,5 | 33,9 | 63,0 |
| Hansen (30.03.2022) | 48,5 | 36,6 | 58,2 |
| Link-Gelles (05.10.2022) | 47,0 | 33,0 | 58,0 |
| Castillo (21.04.2022) | 46,0 | -2,0 | 94,0 |
| Šmíd (25.02.2022) | 45,0 | 29,0 | 54,0 |
| Castillo (21.04.2022) | 44,0 | 29,0 | 59,0 |
| Hansen (30.03.2022) | 42,6 | 26,9 | 54,9 |
| Tartof (30.06.2022) | 42,0 | 31,0 | 52,0 |
| Šmíd (25.02.2022) | 37,0 | 12,0 | 55,0 |
| Šmíd (25.02.2022) | 32,0 | 20,0 | 43,0 |
| Šmíd (25.02.2022) | 29,0 | 21,0 | 37,0 |
| Castillo (21.04.2022) | 28,0 | 5,0 | 50,0 |
| Tartof (30.06.2022) | 27,0 | -11,0 | 52,0 |
| Castillo (21.04.2022) | 1,0 | -23,0 | 25,0 |
| Castillo (21.04.2022) | -9,0 | -69,0 | 51,0 |
| **No information** | | | |
| Carazo (03.05.2022) | 86,0 | 77,0 | 91,0 |
| Tseng (21.04.2022) | 84,5 | 23,0 | 96,9 |
| Yan (18.08.2022) | 80,4 | 40,7 | 93,5 |
| Yan (18.08.2022) | 78,3 | 60,8 | 88,0 |
| Carazo (03.05.2022) | 76,0 | 74,0 | 78,0 |
| Altarawneh (15.06.2022) | 73,5 | 60,5 | 82,2 |
| Risk (16.08.2022) | 68,0 | 53,0 | 78,0 |
| Young-Xu (03.08.2022) | 63,0 | 58,0 | 67,0 |
| Castillo (21.04.2022) | 58,0 | 28,0 | 88,0 |
| Ferdinands (03.10.2022) | 58,0 | 53,0 | 62,0 |
| Castillo (21.04.2022) | 55,0 | 40,0 | 70,0 |
| Carazo (03.05.2022) | 52,0 | 42,0 | 61,0 |
| Castillo (21.04.2022) | 49,0 | 34,0 | 64,0 |
| Adams (14.06.2022) | 47,0 | 30,0 | 60,0 |
| Adams (14.06.2022) | 46,0 | 30,0 | 58,0 |
| Adams (14.06.2022) | 44,0 | 31,0 | 54,0 |
| Adams (14.06.2022) | 41,0 | 9,0 | 62,0 |
| Chemaitelly (02.06.2022) | 40,9 | -199,1 | 88,3 |
| Plumb (12.04.2022) | 33,0 | 15,0 | 47,2 |

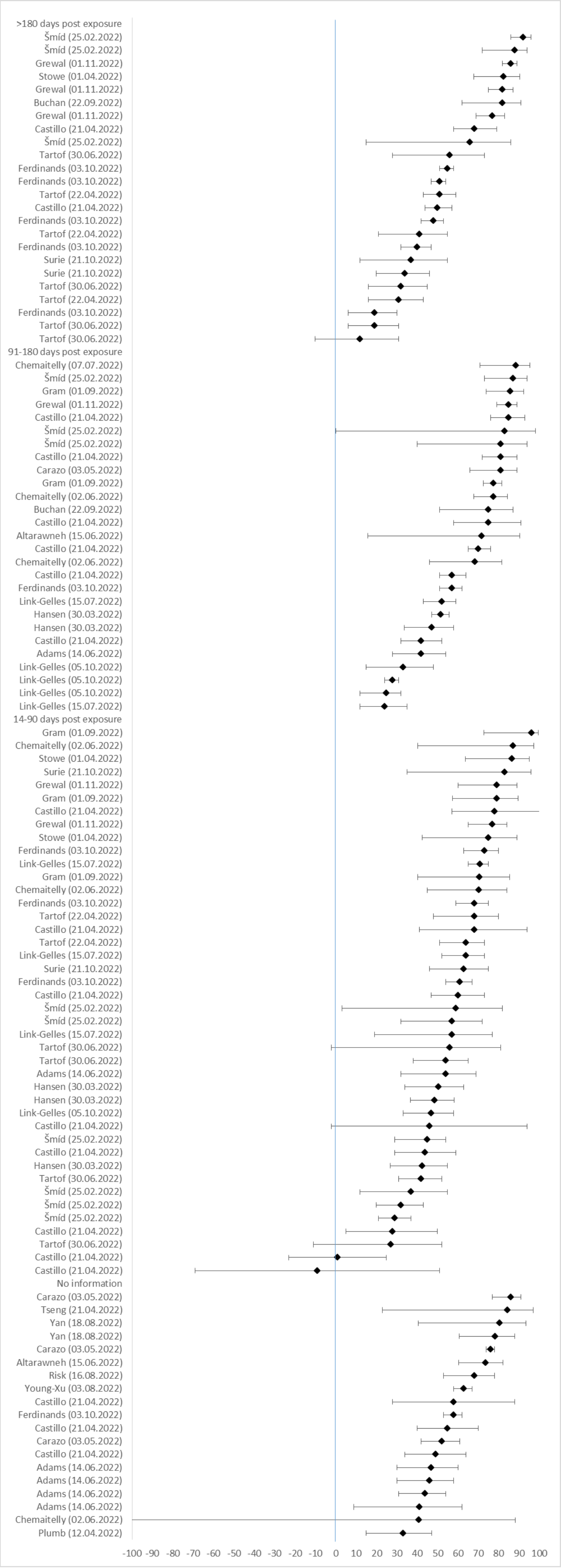
>180 days post exposure
Šmíd (25.02.2022)
Šmíd (25.02.2022)
Grewal (01.11.2022)
Stowe (01.04.2022)
Grewal (01.11.2022)
Buchan (22.09.2022)
Grewal (01.11.2022)
Castillo (21.04.2022)
Šmíd (25.02.2022)
Tartof (30.06.2022)
Ferdinands (03.10.2022)
Ferdinands (03.10.2022)
Tartof (22.04.2022)
Castillo (21.04.2022)
Ferdinands (03.10.2022)
Tartof (22.04.2022)
Ferdinands (03.10.2022)
Surie (21.10.2022)
Surie (21.10.2022)
Tartof (30.06.2022)
Tartof (22.04.2022)
Ferdinands (03.10.2022)
Tartof (30.06.2022)
Tartof (30.06.2022)
91-180 days post exposure
Chemaitelly (07.07.2022)
Šmíd (25.02.2022)
Gram (01.09.2022)
Grewal (01.11.2022)
Castillo (21.04.2022)
Šmíd (25.02.2022)
Šmíd (25.02.2022)
Castillo (21.04.2022)
Carazo (03.05.2022)
Gram (01.09.2022)
Chemaitelly (02.06.2022)
Buchan (22.09.2022)
Castillo (21.04.2022)
Altarawneh (15.06.2022)
Castillo (21.04.2022)
Chemaitelly (02.06.2022)
Castillo (21.04.2022)
Ferdinands (03.10.2022)
Link-Gelles (15.07.2022)
Hansen (30.03.2022)
Hansen (30.03.2022)
Castillo (21.04.2022)
Adams (14.06.2022)
Link-Gelles (05.10.2022)
Link-Gelles (05.10.2022)
Link-Gelles (05.10.2022)
Link-Gelles (15.07.2022)
14-90 days post exposure
Gram (01.09.2022)
Chemaitelly (02.06.2022)
Stowe (01.04.2022)
Surie (21.10.2022)
Grewal (01.11.2022)
Gram (01.09.2022)
Castillo (21.04.2022)
Grewal (01.11.2022)
Stowe (01.04.2022)
Ferdinands (03.10.2022)
Link-Gelles (15.07.2022)
Gram (01.09.2022)
Chemaitelly (02.06.2022)
Ferdinands (03.10.2022)
Tartof (22.04.2022)
Castillo (21.04.2022)
Tartof (22.04.2022)
Link-Gelles (15.07.2022)
Surie (21.10.2022)
Ferdinands (03.10.2022)
Castillo (21.04.2022)
Šmíd (25.02.2022)
Šmíd (25.02.2022)
Link-Gelles (15.07.2022)
Tartof (30.06.2022)
Tartof (30.06.2022)
Adams (14.06.2022)
Hansen (30.03.2022)
Hansen (30.03.2022)
Link-Gelles (05.10.2022)
Castillo (21.04.2022)
Šmíd (25.02.2022)
Castillo (21.04.2022)
Hansen (30.03.2022)
Tartof (30.06.2022)
Šmíd (25.02.2022)
Šmíd (25.02.2022)
Šmíd (25.02.2022)
Castillo (21.04.2022)
Tartof (30.06.2022)
Castillo (21.04.2022)
Castillo (21.04.2022)
No information
Carazo (03.05.2022)
Tseng (21.04.2022)
Yan (18.08.2022)
Yan (18.08.2022)
Carazo (03.05.2022)
Altarawneh (15.06.2022)
Risk (16.08.2022)
Young-Xu (03.08.2022)
Castillo (21.04.2022)
Ferdinands (03.10.2022)
Castillo (21.04.2022)
Carazo (03.05.2022)
Castillo (21.04.2022)
Adams (14.06.2022)
Adams (14.06.2022)
Adams (14.06.2022)
Adams (14.06.2022)
Chemaitelly (02.06.2022)
Plumb (12.04.2022)
-100 -90 -80 -70 -60 -50 -40 -30 -20 -10 0 10 20 30 40 50 60 70 80 90 100

**SF20**: Severe disease: Older adults (60+ years), Exposure 3+, Reference unvaccinated (n=4)

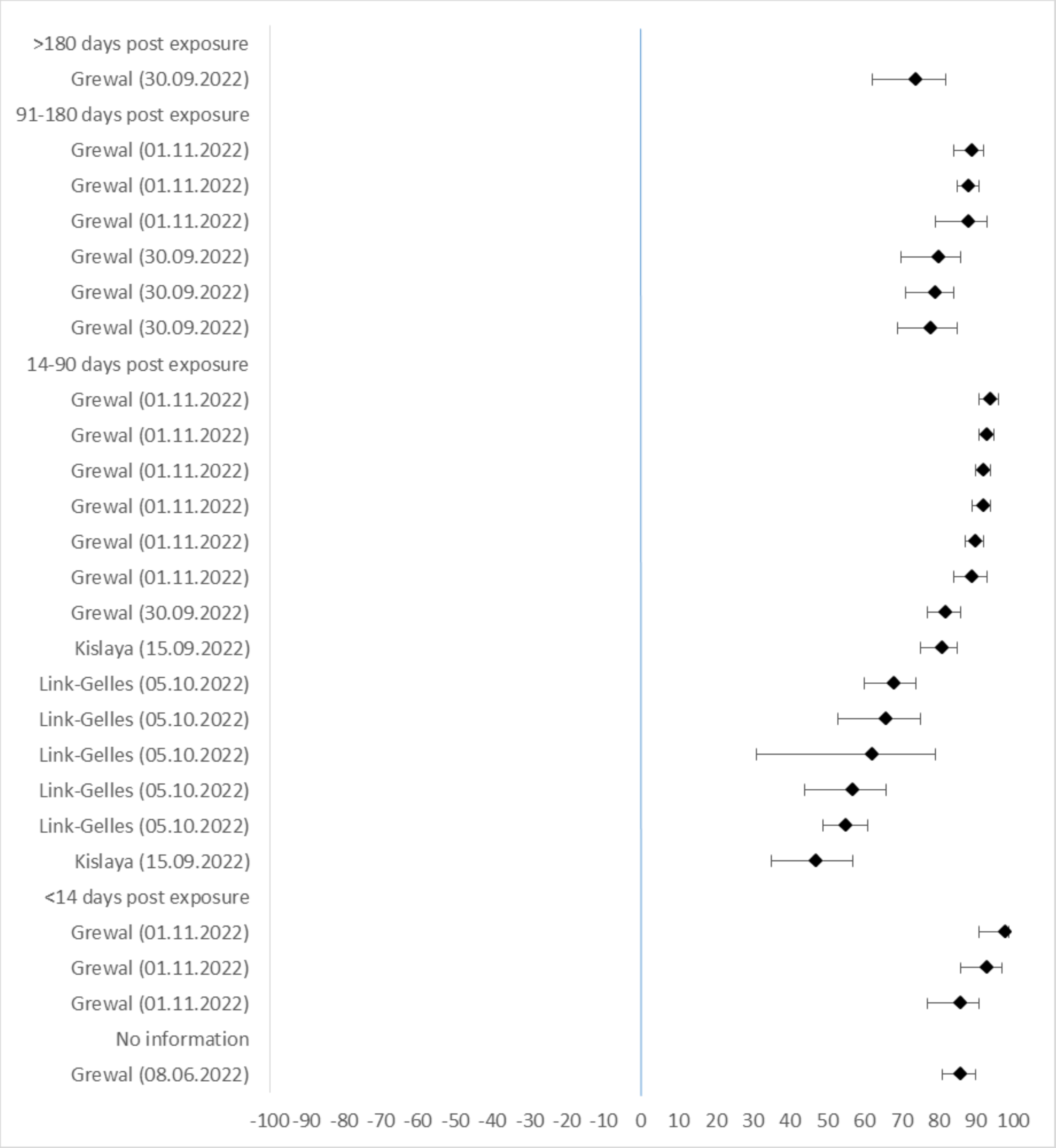

| Author (Date) | VE in % | lower CI 95% | upper CI 95% |
|---|---|---|---|
| **>180 days post exposure** | | | |
| Grewal (30.09.2022) | 74,0 | 62,0 | 82,0 |
| **91-180 days post exposure** | | | |
| Grewal (01.11.2022) | 89,0 | 84,0 | 92,0 |
| Grewal (01.11.2022) | 88,0 | 85,0 | 91,0 |
| Grewal (01.11.2022) | 88,0 | 79,0 | 93,0 |
| Grewal (30.09.2022) | 80,0 | 70,0 | 86,0 |
| Grewal (30.09.2022) | 79,0 | 71,0 | 84,0 |
| Grewal (30.09.2022) | 78,0 | 69,0 | 85,0 |
| **14-90 days post exposure** | | | |
| Grewal (01.11.2022) | 94,0 | 91,0 | 96,0 |
| Grewal (01.11.2022) | 93,0 | 91,0 | 95,0 |
| Grewal (01.11.2022) | 92,0 | 90,0 | 94,0 |
| Grewal (01.11.2022) | 92,0 | 89,0 | 94,0 |
| Grewal (01.11.2022) | 90,0 | 87,0 | 92,0 |
| Grewal (01.11.2022) | 89,0 | 84,0 | 93,0 |
| Grewal (30.09.2022) | 82,0 | 77,0 | 86,0 |
| Kislaya (15.09.2022) | 81,0 | 75,0 | 85,0 |
| Link-Gelles (05.10.2022) | 68,0 | 60,0 | 74,0 |
| Link-Gelles (05.10.2022) | 66,0 | 53,0 | 75,0 |
| Link-Gelles (05.10.2022) | 62,0 | 31,0 | 79,0 |
| Link-Gelles (05.10.2022) | 57,0 | 44,0 | 66,0 |
| Link-Gelles (05.10.2022) | 55,0 | 49,0 | 61,0 |
| Kislaya (15.09.2022) | 47,0 | 35,0 | 57,0 |
| **<14 days post exposure** | | | |
| Grewal (01.11.2022) | 98,0 | 91,0 | 99,0 |
| Grewal (01.11.2022) | 93,0 | 86,0 | 97,0 |
| Grewal (01.11.2022) | 86,0 | 77,0 | 91,0 |
| **No infomation** | | | |
| Grewal (08.06.2022) | 86,0 | 81,0 | 90,0 |

**SF21**: Severe disease: Older adults (60+ years), Exposure 3+, Reference less exposures (3 exposures) (n=10)

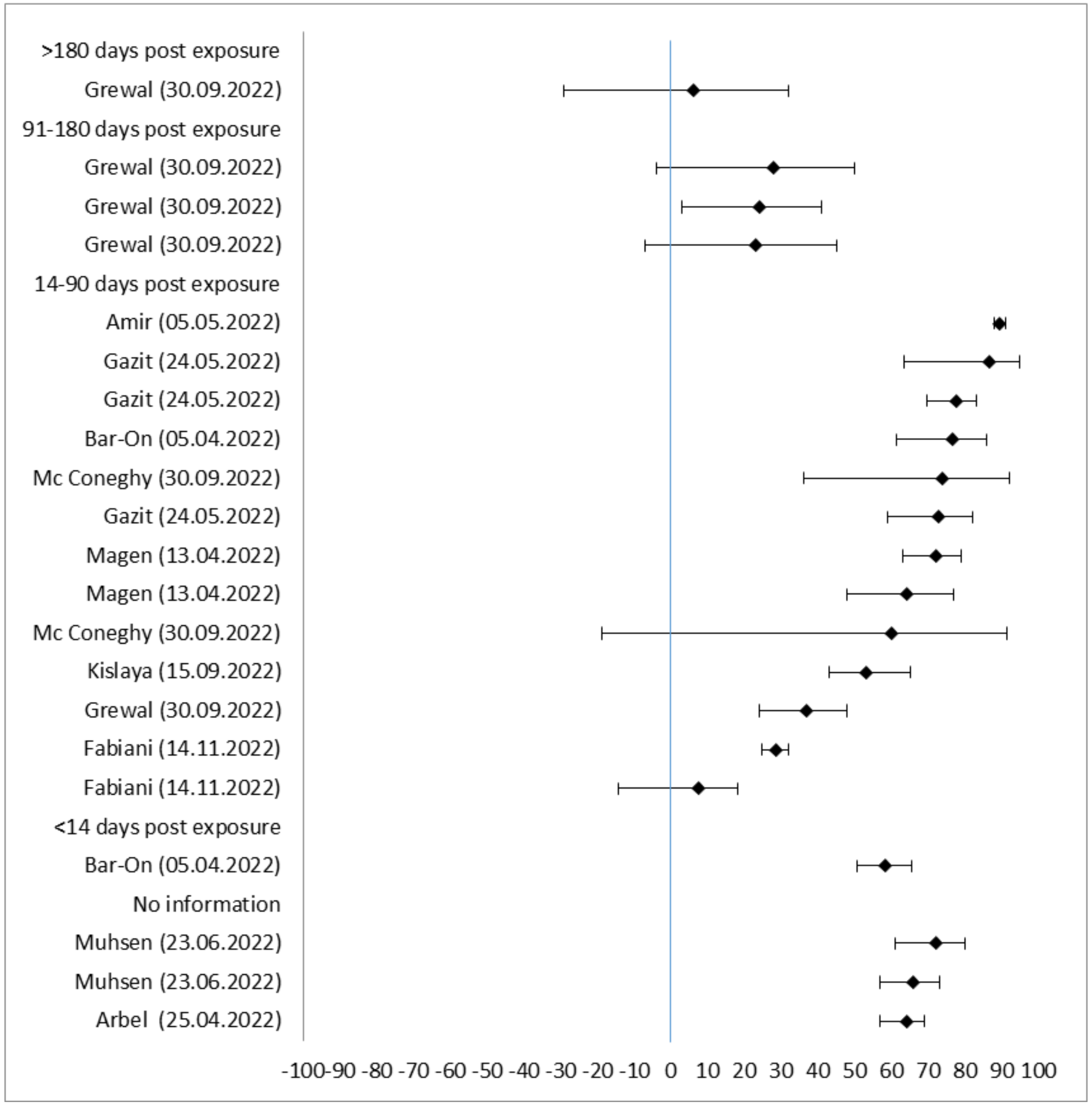

| Author (date) | VE in % | lower CI 95% | upper CI 95% |
|---|---|---|---|
| **>180 days post exposure** | | | |
| Grewal (30.09.2022) | 6,0 | -29,0 | 32,0 |
| **91-180 days post exposure** | | | |
| Grewal (30.09.2022) | 28,0 | -4,0 | 50,0 |
| Grewal (30.09.2022) | 24,0 | 3,0 | 41,0 |
| Grewal (30.09.2022) | 23,0 | -7,0 | 45,0 |
| **14-90 days post exposure** | | | |
| Amir (05.05.2022) | 89,2 | 88,0 | 91,0 |
| Gazit (24.05.2022) | 86,5 | 63,4 | 95,0 |
| Gazit (24.05.2022) | 77,5 | 69,7 | 83,2 |
| Bar-On (05.04.2022) | 76,7 | 61,5 | 85,9 |
| Mc Coneghy (30.09.2022) | 73,9 | 36,1 | 92,2 |
| Gazit (24.05.2022) | 72,8 | 58,8 | 82,1 |
| Magen (13.04.2022) | 72,0 | 63,0 | 79,0 |
| Magen (13.04.2022) | 64,0 | 48,0 | 77,0 |
| Mc Coneghy (30.09.2022) | 60,1 | -18,8 | 91,5 |
| Kislaya (15.09.2022) | 53,0 | 43,0 | 65,0 |
| Grewal (30.09.2022) | 37,0 | 24,0 | 48,0 |
| Fabiani (14.11.2022) | 28,5 | 24,7 | 32,1 |
| Fabiani (14.11.2022) | 7,6 | -14,1 | 18,3 |
| **<14 days post exposure** | | | |
| Bar-On (05.04.2022) | 58,3 | 50,5 | 65,5 |
| **No information** | | | |
| Muhsen (23.06.2022) | 72,0 | 61,0 | 80,0 |
| Muhsen (23.06.2022) | 66,0 | 57,0 | 73,0 |
| Arbel (25.04.2022) | 64,0 | 57,0 | 69,0 |

**SF22:** Severe disease: Older adults (60+ years), Exposure 3, Reference unvaccinated (n=12)

| Author (Date) | VE in % | lower CI 95% | upper CI 95% |
|---|---|---|---|
| **>180 days post exposure** | | | |
| Grewal (01.11.2022) | 85,0 | 78,0 | 90,0 |
| Grewal (01.11.2022) | 82,0 | 77,0 | 87,0 |
| Grewal (01.11.2022) | 79,0 | 71,0 | 85,0 |
| Grewal (01.11.2022) | 79,0 | 73,0 | 84,0 |
| Grewal (01.11.2022) | 76,0 | 68,0 | 82,0 |
| Grewal (01.11.2022) | 75,0 | 68,0 | 80,0 |
| **91-180 days post exposure** | | | |
| Grewal (01.11.2022) | 91,0 | 88,0 | 93,0 |
| Grewal (01.11.2022) | 89,0 | 86,0 | 91,0 |
| Stowe (01.04.2022) | 86,8 | 77,1 | 92,3 |
| Grewal (01.11.2022) | 82,0 | 79,0 | 86,0 |
| Grewal (08.06.2022) | 77,0 | 69,0 | 82,0 |
| Grewal (30.09.2022) | 72,0 | 64,0 | 78,0 |
| Chatzilena (12.09.2022) | 46,9 | 16,3 | 66,4 |
| Link-Gelles (05.10.2022) | 46,0 | 28,0 | 60,0 |
| Link-Gelles (05.10.2022) | 38,0 | 30,0 | 46,0 |
| Link-Gelles (05.10.2022) | 38,0 | 32,0 | 43,0 |
| **14-90 days post exposure** | | | |
| Grewal (01.11.2022) | 98,0 | 97,0 | 98,0 |
| Baum (06.07.2022) | 98,0 | 95,0 | 99,0 |
| Grewal (01.11.2022) | 96,0 | 95,0 | 97,0 |
| Stowe (01.04.2022) | 95,8 | 91,3 | 97,9 |
| Baum (06.07.2022) | 95,0 | 94,0 | 96,0 |
| Grewal (01.11.2022) | 94,0 | 92,0 | 95,0 |
| Stowe (01.04.2022) | 92,8 | 88,4 | 95,6 |
| Stowe (01.04.2022) | 92,5 | 88,1 | 95,2 |
| Grewal (01.11.2022) | 92,0 | 90,0 | 94,0 |
| Wan (17.10.2022) | 92,0 | 89,5 | 93,9 |
| Wan (17.10.2022) | 91,4 | 90,1 | 92,5 |
| Grewal (01.11.2022) | 91,0 | 89,0 | 92,0 |
| Wan (17.10.2022) | 90,4 | 82,9 | 94,6 |
| Wan (17.10.2022) | 86,4 | 62,8 | 95,1 |
| Grewal (01.11.2022) | 86,0 | 84,0 | 89,0 |
| Grewal (08.06.2022) | 80,0 | 72,0 | 86,0 |
| Grewal (30.09.2022) | 77,0 | 68,0 | 83,0 |
| Link-Gelles (05.10.2022) | 73,0 | 55,0 | 84,0 |
| Link-Gelles (05.10.2022) | 56,0 | 39,0 | 68,0 |
| Chatzilena (12.09.2022) | 50,8 | -12,2 | 79,2 |
| **<14 days post exposure** | | | |
| Grewal (01.11.2022) | 95,0 | 91,0 | 97,0 |
| Stowe (01.04.2022) | 94,7 | 71,6 | 99,0 |
| Stowe (01.04.2022) | 89,2 | 63,1 | 96,8 |
| Grewal (01.11.2022) | 88,0 | 81,0 | 92,0 |
| Grewal (01.11.2022) | 66,0 | 48,0 | 77,0 |
| **No information** | | | |
| Kirsbom (01.05.2022) | 90,9 | 88,7 | 92,7 |
| Yan (18.08.2022) | 90,8 | 83,4 | 94,9 |
| Sharma (27.04.2022) | 87,9 | 85,3 | 90,2 |
| Kirsbom (01.05.2022) | 82,3 | 64,2 | 91,3 |
| Sharma (27.04.2022) | 81,8 | 79,2 | 84,2 |
| Kislaya (15.09.2022) | 74,0 | 66,0 | 80,0 |
| Kislaya (15.09.2022) | 63,0 | 55,0 | 70,0 |
| Chatzilena (12.09.2022) | 47,2 | 16,8 | 66,6 |

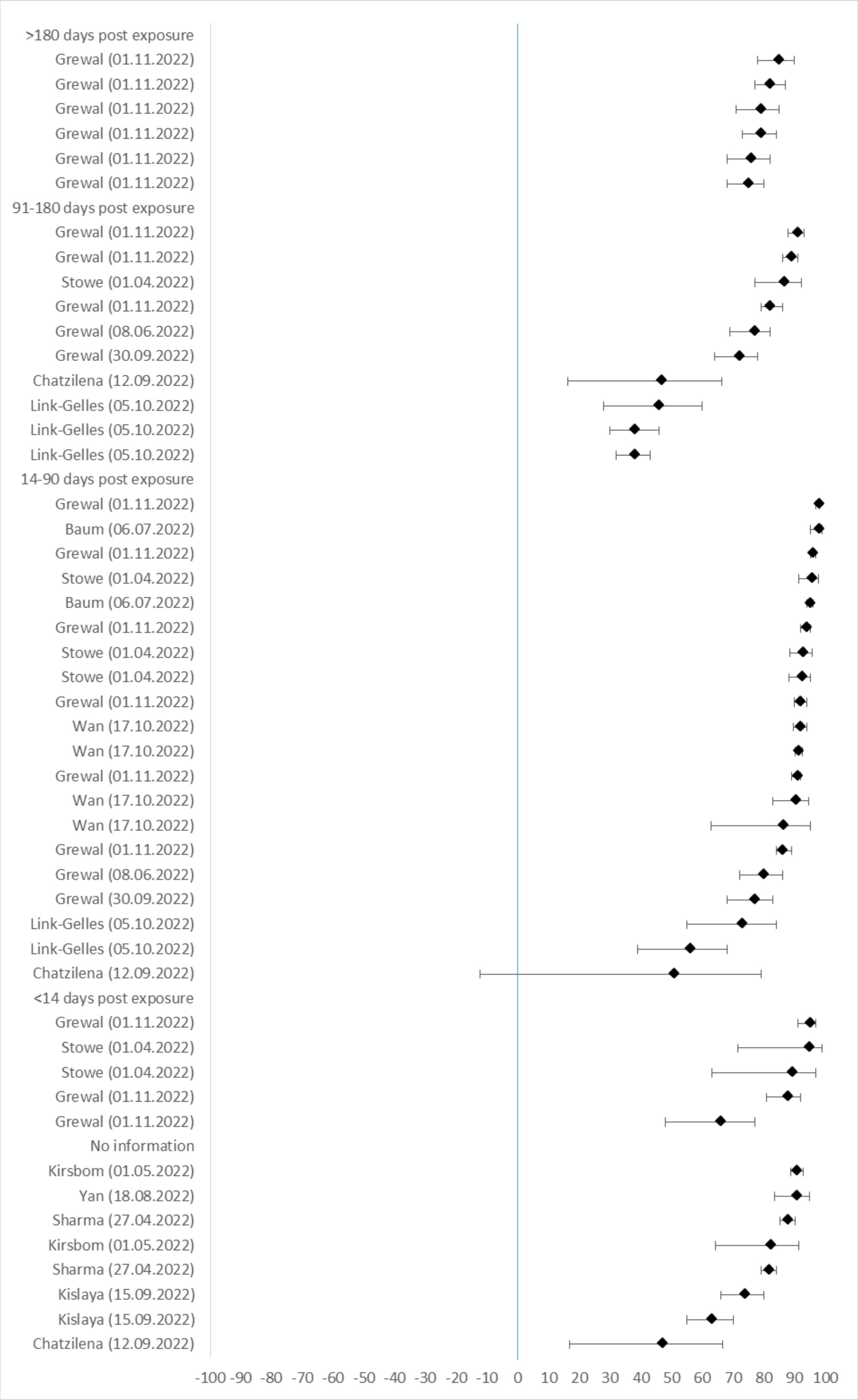
>180 days post exposure
Grewal (01.11.2022)
Grewal (01.11.2022)
Grewal (01.11.2022)
Grewal (01.11.2022)
Grewal (01.11.2022)
Grewal (01.11.2022)
91-180 days post exposure
Grewal (01.11.2022)
Grewal (01.11.2022)
Stowe (01.04.2022)
Grewal (01.11.2022)
Grewal (08.06.2022)
Grewal (30.09.2022)
Chatzilena (12.09.2022)
Link-Gelles (05.10.2022)
Link-Gelles (05.10.2022)
Link-Gelles (05.10.2022)
14-90 days post exposure
Grewal (01.11.2022)
Baum (06.07.2022)
Grewal (01.11.2022)
Stowe (01.04.2022)
Baum (06.07.2022)
Grewal (01.11.2022)
Stowe (01.04.2022)
Stowe (01.04.2022)
Grewal (01.11.2022)
Wan (17.10.2022)
Wan (17.10.2022)
Grewal (01.11.2022)
Wan (17.10.2022)
Wan (17.10.2022)
Grewal (01.11.2022)
Grewal (08.06.2022)
Grewal (30.09.2022)
Link-Gelles (05.10.2022)
Link-Gelles (05.10.2022)
Chatzilena (12.09.2022)
<14 days post exposure
Grewal (01.11.2022)
Stowe (01.04.2022)
Stowe (01.04.2022)
Grewal (01.11.2022)
Grewal (01.11.2022)
No information
Kirsbom (01.05.2022)
Yan (18.08.2022)
Sharma (27.04.2022)
Kirsbom (01.05.2022)
Sharma (27.04.2022)
Kislaya (15.09.2022)
Kislaya (15.09.2022)
Chatzilena (12.09.2022)
-100 -90 -80 -70 -60 -50 -40 -30 -20 -10 0 10 20 30 40 50 60 70 80 90 100

**SF23:** Severe disease: Older adults (60+ years), Exposure 3, Reference less exposures (2 exposures) (n=4)

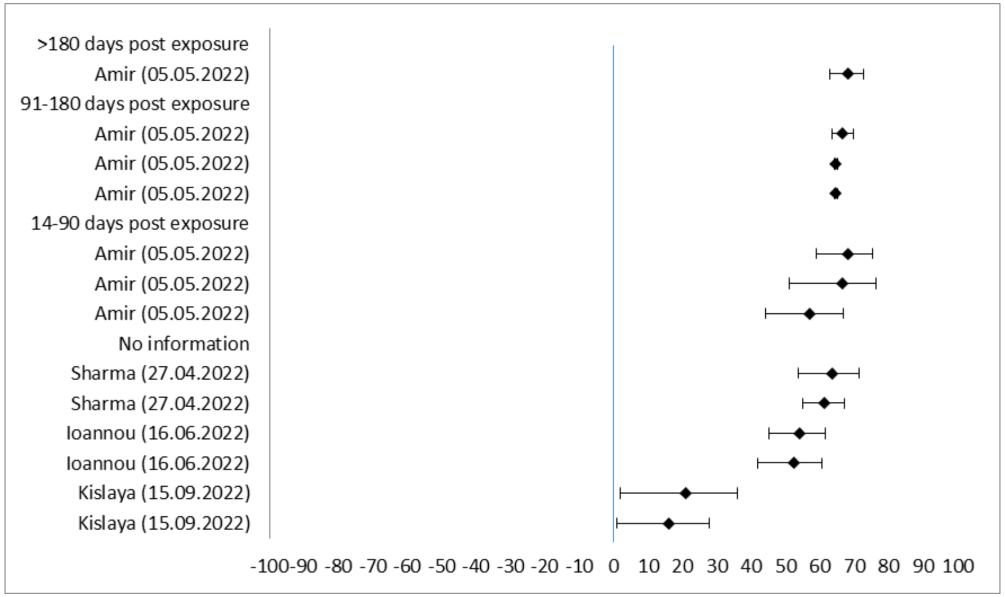

| Author (date) | VE in % | lower CI 95% | upper CI 95% |
|---|---|---|---|
| **>180 days post exposure** | | | |
| Amir (05.05.2022) | 68,1 | 62,8 | 72,6 |
| **91-180 days post exposure** | | | |
| Amir (05.05.2022) | 66,4 | 63,6 | 69,8 |
| Amir (05.05.2022) | 64,7 | 64,2 | 65,1 |
| Amir (05.05.2022) | 64,7 | 64,3 | 65,1 |
| **14-90 days post exposure** | | | |
| Amir (05.05.2022) | 68,1 | 58,9 | 75,5 |
| Amir (05.05.2022) | 66,4 | 51,2 | 76,4 |
| Amir (05.05.2022) | 56,9 | 44,2 | 67,0 |
| **No information** | | | |
| Sharma (27.04.2022) | 63,5 | 53,7 | 71,6 |
| Sharma (27.04.2022) | 61,4 | 55,0 | 67,1 |
| Ioannou (16.06.2022) | 54,2 | 45,4 | 61,6 |
| Ioannou (16.06.2022) | 52,3 | 42,1 | 60,7 |
| Kislaya (15.09.2022) | 21,0 | 2,0 | 36,0 |
| Kislaya (15.09.2022) | 16,0 | 1,0 | 28,0 |

**SF24:** Severe disease: Older adults (60+ years), Exposure 1-3, Reference unvaccinated (n=11)

| Author (Date) | VE in % | lower CI 95% | upper CI 95% |
|---|---|---|---|
| **>180 days post exposure** | | | |
| Grewal (01.11.2022) | 84,0 | 80,0 | 87,0 |
| Grewal (01.11.2022) | 75,0 | 67,0 | 80,0 |
| Stowe (01.04.2022) | 73,4 | 55,1 | 84,3 |
| Grewal (01.11.2022) | 72,0 | 64,0 | 78,0 |
| Grewal (01.11.2022) | 71,0 | 63,0 | 78,0 |
| Grewal (01.11.2022) | 70,0 | 62,0 | 77,0 |
| Grewal (01.11.2022) | 69,0 | 62,0 | 75,0 |
| Grewal (01.11.2022) | 68,0 | 59,0 | 75,0 |
| Grewal (01.11.2022) | 64,0 | 55,0 | 71,0 |
| Kirsbom (01.05.2022) | 61,0 | 49,8 | 69,7 |
| Grewal (01.11.2022) | 46,0 | 34,0 | 56,0 |
| **91-180 days post exposure** | | | |
| Baum (06.07.2022) | 92,0 | 87,0 | 95,0 |
| Baum (06.07.2022) | 85,0 | 82,0 | 87,0 |
| Grewal (01.11.2022) | 79,0 | 73,0 | 84,0 |
| Grewal (01.11.2022) | 76,0 | 67,0 | 82,0 |
| Baum (06.07.2022) | 74,0 | 60,0 | 83,0 |
| Grewal (01.11.2022) | 60,0 | 47,0 | 70,0 |
| Link-Gelles (05.10.2022) | 44,0 | 23,0 | 59,0 |
| Link-Gelles (05.10.2022) | 31,0 | 21,0 | 39,0 |
| Link-Gelles (05.10.2022) | 31,0 | 24,0 | 37,0 |
| **14-90 days post exposure** | | | |
| Baum (06.07.2022) | 98,0 | 95,0 | 99,0 |
| Baum (06.07.2022) | 93,0 | 89,0 | 95,0 |
| Stowe (01.04.2022) | 90,9 | 72,6 | 97,0 |
| Wan (17.10.2022) | 87,1 | 72,3 | 94,0 |
| Grewal (01.11.2022) | 86,0 | 78,0 | 92,0 |
| Grewal (01.11.2022) | 84,0 | 61,0 | 93,0 |
| Grewal (01.11.2022) | 84,0 | 57,0 | 94,0 |
| Wan (17.10.2022) | 82,0 | 79,6 | 84,2 |
| Grewal (01.11.2022) | 81,0 | 69,0 | 88,0 |
| Grewal (01.11.2022) | 80,0 | 62,0 | 90,0 |
| Wan (17.10.2022) | 78,3 | 67,6 | 85,5 |
| Stowe (01.04.2022) | 78,3 | 43,7 | 91,7 |
| Link-Gelles (05.10.2022) | 78,0 | 63,0 | 87,0 |
| Wan (17.10.2022) | 74,9 | 72,6 | 77,0 |
| Wan (17.10.2022) | 63,9 | 58,6 | 68,4 |
| Grewal (01.11.2022) | 57,0 | 27,0 | 75,0 |
| Wan (17.10.2022) | 50,5 | 13,7 | 71,6 |
| Wan (17.10.2022) | 44,8 | 14,8 | 64,3 |
| Wan (17.10.2022) | 44,8 | 38,4 | 50,5 |
| **No information** | | | |
| Yan (18.08.2022) | 82,1 | 74,6 | 87,3 |
| Kislaya (15.09.2022) | 67,0 | 56,0 | 75,0 |
| Sharma (27.04.2022) | 66,7 | 61,4 | 71,6 |
| Kislaya (15.09.2022) | 56,0 | 46,0 | 65,0 |
| Sharma (27.04.2022) | 52,9 | 47,8 | 57,6 |
| Grewal (08.06.2022) | 52,0 | 33,0 | 65,0 |
| Grewal (30.09.2022) | 43,0 | 24,0 | 58,0 |
| Yan (18.08.2022) | 39,1 | 13,0 | 57,4 |

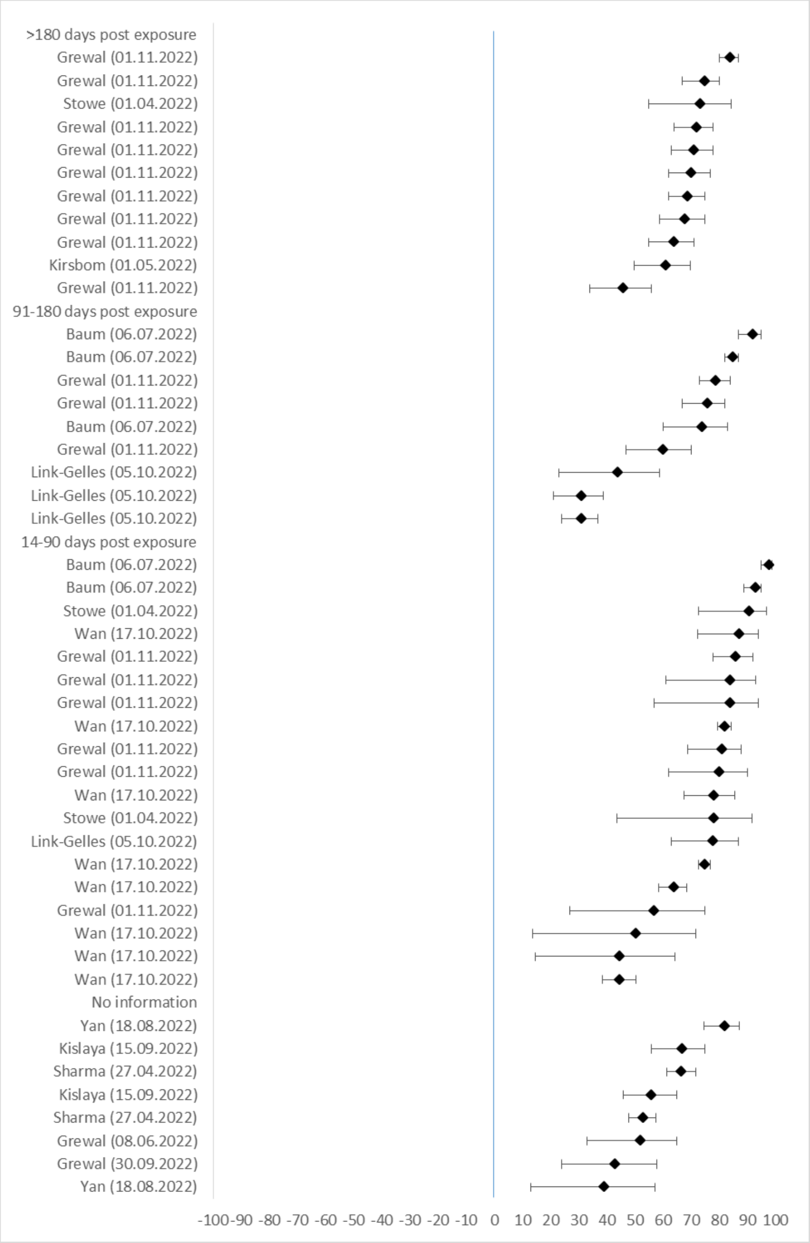
>180 days post exposure
Grewal (01.11.2022)
Grewal (01.11.2022)
Stowe (01.04.2022)
Grewal (01.11.2022)
Grewal (01.11.2022)
Grewal (01.11.2022)
Grewal (01.11.2022)
Grewal (01.11.2022)
Grewal (01.11.2022)
Kirsbom (01.05.2022)
Grewal (01.11.2022)
91-180 days post exposure
Baum (06.07.2022)
Baum (06.07.2022)
Grewal (01.11.2022)
Grewal (01.11.2022)
Baum (06.07.2022)
Grewal (01.11.2022)
Link-Gelles (05.10.2022)
Link-Gelles (05.10.2022)
Link-Gelles (05.10.2022)
14-90 days post exposure
Baum (06.07.2022)
Baum (06.07.2022)
Stowe (01.04.2022)
Wan (17.10.2022)
Grewal (01.11.2022)
Grewal (01.11.2022)
Grewal (01.11.2022)
Wan (17.10.2022)
Grewal (01.11.2022)
Grewal (01.11.2022)
Wan (17.10.2022)
Stowe (01.04.2022)
Link-Gelles (05.10.2022)
Wan (17.10.2022)
Wan (17.10.2022)
Grewal (01.11.2022)
Wan (17.10.2022)
Wan (17.10.2022)
Wan (17.10.2022)
No information
Yan (18.08.2022)
Kislaya (15.09.2022)
Sharma (27.04.2022)
Kislaya (15.09.2022)
Sharma (27.04.2022)
Grewal (08.06.2022)
Grewal (30.09.2022)
Yan (18.08.2022)
-100 -90 -80 -70 -60 -50 -40 -30 -20 -10 0 10 20 30 40 50 60 70 80 90 100

**SF25:** Severe disease: Children and adolescents (5-17 years), Exposure 3, Reference vaccinated (n=4)

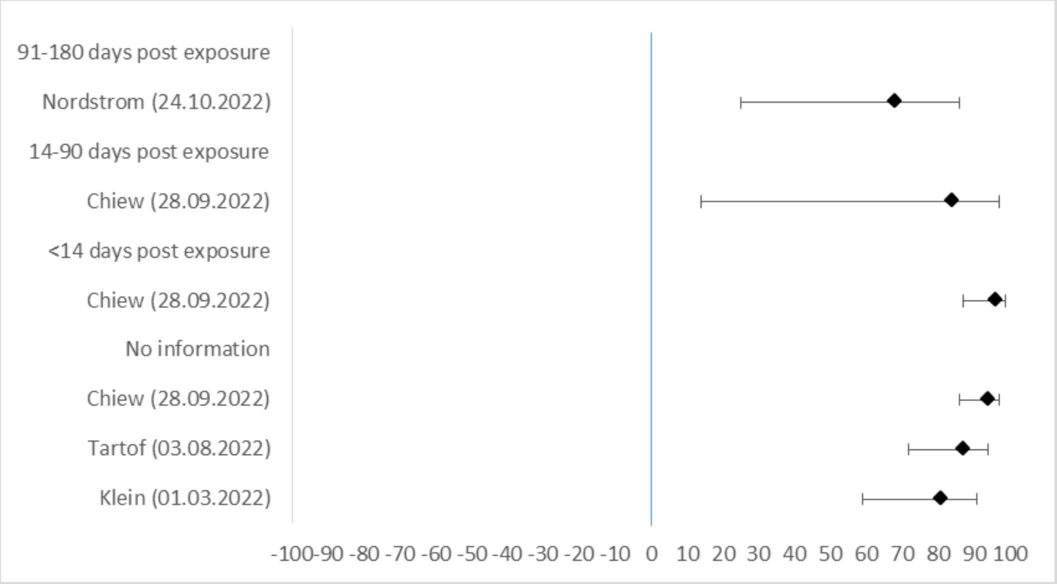

| Author (Date) | VE in % | lower CI 95% | upper CI 95% |
|---|---|---|---|
| **91-180 days post exposure** | | | |
| Nordstrom (24.10.2022) | 68,0 | 25,0 | 86,0 |
| **14-90 days post exposure** | | | |
| Chiew (28.09.2022) | 84,0 | 14,0 | 97,0 |
| **<14 days post exposure** | | | |
| Chiew (28.09.2022) | 96,0 | 87,0 | 99,0 |
| **No information** | | | |
| Chiew (28.09.2022) | 94,0 | 86,0 | 97,0 |
| Tartof (03.08.2022) | 87,0 | 72,0 | 94,0 |
| Klein (01.03.2022) | 81,0 | 59,0 | 91,0 |

**SF26:** Severe disease: Children and adolescents (5-17 years), Exposure 1-3, Reference unvaccinated (n=11)

| Author (Date) | VE in % | lower CI 95% | upper CI 95% |
|---|---|---|---|
| **>180 days post exposure** | | | |
| Buchan (07.04.2022) | 88,0 | 77,0 | 94,0 |
| Price (30.03.2022) | 38,0 | -3,0 | 62,0 |
| Tartof (03.08.2022) | 16,0 | -7,0 | 34,0 |
| **91-180 days post exposure** | | | |
| Zambrano (04.08.2022) | 92,0 | 71,0 | 98,0 |
| Florentino (08.08.2022) | 82,7 | 68,8 | 90,4 |
| Buchan (07.04.2022) | 82,0 | 64,0 | 91,0 |
| Tartof (03.08.2022) | 45,0 | 28,0 | 57,0 |
| Klein (01.03.2022) | -2,0 | -25,0 | 17,0 |
| Klein (01.03.2022) | -3,0 | -30,0 | 18,0 |
| **14-90 days post exposure** | | | |
| Piche-Renaud (01.08.2022) | 94,0 | 56,0 | 99,0 |
| Florentino (08.08.2022) | 86,4 | 75,2 | 92,6 |
| Nordstrom (24.10.2022) | 86,0 | 63,0 | 95,9 |
| Florentino (08.08.2022) | 84,2 | 76,3 | 89,5 |
| Florentino (08.08.2022) | 83,7 | 76,0 | 88,9 |
| Buchan (07.04.2022) | 83,0 | 55,0 | 93,0 |
| Florentino (08.08.2022) | 82,8 | 72,1 | 89,4 |
| Florentino (08.08.2022) | 82,0 | 72,6 | 88,2 |
| Zambrano (04.08.2022) | 78,0 | 48,0 | 90,0 |
| Buchan (07.04.2022) | 76,0 | -10,0 | 95,0 |
| Florentino (08.08.2022) | 75,6 | 58,1 | 85,8 |
| Piche-Renaud (01.08.2022) | 74,0 | 44,0 | 88,0 |
| Tartof (03.08.2022) | 73,0 | 54,0 | 84,0 |
| Klein (01.03.2022) | 51,0 | 30,0 | 65,0 |
| Klein (01.03.2022) | 45,0 | 30,0 | 57,0 |
| Price (30.03.2022) | 43,0 | -1,0 | 68,0 |
| Tartof (03.08.2022) | 38,0 | 14,0 | 56,0 |
| Klein (01.03.2022) | 34,0 | 8,0 | 53,0 |
| **<14 days post exposure** | | | |
| Florentino (08.08.2022) | 65,0 | 37,2 | 80,5 |
| Florentino (08.08.2022) | 62,4 | -22,2 | 88,5 |
| Florentino (08.08.2022) | 20,6 | -152,2 | 75,0 |
| **No information** | | | |
| Buchan (07.04.2022) | 85,0 | 74,0 | 91,0 |
| Tan (11.08.2022) | 82,7 | 74,8 | 88,2 |
| Piche-Renaud (01.08.2022) | 81,0 | 64,0 | 90,0 |
| Gonzalez (16.07.2022) | 78,2 | 42,0 | 90,3 |
| Nordstrom (24.10.2022) | 75,0 | 54,0 | 86,0 |
| Chiew (28.09.2022) | 75,0 | 56,0 | 86,0 |
| Price (30.03.2022) | 68,0 | 42,0 | 82,0 |
| Florentino (08.08.2022) | 56,3 | 45,9 | 64,6 |
| Tan (11.08.2022) | 42,3 | 24,9 | 55,7 |
| Chiew (28.09.2022) | 33,0 | 0,0 | 76,0 |

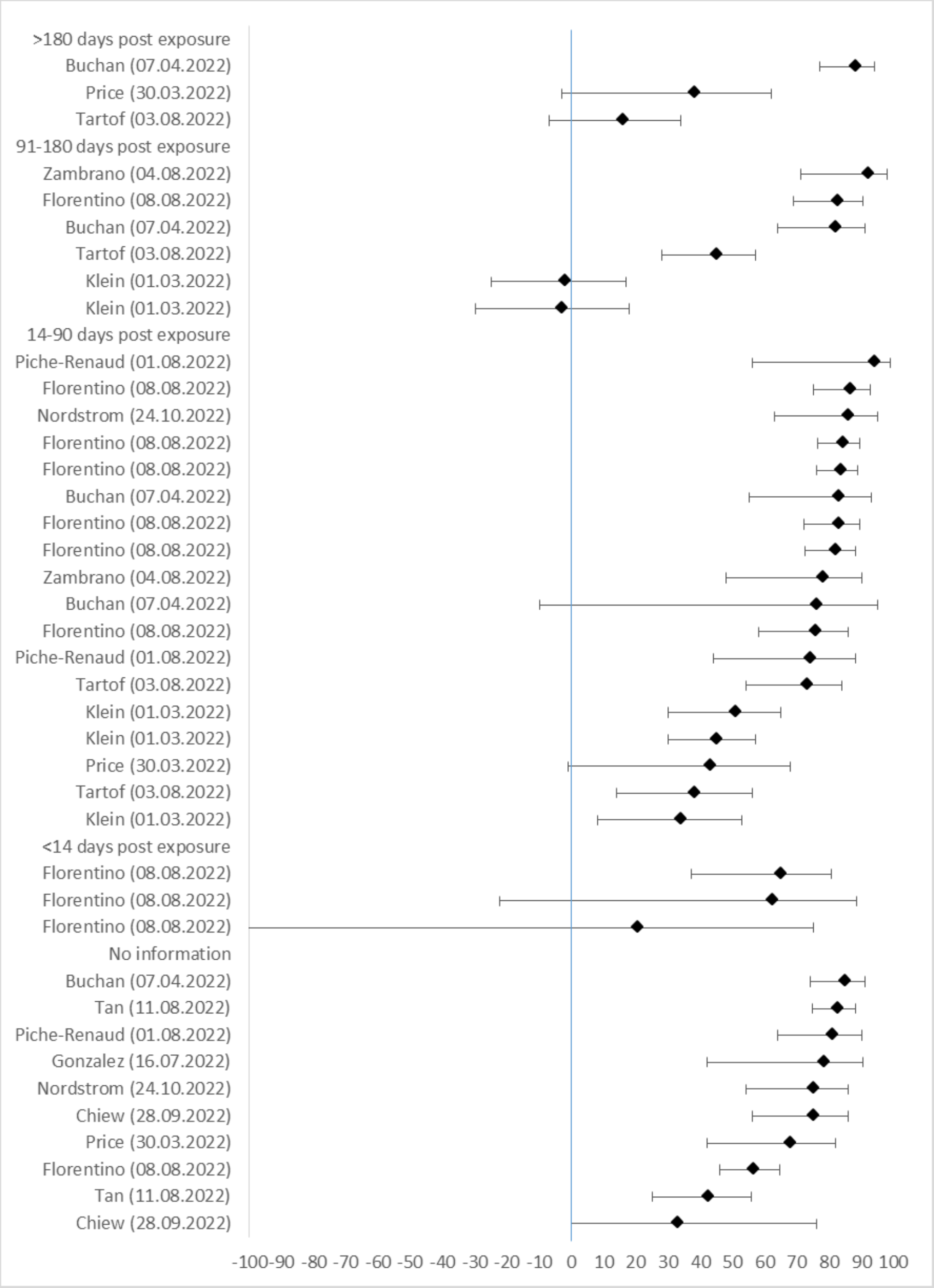
>180 days post exposure
Buchan (07.04.2022)
Price (30.03.2022)
Tartof (03.08.2022)
91-180 days post exposure
Zambrano (04.08.2022)
Florentino (08.08.2022)
Buchan (07.04.2022)
Tartof (03.08.2022)
Klein (01.03.2022)
Klein (01.03.2022)
14-90 days post exposure
Piche-Renaud (01.08.2022)
Florentino (08.08.2022)
Nordstrom (24.10.2022)
Florentino (08.08.2022)
Florentino (08.08.2022)
Buchan (07.04.2022)
Florentino (08.08.2022)
Florentino (08.08.2022)
Zambrano (04.08.2022)
Buchan (07.04.2022)
Florentino (08.08.2022)
Piche-Renaud (01.08.2022)
Tartof (03.08.2022)
Klein (01.03.2022)
Klein (01.03.2022)
Price (30.03.2022)
Tartof (03.08.2022)
Klein (01.03.2022)
<14 days post exposure
Florentino (08.08.2022)
Florentino (08.08.2022)
Florentino (08.08.2022)
No information
Buchan (07.04.2022)
Tan (11.08.2022)
Piche-Renaud (01.08.2022)
Gonzalez (16.07.2022)
Nordstrom (24.10.2022)
Chiew (28.09.2022)
Price (30.03.2022)
Florentino (08.08.2022)
Tan (11.08.2022)
Chiew (28.09.2022)
-100 -90 -80 -70 -60 -50 -40 -30 -20 -10 0 10 20 30 40 50 60 70 80 90 100

**SF27:** Death: General population (adults 18+ years), Exposure 3+, Reference unvaccinated (n=1)

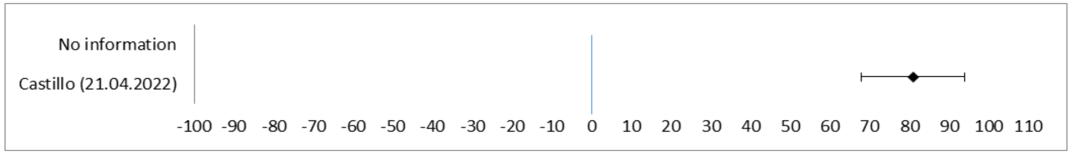

| Author (Date) | VE in % | lower CI 95% | upper CI 95% |
|---|---|---|---|
| **No information** | | | |
| Castillo (21.04.2022) | 81,0 | 68,0 | 94,0 |

**SF28:** Death: General population (adults 18+ years), Exposure 3, Reference unvaccinated (n=6)

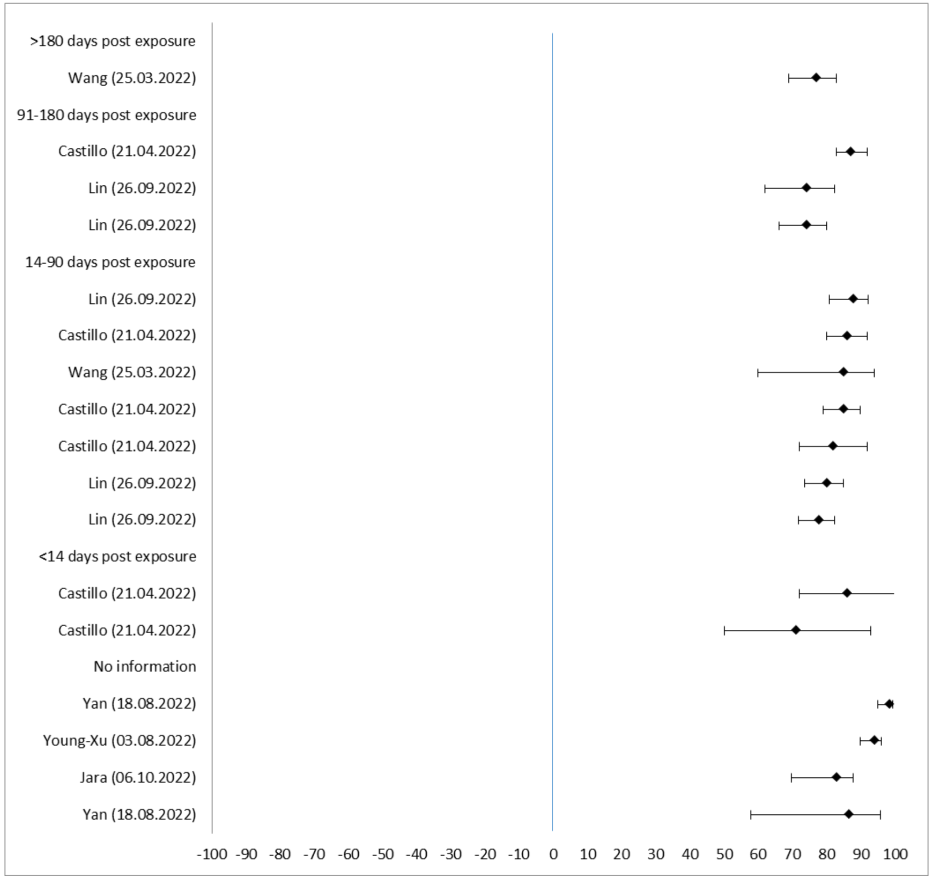

| Author (Date) | VE in % | lower CI 95% | upper CI 95% |
|---|---|---|---|
| **>180 days post exposure** | | | |
| Wang (25.03.2022) | 77,0 | 69,0 | 83,0 |
| **91-180 days post exposure** | | | |
| Castillo (21.04.2022) | 87,0 | 83,0 | 92,0 |
| Lin (26.09.2022) | 74,1 | 62,0 | 82,3 |
| Lin (26.09.2022) | 74,1 | 66,2 | 80,1 |
| **14-90 days post exposure** | | | |
| Lin (26.09.2022) | 87,7 | 80,9 | 92,1 |
| Castillo (21.04.2022) | 86,0 | 80,0 | 92,0 |
| Wang (25.03.2022) | 85,0 | 60,0 | 94,0 |
| Castillo (21.04.2022) | 85,0 | 79,0 | 90,0 |
| Castillo (21.04.2022) | 82,0 | 72,0 | 92,0 |
| Lin (26.09.2022) | 80,0 | 73,6 | 84,9 |
| Lin (26.09.2022) | 77,8 | 71,8 | 82,5 |
| **<14 days post exposure** | | | |
| Castillo (21.04.2022) | 86,0 | 72,0 | 100,0 |
| Castillo (21.04.2022) | 71,0 | 50,0 | 93,0 |
| **No information** | | | |
| Yan (18.08.2022) | 98,4 | 95,0 | 99,5 |
| Young-Xu (03.08.2022) | 94,0 | 90,0 | 96,0 |
| Jara (06.10.2022) | 83,0 | 69,7 | 87,7 |
| Yan (18.08.2022) | 86,6 | 57,9 | 95,7 |

**SF29**: Death: General population (adults 18+ years), Exposure 3, Reference less exposures (2 exposures) (n=3)

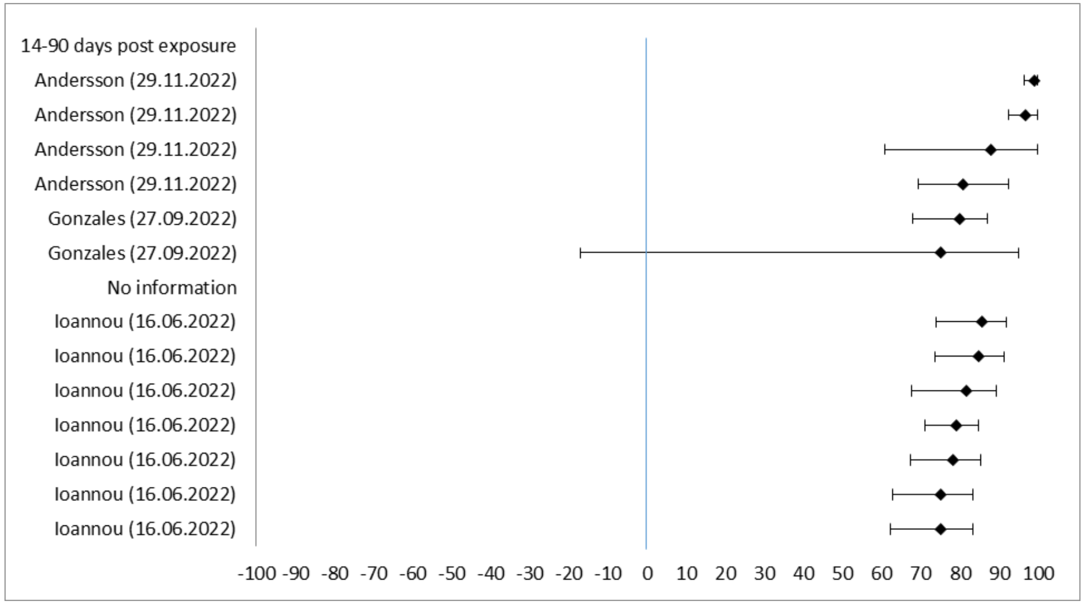

| Author (date) | VE in % | Lower CI 95% | Upper CI 95% |
|---|---|---|---|
| **14-90 days post exposure** | | | |
| Andersson (29.11.2022) | 99,0 | 96,6 | 100 |
| Andersson (29.11.2022) | 96,8 | 92,6 | 100 |
| Andersson (29.11.2022) | 87,8 | 60,7 | 100 |
| Andersson (29.11.2022) | 80,9 | 69,5 | 92,4 |
| Gonzales (27.09.2022) | 80,0 | 68,0 | 87,0 |
| Gonzales (27.09.2022) | 75,0 | -17,0 | 95,0 |
| **No information** | | | |
| Ioannou (16.06.2022) | 85,5 | 73,9 | 92,0 |
| Ioannou (16.06.2022) | 84,8 | 73,7 | 91,2 |
| Ioannou (16.06.2022) | 81,6 | 67,8 | 89,4 |
| Ioannou (16.06.2022) | 79,1 | 71,2 | 84,9 |
| Ioannou (16.06.2022) | 78,1 | 67,5 | 85,3 |
| Ioannou (16.06.2022) | 75,2 | 62,9 | 83,4 |
| Ioannou (16.06.2022) | 75,0 | 62,3 | 83,4 |

**SF30:** Death: General population (adults 18+ years), Exposure 1-3, Reference unvaccinated (n=4)

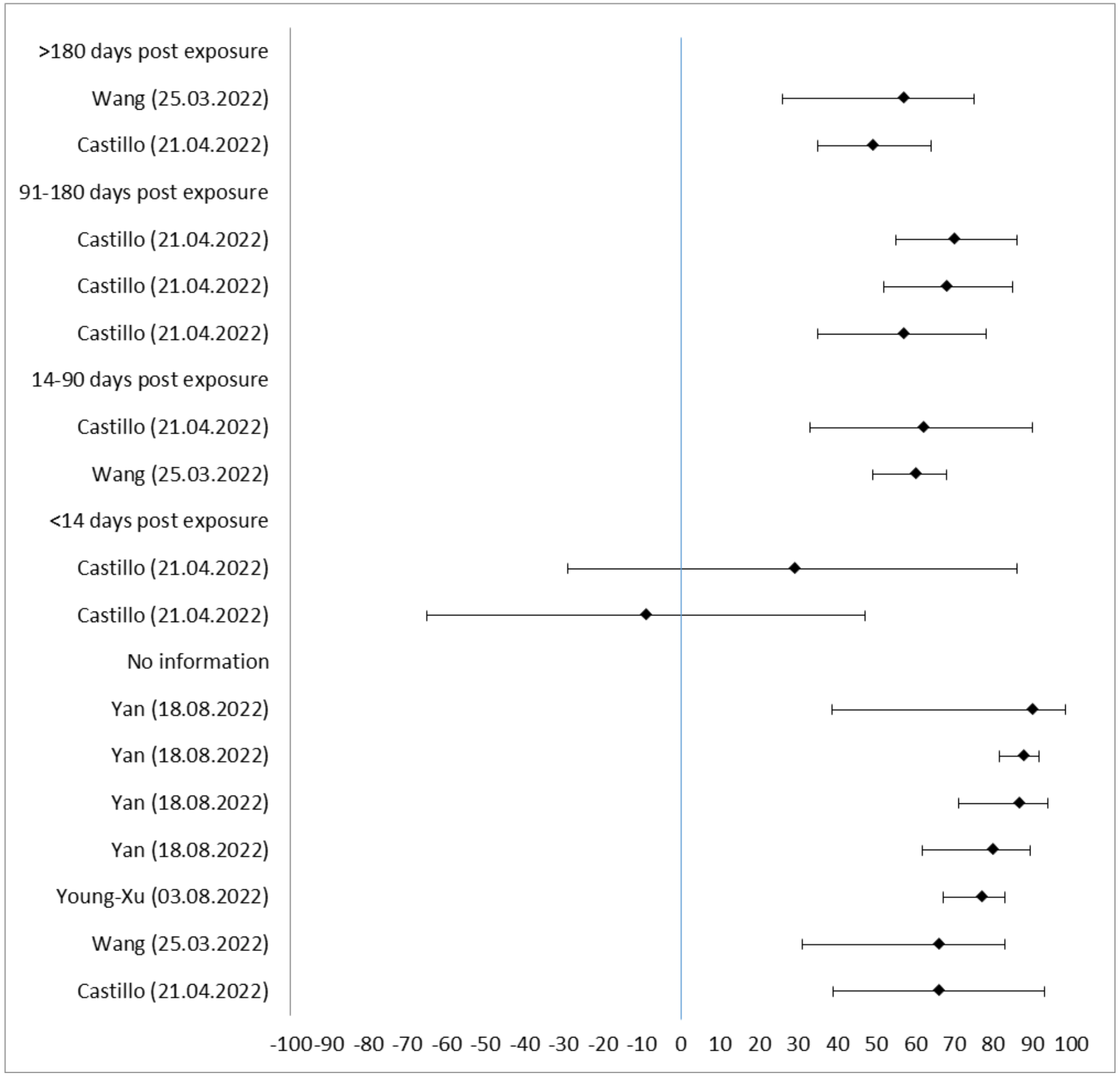

| **Author (Date)** | **VE in %** | **lower CI 95%** | **upper CI 95%** |
|---|---|---|---|
| **>180 days post exposure** | | | |
| Wang (25.03.2022) | 57,0 | 26,0 | 75,0 |
| Castillo (21.04.2022) | 49,0 | 35,0 | 64,0 |
| **91-180 days post exposure** | | | |
| Castillo (21.04.2022) | 70,0 | 55,0 | 86,0 |
| Castillo (21.04.2022) | 68,0 | 52,0 | 85,0 |
| Castillo (21.04.2022) | 57,0 | 35,0 | 78,0 |
| **14-90 days post exposure** | | | |
| Castillo (21.04.2022) | 62,0 | 33,0 | 90,0 |
| Wang (25.03.2022) | 60,0 | 49,0 | 68,0 |
| **<14 days post exposure** | | | |
| Castillo (21.04.2022) | 29,0 | -29,0 | 86,0 |
| Castillo (21.04.2022) | -9,0 | -65,0 | 47,0 |
| **No information** | | | |
| Yan (18.08.2022) | 89,9 | 38,6 | 98,3 |
| Yan (18.08.2022) | 87,6 | 81,4 | 91,8 |
| Yan (18.08.2022) | 86,6 | 71,0 | 93,8 |
| Yan (18.08.2022) | 79,9 | 61,7 | 89,5 |
| Young-Xu (03.08.2022) | 77,0 | 67,0 | 83,0 |
| Wang (25.03.2022) | 66,0 | 31,0 | 83,0 |
| Castillo (21.04.2022) | 66,0 | 39,0 | 93,0 |

**SF31:** Death: Older adults (60+ years), Exposure 3+, Reference unvaccinated (n=1)

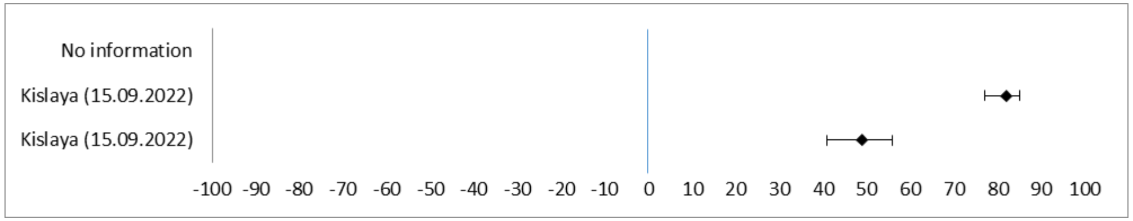

| Author (Date) | VE in % | lower CI 95% | upper CI 95% |
|---|---|---|---|
| **No information** | | | |
| Kislaya (15.09.2022) | 82,0 | 77,0 | 85,0 |
| Kislaya (15.09.2022) | 49,0 | 41,0 | 56,0 |

**SF32:** Death: Older adults (60+ years), Exposure 3+, Reference less exposures (3 exposures) (n=5)

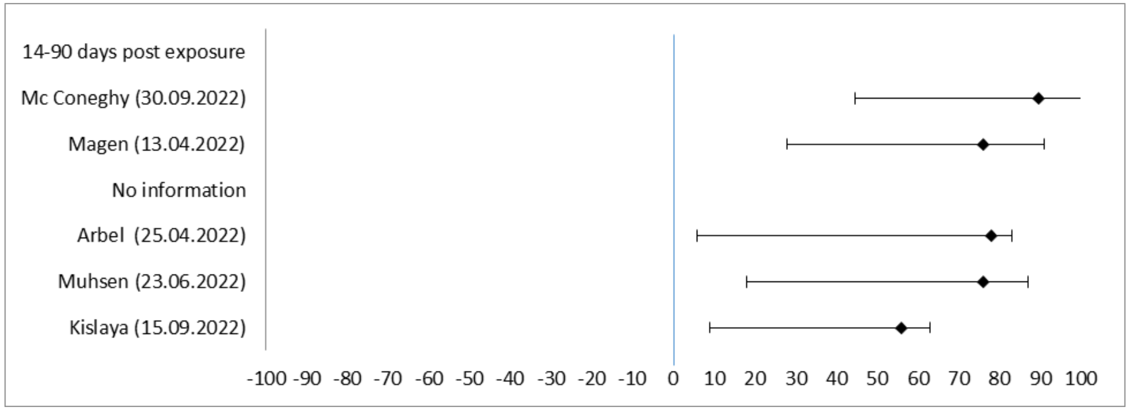

| Author (date) | VE in % | lower CI 95% | upper CI 95% |
|---|---|---|---|
| **14-90 days post exposure** | | | |
| Mc Coneghy (30.09.2022) | 89,6 | 45,0 | 100,0 |
| Magen (13.04.2022) | 76,0 | 48,0 | 91,0 |
| **No information** | | | |
| Arbel (25.04.2022) | 78,0 | 72,0 | 83,0 |
| Muhsen (23.06.2022) | 76,0 | 58,0 | 87,0 |
| Kislaya (15.09.2022) | 56,0 | 47,0 | 63,0 |

**SF33:** Death: Older adults (60+ years), Exposure 3, Reference unvaccinated (n=4)

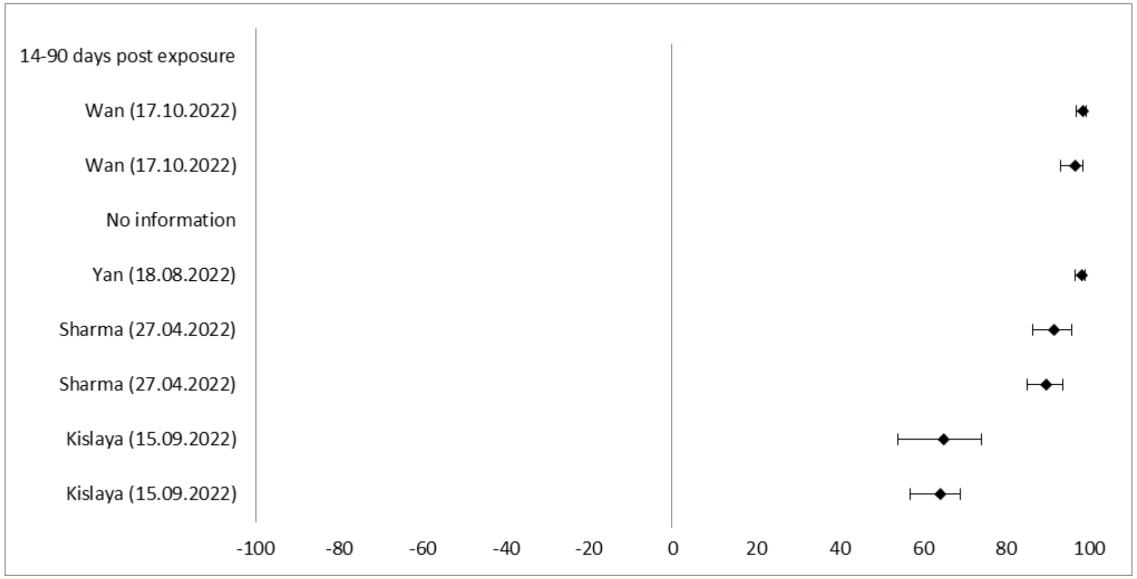

| Author (Date) | VE in % | lower CI 95% | upper CI 95% |
|---|---|---|---|
| **14-90 days post exposure** | | | |
| Wan (17.10.2022) | 98,4 | 96,6 | 99,2 |
| Wan (17.10.2022) | 96,4 | 92,9 | 98,2 |
| **No information** | | | |
| Yan (18.08.2022) | 98,0 | 96,5 | 98,9 |
| Sharma (27.04.2022) | 91,4 | 86,4 | 95,6 |
| Sharma (27.04.2022) | 89,6 | 85,0 | 93,6 |
| Kislaya (15.09.2022) | 65,0 | 54,0 | 74,0 |
| Kislaya (15.09.2022) | 64,0 | 57,0 | 69,0 |

**SF34:** Death: Older adults (60+ years), Exposure 3, Reference less exposures (2 exposures) (n=3)

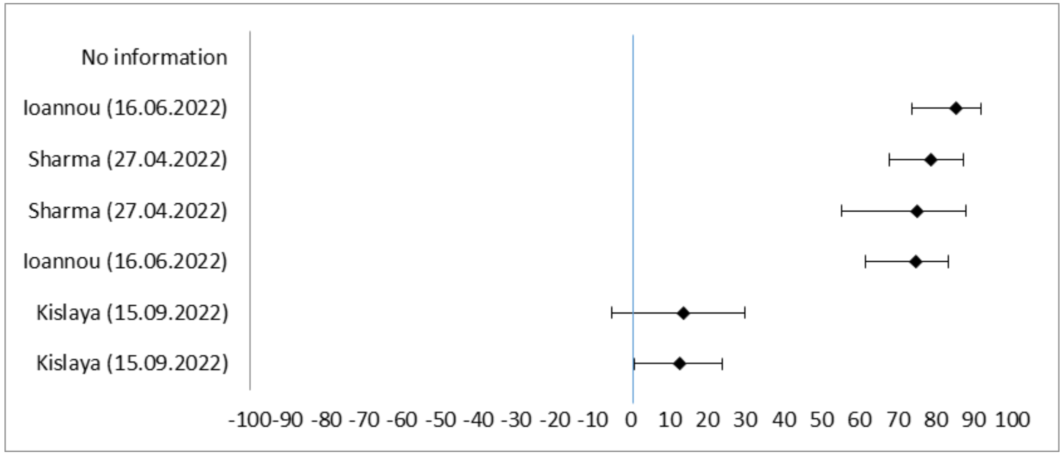

| Author (date) | VE in % | lower CI 95% | upper CI 95% |
|---|---|---|---|
| **No information** | | | |
| Ioannou (16.06.2022) | 85,5 | 73,7 | 92,0 |
| Sharma (27.04.2022) | 78,8 | 67,9 | 87,5 |
| Sharma (27.04.2022) | 75,0 | 55,4 | 88,0 |
| Ioannou (16.06.2022) | 74,8 | 61,8 | 83,3 |
| Kislaya (15.09.2022) | 14,0 | -5,0 | 30,0 |
| Kislaya (15.09.2022) | 13,0 | 1,0 | 24,0 |

**SF35:** Death: Older adults (60+ years), Exposure 1-3, Reference unvaccinated (n=4)

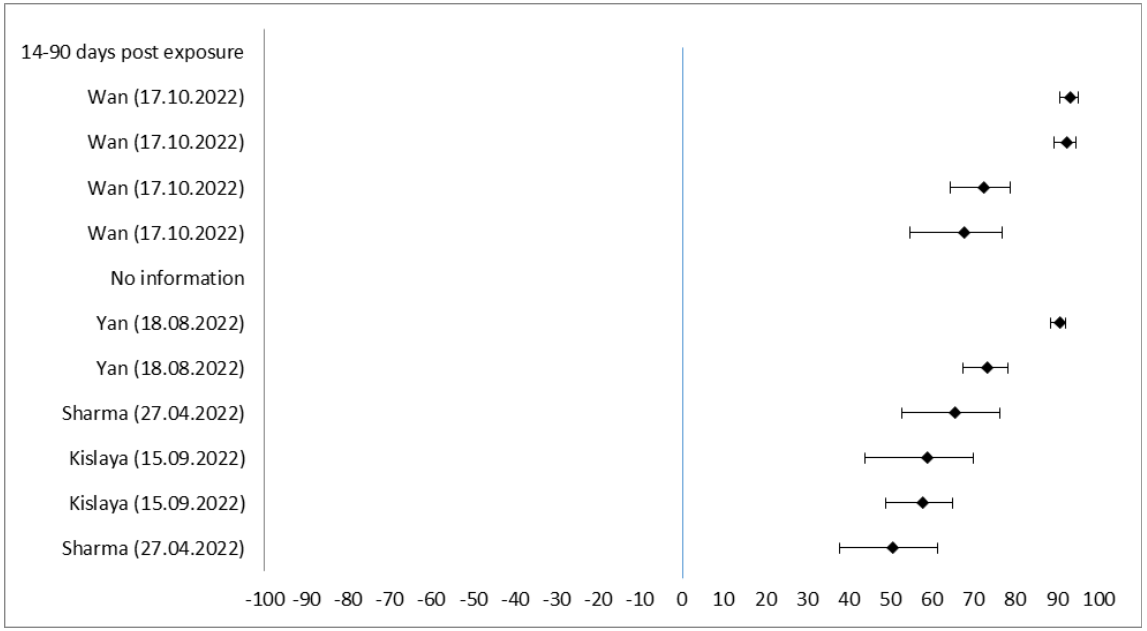

| Author (Date) | VE in % | lower CI 95% | upper CI 95% |
|---|---|---|---|
| **14-90 days post exposure** | | | |
| Wan (17.10.2022) | 93,4 | 90,7 | 95,3 |
| Wan (17.10.2022) | 92,5 | 89,3 | 94,7 |
| Wan (17.10.2022) | 72,6 | 64,4 | 79,0 |
| Wan (17.10.2022) | 67,9 | 54,9 | 77,1 |
| **No information** | | | |
| Yan (18.08.2022) | 90,7 | 88,6 | 92,3 |
| Yan (18.08.2022) | 73,5 | 67,7 | 78,3 |
| Sharma (27.04.2022) | 65,6 | 52,8 | 76,3 |
| Kislaya (15.09.2022) | 59,0 | 44,0 | 70,0 |
| Kislaya (15.09.2022) | 58,0 | 49,0 | 65,0 |
| Sharma (27.04.2022) | 50,7 | 37,9 | 61,6 |

**SF36:** Death: Children and adolescents (5-17 years), Exposure 1-3, Reference unvaccinated (n=1)

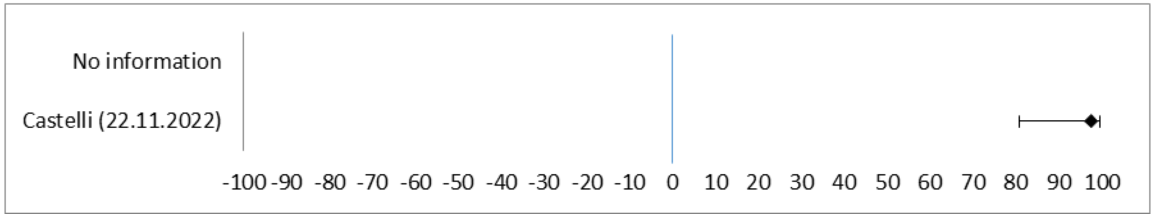

| Author (Date) | VE in % | lower CI 95% | lower CI 95% |
|---|---|---|---|
| **No information** | | | |
| Castelli (22.11.2022) | 97,6 | 81,0 | 99,7 |

### Literature

### Included primary studies (n=90)

identified via:

International Vaccine Access Center, Johns Hopkins Bloomberg School of Public Health, World Health Organization, Coalition for Epidemic Preparedness Innovations. Results of COVID-19 Vaccine Effectiveness Studies: An Ongoing Systematic Review: Weekly Summary Tables. Updated December 1, 2022. Available from: URL: https://view-hub.org/covid-19/effectiveness-studies

1. Abu-Raddad, Laith J.; Chemaitelly, Hiam; Ayoub, Houssein H.; AlMukdad, Sawsan; Yassine, Hadi M.; Al-Khatib, Hebah A. et al. (2022): Effect of mRNA Vaccine Boosters against SARS-CoV-2 Omicron Infection in Qatar. In: N Engl J Med 386 (19), S. 1804–1816. DOI: 10.1056/NEJMoa2200797.
2. Adams, Katherine; Rhoads, Jillian P.; Surie, Diya; Gaglani, Manjusha; Ginde, Adit A.; McNeal, Tresa et al. (2022): Vaccine Effectiveness of Primary Series and Booster Doses against Omicron Variant COVID-19-Associated Hospitalization in the United States. In: medRxiv : the preprint server for health sciences. DOI: 10.1101/2022.06.09.22276228.
3. Altarawneh, Heba N.; Chemaitelly, Hiam; Ayoub, Houssein H.; Tang, Patrick; Hasan, Mohammad R.; Yassine, Hadi M. et al. (2022): Effects of Previous Infection and Vaccination on Symptomatic Omicron Infections. In: The New England journal of medicine 387 (1), S. 21–34. DOI: 10.1056/NEJMoa2203965.
4. Amir, Ofra; Goldberg, Yair; Mandel, Micha; Bar-On, Yinon M.; Bodenheimer, Omri; Freedman, Laurence et al. (2022): Protection against omicron severe disease 0-7 months after BNT162b2 booster. DOI: 10.1101/2022.05.04.22274647.
5. Amir, Ofra; Goldberg, Yair; Mandel, Micha; Bar-On, Yinon M.; Bodenheimer, Omri; Freedman, Laurence et al. (2022): Initial protection against Omicron in children and adolescents by BNT162b2. DOI: 10.1101/2022.05.22.22275323.
6. Andersson, Niklas Worm; Thiesson, Emilia Myrup; Baum, Ulrike; Pihlström, Nicklas; Starrfelt, Jostein; Faksová, Kristýna et al. (2022): Comparative effectiveness of heterologous booster schedules with AZD1222, BNT162b2, or mRNA-1273 vaccines against COVID-19 during omicron predominance in the Nordic countries. DOI: 10.1101/2022.11.24.22282651.
7. Arashiro, Takeshi; Arima, Yuzo; Muraoka, Hirokazu; Sato, Akihiro; Oba, Kunihiro; Uehara, Yuki et al. (2022): COVID-19 vaccine effectiveness against symptomatic SARS-CoV-2 infection during Delta-dominant and Omicron-dominant periods in Japan: a multi-center prospective case-control study (FASCINATE study). In: Clinical infectious diseases : an official publication of the Infectious Diseases Society of America. DOI: 10.1093/cid/ciac635.
8. Arbel, Ronen; Sergienko, Ruslan; Friger, Michael; Peretz, Alon; Beckenstein, Tanya; Yaron, Shlomit et al. (2022): Effectiveness of a second BNT162b2 booster vaccine against hospitalization and death from COVID-19 in adults aged over 60 years. In: Nature medicine 28 (7), S. 1486–1490. DOI: 10.1038/s41591-022-01832-0.
9. Bar-On, Yinon M.; Goldberg, Yair; Mandel, Micha; Bodenheimer, Omri; Amir, Ofra; Freedman, Laurence et al. (2022): Protection by 4th dose of BNT162b2 against Omicron in Israel. DOI: 10.1101/2022.02.01.22270232.
10. Baum, Ulrike; Poukka, Eero; Leino, Tuija; Kilpi, Terhi; Nohynek, Hanna; Palmu, Arto A. (2022): High vaccine effectiveness against severe Covid-19 in the elderly in Finland before and after the emergence of Omicron. DOI: 10.1101/2022.03.11.22272140.
11. Breznik, Jessica A.; Rahim, Ahmad; Kajaks, Tara; Hagerman, Megan; Bilaver, Lucas; Colwill, Karen et al. (2022): Protection from Omicron infection in residents of nursing and retirement homes in Ontario, Canada. DOI: 10.1101/2022.06.28.22277016.
12. Britton, Amadea; Embi, Peter J.; Levy, Matthew E.; Gaglani, Manjusha; DeSilva, Malini B.; Dixon, Brian E. et al. (2022): Effectiveness of COVID-19 mRNA Vaccines Against COVID-19-Associated Hospitalizations Among Immunocompromised Adults During SARS-CoV-2 Omicron Predominance - VISION Network, 10 States, December 2021-August 2022. In: MMWR Morb. Mortal. Wkly. Rep. 71 (42), S. 1335–1342. DOI: 10.15585/mmwr.mm7142a4.
13. Buchan, Sarah A.; Chung, Hannah; Brown, Kevin A.; Austin, Peter C.; Fell, Deshayne B.; Gubbay, Jonathan B. et al. (2022): Estimated Effectiveness of COVID-19 Vaccines Against Omicron or Delta Symptomatic Infection and Severe Outcomes. In: JAMA network open 5 (9), e2232760. DOI: 10.1001/jamanetworkopen.2022.32760.
14. Buchan, Sarah A.; Nguyen, Lena; Wilson, Sarah E.; Kitchen, Sophie A.; Kwong, Jeffrey C. (2022): Vaccine effectiveness of BNT162b2 against Omicron and Delta outcomes in adolescents. DOI: 10.1101/2022.04.07.22273319.
15. Butt, Adeel A.; Talisa, Victor B.; Shaikh, Obaid S.; Omer, Saad B.; Mayr, Florian B. (2022): Relative Vaccine Effectiveness of a SARS-CoV-2 mRNA Vaccine Booster Dose Against the Omicron Variant. In: Clinical infectious diseases : an official publication of the Infectious Diseases Society of America. DOI: 10.1093/cid/ciac328.
16. Carazo, Sara; Skowronski, Danuta M.; Brisson, Marc; Sauvageau, Chantal; Brousseau, Nicholas; Gilca, Rodica et al. (2022): Protection against Omicron re-infection conferred by prior heterologous SARS-CoV-2 infection, with and without mRNA vaccination. DOI: 10.1101/2022.04.29.22274455.
17. Castelli, Juan Manuel; Rearte, Analia; Olszevicki, Santiago; Voto, Carla; Del Valle Juarez, María; Pesce, Martina et al. (2022): Effectiveness of mRNA-1273, BNT162b2, and BBIBP-CorV vaccines against infection and mortality in children in Argentina, during predominance of delta and omicron covid-19 variants: test negative, case-control study. In: BMJ (Clinical research ed.) 379, e073070. DOI: 10.1136/bmj-2022-073070.
18. Castillo Suarez, Milena; Khaoua, Hamid; Courtejoie, Noémie (2022): Vaccine-induced and naturally-acquired protection against Omicron and Delta symptomatic infection and severe COVID-19 outcomes, France, December 2021 to January 2022. In: Euro surveillance : bulletin Europeen sur les maladies transmissibles = European communicable disease bulletin 27 (16). DOI: 10.2807/1560-7917.ES.2022.27.16.2200250.
19. Cerqueira-Silva, Thiago; Shah, Syed Ahmar; Robertson, Chris; Sanchez, Mauro Niskier; Katikireddi, Srinivasa Vittal; Araújo Oliveira, Vinicius de et al. (2022): Waning of mRNA Boosters after Homologous Primary Series with BNT162b2 or ChadOx1 Against Symptomatic Infection and Severe COVID-19 in Brazil and Scotland: A Test-Negative Design Case-Control Study. In: SSRN Journal. DOI: 10.2139/ssrn.4082927.

20. Chatzilena, Anastasia; Hyams, Catherine; Challen, Robert; Marlow, Robin; King, Jade; Adegbite, David et al. (2022): Effectiveness of BNT162b2 COVID-19 Vaccination in Prevention of Hospitalisations and Severe Disease in Adults with Delta (B.1.617.2) and Omicron (B.1.1.529) Variant SARS-CoV-2 Infection: A Prospective Test Negative Case-Control Study. In: SSRN Journal. DOI: 10.2139/ssrn.4216690.
21. Chemaitelly, Hiam; AlMukdad, Sawsan; Ayoub, Houssein H.; Altarawneh, Heba N.; Coyle, Peter; Tang, Patrick et al. (2022): Effectiveness of the BNT162b2 vaccine against SARS-CoV-2 infection among children and adolescents in Qatar. DOI: 10.1101/2022.07.26.22278045.
22. Chemaitelly, Hiam; Ayoub, Houssein H.; AlMukdad, Sawsan; Coyle, Peter; Tang, Patrick; Yassine, Hadi M. et al. (2022): Duration of mRNA vaccine protection against SARS-CoV-2 Omicron BA.1 and BA.2 subvariants in Qatar. In: Nature communications 13 (1), S. 3082. DOI: 10.1038/s41467-022-30895-3.
23. Chemaitelly, Hiam; Ayoub, Houssein H.; Tang, Patrick; Coyle, Peter; Yassine, Hadi M.; Al Thani, Asmaa A. et al. (2022): COVID-19 primary series and booster vaccination and immune imprinting. DOI: 10.1101/2022.10.31.22281756.
24. Chemaitelly, Hiam; Ayoub, Houssein H.; Tang, Patrick; Coyle, Peter; Yassine, Hadi M.; Al Thani, Asmaa A. et al. (2022): Long-term COVID-19 booster effectiveness by infection history and clinical vulnerability and immune imprinting. DOI: 10.1101/2022.11.14.22282103.
25. Chemaitelly, Hiam; Nagelkerke, Nico; Ayoub, Houssein H.; Coyle, Peter; Tang, Patrick; Yassine, Hadi M. et al. (2022): Duration of immune protection of SARS-CoV-2 natural infection against reinfection in Qatar. DOI: 10.1101/2022.07.06.22277306.
26. Chiew, Calvin J.; Premikha, M.; Chong, Chia Yin; Wei, Wycliffe E.; Ong, Benjamin; Lye, David Chien et al. (2022): Effectiveness of primary series and booster vaccination against SARS-CoV-2 infection and hospitalisation among adolescents aged 12–17 years in Singapore: a national cohort study. In: The Lancet Infectious Diseases. DOI: 10.1016/S1473-3099(22)00573-4.
27. Cocchio, Silvia; Zabeo, Federico; Tremolada, Giulia; Facchin, Giacomo; Venturato, Giovanni; Marcon, Thomas et al. (2022): COVID-19 Vaccine Effectiveness against Omicron Variant among Underage Subjects: The Veneto Region's Experience. In: Vaccines 10 (8). DOI: 10.3390/vaccines10081362.
28. Cohen-Stavi, Chandra J.; Magen, Ori; Barda, Noam; Yaron, Shlomit; Peretz, Alon; Netzer, Doron et al. (2022): BNT162b2 Vaccine Effectiveness against Omicron in Children 5 to 11 Years of Age. In: N Engl J Med 387 (3), S. 227–236. DOI: 10.1056/NEJMoa2205011.
29. Fabiani, Massimo; Mateo-Urdiales, Alberto; Sacco, Chiara; Rota, Maria Cristina; Petrone, Daniele; Bressi, Marco et al. (2022): Relative effectiveness of a 2nd booster dose of COVID-19 mRNA vaccine up to four months post administration in individuals aged 80 years or more in Italy: A retrospective matched cohort study. In: Vaccine. DOI: 10.1016/j.vaccine.2022.11.013.
30. Ferdinands, Jill M.; Rao, Suchitra; Dixon, Brian E.; Mitchell, Patrick K.; DeSilva, Malini B.; Irving, Stephanie A. et al. (2022): Waning of vaccine effectiveness against moderate and severe covid-19 among adults in the US from the VISION network: test negative, case-control study. In: BMJ 379, e072141. DOI: 10.1136/bmj-2022-072141.
31. Florentino, Pilar T. V.; Millington, Tristan; Cerqueira-Silva, Thiago; Robertson, Chris; Araújo Oliveira, Vinicius de; Júnior, Juracy B. S. et al. (2022): Vaccine effectiveness of two-dose BNT162b2 against symptomatic and severe COVID-19 among adolescents in Brazil and Scotland over time: a test-negative case-control study. In: The Lancet Infectious Diseases. DOI: 10.1016/S1473-3099(22)00451-0.
32. Fowlkes, Ashley L.; Yoon, Sarang K.; Lutrick, Karen; Gwynn, Lisa; Burns, Joy; Grant, Lauren et al. (2022): Effectiveness of 2-Dose BNT162b2 (Pfizer BioNTech) mRNA Vaccine in Preventing SARS-CoV-2 Infection Among Children Aged 5-11 Years and Adolescents Aged 12-15 Years - PROTECT Cohort, July 2021-February 2022. In: MMWR Morb. Mortal. Wkly. Rep. 71 (11), S. 422–428. DOI: 10.15585/mmwr.mm7111e1.
33. Gazit, Sivan; Saciuk, Yaki; Perez, Galit; Peretz, Asaf; Pitzer, Virginia E.; Patalon, Tal (2022): Short term, relative effectiveness of four doses versus three doses of BNT162b2 vaccine in people aged 60 years and older in Israel: retrospective, test negative, case-control study. In: BMJ 377, e071113. DOI: 10.1136/bmj-2022-071113.
34. González, Soledad; Olszevicki, Santiago; Gaiano, Alejandra; Baino, Ana Nina Varela; Regairaz, Lorena; Salazar, Martín et al. (2022): Effectiveness of BBIBP-CorV, BNT162b2 and mRNA-1273 vaccines against hospitalisations among children and adolescents during the Omicron outbreak in Argentina: A retrospective cohort study. In: Lancet Regional Health. Americas 13, S. 100316. DOI: 10.1016/j.lana.2022.100316.
35. González, Soledad; Olszevicki, Santiago; Gaiano, Alejandra; Salazar, Martín; Regairaz, Lorena; Varela Baino, Ana Nina et al. (2022): Protection of homologous and heterologous boosters after primary schemes of rAd26-rAd5, ChAdOx1 nCoV-19 and BBIBP-CorV during the Omicron outbreak in adults of 50 years and older in Argentina: a test-negative case-control study. DOI: 10.1101/2022.09.25.22280341.
36. Gram, Mie Agermose; Emborg, Hanne-Dorthe; Schelde, Astrid Blicher; Friis, Nikolaj Ulrik; Nielsen, Katrine Finderup; Moustsen-Helms, Ida Rask et al. (2022): Vaccine effectiveness against SARS-CoV-2 infection or COVID-19 hospitalization with the Alpha, Delta, or Omicron SARS-CoV-2 variant: A nationwide Danish cohort study. In: PLoS medicine 19 (9), e1003992. DOI: 10.1371/journal.pmed.1003992.
37. Grewal, Ramandip; Kitchen, Sophie A.; Nguyen, Lena; Buchan, Sarah A.; Wilson, Sarah E.; Costa, Andrew P.; Kwong, Jeffrey C. (2022): Effectiveness of a fourth dose of covid-19 mRNA vaccine against the omicron variant among long term care residents in Ontario, Canada: test negative design study. In: BMJ 378, e071502. DOI: 10.1136/bmj-2022-071502.
38. Grewal, Ramandip; Nguyen, Lena; Buchan, Sarah A.; Wilson, Sarah E.; Costa, Andrew P.; Kwong, Jeffrey C. (2022): Effectiveness and Duration of Protection of a Fourth Dose of COVID-19 mRNA Vaccine among Long-Term Care Residents in Ontario, Canada. DOI: 10.1101/2022.09.29.22280526.
39. Grewal, Ramandip; Nguyen, Lena; Buchan, Sarah A.; Wilson, Sarah E.; Nasreen, Sharifa; Austin, Peter C. et al. (2022): Effectiveness of mRNA COVID-19 vaccine booster doses against Omicron severe outcomes. DOI: 10.1101/2022.10.31.22281766.
40. Hansen, Christian Holm; Friis, Nikolaj Ulrik; Bager, Peter; Stegger, Marc; Fonager, Jannik; Fomsgaard, Anders et al. (2022): Risk of Reinfection, Vaccine Protection, and Severity of Infection with the BA.5 Omicron Subvariant: A Danish Nation-Wide Population-Based Study. In: SSRN Journal. DOI: 10.2139/ssrn.4165630.
41. Hansen, Christian; Schelde, Astrid; Moustsen-Helm, Ida; Embor, Hanne-Dorthe; Eriksen, Rasmus; Stegger, Marc et al. (2022): Vaccine effectiveness against infection and COVID-19-associated hospitalisation with the Omicron (B.1.1.529) variant after vaccination with the BNT162b2 or mRNA-1273 vaccine: A nationwide Danish cohort study. DOI: 10.21203/rs.3.rs-1486018/v1.

42. Ioannou, George N.; Bohnert, Amy S. B.; O'Hare, Ann M.; Boyko, Edward J.; Maciejewski, Matthew L.; Smith, Valerie A. et al. (2022): Effectiveness of mRNA COVID-19 vaccine boosters against infection, hospitalization and death: a target trial emulation in the omicron (B.1.1.529) variant era. DOI: 10.1101/2022.06.15.22276466.
43. Ionescu, Iulia G.; Skowronski, Danuta M.; Sauvageau, Chantal; Chuang, Erica; Ouakki, Manale; Kim, Shinhye; Serres, Gaston de (2022): BNT162b2 effectiveness against Delta & Omicron variants in teens by dosing interval and duration. DOI: 10.1101/2022.06.27.22276790.
44. Jara, Alejandro; Cuadrado, Cristobal; Undurraga, Eduardo A.; García, Christian; Nájera, Manuel; Bertoglia, María Paz et al. (2022): Effectiveness and duration of a second COVID-19 vaccine booster. DOI: 10.1101/2022.10.03.22280660.
45. Kim, Sara S.; Chung, Jessie R.; Talbot, H. Keipp; Grijalva, Carlos G.; Wernli, Karen J.; Kiniry, Erika et al. (2022): Effectiveness of 2 and 3 mRNA COVID-19 Vaccines Doses against Omicron and Delta-Related Outpatient Illness among Adults, October 2021 – February 2022 7. DOI: 10.1101/2022.04.06.22273535.
46. Kirsebom, Freja; Andrews, Nick; Sachdeva, Ruchira; Stowe, Julia; Ramsay, Mary; Bernal, Jamie Lopez (2022): Effectiveness of ChAdOx1-S COVID-19 Booster Vaccination against the Omicron and Delta variants in England. DOI: 10.1101/2022.04.29.22274483.
47. Kislaya, Irina; Machado, Ausenda; Magalhães, Sarah; Rodrigues, Ana Paula; Franco, Rafael; Leite, Pedro Pinto et al. (2022): COVID-19 mRNA vaccine effectiveness (second and first booster dose) against hospitalisation and death during Omicron BA.5 circulation: cohort study based on electronic health records, Portugal, May to July 2022. In: Euro surveillance : bulletin Europeen sur les maladies transmissibles = European communicable disease bulletin 27 (37). DOI: 10.2807/1560-7917.ES.2022.27.37.2200697.
48. Klein, Nicola P.; Stockwell, Melissa S.; Demarco, Maria; Gaglani, Manjusha; Kharbanda, Anupam B.; Irving, Stephanie A. et al. (2022): Effectiveness of COVID-19 Pfizer-BioNTech BNT162b2 mRNA Vaccination in Preventing COVID-19-Associated Emergency Department and Urgent Care Encounters and Hospitalizations Among Nonimmunocompromised Children and Adolescents Aged 5-17 Years - VISION Network, 10 States, April 2021-January 2022. In: MMWR Morb. Mortal. Wkly. Rep. 71 (9), S. 352–358. DOI: 10.15585/mmwr.mm7109e3.
49. Lauring, Adam S.; Tenforde, Mark W.; Chappell, James D.; Gaglani, Manjusha; Ginde, Adit A.; McNeal, Tresa et al. (2022): Clinical severity of, and effectiveness of mRNA vaccines against, covid-19 from omicron, delta, and alpha SARS-CoV-2 variants in the United States: prospective observational study. In: BMJ 376, e069761. DOI: 10.1136/bmj-2021-069761.
50. Lin, Dan-Yu; Gu, Yu; Xu, Yangjianchen; Wheeler, Bradford; Young, Hayley; Sunny, Shadia Khan et al. (2022): Association of Primary and Booster Vaccination and Prior Infection With SARS-CoV-2 Infection and Severe COVID-19 Outcomes. In: JAMA 328 (14), S. 1415–1426. DOI: 10.1001/jama.2022.17876.
51. Lind, Margaret L.; Robertson, Alexander James; Silva, Julio; Warner, Frederick; Coppi, Andreas C.; Price, Nathan et al. (2022): Effectiveness of Primary and Booster COVID-19 mRNA Vaccination against Omicron Variant SARS-CoV-2 Infection in People with a Prior SARS-CoV-2 Infection 397. DOI: 10.1101/2022.04.19.22274056.
52. Link-Gelles, Ruth; Ciesla, Allison Avrich; Fleming-Dutra, Katherine E.; Smith, Zachary R.; Britton, Amadea; Wiegand, Ryan E. et al. (2022): Effectiveness of Bivalent mRNA Vaccines in Preventing Symptomatic SARS-CoV-2 Infection - Increasing Community Access to Testing Program, United States, September-November 2022. In: MMWR Morb. Mortal. Wkly. Rep. 71 (48), S. 1526–1530. DOI: 10.15585/mmwr.mm7148e1.
53. Link-Gelles, Ruth; Levy, Matthew E.; Gaglani, Manjusha; Irving, Stephanie A.; Stockwell, Melissa; Dascomb, Kristin et al. (2022): Effectiveness of 2, 3, and 4 COVID-19 mRNA Vaccine Doses Among Immunocompetent Adults During Periods when SARS-CoV-2 Omicron BA.1 and BA.2/BA.2.12.1 Sublineages Predominated - VISION Network, 10 States, December 2021-June 2022. In: MMWR. Morbidity and mortality weekly report 71 (29), S. 931–939. DOI: 10.15585/mmwr.mm7129e1.
54. Link-Gelles, Ruth; Levy, Matthew E.; Natarajan, Karthik; Reese, Sarah E.; Naleway, Allison L.; Grannis, Shaun J. et al. (2022): Association between COVID-19 mRNA vaccination and COVID-19 illness and severity during Omicron BA.4 and BA.5 sublineage periods. DOI: 10.1101/2022.10.04.22280459.
55. Liu, Bette; Gidding, Heather; Stepien, Sandrine; Cretikos, Michelle; Macartney, Kristine (2022): Relative Effectiveness of COVID-19 Vaccination with 3 Compared to 2 Doses Against SARS-CoV-2 B.1.1.529 (Omicron) Among an Australian Population with Low Prior Rates of SARS-CoV-2 Infection. In: SSRN Journal. DOI: 10.2139/ssrn.4142075.
56. Magen, Ori; Waxman, Jacob G.; Makov-Assif, Maya; Vered, Roni; Dicker, Dror; Hernán, Miguel A. et al. (2022): Fourth Dose of BNT162b2 mRNA Covid-19 Vaccine in a Nationwide Setting. In: N Engl J Med 386 (17), S. 1603–1614. DOI: 10.1056/NEJMoa2201688.
57. McConeghy, Kevin W.; White, Elizabeth M.; Blackman, Carolyn; Santostefano, Christopher M.; Lee, Yoojin; Rudolph, James L. et al. (2022): Effectiveness of a Second COVID-19 Vaccine Booster Dose Against Infection, Hospitalization, or Death Among Nursing Home Residents - 19 States, March 29-July 25, 2022. In: MMWR Morb. Mortal. Wkly. Rep. 71 (39), S. 1235–1238. DOI: 10.15585/mmwr.mm7139a2.
58. Mimura, Wataru; Ishiguro, Chieko; Maeda, Megumi; Murata, Fumiko; Fukuda, Haruhisa (2022): Effectiveness of messenger RNA vaccines against infection with SARS-CoV-2 during the periods of Delta and Omicron variant predominance in Japan: the Vaccine Effectiveness, Networking, and Universal Safety (VENUS) study. In: International journal of infectious diseases : IJID : official publication of the International Society for Infectious Diseases 125, S. 58–60. DOI: 10.1016/j.ijid.2022.10.001.
59. Monge, Susana; Rojas-Benedicto, Ayelén; Olmedo, Carmen; Mazagatos, Clara; José Sierra, María; Limia, Aurora et al. (2022): Effectiveness of mRNA vaccine boosters against infection with the SARS-CoV-2 omicron (B.1.1.529) variant in Spain: a nationwide cohort study. In: The Lancet Infectious Diseases 22 (9), S. 1313–1320. DOI: 10.1016/S1473-3099(22)00292-4.
60. Montez-Rath, Maria E.; Garcia, Pablo; Han, Jialin; Cadden, LinaCel; Hunsader, Patti; Morgan, Curt et al. (2022): SARS-CoV-2 infection during the Omicron surge among patients receiving dialysis: the role of circulating receptor-binding domain antibodies and vaccine doses. In: medRxiv. DOI: 10.1101/2022.03.15.22272426.
61. Muhsen, Khitam; Maimon, Nimrod; Mizrahi, Amiel Yaron; Boltyansky, Boris; Bodenheimer, Omri; Diamant, Zafrira Hillel et al. (2022): Association of Receipt of the Fourth BNT162b2 Dose With Omicron Infection and COVID-19 Hospitalizations Among Residents of Long-term Care Facilities. In: INTEMED 182 (8), S. 859–867. DOI: 10.1001/jamainternmed.2022.2658.
62. Ng, Oon Tek; Marimuthu, Kalisvar; Lim, Nigel; Lim, Ze Qin; Thevasagayam, Natascha May; Koh, Vanessa et al. (2022): Analysis of COVID-19 Incidence and Severity Among Adults Vaccinated With 2-Dose mRNA COVID-19 or Inactivated SARS-CoV-2 Vaccines With and Without Boosters in Singapore. In: JAMA network open 5 (8), e2228900. DOI: 10.1001/jamanetworkopen.2022.28900.

63. Norddahl, Gudmundur L.; Melsted, Pall; Gunnarsdottir, Kristbjorg; Halldorsson, Gisli H.; Olafsdottir, Thorunn A.; Gylfason, Arnaldur et al. (2022): Effect of booster vaccination against Delta and Omicron variants in Iceland. DOI: 10.1101/2022.02.26.22271509.
64. Nordström, Peter; Ballin, Marcel; Nordström, Anna (2022): Safety and effectiveness of COVID-19 mRNA vaccination and risk factors for hospitalisation caused by the omicron variant in 0.8 million adolescents: A nationwide cohort study in Sweden. DOI: 10.1101/2022.10.19.22281286.
65. Patalon, Tal; Saciuk, Yaki; Peretz, Asaf; Perez, Galit; Lurie, Yoav; Maor, Yasmin; Gazit, Sivan (2022): Waning effectiveness of the third dose of the BNT162b2 mRNA COVID-19 vaccine. In: Nature communications 13 (1), S. 3203. DOI: 10.1038/s41467-022-30884-6.
66. Petrie, Joshua G.; King, Jennifer P.; McClure, David L.; Rolfes, Melissa A.; Meece, Jennifer K.; Belongia, Edward A.; McLean, Huong Q. (2022): Effectiveness of COVID-19 mRNA vaccine booster dose relative to primary series during a period of Omicron circulation. DOI: 10.1101/2022.04.15.22273915.
67. Piché-Renaud, Pierre-Philippe; Swayze, Sarah; Buchan, Sarah; Wilson, Sarah; Austin, Peter C.; Morris, Shaun K. et al. (2022): Vaccine Effectiveness of BNT162b2 Against Omicron in Children Aged 5-11 Years: A Test-Negative Design. In: SSRN Journal. DOI: 10.2139/ssrn.4176388.
68. Plumb, Ian D.; Feldstein, Leora R.; Barkley, Eric; Posner, Alexander B.; Bregman, Howard S.; Hagen, Melissa Briggs; Gerhart, Jacqueline L. (2022): Effectiveness of COVID-19 mRNA Vaccination in Preventing COVID-19-Associated Hospitalization Among Adults with Previous SARS-CoV-2 Infection - United States, June 2021-February 2022. In: MMWR. Morbidity and mortality weekly report 71 (15), S. 549–555. DOI: 10.15585/mmwr.mm7115e2.
69. Powell, Annabel A.; Kirsebom, Freja; Stowe, Julia; Ramsay, Mary E.; Lopez-Bernal, Jamie; Andrews, Nick; Ladhani, Shamez N. (2022): Protection against symptomatic disease with the delta and omicron BA.1/BA.2 variants of SARS-CoV-2 after infection and vaccination in adolescents: national observational test-negative case control study, August 2021 to March 2022, England. DOI: 10.1101/2022.08.19.22278987.
70. Price, Ashley M.; Olson, Samantha M.; Newhams, Margaret M.; Halasa, Natasha B.; Boom, Julie A.; Sahni, Leila C. et al. (2022): BNT162b2 Protection against the Omicron Variant in Children and Adolescents. In: N Engl J Med 386 (20), S. 1899–1909. DOI: 10.1056/NEJMoa2202826.
71. Rennert, Lior; Ma, Zichen; McMahan, Christopher S.; Dean, Delphine (2022): Covid-19 vaccine effectiveness against general SARS-CoV-2 infection from the omicron variant: A retrospective cohort study. DOI: 10.1101/2022.05.06.22274771.
72. Risk, Malcolm; Hayek, Salim S.; Schiopu, Elena; Yuan, Liyang; Shen, Chen; Shi, Xu et al. (2022): COVID-19 vaccine effectiveness against omicron (B.1.1.529) variant infection and hospitalisation in patients taking immunosuppressive medications: a retrospective cohort study. In: The Lancet Rheumatology 383, S. 2603. DOI: 10.1016/S2665-9913(22)00216-8.
73. Rudan, Igor; Millington, Tristan; Antal, Karen; Grange, Zoe; Fenton, Lynda; Sullivan, Christopher et al. (2022): BNT162b2 COVID-19 vaccination uptake, safety, effectiveness and waning in children and young people aged 12-17 years in Scotland. In: The Lancet regional health. Europe 23, S. 100513. DOI: 10.1016/j.lanepe.2022.100513.
74. Sharma, Aditya; Oda, Gina; Holodniy, Mark (2022): Effectiveness of mRNA-based vaccines during the emergence of SARS-CoV-2 Omicron variant. In: Clinical infectious diseases : an official publication of the Infectious Diseases Society of America. DOI: 10.1093/cid/ciac325.
75. Šmíd, Martin; Berec, Luděk; Májek, Ondřej; Pavlík, Tomáš; Jarkovský, Jiří; Weiner, Jakub et al. (2022): Protection by vaccines and previous infection against the Omicron variant of SARS-CoV-2. DOI: 10.1101/2022.02.24.22271396.
76. Stowe, Julia; Andrews, Nick; Kirsebom, Freja; Ramsay, Mary; Bernal, Jamie Lopez (2022): Effectiveness of COVID-19 vaccines against Omicron and Delta hospitalisation: test negative case-control study 386. DOI: 10.1101/2022.04.01.22273281.
77. Surie, Diya; Bonnell, Levi; Adams, Katherine; Gaglani, Manjusha; Ginde, Adit A.; Douin, David J. et al. (2022): Effectiveness of Monovalent mRNA Vaccines Against COVID-19-Associated Hospitalization Among Immunocompetent Adults During BA.1/BA.2 and BA.4/BA.5 Predominant Periods of SARS-CoV-2 Omicron Variant in the United States - IVY Network, 18 States, December 26, 2021-August 31, 2022. In: MMWR Morb. Mortal. Wkly. Rep. 71 (42), S. 1327–1334. DOI: 10.15585/mmwr.mm7142a3.
78. Tan, Celine Y.; Chiew, Calvin J.; Lee, Vernon J.; Ong, Benjamin; Lye, David Chien; Tan, Kelvin Bryan (2022): Effectiveness of a Fourth Dose of COVID-19 mRNA Vaccine Against Omicron Variant Among Elderly People in Singapore. In: Annals of internal medicine. DOI: 10.7326/M22-2042.
79. Tartof, Sara Y.; Frankland, Timothy B.; Slezak, Jeff M.; Puzniak, Laura; Hong, Vennis; Xie, Fagen et al. (2022): Effectiveness Associated With BNT162b2 Vaccine Against Emergency Department and Urgent Care Encounters for Delta and Omicron SARS-CoV-2 Infection Among Adolescents Aged 12 to 17 Years. In: JAMA network open 5 (8), e2225162. DOI: 10.1001/jamanetworkopen.2022.25162.
80. Tartof, Sara Y.; Slezak, Jeff M.; Puzniak, Laura; Hong, Vennis; Xie, Fagen; Ackerson, Bradley K. et al. (2022): BNT162b2 Effectiveness and Durability Against BA.1 and BA.2 Hospital and Emergency Department Admissions in a Large US Health System: A Test-Negative Design. In: SSRN Journal. DOI: 10.2139/ssrn.4150500.
81. Tartof, Sara Y.; Slezak, Jeff M.; Puzniak, Laura; Hong, Vennis; Xie, Fagen; Ackerson, Bradley K. et al. (2022): Durability of BNT162b2 vaccine against hospital and emergency department admissions due to the omicron and delta variants in a large health system in the USA: a test-negative case–control study. In: The Lancet Respiratory Medicine 10 (7), S. 689–699. DOI: 10.1016/S2213-2600(22)00101-1.
82. Tsang, Nicole Ngai Yung; So, Hau Chi; Cowling, Benjamin J.; Leung, Gabriel; Ip, Dennis Kai Ming (2022): Effectiveness of BNT162b2 and CoronaVac COVID-19 Vaccination Against Asymptomatic and Symptomatic Infection of SARS-CoV-2 Omicron BA.2 in Hong Kong. In: SSRN Journal. DOI: 10.2139/ssrn.4200539.
83. Tseng, Hung Fu; Ackerson, Bradley K.; Bruxvoort, Katia J.; Sy, Lina S.; Tubert, Julia E.; Lee, Gina S. et al. (2022): Effectiveness of mRNA-1273 against infection and COVID-19 hospitalization with SARS-CoV-2 Omicron subvariants: BA.1, BA.2, BA.2.12.1, BA.4, and BA.5. DOI: 10.1101/2022.09.30.22280573.
84. Tseng, Hung Fu; Ackerson, Bradley K.; Luo, Yi; Sy, Lina S.; Talarico, Carla A.; Tian, Yun et al. (2022): Effectiveness of mRNA-1273 against SARS-CoV-2 Omicron and Delta variants. In: Nature medicine 28 (5), S. 1063–1071. DOI: 10.1038/s41591-022-01753-y.
85. Veneti, Lamprini; Berild, Jacob Dag; Watle, Sara Viksmoen; Starrfelt, Jostein; Greve-Isdahl, Margrethe; Langlete, Petter et al. (2022): Vaccine effectiveness with BNT162b2 (Comirnaty, Pfizer-BioNTech) vaccine against reported SARS-CoV-2 Delta and Omicron infection among adolescents, Norway, August 2021 to January 2022. DOI: 10.1101/2022.03.24.22272854.

86. Wan, Eric Yuk Fai; Mok, Anna Hoi Ying; Yan, Vincent Ka Chun; Ying Chan, Cheyenne I.; Wang, Boyuan; Lai, Francisco Tsz Tsun et al. (2022): Effectiveness of BNT162b2 and CoronaVac vaccinations against SARS-CoV-2 omicron infection in people aged 60 years or above: a case-control study. In: Journal of travel medicine. DOI: 10.1093/jtm/taac119.
87. Wang, Xiaofeng; Zein, Joe; Ji, Xinge; Lin, Dan-Yu (2022): Impact of Vaccination, Prior Infection, and Therapy on Delta and Omicron Variants 62. DOI: 10.1101/2022.03.24.22272901.
88. Yan, Vincent Ka Chun; Wan, Eric Yuk Fai; Ye, Xuxiao; Mok, Anna Hoi Ying; Lai, Francisco Tsz Tsun; Chui, Celine Sze Ling et al. (2022): Effectiveness of BNT162b2 and CoronaVac vaccinations against mortality and severe complications after SARS-CoV-2 Omicron BA.2 infection: a case-control study. In: Emerging microbes & infections, S. 1–48. DOI: 10.1080/22221751.2022.2114854.
89. Young-Xu, Yinong; Zwain, Gabrielle M.; Izurieta, Hector S.; Korves, Caroline; Powell, Ethan I.; Smith, Jeremy et al. (2022): Effectiveness of mRNA COVID-19 vaccines against Omicron and Delta variants in a matched test-negative case–control study among US veterans. In: BMJ Open 12 (8). DOI: 10.1136/bmjopen-2022-063935.
90. Zambrano, Laura D.; Newhams, Margaret M.; Olson, Samantha M.; Halasa, Natasha B.; Price, Ashley M.; Orzel, Amber O. et al. (2022): BNT162b2 mRNA Vaccination Against COVID-19 is Associated with Decreased Likelihood of Multisystem Inflammatory Syndrome in U.S. Children Ages 5-18 Years. In: Clinical infectious diseases: an official publication of the Infectious Diseases Society of America. DOI: 10.1093/cid/ciac637.

Supplement 2 Table S8: Excluded primary studies with reason for exclusion (n=48)

| No. | Author (date) | Reason for exclusion | Category |
|---|---|---|---|
| 1 | Altarawneh (06.01.2022) | Date (inclusion from 12.02.2022, RKI Review) | Date |
| 2 | Buchan (28.01.2022) | Date (inclusion from 12.02.2022, RKI Review) | Date |
| 3 | Canetti (09.11.2022) | Editorial | Article Format |
| 4 | Carazo (27.06.2022) | HCW | Study population |
| 5 | Carlsen (01.06.2022) | Newborns/Infants | Study population |
| 6 | Cerqueira-Silva (01.07.2022) | Correspondence | Article Format |
| 7 | Cerquerira-Silva (18.07.2022) | Vaccine CoronaVac | Vaccine type |
| 8 | Cheng (11.08.2022) | Letter | Article format |
| 9 | Chin (27.05.2022) | Prison Inmates | Study population |
| 10 | Cohen (02.08.2022) | HCW | Study population |
| 11 | Collie (14.09.2022) | Editorial | Article format |
| 12 | Collie (29.12.2021) | Date (inclusion from 12.02.2022, RKI Review) | Date |
| 13 | Consonni (19.10.2022) | HCW | Study population |
| 14 | Ferdinands (11.02.2022) | Date (inclusion from 12.02.2022, RKI Review) | Date |
| 15 | Florentino (13.08.2022) | Vaccine CoronaVac | Vaccine type |
| 16 | Glatman-Freedman (31.03.2022) | not Omicron-specific | Study population |
| 17 | Gray (09.06.2022) | HCW | Study population |
| 18 | Guedalia (11.07.2022) | Pregnant women | Study population |
| 19 | Halasa (22.06.2022) | Infants | Study population |
| 20 | Hansen (23.12.2021) | Date (inclusion from 02/12/2022, RKI review) | Date |
| 21 | Hertz (15.08.2022) | HCW | Study population |
| 22 | Huang (09.09.2022) | Vaccine Ad5-nCoV, BBIBP-CorV, CoronaVac | Vaccine type |
| 23 | Jara (23.05.2022) | CoronaVac vaccine | Vaccine type |
| 24 | Jorgensen (08.11.2022) | Newborns/infants | Study population |
| 25 | Kim (29.08.2022) | HCW | Study population |
| 26 | Moghnieh (22.09.2022) | Airline employees | Study population |
| 27 | Natarajan (29.03.2022) | Report | Article format |
| 28 | Penayo (06/2022) | No access to full text | Article format |
| 29 | Powell (21.03.2022) | Comment | Article format |
| 30 | Ranzani (16.08.2022) | Vaccine CoronaVac | Vaccine type |
| 31 | Regev-Yochay (15.02.2022) | HCW | Study population |
| 32 | Richterman (06.06.2022) | HCW | Study population |
| 33 | Risk (08.10.2022) | No access to full text | Article format |

| | | | |
|---|---|---|---|
| 34 | Schrag (26.09.2022) | Pregnant | Study population |
| 35 | Simwanza (08.06.2022) | Prison inmates | Study population |
| 36 | Spensley (26.01.2022) | Date (inclusion from 12.02.2022, RKI Review). | Date |
| 37 | Stirrup (09.08.2022) | HCW | Study population |
| 38 | Suah (02.05.2022) | Letter | Article Format |
| 39 | Tan (13.09.2022) | Letter | Article format |
| 40 | Tartof (25.10.2022) | Correspondence | Article format |
| 41 | Tenforde (25.03.2022) | Report | Article format |
| 42 | Thompson (21.01.2022) | Date (Inclusion from 12.02.2022, RKI Review) | Date |
| 43 | UKHSA (03.11.2022) | Report | Article format |
| 44 | UKHSA (14.01.2022) | Report | Article format |
| 45 | Wan (17.08.2022) | Letter | Article format |
| 46 | Willet (26.01.2022) | Date (inclusion from 12.02.2022, RKI Review) | Date |
| 47 | Yoon (06.04.2022) | HCW | Study population |
| 48 | Zerbo (18.10.2022) | Newborns/infants | Study population |

1. Altarawneh, Heba; Chemaitelly, Hiam; Tang, Patrick; Hasan, Mohammad R.; Qassim, Suelen; Ayoub, Houssein H. et al. (2022): Protection afforded by prior infection against SARS-CoV-2 reinfection with the Omicron variant. DOI: 10.1101/2022.01.05.22268782.
2. Buchan, Sarah A.; Chung, Hannah; Brown, Kevin A.; Austin, Peter C.; Fell, Deshayne B.; Gubbay, Jonathan B. et al. (2022): Effectiveness of COVID-19 vaccines against Omicron or Delta symptomatic infection and severe outcomes. DOI: 10.1101/2021.12.30.21268565.
3. Canetti, Michal; Barda, Noam; Gilboa, Mayan; Indenbaum, Victoria; Asraf, Keren; Gonen, Tal et al. (2022): Six-Month Follow-up after a Fourth BNT162b2 Vaccine Dose. In: *N Engl J Med* 387 (22), S. 2092–2094. DOI: 10.1056/NEJMc2211283.
4. Carazo, Sara; Skowronski, Danuta M.; Brisson, Marc; Barkati, Sapha; Sauvageau, Chantal; Brousseau, Nicholas et al. (2022): Protection against Omicron BA.2 reinfection conferred by primary Omicron or pre-Omicron infection with and without mRNA vaccination. DOI: 10.1101/2022.06.23.22276824.
5. Carlsen, Ellen Øen; Magnus, Maria C.; Oakley, Laura; Fell, Deshayne B.; Greve-Isdahl, Margrethe; Kinge, Jonas Minet; Håberg, Siri E. (2022): Association of COVID-19 Vaccination During Pregnancy With Incidence of SARS-CoV-2 Infection in Infants. In: *INTEMED* 182 (8), S. 825–831. DOI: 10.1001/jamainternmed.2022.2442.
6. Cerqueira-Silva, Thiago; Araujo Oliveira, Vinicius de; Paixão, Enny S.; Florentino, Pilar Tavares Veras; Penna, Gerson O.; Pearce, Neil et al. (2022): Vaccination plus previous infection: protection during the omicron wave in Brazil. In: *The Lancet Infectious Diseases* 22 (7), S. 945–946. DOI: 10.1016/S1473-3099(22)00288-2.
7. Cerqueira-Silva, Thiago; Araujo Oliveira, Vinicius de; Paixão, Enny S.; Júnior, Juracy Bertoldo; Penna, Gerson O.; Werneck, Guilherme L. et al. (2022): Duration of protection of CoronaVac plus heterologous BNT162b2 booster in the Omicron period in Brazil. In: *Nature communications* 13 (1), S. 4154. DOI: 10.1038/s41467-022-31839-7.
8. Cheng, Franco Wing Tak; Fan, Min; Wong, Carlos King Ho; Chui, Celine Sze Ling; Lai, Francisco Tsz Tsun; Li, Xue et al. (2022): The effectiveness and safety of mRNA (BNT162b2) and inactivated (CoronaVac) COVID-19 vaccines among individuals with chronic kidney diseases. In: Kidney international 102 (4), S. 922–925. DOI: 10.1016/j.kint.2022.07.018.
9. Chin, Elizabeth T.; Leidner, David; Lamson, Lauren; Lucas, Kimberley; Studdert, David M.; Goldhaber-Fiebert, Jeremy D. et al. (2022): Protection against Omicron conferred by mRNA primary vaccine series, boosters, and prior infection. In: medRxiv. DOI: 10.1101/2022.05.26.22275639.
10. Cohen, Matan J.; Oster, Yonatan; Moses, Allon E.; Spitzer, Avishay; Benenson, Shmuel (2022): Association of Receiving a Fourth Dose of the BNT162b Vaccine With SARS-CoV-2 Infection Among Health Care Workers in Israel. In: JAMA network open 5 (8), e2224657. DOI: 10.1001/jamanetworkopen.2022.24657.
11. Collie, Shirley; Champion, Jared; Moultrie, Harry; Bekker, Linda-Gail; Gray, Glenda (2022): Effectiveness of BNT162b2 Vaccine against Omicron Variant in South Africa. In: N Engl J Med 386 (5), S. 494–496. DOI: 10.1056/NEJMc2119270.
12. Collie, Shirley; Nayager, Jiren; Bamford, Lesley; Bekker, Linda-Gail; Zylstra, Matt; Gray, Glenda (2022): Effectiveness and Durability of the BNT162b2 Vaccine against Omicron Sublineages in South Africa. In: N Engl J Med. DOI: 10.1056/NEJMc2210093.
13. Consonni, Dario; Lombardi, Andrea; Mangioni, Davide; Bono, Patrizia; Oggioni, Massimo; Uceda Renteria, Sara et al. (2022): Immunogenicità ed efficacia del vaccino anti-COVID-19 BNT162b2 in una coorte di operatori sanitari a Milano. In: *Epidemiologia e prevenzione* 46 (4), S. 250–258. DOI: 10.19191/EP22.4.A513.065.
14. Ferdinands, Jill M.; Rao, Suchitra; Dixon, Brian E.; Mitchell, Patrick K.; DeSilva, Malini B.; Irving, Stephanie A. et al. (2022): Waning 2-Dose and 3-Dose Effectiveness of mRNA Vaccines Against COVID-19-Associated Emergency Department and Urgent Care Encounters and Hospitalizations Among Adults During Periods of Delta and Omicron Variant Predominance - VISION Network, 10 States, August 2021-January 2022. In: MMWR Morb. Mortal. Wkly. Rep. 71 (7), S. 255–263. DOI: 10.15585/mmwr.mm7107e2.
15. Florentino, Pilar T. V.; Alves, Flávia J. O.; Cerqueira-Silva, Thiago; Oliveira, Vinicius de Araújo; Júnior, Juracy B. S.; Jantsch, Adelson G. et al. (2022): Vaccine effectiveness of CoronaVac against COVID-19 among children in Brazil during the Omicron period. In: Nature communications 13 (1), S. 4756. DOI: 10.1038/s41467-022-32524-5.

16. Glatman-Freedman, Aharona; Bromberg, Michal; Hershkovitz, Yael; Sefty, Hanna; Kaufman, Zalman; Dichtiar, Rita; Keinan-Boker, Lital (2022): Effectiveness of BNT162b2 Vaccine Booster against SARS-CoV-2 Infection and Breakthrough Complications, Israel. In: Emerging infectious diseases 28 (5), S. 948–956. DOI: 10.3201/eid2805.220141.
17. Gray, Glenda; Collie, Shirley; Goga, Ameena; Garrett, Nigel; Champion, Jared; Seocharan, Ishen et al. (2022): Effectiveness of Ad26.COV2.S and BNT162b2 Vaccines against Omicron Variant in South Africa. In: N Engl J Med 386 (23), S. 2243–2245. DOI: 10.1056/NEJMc2202061.
18. Guedalia, Joshua; Lipschuetz, Michal; Calderon-Margalit, Ronit; Cohen, Sarah M.; Goldman-Wohl, Debra; Kaminer, Tali et al. (2022): Effectiveness of BNT162b2 mRNA COVID-19 Third Vaccines During Pregnancy: A National Observational Study in Israel. In: SSRN Journal. DOI: 10.2139/ssrn.4159559.
19. Halasa, Natasha B.; Olson, Samantha M.; Staat, Mary A.; Newhams, Margaret M.; Price, Ashley M.; Pannaraj, Pia S. et al. (2022): Maternal Vaccination and Risk of Hospitalization for Covid-19 among Infants. In: N Engl J Med 387 (2), S. 109–119. DOI: 10.1056/NEJMoa2204399.
20. Hansen, Christian Holm; Schelde, Astrid Blicher; Moustsen-Helm, Ida Rask; Emborg, Hanne-Dorthe; Krause, Tyra Grove; Mølbak, Kåre; Valentiner-Branth, Palle (2021): Vaccine effectiveness against SARS-CoV-2 infection with the Omicron or Delta variants following a two-dose or booster BNT162b2 or mRNA-1273 vaccination series: A Danish cohort study.
21. Hertz, Tomer; Levy, Shlomia; Ostrovsky, Daniel; Oppenheimer, Hannah; Zismanov, Shosh; Kuzmina, Alona et al. (2022): Correlates of protection for booster doses of the BNT162b2 vaccine. DOI: 10.21203/rs.3.rs-1915346/v1.
22. Huang, Zhuoying; Xu, Shuangfei; Liu, Jiechen; Wu, Linlin; Qiu, Jing; Wang, Nan et al. (2022): Effectiveness of inactivated and Ad5-nCoV COVID-19 vaccines against SARS-CoV-2 Omicron BA. 2 variant infection, severe illness, and death. DOI: 10.1101/2022.09.04.22279587.
23. Jara, Alejandro; Undurraga, Eduardo A.; Zubizarreta, José R.; González, Cecilia; Acevedo, Johanna; Pizarro, Alejandra et al. (2022): Effectiveness of CoronaVac in children 3-5 years of age during the SARS-CoV-2 Omicron outbreak in Chile. In: Nature medicine 28 (7), S. 1377–1380. DOI: 10.1038/s41591-022-01874-4.
24. Jorgensen, Sarah C. J.; Hernandez, Alejandro; Fell, Deshayne; Austin, Peter C.; D'Souza, Rohan; Guttmann, Astrid et al. (2022): Effectiveness of Maternal mRNA COVID-19 Vaccination During Pregnancy Against Delta and Omicron SARS-CoV-2 Infection and Hospitalization in Infants: A Test-Negative Design Study. In: SSRN Journal. DOI: 10.2139/ssrn.4246651.
25. Kim, Sung Ran; Kang, Hyeon Jeong; Jeong, Hye Rin; Jang, Su Yeon; Lee, Jae Eun; Da Kim, Eun et al. (2022): Relative Effectiveness of COVID-19 Vaccination in Healthcare Workers: 3-Dose Versus 2-Dose Vaccination. In: Journal of Korean medical science 37 (35), e267. DOI: 10.3346/jkms.2022.37.e267.
26. Moghnieh, Rima; El Hajj, Claude; Abdallah, Dania; Jbeily, Nayla; Bizri, Abdul Rahman; Sayegh, Mohamed H. (2022): Immunogenicity and Effectiveness of Primary and Booster Vaccine Combination Strategies during Periods of SARS-CoV-2 Delta and Omicron Variants. In: Vaccines 10 (10). DOI: 10.3390/vaccines10101596.
27. Natarajan, Karthik; Prasad, Namrata; Dascomb, Kristin; Irving, Stephanie A.; Yang, Duck-Hye; Gaglani, Manjusha et al. (2022): Effectiveness of Homologous and Heterologous COVID-19 Booster Doses Following 1 Ad.26.COV2.S (Janssen Johnson & Johnson) Vaccine Dose Against COVID-19-Associated Emergency Department and Urgent Care Encounters and Hospitalizations Among Adults - VISION Network, 10 States, December 2021-March 2022. In: MMWR Morb. Mortal. Wkly. Rep. 71 (13), S. 495–502. DOI: 10.15585/mmwr.mm7113e2.
28. Kein Zugriff
29. Powell, Annabel A.; Kirsebom, Freja; Stowe, Julia; McOwat, Kelsey; Saliba, Vanessa; Ramsay, Mary E. et al. (2022): Effectiveness of BNT162b2 against COVID-19 in adolescents. In: The Lancet Infectious Diseases 22 (5), S. 581–583. DOI: 10.1016/S1473-3099(22)00177-3.
30. Ranzani, Otavio T.; Hitchings, Matt D.T.; Leite de Melo, Rosana; França, Giovanny V. A. de; Fernandes, Cássia de Fátima R.; Lind, Margaret L. et al. (2022): Effectiveness of an Inactivated Covid-19 Vaccine with Homologous and Heterologous Boosters against Omicron in Brazil. DOI: 10.1101/2022.03.30.22273193.
31. Regev-Yochay, Gili; Gonen, Tal; Gilboa, Mayan; Mandelboim, Michal; Indenbaum, Victoria; Amit, Sharon et al. (2022): 4th Dose COVID mRNA Vaccines' Immunogenicity & Efficacy Against Omicron VOC. DOI: 10.1101/2022.02.15.22270948.
32. Richterman, Aaron; Behrman, Amy; Brennan, Patrick J.; O'Donnell, Judith A.; Snider, Christopher K.; Chaiyachati, Krisda H. (2022): Durability of SARS-CoV-2 mRNA Booster Vaccine Protection Against Omicron Among Health Care Workers with a Vaccine Mandate. In: Clinical infectious diseases : an official publication of the Infectious Diseases Society of America. DOI: 10.1093/cid/ciac454.
33. Risk, Malcolm; Miao, Heidi; Freed, Gary; Shen, Chen (2022): Vaccine Effectiveness, School Reopening, and Risk of Omicron Infection Among Adolescents Aged 12-17 Years. In: The Journal of adolescent health : official publication of the Society for Adolescent Medicine. DOI: 10.1016/j.jadohealth.2022.09.006.
34. Schrag, Stephanie J.; Verani, Jennifer R.; Dixon, Brian E.; Page, Jessica M.; Butterfield, Kristen A.; Gaglani, Manjusha et al. (2022): Estimation of COVID-19 mRNA Vaccine Effectiveness Against Medically Attended COVID-19 in Pregnancy During Periods of Delta and Omicron Variant Predominance in the United States. In: JAMA network open 5 (9), e2233273. DOI: 10.1001/jamanetworkopen.2022.33273.
35. Simwanza, John; Hines, Jonas Z.; Sinyange, Danny; Sinyange, Nyambe; Mulenga, Chilufya; Hanyinza, Sarah et al. (2022): COVID-19 vaccine effectiveness during a prison outbreak when the Omicron was the dominant circulating variant— Zambia, December 2021. DOI: 10.1101/2022.05.06.22274701.
36. Spensley, Katrina J.; Gleeson, Sarah; Martin, Paul; Thomson, Tina; Clarke, Candice L.; Pickard, Graham et al. (2022): Comparison of Vaccine Effectiveness Against the Omicron (B.1.1.529) Variant in Hemodialysis Patients. In: Kidney international reports 7 (6), S. 1406–1409. DOI: 10.1016/j.ekir.2022.04.005.
37. Stirrup, Oliver; Shrotri, Madhumita; Adams, Natalie L.; Krutikov, Maria; Nacer-Laidi, Hadjer; Azmi, Borscha et al. (2022): Clinical effectiveness of SARS-CoV-2 booster vaccine against Omicron infection in residents and staff of Long-Term Care Facilities: a prospective cohort study (VIVALDI). DOI: 10.1101/2022.08.08.22278532.
38. Suah, Jing Lian; Tng, Boon Hwa; Tok, Peter Seah Keng; Husin, Masliyana; Thevananthan, Thevesh; Peariasamy, Kalaiarasu M.; Sivasampu, Sheamini (2022): Real-world effectiveness of homologous and heterologous BNT162b2, CoronaVac, and AZD1222 booster vaccination against Delta and Omicron SARS-CoV-2 infection. In: *Emerging microbes & infections* 11 (1), S. 1343–1345. DOI: 10.1080/22221751.2022.2072773.

39. Tan, Celine Y.; Chiew, Calvin J.; Lee, Vernon J.; Ong, Benjamin; Lye, David Chien; Tan, Kelvin Bryan (2022): Effectiveness of a Fourth Dose of COVID-19 mRNA Vaccine Against Omicron Variant Among Elderly People in Singapore. In: *Annals of internal medicine*. DOI: 10.7326/M22-2042.
40. Tartof, Sara Y.; Slezak, Jeff M.; Puzniak, Laura; Hong, Vennis; Frankland, Timothy B.; Ackerson, Bradley K. et al. (2022): BNT162b2 vaccine effectiveness against SARS-CoV-2 omicron BA.4 and BA.5. In: The Lancet. Infectious diseases 22 (12), S. 1663–1665. DOI: 10.1016/S1473-3099(22)00692-2.
41. Tenforde, Mark W.; Self, Wesley H.; Gaglani, Manjusha; Ginde, Adit A.; Douin, David J.; Talbot, H. Keipp et al. (2022): Effectiveness of mRNA Vaccination in Preventing COVID-19-Associated Invasive Mechanical Ventilation and Death - United States, March 2021-January 2022. In: MMWR Morb. Mortal. Wkly. Rep. 71 (12), S. 459–465. DOI: 10.15585/mmwr.mm7112e1.
42. Thompson, Mark G.; Natarajan, Karthik; Irving, Stephanie A.; Rowley, Elizabeth A.; Griggs, Eric P.; Gaglani, Manjusha et al. (2022): Effectiveness of a Third Dose of mRNA Vaccines Against COVID-19-Associated Emergency Department and Urgent Care Encounters and Hospitalizations Among Adults During Periods of Delta and Omicron Variant Predominance - VISION Network, 10 States, August 2021-January 2022. In: MMWR Morb. Mortal. Wkly. Rep. 71 (4), S. 139–145. DOI: 10.15585/mmwr.mm7104e3.
43. UKHSA (2022): COVID-19 vaccine surveillance report. Week 44. Online verfügbar unter https://assets.publishing.service.gov.uk/government/uploads/system/uploads/attachment_data/file/1115385/Vaccine_surveillance_report___week-44.pdf.
44. UKHSA (2022): COVID-19 vaccine surveillance report. Week 4. Online verfügbar unter https://assets.publishing.service.gov.uk/government/uploads/system/uploads/attachment_data/file/1050721/Vaccine-surveillance-report-week-4.pdf.
45. Wan, Eric Yuk Fai; Mok, Anna Hoi Ying; Yan, Vincent Ka Chun; Wang, Boyuan; Zhang, Ran; Hong, Sabrina Nan et al. (2022): Vaccine effectiveness of BNT162b2 and CoronaVac against SARS-CoV-2 Omicron BA.2 infection, hospitalisation, severe complications, cardiovascular disease and mortality in patients with diabetes mellitus: A case control study. In: The Journal of infection 85 (5), e140-e144. DOI: 10.1016/j.jinf.2022.08.008.
46. Willett, Brian J.; Grove, Joe; MacLean, Oscar A.; Wilkie, Craig; Logan, Nicola; Lorenzo, Giuditta de et al. (2022): The hyper-transmissible SARS-CoV-2 Omicron variant exhibits significant antigenic change, vaccine escape and a switch in cell entry mechanism. DOI: 10.1101/2022.01.03.21268111.
47. Yoon, Sarang K.; Hegmann, Kurt T.; Thiese, Matthew S.; Burgess, Jefferey L.; Ellingson, Katherine; Lutrick, Karen et al. (2022): Protection with a Third Dose of mRNA Vaccine against SARS-CoV-2 Variants in Frontline Workers. In: N Engl J Med 386 (19), S. 1855–1857. DOI: 10.1056/NEJMc2201821.
48. Zerbo, Ousseny; Ray, G. Thomas; Fireman, Bruce; Layefsky, Evan; Goddard, Kristin; Lewis, Edwin et al. (2022): Maternal SARS-CoV-2 Vaccination and Infant Protection Against SARS-CoV-2 During the First 6 Months of Life. In: Research square. DOI: 10.21203/rs.3.rs-2143552/v1.
